# Defining the Proteomic and Phosphoproteomic Landscape of Circulating Extracellular Vesicles in the Diabetes Spectrum

**DOI:** 10.1101/2021.10.31.21265724

**Authors:** Yury O. Nunez Lopez, Anton Iliuk, Alejandra Petrilli, Carley Glass, Anna Casu, Richard E. Pratley

## Abstract

The purpose of this study was to characterize the proteomic and phosphoproteomic profiles of circulating extracellular vesicles (EVs) from people with normal glucose tolerance (NGT), prediabetes (PDM), and diabetes (T2DM). Archived serum samples from 30 human subjects (N=10 per group, ORIGINS study, ClinicalTrials.gov NCT02226640) were used. EVs were isolated using EVTRAP (Tymora). Mass spectrometry (LC-MS)-based methods were used to detect the global EV proteome and phosphoproteome. Differentially expressed features, correlation networks, enriched pathways, and enriched tissue-specific protein sets were identified using custom R scripts. A total of 2372 unique EV proteins and 716 unique EV phosphoproteins were identified. Unsupervised clustering of the differentially expressed (fold change≥2, P<0.05, FDR<0.05) proteins and, particularly, phosphoproteins, showed excellent discrimination among the three groups. Among characteristic changes in the PDM and T2DM EVs, “integrins switching” appeared to be a central feature. Proteins involved in oxidative phosphorylation (OXPHOS), known to be reduced in various tissues in diabetes, were significantly increased in EVs from PDM and T2DM, which suggests that an abnormally elevated EV-mediated secretion of OXPHOS components may underlie development of diabetes. We also detected a highly enriched signature of liver-specific markers among the downregulated EV proteins and phosphoproteins in both PDM and T2DM groups. This suggests that an alteration in liver EV composition and/or secretion may occur early in prediabetes. Levels of signaling molecules involved in cell death pathways were significantly altered in the circulating EVs. Consistent with the fact that patients with T2DM have abnormalities in platelet function, we detected a significant enrichment (FDR<<0.01) for upregulated EV proteins and phosphoproteins that play a role in platelet activation, coagulation, and chemokine signaling pathways in PDM and T2DM. Overall, this pilot study demonstrates the potential of EV proteomic and phosphoproteomic signatures to provide insight into the pathobiology of diabetes and its complications. These insights could lead to the development of new biomarkers of disease risk, classification, progression, and response to interventions that could allow personalization of interventions to improve outcomes.

## INTRODUCTION

Type 2 Diabetes mellitus (T2DM) affects 31 million people in the United States and 463 million globally [1], with high risk for chronic complications of cardiovascular disease (CVD), chronic kidney disease (CKD) and heart failure [2]. T2DM can be prevented, with lifestyle interventions and pharmacologic therapies targeting those at high risk of progressing [3; 4]. These interventions are not effectively employed, however, suggesting a need to personalize therapies. There is now evidence that classic T2DM is, in fact, genetically and phenotypically heterogeneous [5]. Thus, despite a large number of therapies are available to improve glucose levels in T2DM, there is a need for better biomarkers to select optimal therapies to improve outcomes and decrease morbidity, mortality and costs from diabetes.

Although good progress has been achieved in the development of biomarkers with proven clinical utility for diseases like cancer, the development and clinical implementation of biomarkers to personalize therapy in diabetes is lagging behind. Accumulating evidence indicates that extracellular vesicles (EVs) are key players in cell-to-cell communication and inter-organ crosstalk [6-9]. EVs carry unique signatures (*e*.*g*., proteins, lipids, nucleic acids) that are cell and condition specific [10-12]. Once released from cells, EVs can make their way into blood, urine, and other bodily fluids [11; 13]. Because extracellular vesicles (EVs) are abundant in bodily fluids and carry a variety of molecules such as proteins, miRNAs, and lipids involved in both physiological and pathological processes, they are particularly attractive as biomarkers. Protein phosphorylation is a major regulatory mechanism in living cells and might provide important insight into function, but a number of challenges have limited the exploration of phosphoproteins as biomarkers, including the difficulty of reliably purifying and quantifying low-abundance phosphoproteins and interference from proteins and metabolites in the biofluids [14; 15]. Recent advances in the development of EV-based technologies (i.e., using Extracellular Vesicle Total Recovery and Purification (EVTRAP) beads followed by Polymer-based Metal Affinity Capture (PolyMAC), developed by Tymora Analytical Operations, Inc) for the characterization of the EV phosphoproteome may circumvent some of these limitations [16-18]. EVTRAP is a recently developed, broad non-antibody-based affinity isolation method that produces quantitative EV yields that have proven advantageous for MS/MS proteomic and phosphoproteomic studies [16-18].

Recent evidence supports a role for EVs in the pathogenesis of T2DM [19-23]. However, little is known about the evolution of changes in EVs in the early stages of the human disease (i.e., PDM) and no characterization of the paired EV proteome and phosphoproteome across the diabetes spectrum exists.

## RESULTS

### Study design and clinical characteristics of the study cohort

A balanced subset of patients from the ORIGINS study (ClinicalTrials.gov, ID: NCT02226640) was selected for this proteomics study (Table 1). Details from the parent study cohort have been previously described [24]. For this study, a total of 30 participants (N=10 per group) were specifically selected as a subgroup that was not confounded by differences in sex, age, and obesity, which are known to affect metabolic function. Table 1 describes the summary of clinical characteristics of the study cohort.

**Table 1.**
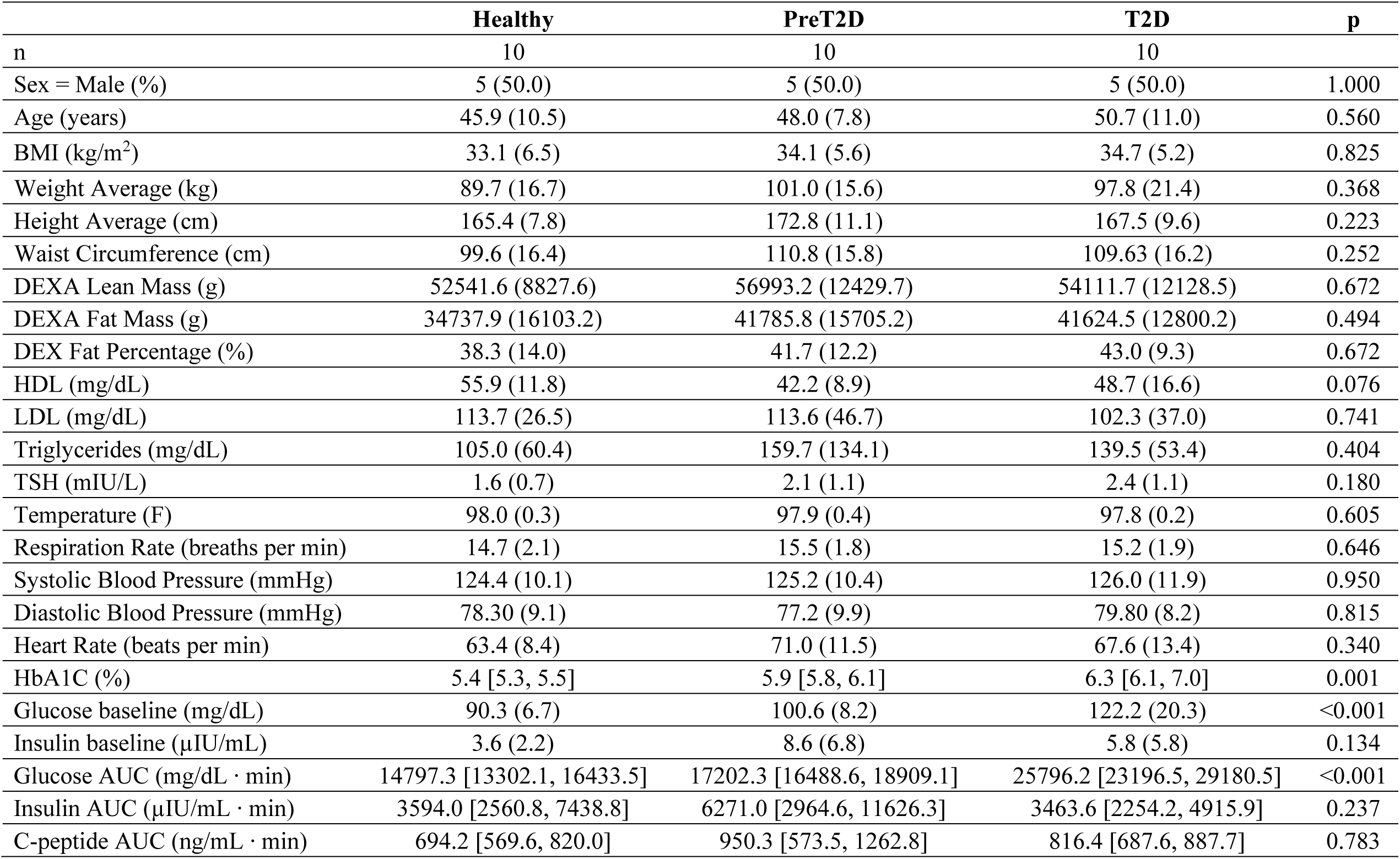
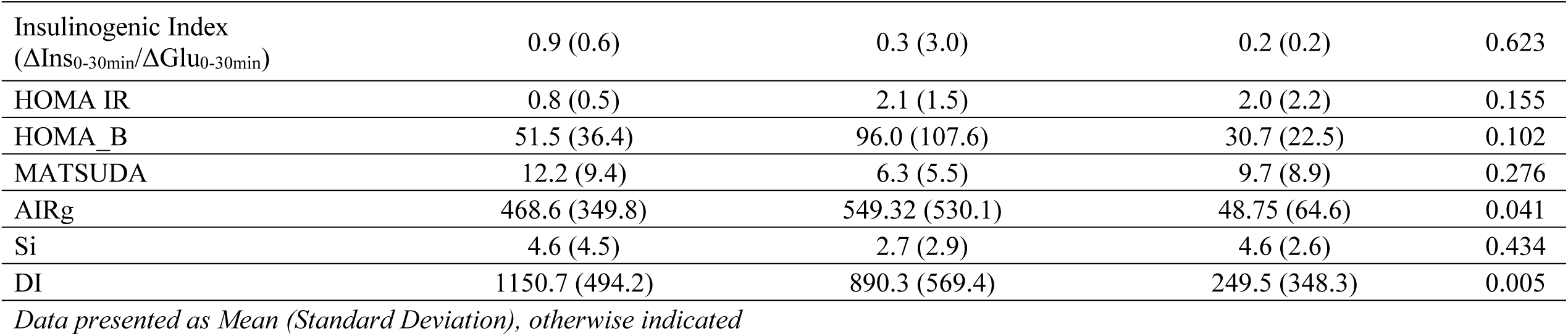
Clinical characteristics of the study cohort

|  | **Healthy** | **PreT2D** | **T2D** | **p** |
| --- | --- | --- | --- | --- |
| n | 10 | 10 | 10 |  |
| Sex = Male (%) | 5 (50.0) | 5 (50.0) | 5 (50.0) | 1.000 |
| Age (years) | 45.9 (10.5) | 48.0 (7.8) | 50.7 (11.0) | 0.560 |
| BMI (kg/m<sup>2</sup>) | 33.1 (6.5) | 34.1 (5.6) | 34.7 (5.2) | 0.825 |
| Weight Average (kg) | 89.7 (16.7) | 101.0 (15.6) | 97.8 (21.4) | 0.368 |
| Height Average (cm) | 165.4 (7.8) | 172.8 (11.1) | 167.5 (9.6) | 0.223 |
| Waist Circumference (cm) | 99.6 (16.4) | 110.8 (15.8) | 109.63 (16.2) | 0.252 |
| DEXA Lean Mass (g) | 52541.6 (8827.6) | 56993.2 (12429.7) | 54111.7 (12128.5) | 0.672 |
| DEXA Fat Mass (g) | 34737.9 (16103.2) | 41785.8 (15705.2) | 41624.5 (12800.2) | 0.494 |
| DEX Fat Percentage (%) | 38.3 (14.0) | 41.7 (12.2) | 43.0 (9.3) | 0.672 |
| HDL (mg/dL) | 55.9 (11.8) | 42.2 (8.9) | 48.7 (16.6) | 0.076 |
| LDL (mg/dL) | 113.7 (26.5) | 113.6 (46.7) | 102.3 (37.0) | 0.741 |
| Triglycerides (mg/dL) | 105.0 (60.4) | 159.7 (134.1) | 139.5 (53.4) | 0.404 |
| TSH (mIU/L) | 1.6 (0.7) | 2.1 (1.1) | 2.4 (1.1) | 0.180 |
| Temperature (F) | 98.0 (0.3) | 97.9 (0.4) | 97.8 (0.2) | 0.605 |
| Respiration Rate (breaths per min) | 14.7 (2.1) | 15.5 (1.8) | 15.2 (1.9) | 0.646 |
| Systolic Blood Pressure (mmHg) | 124.4 (10.1) | 125.2 (10.4) | 126.0 (11.9) | 0.950 |
| Diastolic Blood Pressure (mmHg) | 78.30 (9.1) | 77.2 (9.9) | 79.80 (8.2) | 0.815 |
| Heart Rate (beats per min) | 63.4 (8.4) | 71.0 (11.5) | 67.6 (13.4) | 0.340 |
| HbA1C (%) | 5.4 [5.3, 5.5] | 5.9 [5.8, 6.1] | 6.3 [6.1, 7.0] | 0.001 |
| Glucose baseline (mg/dL) | 90.3 (6.7) | 100.6 (8.2) | 122.2 (20.3) | <0.001 |
| Insulin baseline (μIU/mL) | 3.6 (2.2) | 8.6 (6.8) | 5.8 (5.8) | 0.134 |
| Glucose AUC (mg/dL · min) | 14797.3 [13302.1, 16433.5] | 17202.3 [16488.6, 18909.1] | 25796.2 [23196.5, 29180.5] | <0.001 |
| Insulin AUC (μIU/mL · min) | 3594.0 [2560.8, 7438.8] | 6271.0 [2964.6, 11626.3] | 3463.6 [2254.2, 4915.9] | 0.237 |
| C-peptide AUC (ng/mL · min) | 694.2 [569.6, 820.0] | 950.3 [573.5, 1262.8] | 816.4 [687.6, 887.7] | 0.783 |
| Insulinogenic Index<br>($\Delta\text{Ins}_{0-30\text{min}}/\Delta\text{Glu}_{0-30\text{min}}$) | 0.9 (0.6) | 0.3 (3.0) | 0.2 (0.2) | 0.623 |
| HOMA IR | 0.8 (0.5) | 2.1 (1.5) | 2.0 (2.2) | 0.155 |
| HOMA_B | 51.5 (36.4) | 96.0 (107.6) | 30.7 (22.5) | 0.102 |
| MATSUDA | 12.2 (9.4) | 6.3 (5.5) | 9.7 (8.9) | 0.276 |
| AIRg | 468.6 (349.8) | 549.32 (530.1) | 48.75 (64.6) | 0.041 |
| Si | 4.6 (4.5) | 2.7 (2.9) | 4.6 (2.6) | 0.434 |
| DI | 1150.7 (494.2) | 890.3 (569.4) | 249.5 (348.3) | 0.005 |

*Data presented as Mean (Standard Deviation), otherwise indicated*

### EVs from serum are highly enriched with exosomal proteins and phosphoproteins

A total of 2372 unique EV proteins and 716 unique EV phosphoproteins were identified from 1 mL of fasting serum. Most of these proteins and phosphoproteins have been reported to be present in extracellular vesicles (by cross-referencing to the Vesiclepedia database, Figure 1A) and specifically in exosomes (Figure 1B,C). Importantly, 91% of the top 100 exosomal proteins reported as best markers of exosomes were readily identified by our LC/MS approach. Scanning electron microscopy (SEM) of these preparations confirmed the presence of particles with morphology and dimensions consistent with those of exosomes (Figure 1D). Altogether, this data demonstrated that the EVTRAP method significantly enriched the preparations with exosomes.

**Figure 1.**
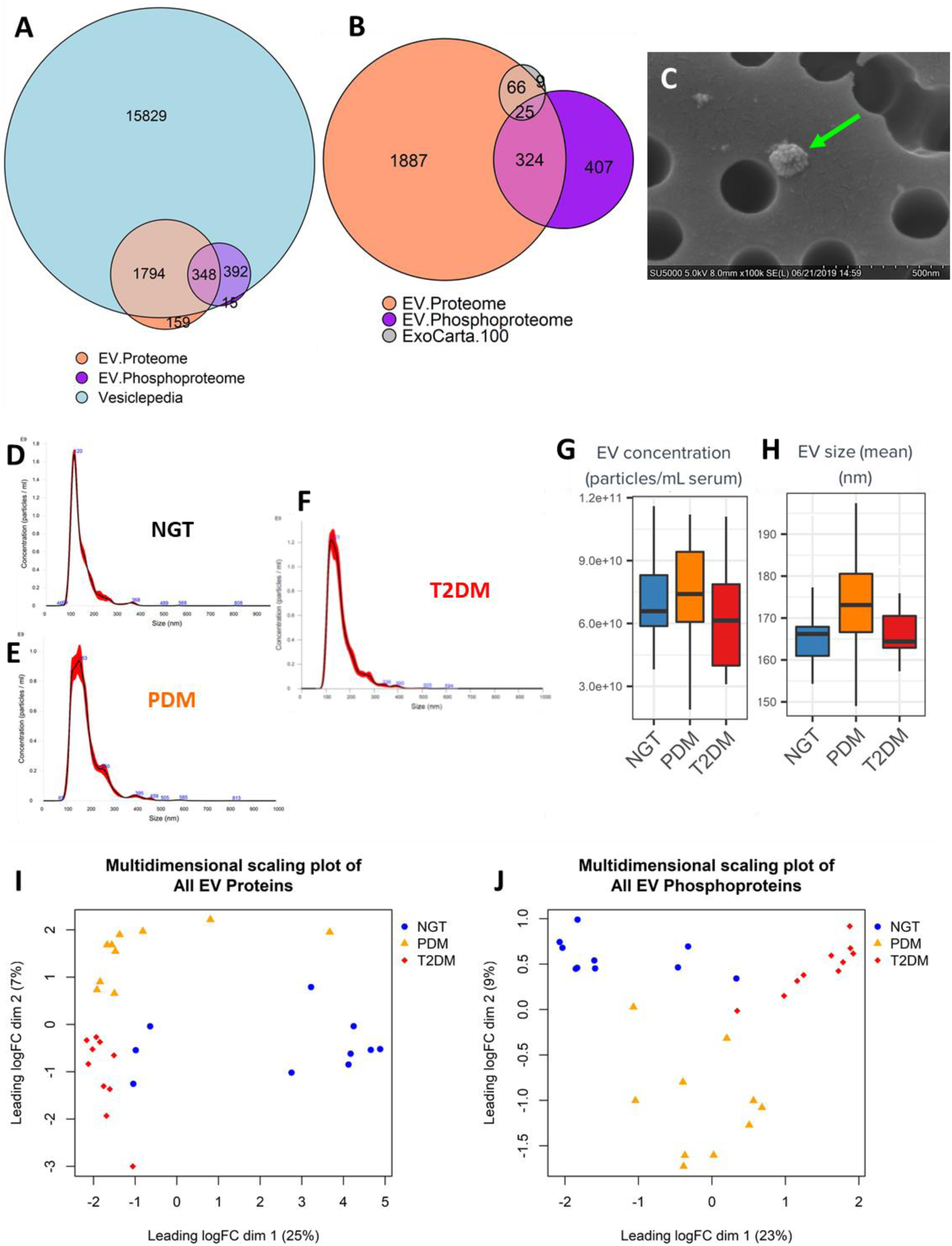
Characterization of EVs isolated from human serum from people with normal glucose tolerance (NGT), prediabetes (PDM) and type 2 diabetes mellitus (T2DM). **(A**,**B)** Euler diagram cross-referencing all reliably detected EV proteins and EV phosphoproteins to the Vesiclepedia database and to the list of best exosomal markers, as reported by Exocarta Top 100 database. **(C)** Representative scanning electron microscopy of circulating EV, **(D-F)** Representative nanoparticle tracking analysis (NTA) traces of circulating EVs, depicting size distribution of particles in the 3 study groups. **(G**,**H)** Boxplots of total EV concentration and mean EV size, respectively. **(I**,**J)** Multidimensional scaling plots using all reliably detected EV proteins and phosphoproteins, respectively.

We further characterized the distribution of particles by nanoparticle tracking analysis (NTA) and detected no significant differences in the total number of circulating EV-like nanoparticles among the three study groups (Figures 1E-I).

### Differential expression analysis provides insight on potential tissue-specific mechanisms

To gain insight into the biology underlying the development of diabetes and to identify potential circulating EV biomarkers of the disease, we implemented a multi-omic (proteomics and phosphoproteomics) approach to characterize the EV composition in serum. Multidimensional scaling analysis of all the proteomic and phosphoproteomic data (Figure 1J,K) revealed that each study group presented relatively homogeneous profiles of EV proteins and phosphoproteins that were also distinguishable from the other groups. Consequently, we were able to identify 196 and 309 differentially expressed proteins and 53 and 191 differentially expressed phosphoproteins in PDM and T2DM, respectively, as compared to NGT subjects (Supplementary Tables ST1-ST4). Using these circulating EV signatures and unsupervised clustering, we were able to correctly assign study participants to their respective groups with close to 100% accuracy (Figure 2,3). Notably, the EV proteome and phosphoproteome displayed a relatively small number of commonly detected features (specifically, 245 proteins were also found as phosphorylated proteins in the circulating EVs) and a rather low correlation between the corresponding common features (Figure 4A-C). Only 25 features were differentially expressed in both protein and phosphoprotein states, but often in opposite directions, with AKT1 being a notable example (Figure 4B,C, Supplementary Tables ST5-ST6). Supporting the relevance of this finding, the change in phosphorylated AKT1 in circulating EVs significantly negatively correlated with the change in multiple relevant clinical measures including acute insulin response to glucose (AIRg), disposition index (DI), insulin secretion (HOMA-B), fasting plasma glucose (FPG), glucose AUC, and HbA1c (absolute r ≥ 0.37, P≤0.05, Supplementary Table ST7). Similarly, the change in total AKT1 protein significantly negatively correlated with FPG, glucose AUC, and HbA1c (r ≤ -0.39, P < 0.013, Supplementary Table ST8).

**Figure 2.**
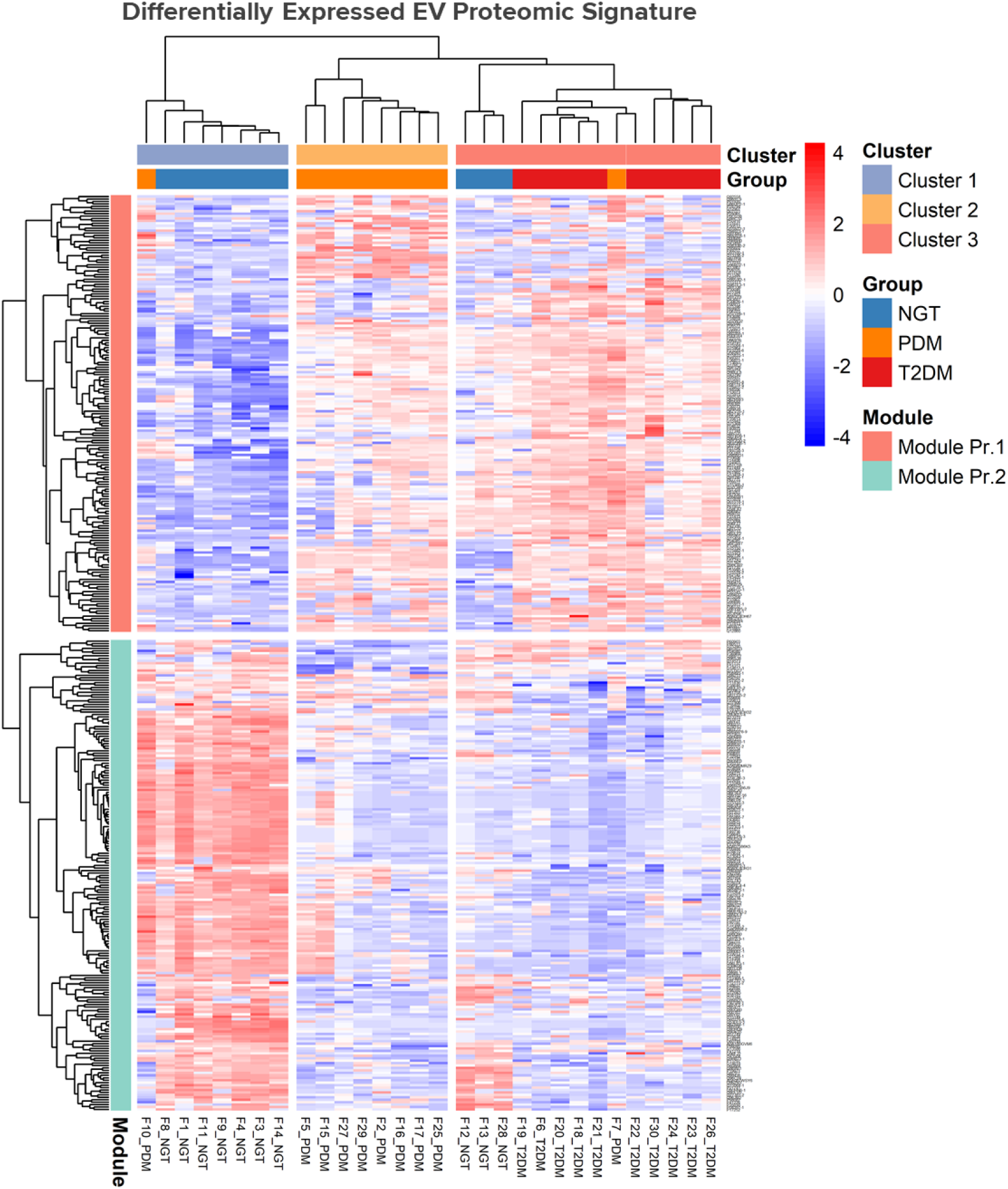
Heatmap of differentially expressed EV proteins reliably discriminate the 3 study groups.

**Figure 3.**
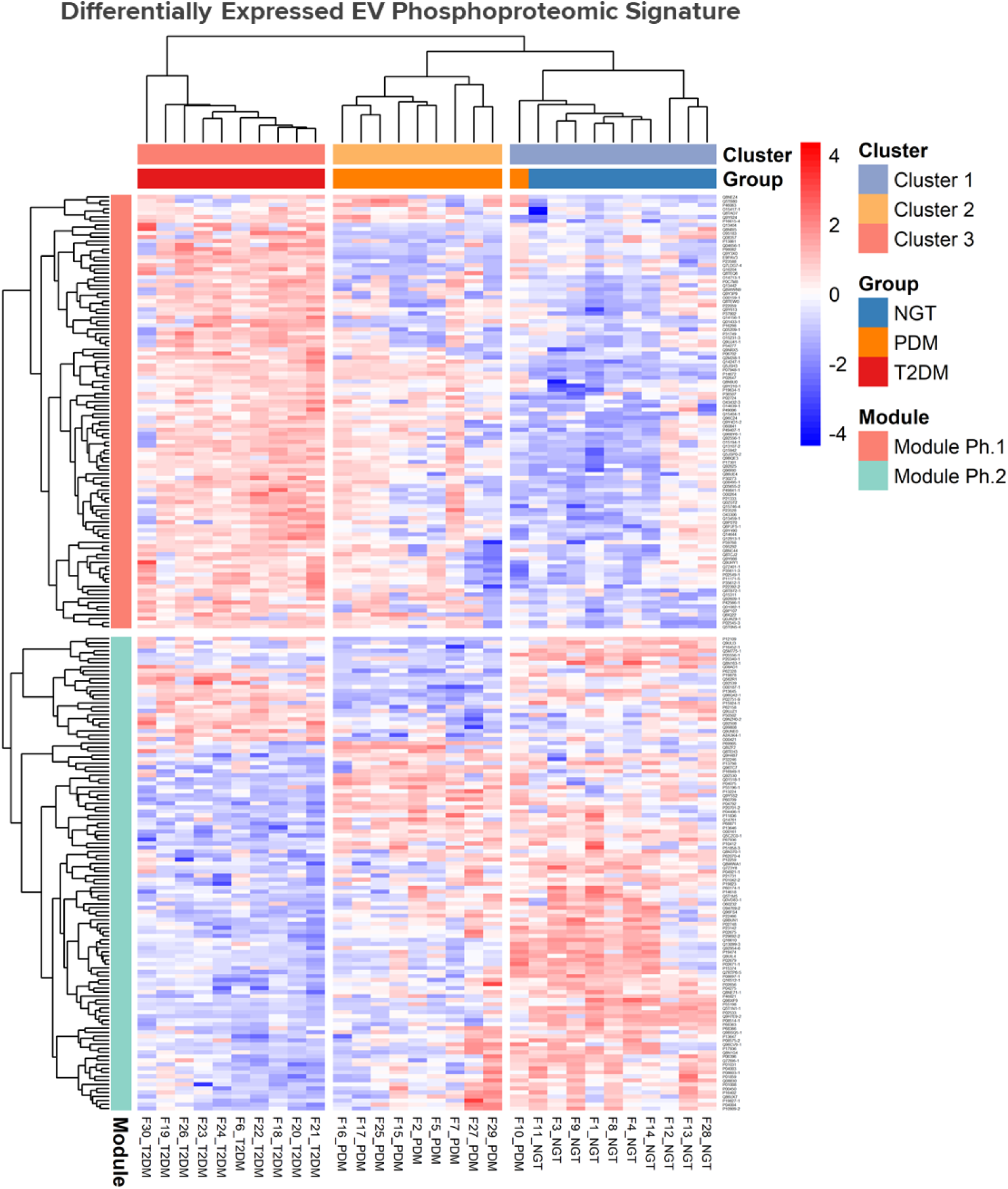
Heatmap of differentially expressed EV phosphoproteins discriminate the 3 study groups with almost 100% accuracy.

**Figure 4.**
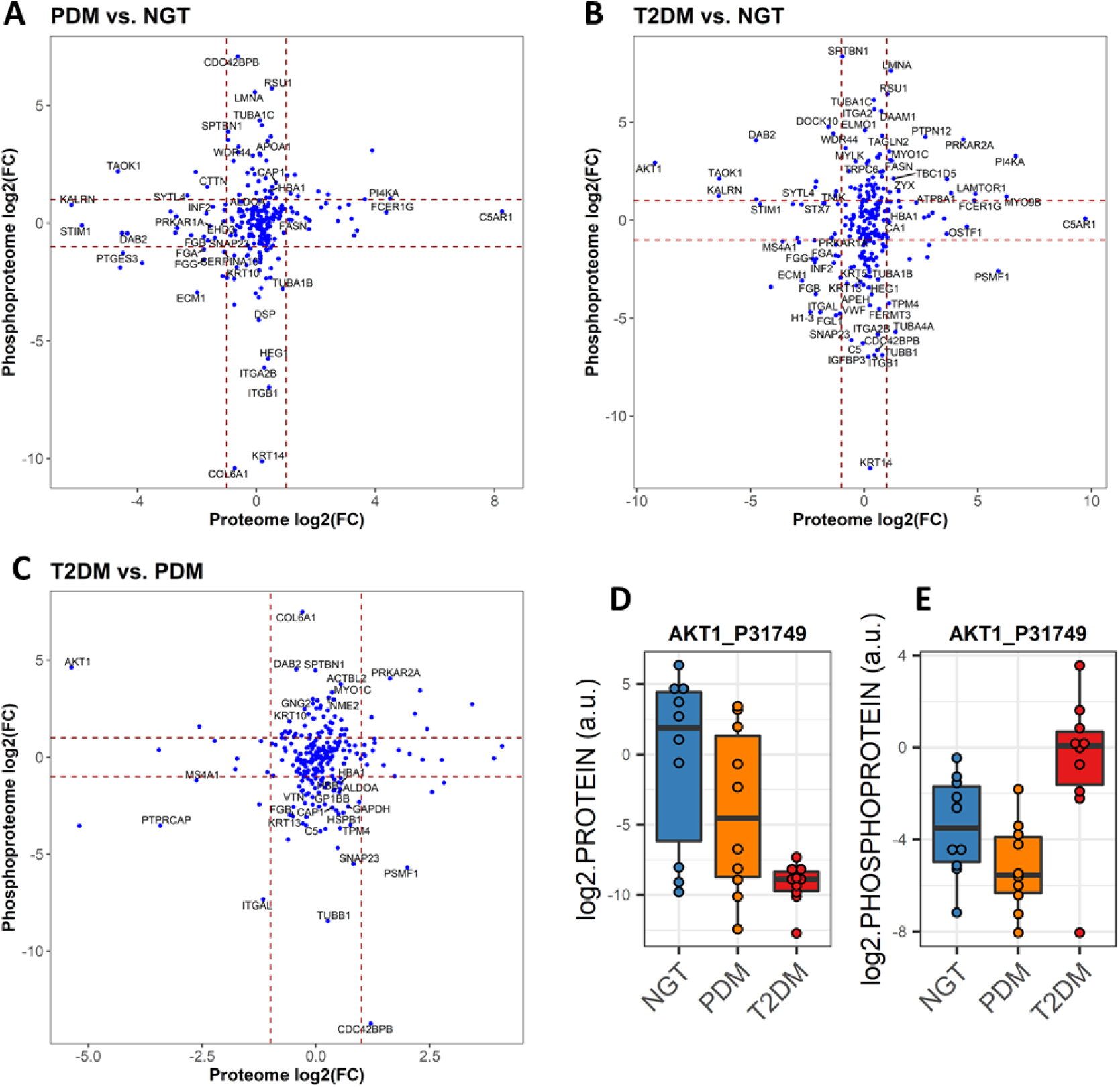
Global comparison of changes in the EV proteome and the EV phosphoproteome. **(A)** Correlation of changes in PDM, as compared to NGT. **(B)** Correlation of changes in T2DM, as compared to NGT. **(C)** Correlation of changes in T2DM, as compared to PDM. **(D**,**E)** Boxplots for total EV AKT1 protein and phosphorylated EV AKT1.

In addition, signaling kinases including AKT1, GSK3B, LYN, MAP2K2, and PRKCD were all among the significantly upregulated EV phosphoproteins that highlighted a network enriched for immune-related pathways including chemokine signaling, Fc gamma R-mediated phagocytosis, and B cell receptor signaling, among others (Figure 5C, Figure 6A-D). Similar to phosphorylated AKT1, the change in phosphorylated LYN and PRKCD kinases also significantly correlated with the change in FPG, glucose AUC, HbA1c, and the acute insulin response to glucose (AIRg) (absolute |r| ≥ 0.5, P < 0.12, Supplementary Table ST7). On the other hand, the enrichment analysis of the differentially expressed EV proteins highlighted respective networks in both PDM and T2DM with significant enrichment in oxidative phosphorylation (OXPHOS) signaling among the upregulated proteins (Figure 5D,F). An enrichment in upregulated proteins involved in immune cell mediate cytotoxicity was also detected in the PDM group (Figure 5D).

**Figure 5.**
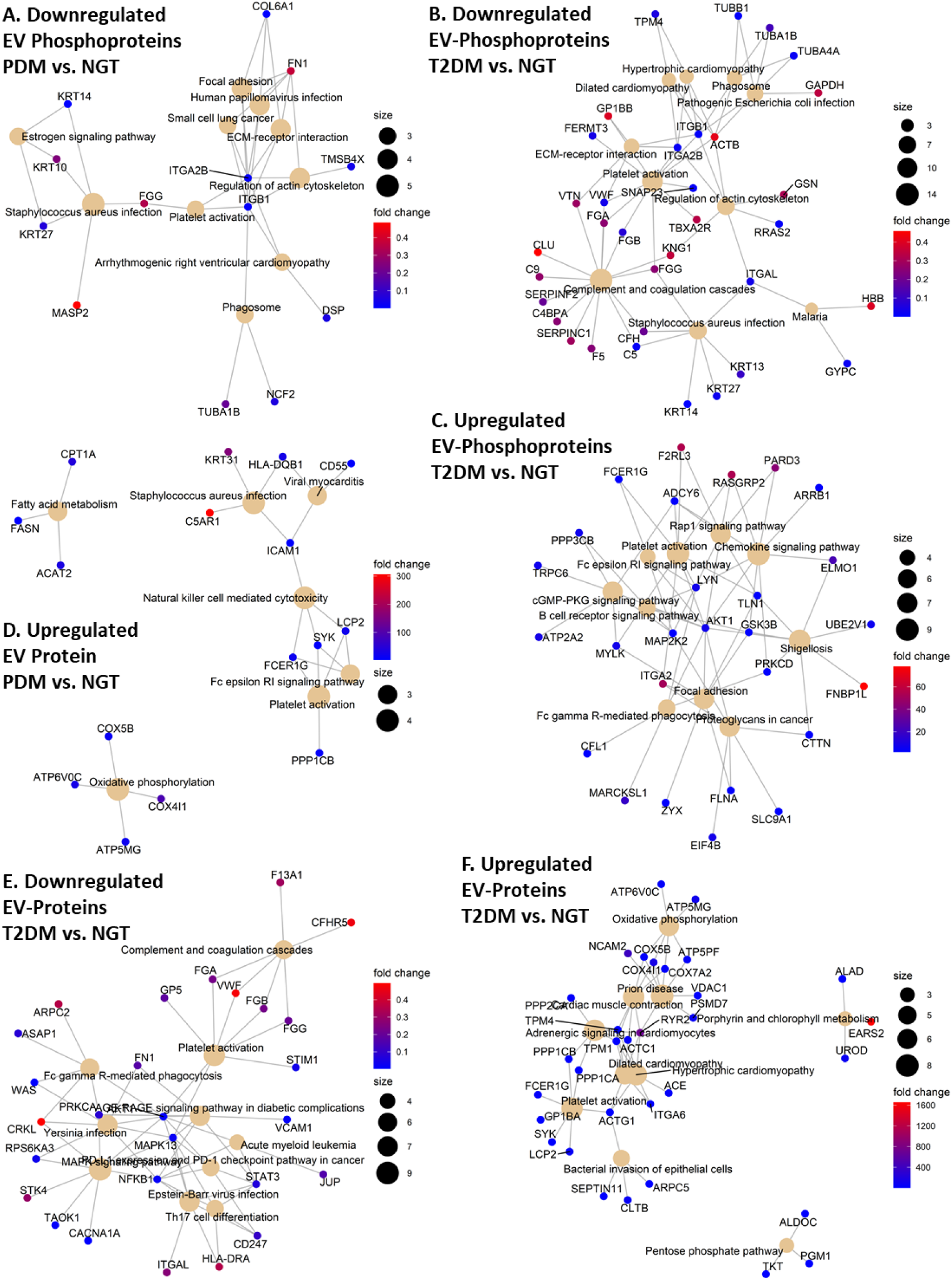
Pathway enrichment networks. **(A**,**B)** Network of enriched KEGG pathways in EV phosphoproteins that are downregulated in PDM compared to NGT (A) and in T2DM compared to NGT (B). **(C)** Network of enriched KEGG pathways in EV phosphoproteins that are upregulated in T2DM compared to NGT. **(D)** Network of enriched KEGG pathways in EV proteins that are upregulated in PDM compared to NGT. **(E)** Network of enriched KEGG pathways in EV proteins that are downregulated in T2DM compared to NGT. **(F)** Network of enriched KEGG pathways in EV proteins that are upregulated in T2DM compared to NGT.

**Figure 6.**
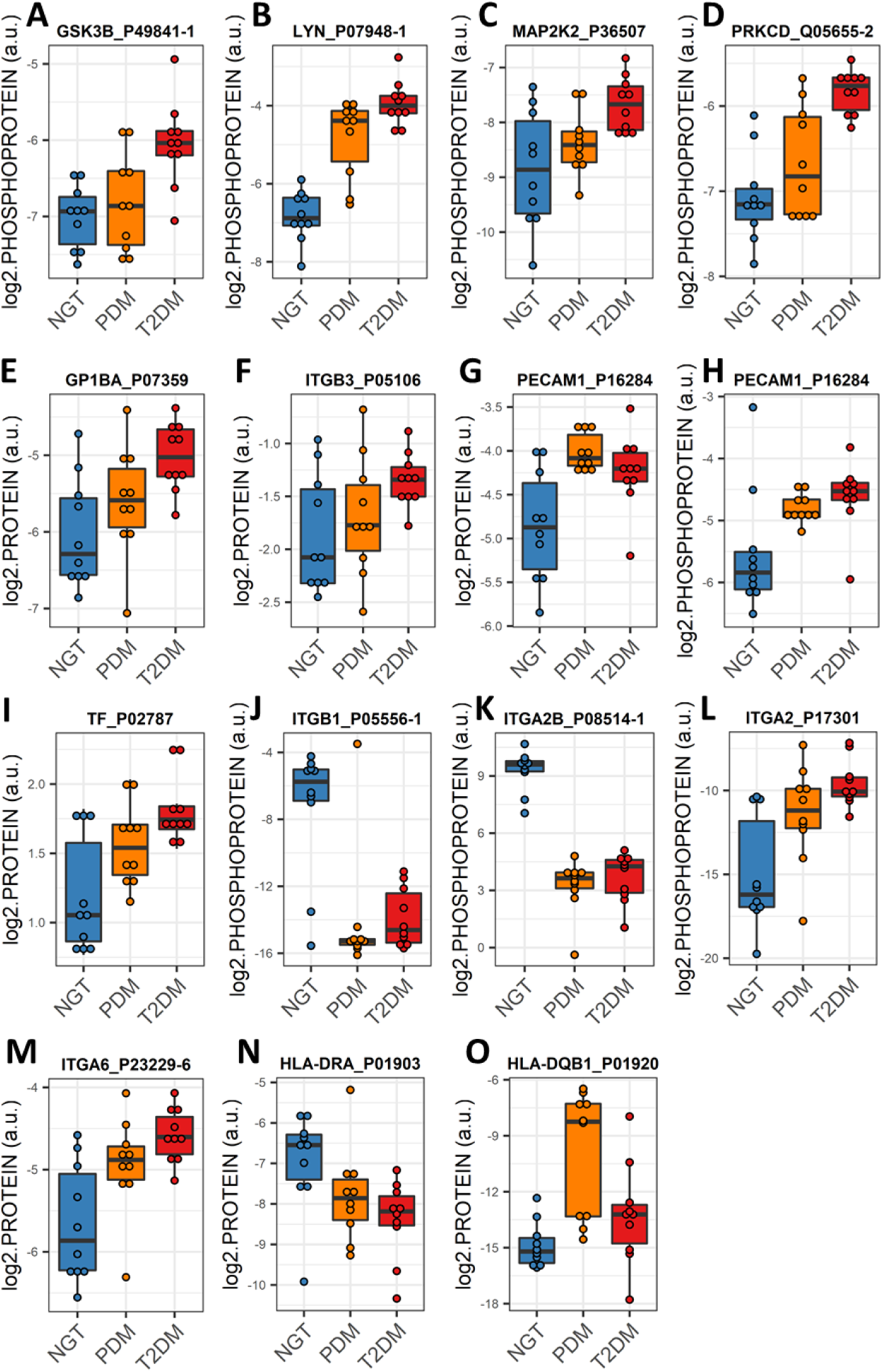
Expression profiles of select EV proteins and phosphoproteins. **(A-D)** Boxplots of differentially expressed EV proteins that are surface markers (A,B) and activation markers (C,D) of platelets. **(E)** Boxplot of differentially expressed tissue factor (TF), primary initiator of coagulation. **(F-I)** Boxplots of differentially expressed integrins. **(J**,**K)** Boxplots of differentially expressed major histocompatibility complex proteins in EVs.

### Altered expression of platelet and immune activation and coagulation markers in circulating EVs is common in PDM and T2DM

Proteins and phosphoproteins that play a role in chemokine signaling pathways in PDM and in platelet activation and coagulation in PDM and T2DM (Figure 5) were enriched among the differentially expressed EV proteins and phosphoproteins. Of note, significantly increased levels of the platelet surface markers GP1BA and integrin ITGB3 and the activation marker PCAM1, were common in EV preparations from both PDM and T2DM groups (Figure 6E-H). In addition, significant upregulation of tissue factor (TF) was also common in the circulating EVs from both groups (Figure 6I). TF in complex with coagulation factor F7a is the primary initiator of blood coagulation and has been reported to be released in EVs from platelets, monocytes, and pancreatic tumor cells contributing to thrombus formation [25].

### An integrin switch signature in circulating EVs is characteristic of PDM and T2DM

The networks of KEGG pathway-enriched downregulated EV phosphoproteins additionally highlighted a central role for integrins ITGB1 and ITGA2B in both the PDM and T2DM networks, as compared to the NGT group (Figure 5A,B; Figure 6J,K). Consequently, the modulation of interactions between the extracellular matrix (ECM) and cell receptors, the regulation of the actin cytoskeleton, and the regulation of phagosome functions were also among key pathways enriched among the downregulated EV phosphoproteins (Figure 5A,B). On the other hand, phosphorylated ITGA2 and total ITGA6 protein were highly upregulated in the T2DM group (Figure 5C,F; Figure 6M,N), which suggests that T2DM might be associated with a switch of integrin surface molecules in EVs and the originating cells. HLA proteins, which are reported to modulate the expression of integrins [26] were also among the differentially expressed EV proteins highlighted by the functional enrichment networks (i.e., HLA-DQB1 and HLA-DRA, Figure 5D,E, Figure 6N,O). Remarkably, these phosphorylated integrins correlated with important clinical measures of body composition (i.e., fat mass and waist circumference), glycemic control (i.e., FPG, glucose AUC, and HbA1c), glucose disposition (i.e., DI), insulin action (i.e., HOMA-IR), and beta cell function (i.e., fasting plasma insulin –FPI) (absolute |r| ≥ 0.4, P < 0.05, Supplementary Table ST7).

### Signatures of liver proteins and phosphoproteins are downregulated in EVs as early as the prediabetes stage

To gain insight into which organs or cell types might be significantly contributing to the differences in EV protein and phosphoprotein cargo in PDM and T2DM, we conducted enrichment analyses for cell-type specific signatures extracted from the Human Protein Atlas (HPA). As shown in Figure 7, we detected a highly enriched (FDR <<< 0.05) signature of liver-specific markers among the downregulated EV proteins and phosphoproteins in both the PDM and T2DM groups, as compared to NGT. Moreover, by conducting downstream effects analysis (DEA) using Ingenuity Pathway Analysis (IPA) software, we observed that the differentially expressed EV proteome in PDM appears to code for suppression of liver cell death and hyperproliferation functions with concomitant activation of liver inflammatory processes (Figure 8). On the other hand, the same EV proteome seems to code for the downstream activation of renal damage and necrotic cell death processes in connection with renal nephritis and kidney failure pathways (Figure 8).

**Figure 7.**
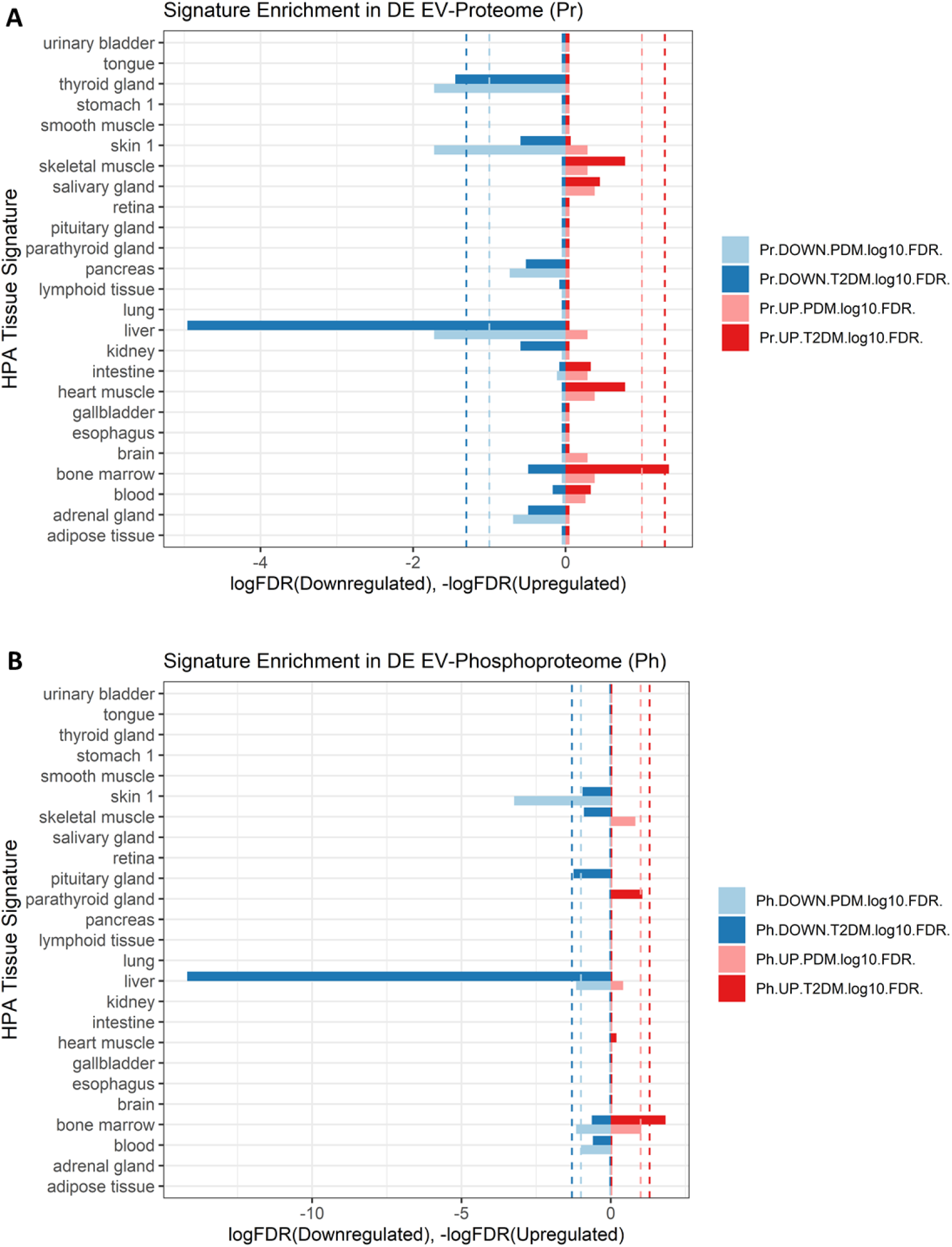
Enrichment for differentially expressed (DE) tissue-specific proteins (A) and phosphoproteins (B) in circulating EVs in people with prediabetes (PDM) and type 2 diabetes (T2DM), as compared to people with normal glucose tolerance (NGT). Pr: proteome; Ph: phosphoproteome; light blue color: downregulated features in PDM, dark blue color: downregulated features in T2DM; light red color: upregulated features in PDM, dark red color: upregulated features in T2DM.

**Figure 8.**
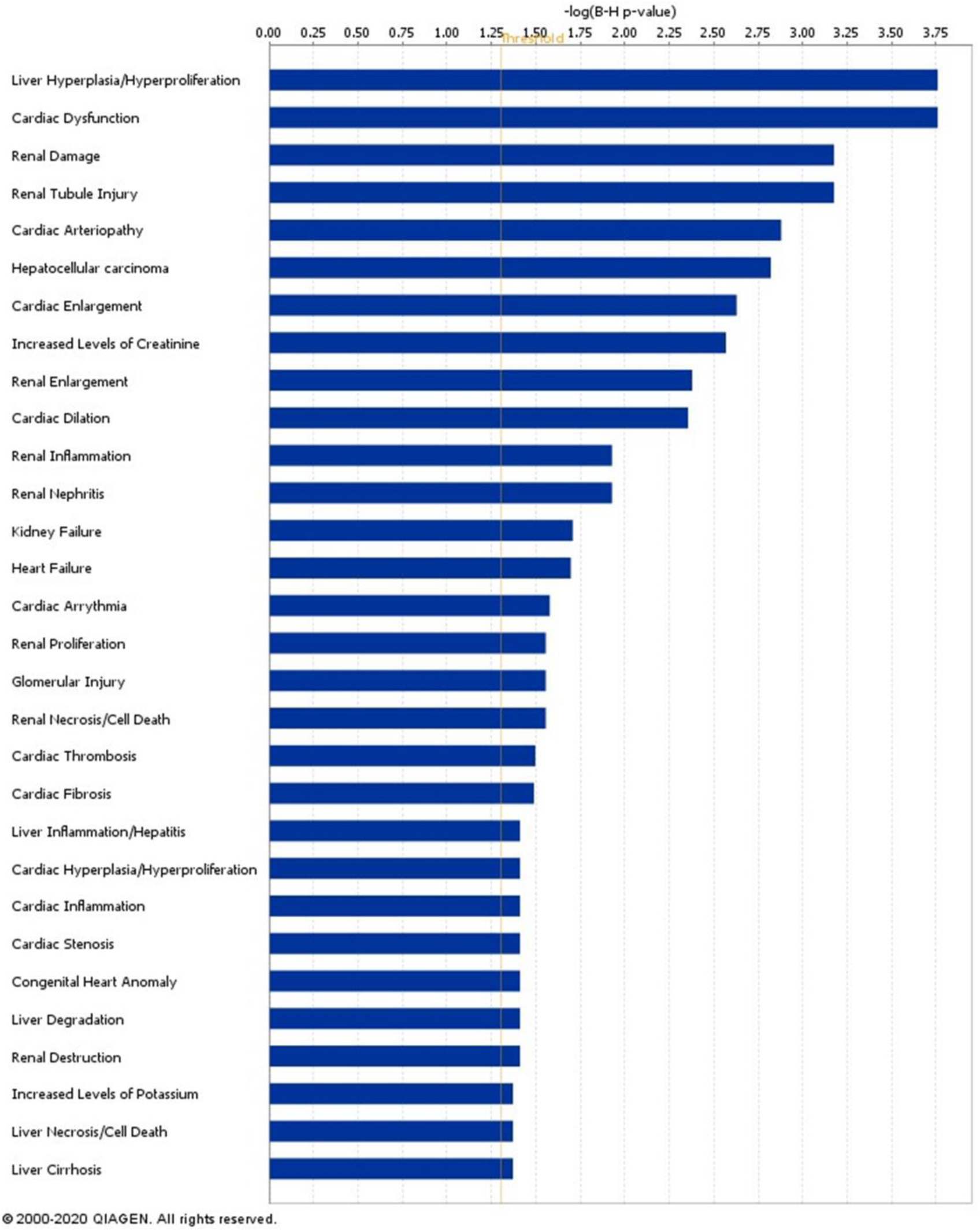
Downstream effects analysis (DEA) for differentially expressed proteins in PDM, as compared to NGT, using the Ingenuity Pathway Analysis (IPA) software.

## DISCUSSION

Understanding of the role of exosomes and EVs in type 2 diabetes has evolved in the last decade. Most EV profiling studies to date have focused on the miRNA cargo. Importantly, no study has addressed the quantification of circulating EV proteins and phosphoproteins in humans with prediabetes. Most studies on EV proteomics and phosphoproteomics have been conducted in the cancer field using cultured cells, with a small number of studies characterizing the phosphoproteome of EVs from urine or plasma/serum samples from healthy people or people with other conditions [16; 18; 27-30]. Thus, our work makes an important contribution by defining the proteomic and phosphoproteomic landscape of circulating EVs in prediabetes and diabetes. As shown in Figure 1, unsupervised clustering of select EV protein and phosphoprotein signatures could accurately separate the study participant samples based on their disease stage.

In addition to identifying EV proteomic and phosphoproteomic signatures across the spectrum of diabetes, our data suggest potential EV-mediated mechanisms that might underlie the development of prediabetes and diabetes and its complications. These associations are, of course, limited by the cross-sectional nature of this study. An interesting finding from our work is that in people with T2DM (compared to both NGT and PDM groups), AKT1 was found to be highly downregulated among the EV proteins, but highly upregulated in the phosphorylated state (Figure 2). This finding is significant because phospho-AKT is a key signaling molecule downstream of the insulin receptor and “the substrates of AKT are intimately linked to the various physiological functions of insulin and are often specific to a particular cell type” [31]. Because we did not observe a significant change in the concentration of EVs circulating in the human serum of the study participants, this result suggests that an increase in cytoplasmic AKT phosphorylation in T2DM might be accompanied by a reduction in AKT secretion via EVs. Alternatively, EV secretion may not be affected but the amount of total AKT in the cells might be dramatically reduced in T2DM, along with higher phosphorylation levels than in NGT subjects. The fact that the change in EV expression of phosphorylated and total AKT correlated with multiple clinical measures related to glycemic control and beta cell function underscores the importance of these changes during diabetes development. Interestingly, impaired translocation and activation of mitochondrial AKT1 reduced the activity of mitochondrial OXPHOS Complex V in diabetic myocardium [32; 33] and oxidative phosphorylation was found to be overrepresented among the differentially expressed EV proteins and phosphoproteins in our study (Figure 4C,D). Notably, oxidative stress has been demonstrated to be a causal factor in the impairment of adipose tissue metabolism and insulin resistance [34; 35]. Proteins involved in oxidative phosphorylation are known to be reduced in various tissues in diabetes [36-39] but, surprisingly, we detected them upregulated in the circulating EVs. We reason, then, that an abnormal removal of OXPHOS-related proteins and phosphoproteins via EV secretion (e.g., in adipocytes from people with obesity) might underlie the development of diabetes. It is tempting to speculate that abnormal EV mediated disposal function may be diverting phospho-AKT1 from translocating to the mitochondria.

Other significantly upregulated phosphorylated kinases in the same pathway-enriched network as AKT1 included GSK3B, LYN, MAP2K2, and PRKCD. These kinases coordinate signaling of multiple pathways involved in immune cell activation, which were also overrepresented among differentially expressed EV proteins networks. Supporting our findings, these pathways have been shown to play important roles in obesity associated insulin resistance, among other diabetes-associated phenomena [40-42].

The differentially expressed proteins and phosphoproteins were significantly enriched in platelet activation and coagulation effectors. This is consistent with the known crosstalk between coagulation and inflammation in diabetes, which is a key determinant in the development of complications such as cardiovascular disease, diabetic nephropathy and retinopathy, among others [43-45]. Interestingly, it has recently been reported that differentially expressed EV proteins enriched in complement and coagulation cascade signaling are among the top dysregulated pathways in patients with COVID-19 [46; 47]. Lam and colleagues also found that the liver may increase EV secretion to systemically promote coagulopathy during the course of COVID-19 and this consequently contributes to the prevalent COVID-19-induced liver damage [46]. In the context of our findings highlighting a defect in EV cargo composition or EV release from the liver in people with prediabetes and diabetes, the presumable “overload” of the liver exosomal compartment during COVID-19 in the presence of diabetes likely contribute to the exacerbation of the liver damage and the severity of the diseases under interaction.

We also detected a significant reduction in phosphorylated integrins ITGB1 and ITGA2B in the circulating EVs, while phosphorylated ITGA2 and total ITGA6 protein were significantly upregulated. Of note, HLA proteins have been reported to modulate the expression of integrins [26] and we found some (i.e., HLA-) coordinately and significantly changing and highlighted in the functional enrichment networks. We speculate that these changes may indicate a switch of integrin surface markers in cells and their released EVs, which may consequently change their interaction patterns with the surrounding ECM and tropism for the circulating EVs, respectively [48; 49]. The fact that these EV proteins and phosphoproteins were identified as central hubs in their respective pathway enrichment networks and that their changes significantly correlated with changes in relevant clinical variables including measures of body composition, glycemic control, insulin action, and beta cell function suggests that their presence and changes in abundance in the circulating EVs is important in the development of T2DM. However, our evidence is only correlative, therefore we cannot determine causality. Remarkably, ECM-integrin signaling (e.g., due to increased density of the collagen-binding integrin α2β1 dimer, a key collagen-binding receptor in the plasma membrane of platelets and muscle cells, among others) has been associated with muscle insulin resistance [50; 51], inflammation [52], angiogenesis [53], and cardiovascular risk [54], among others. Our study did not aim to quantify the levels of integrin heterodimers but the fact that the phosphorylated α2 (ITGA2) subunit was elevated in the circulating EVs of both PDM and T2DM subjects may indicate elevated density and/or activity of its heterodimers in specific tissues and secreted EVs. Consistent with its role in platelet function, platelet activation was among the pathways most commonly and significantly enriched among the differentially expressed proteins and phosphoproteins detected in this study in both PDM and T2DM subjects. On the other hand, altered density or activity of integrin molecules such as integrin β1 (ITGB1) have been implicated in a variety of diabetic complications, particularly diabetic nephropathy [55-57]. ITGB1 has been found to be particularly important in podocytes, where it mediates signaling in the axis IGFBP-1/ITGB1/FAK and contributes to essential podocyte functions by promoting cell adhesion, motility, and survival [58]. The activity of this axis was found to be controlled by FOXO1, which was in turn inhibited by activated insulin-PI3K-AKT signaling [58]. Interestingly, we found phosphorylated AKT upregulated in the circulating EVs from the T2DM group. Moreover, the activity of the axis was demonstrated to be reduced in glomeruli from humans with early type 2 diabetic kidney disease (DKD). Supporting the key role of ITGB1 in the kidneys, the β1-integrin-knockout mice develops severe proteinuric kidney disease from birth [59] and patients with Abatacept-stabilized β1-integrin activation are protected from B7-1-positive proteinuric kidney disease [60]. Remarkably, Ingenuity DEA analysis suggested that the differentially expressed EV proteome identified in people with PDM in our study, is associated with the activation of pathways involved in renal damage and necrotic cell death, as well as in renal nephritis and kidney failure. Altogether, our data now suggest that profiling the integrin composition of circulating EVs could aid in the early detection of diabetic complications.

Another novel and important finding from our study is the highly significant downregulation of circulating EV proteins and phosphoproteins from the liver in both the PDM and T2DM groups. This defect may be caused by (1) decreased expression of proteins and phosphoproteins in liver cells, hence reduced levels in the liver-derived EVs, (2) by decreased packaging of proteins and phosphoproteins into EVs, (3) by reduced secretion of EVs with neither a defect in EV packaging nor in cytoplasmic levels of the specific proteins and phosphoproteins, or (4) by some combination of the previous three defects. With the data at hand, we are unable to dissect the specific cause for the alterations in EV cargo composition in the liver or any other tissue. However, the fact that additional analyses (i.e., DEA) suggested that the differentially expressed EV proteins are involved in suppression of liver cell death and hyperproliferation functions with concomitant activation of liver inflammatory processes in PDM, supports an important early role for the liver (potentially mediated by alterations in EV cargo and inter-organ cross-communication) during diabetes development. Indeed, the important role of the liver in diabetes is well established and elegantly incorporated in the Twin Cycle Hypothesis [61; 62]. Our study now suggests that alterations in the cargo and/or in the number of liver-derived circulating EVs represent early pathophysiological changes that could serve as biomarkers of disease development. These EV protein dynamics, identifiable as early as in prediabetes, appear to represent early events contributing to the known increased risk of developing steatosis and renal fibrosis in subjects with type 2 diabetes [63-65]. Notably, important links between liver, kidney, and heart pathologies have been reported and the pathogenic crosstalk between the liver and the inflamed adipose tissue is widely accepted [66; 67]. Our data adds support for a role of circulating EVs in this pathogenic crosstalk pointing at the liver and kidneys as early partners in crime.

Contrasting with our finding that no significant changes occur in the total concentration of circulating EVs in diabetes, other authors have reported increased numbers of circulating particles [21]. However, their EV isolation methods (i.e., polymer precipitation and ultracentrifugation) are less specific than the EVTRAP method employed in this study. Polymer precipitation and ultracentrifugation are known to isolate other types of contaminating particles that could mistakenly be counted as EVs. On the other hand, using flow cytometry quantification of blood cell-specific markers, the same authors reported a significant increase in erythrocyte-derived EVs in diabetes. This latter finding is in agreement with the significant enrichment that we observe in bone marrow cell-specific markers among the upregulated EV proteins and phosphoproteins in people with T2DM (Figure 7).

Study limitations include the small sample size, the cross-sectional nature, the lack of functional measures or imaging of the liver and kidneys, and the fact that we cannot separate out tissue specific EVs. Despite these limitations, our study has several strengths, including careful control for confounding factors, balanced sex, and the use of state-of-the-art methods for broad non-antibody-based specific EV isolation, proteomics, phosphoproteomics, and EV bioinformatics.

## CONCLUSIONS

This work makes an important contribution towards defining the proteomic and phosphoproteomic landscape of circulating EVs across the diabetes disease spectrum. Among key findings, our data indicate that reduced levels of AKT1 protein but with increased phosphorylation status in circulating EVs may underlie the development of pathogenic events during diabetes. Among characteristic common changes in the prediabetic and diabetic EVs, “integrins switching” appear to be a central feature of functional enrichment networks with potential impact in disease development and the known increased risk for complications. In addition, a highly significant signature of downregulated liver-specific EV proteins is demonstrated in both the EV proteome and phosphoproteome as early as prediabetes. This suggests a reduced EV output from the liver, among other possible causes, due to an impaired endocytic secretory pathway in early stages of disease development. Suppressed liver cell death functions contrasted by activated cell death functions in kidneys in prediabetes may represent early events contributing to the known increased risk for steatosis/NASH and renal fibrosis/diabetic nephropathy comorbidities in people with type 2 diabetes. We further demonstrated that upregulated EV proteins and phosphoproteins involved in platelet activation, coagulation, chemokine signaling, and oxidative phosphorylation pathways are evident early in the course of the development of diabetes.

## MATERIALS & METHODS

### Samples

All procedures were approved by the AdventHealth Translational Research Institute (AH/TRI) Institutional Review Board (IRB). Informed consent was obtained from all volunteers before initiation of the study. Archived serum samples from 30 human subjects (N=10 per group, ORIGINS study, ClinicalTrials.gov NCT02226640). The groups were selected, according to ADA guidelines [68] to have either normal glucose tolerance (NGT), prediabetes PDM) of T2DM. Subjects with type 1 diabetes or other types of diabetes were not included in the ORIGINS study. The inclusion and exclusion criteria of the subjects were described previously (ClinicalTrials.gov, ID: NCT02226640). Participants were specifically selected as a subgroup that was relatively well balanced for sex, age, and obesity. This selection was partially automated using custom script based on the *MatchIt* package [69] in the R programing environment through a series of recursive pairwise propensity score matching cycles (between two of the study groups at a time), until all pairwise comparisons were exhausted and the desired sample size was achieved).

### Clinical and metabolic measurements

Anthropometric measures were performed according to standardized protocols. Body composition was measured using a GE Lunar iDEXA whole-body scanner (GE, Madison, WI). Fasting blood samples were obtained and subjects underwent a 2-hour 75 g oral glucose tolerance test (OGTT). On a different visit, an insulin-modified frequently-sampled intravenous glucose tolerance test (FSIVGTT) [70] was performed. Plasma glucose concentrations were measured using the glucose oxidase method with a YSI 2300 STAT Plus Analyzer (YSI Life Sciences, Yellow Springs, OH). Plasma insulin and C-peptide concentrations were determined using the MSD human insulin assay kit and C-peptide kit, respectively (MSD, Rockville, MD). HbA1c levels were measured using a Cobas Integra 800 Analyzer (Roche, Basel, Switzerland). The β cell function was assessed by calculating HOMA-B, the insulinogenic index [ΔIns_0-30’_/ΔGluc_0-30’_] and the insulin and C-peptide areas under the curve (AUC) in response to OGTT. Insulin activity was assessed by calculating HOMA-IR as described elsewhere. Data from the FSIVGTT were used to calculate insulin sensitivity (Si) and acute insulin response to glucose (AIRg) using the Minimal Model method of Bergman (MINMOD-Millennium,© R. Bergman [71]).

### EV purification using EVTRAP™, a non-antibody-based affinity technology developed by Tymora to specifically and quantitatively isolate EVs

Frozen serum samples were thawed, and any large debris removed by centrifugation at 2,500 × g for 10 minutes. The pre-cleared plasma samples were then diluted 20-fold in PBS and incubated with EVTRAP beads for 30 min [17]. After supernatant removal using a magnetic separator rack, the beads were washed with PBS, and the EVs were eluted by a 10 min incubation with 200 mM triethylamine (TEA, Millipore-Sigma) and the resulting EV samples fully dried in a vacuum centrifuge.

### Mass spectrometry (LC-MS)-based methods developed by Tymora used to detect the global EV proteome and phosphoproteome

Isolated EV samples were dried and lysed to extract proteins using the phase-transfer surfactant aided procedure [16]. For this, EVs were solubilized in the lysis solution containing 12 mM sodium deoxycholate, 12 mM sodium lauroyl sarcosinate, 10 mM TCEP, 40 mM CAA, and phosphatase inhibitor cocktail (Millipore-Sigma) in 50 mM Tris·HCl, pH 8.5 by incubating 10 min at 95°C. This step also denatured, reduced and alkylated the proteins. The samples were then diluted fivefold with 50 mM triethylammonium bicarbonate and digested with Lys-C (Wako) at 1:100 (wt/wt) enzyme-to-protein ratio for 3 h at 37°C. Trypsin was added to a final 1:50 (wt/wt) enzyme-to-protein ratio for overnight digestion at 37°C. The samples were acidified with trifluoroacetic acid (TFA) to a final concentration of 1% TFA. Ethyl acetate solution was added at 1:1 ratio to the samples. The mixture was vortexed for 2 min and then centrifuged at 20,000 × g for 2 min to obtain aqueous and organic phases. The organic phase (top layer) was removed, and the aqueous phase was collected, dried down to <10% original volume in a vacuum centrifuge, and desalted using Top-Tip C18 tips (Glygen) according to manufacturer’s instructions. Each sample was split into 99% and 1% aliquots for phosphoproteomic and proteomic experiments respectively. The samples were dried completely in a vacuum centrifuge and stored at -80°C. For phosphoproteome analysis, the 99% portion of each sample was subjected to phosphopeptide enrichment using PolyMAC Phosphopeptide Enrichment kit (Tymora Analytical) according to manufacturer’s instructions, and the eluted phosphopeptides dried completely in a vacuum centrifuge. For phosphoproteomics analysis the whole enriched sample was used, while for proteomics only 50% of the sample was loaded onto LC-MS.

### Nanoparticle Tracking Analysis (NTA)

The size distribution and concentration of particles in EV preparations were analyzed using dynamic light-scattering technology with a NanoSight NS300 instrument and NTA-3.4 software (Malvern Panalytical, Malvern). The instrument was equipped with a 488 nm blue laser module, flow-cell top plate, integrated temperature control, and a single-syringe pump module. Samples were diluted using cell culture grade water (Corning cat# 25-005-CI) to produce an optimal particle concentration for final measurement in the range of 10^7^ to 10^9^ particles/ml. Dilutions were initially assessed with a single quick static measurement of 30 second to identify the optimal dilution (which represented approximately 20 to 100 particles in the instrument’s field of view, per video frame). For final, more accurate quantification, 5 standard measurements of 1 minute of duration each were taken at a controlled temperature of 25 °C and under constant automatic flow (continuous syringe pump speed set to 50 arbitrary units). Camera level for video capture was set to 12 and detection threshold to 5 for all sample measurements.

### Scanning Electron Microscopy (SEM)

Representative SEM images of EV samples were obtained at the Interdisciplinary Center for Biotechnology Research (ICBR) Electron Microscopy Core Laboratory (RRID:SCR_019146), at University of Florida. In short, EV preps were fixed in Trump’s fixative for 30 minutes at room temperature then spread onto Isopore 0.2 μm GTTP filters (Merck Millipore, Tullagreen, Ireland) that had been pre-treated with 0.01% poly-L-lysine solution. The filters were washed 3 times with 1x PBS and 3 times with filter-sterilized deionized water. The fixed EVs were microwave-stabilized in a Ted Pella Pelco Biowave, then dehydrated through an ethanol series at 10, 25, 50, 75, 90 and 100%, followed by critical-point drying (CPD) using a Tousimus CPD system. Samples were sputter coated with gold-palladium and imaged with a Hitachi SU5000 Schottky Field-Emission SEM.

### Detection of tissue-specific signatures and KEGG pathway enrichment analysis

The lists (signatures) of tissue-specific proteins were downloaded from the Human Protein Atlas [72] (https://www.proteinatlas.org/humanproteome/tissue/tissue+specific). Enrichment for the tissue-specific signatures among the lists of differentially expressed EV proteins and phosphoproteins was then assessed via implementation of the hypergeometric test using the *phyper()* function from the *stats* package in the R environment. The complete list of proteins reported by Vesiclepedia [73] (http://microvesicles.org/Archive/VESICLEPEDIA_PROTEIN_MRNA_DETAILS_4.1.txt) plus the additional novel EV proteins detected by our proteomics experiments were used as background (universe) for the hypergeometric tests. Enrichment of KEGG pathway annotations among the sets of differentially expressed proteins and phosphoproteins was assessed using the R package *clusterProfiler* [74] with a P < 0.05 and adjusted P < 0.1 as thresholds for statistical significance.

### Statistical analysis

Data normality was tested using the Shapiro-Wilk test, and nonnormal data was log-transformed to approximate normality. Differences in baseline clinical characteristics were assessed using the Welch two-sample t test (for continuous variables) or the Fisher exact test (for categorical variables). For assessment of differential expression in EV-shuttled proteins and phosphoproteins, linear models using the *limma* R package [75] were implemented. The *limma* package was originally developed for the analysis of microarray data but later adapted for RNA-seq analysis and more recently used for proteomics experiments [76-79]. Because *limma*’s empirical Bayes approach improves statistical power, its use has been proposed to be beneficial for proteomic experiments, which often have relatively small sample sizes [80]. The empirical Bayes approach specifically allows for a realistic distribution of biological variances by using the full data to shrink the observed sample variances towards a pooled estimate [75; 80]. Our models included age, sex, and BMI as established covariates. Although these variables were balanced in our study cohort, we had a priori decided to include them in the models to account for any residual confounding effects, adding stringency to the analysis. Partial correlations were also calculated in the R environment adjusting for the same covariates. Post-hoc analysis was performed using the *phia* package. Calculated effects and correlations with two-tailed P values < 0.05 were considered significant. False discovery rates (FDR) correcting for multiple testing were calculated using the Benjamini-Hochberg (BH) correction as implemented for the *p*.*adjust* function in the *stats* R package.

## Data Availability

Data produced in the present study are available upon reasonable request to the authors

## Acknowledgements

The authors thank Dr. Steven R. Smith (AdventHealth, Translational Research Institute) for providing access to the archived samples from the ORIGINS study.

## REFERENCES

[1] Saeedi, P., Petersohn, I., Salpea, P., Malanda, B., Karuranga, S., Unwin, N., et al., 2019. Global and regional diabetes prevalence estimates for 2019 and projections for 2030 and 2045: Results from the International Diabetes Federation Diabetes Atlas, 9th edition. Diabetes Research and Clinical Practice 157:107843.

[2] Wan, E.Y.F., Chin, W.Y., Yu, E.Y.T., Wong, I.C.K., Chan, E.W.Y., Li, S.X., et al., 2020. The Impact of Cardiovascular Disease and Chronic Kidney Disease on Life Expectancy and Direct Medical Cost in a 10-Year Diabetes Cohort Study. Diabetes Care 43(8):1750–1758.

[3] Tuomilehto, J., Schwarz, P., Lindström, J., 2011. Long-Term Benefits From Lifestyle Interventions for Type 2 Diabetes Prevention. Time to expand the efforts 34(Supplement 2):S210-S214.

[4] DeFronzo, R.A., Abdul-Ghani, M., 2011. Type 2 Diabetes Can Be Prevented With Early Pharmacological Intervention. Diabetes Care 34(Supplement 2):S202–S209.

[5] Pearson, E.R., 2019. Type 2 diabetes: a multifaceted disease. Diabetologia 62(7):1107–1112.

[6] Li, C.-J., Fang, Q.-H., Liu, M.-L., Lin, J.-N., 2020. Current understanding of the role of Adipose-derived Extracellular Vesicles in Metabolic Homeostasis and Diseases: Communication from the distance between cells/tissues. Theranostics 10(16):7422–7435.

[7] Romero, A., Eckel, J., 2021. Organ Crosstalk and the Modulation of Insulin Signaling. Cells 10(8):2082.

[8] Guay, C., Regazzi, R., 2017. Exosomes as new players in metabolic organ cross-talk. Diabetes, Obesity and Metabolism 19(S1):137-146.

[9] Huang, Z., Xu, A., 2021. Adipose Extracellular Vesicles in Intercellular and Inter-Organ Crosstalk in Metabolic Health and Diseases. Frontiers in immunology 12:608680–608680.

[10] Zhang, Y., Liu, Y., Liu, H., Tang, W.H., 2019. Exosomes: biogenesis, biologic function and clinical potential. Cell & bioscience 9:19–19.

[11] Yáñez-Mó, M., Siljander, P.R.M., Andreu, Z., Zavec, A.B., Borràs, F.E., Buzas, E.I., et al., 2015. Biological properties of extracellular vesicles and their physiological functions. Journal of Extracellular Vesicles 4:27066–27066.

[12] Kowal, J., Arras, G., Colombo, M., Jouve, M., Morath, J.P., Primdal-Bengtson, B., et al., 2016. Proteomic comparison defines novel markers to characterize heterogeneous populations of extracellular vesicle subtypes. Proceedings of the National Academy of Sciences 113(8):E968–E977.

[13] Boukouris, S., Mathivanan, S., 2015. Exosomes in bodily fluids are a highly stable resource of disease biomarkers. PROTEOMICS – Clinical Applications 9(3-4):358–367.

[14] Iliuk, A.B., Tao, W.A., 2013. Is phosphoproteomics ready for clinical research? Clin Chim Acta 420:23–27.

[15] Andaluz Aguilar, H., Iliuk, A.B., Chen, I.H., Tao, W.A., 2020. Sequential phosphoproteomics and N-glycoproteomics of plasma-derived extracellular vesicles. Nat Protoc 15(1):161–180.

[16] Iliuk, A., Wu, X., Li, L., Sun, J., Hadisurya, M., Boris, R.S., et al., 2020. Plasma-Derived Extracellular Vesicle Phosphoproteomics through Chemical Affinity Purification. J Proteome Res 19(7):2563–2574.

[17] Wu, X., Li, L., Iliuk, A., Tao, W.A., 2018. Highly Efficient Phosphoproteome Capture and Analysis from Urinary Extracellular Vesicles. J Proteome Res 17(9):3308–3316.

[18] Chen, I.H., Xue, L., Hsu, C.C., Paez, J.S., Pan, L., Andaluz, H., et al., 2017. Phosphoproteins in extracellular vesicles as candidate markers for breast cancer. Proc Natl Acad Sci U S A 114(12):3175–3180.

[19] Noren Hooten, N., Evans, M.K., 2020. Extracellular vesicles as signaling mediators in type 2 diabetes mellitus. Am J Physiol Cell Physiol 318(6):C1189–C1199.

[20] Guay, C., Menoud, V., Rome, S., Regazzi, R., 2015. Horizontal transfer of exosomal microRNAs transduce apoptotic signals between pancreatic beta-cells. Cell Commun Signal 13:17.

[21] Freeman, D.W., Noren Hooten, N., Eitan, E., Green, J., Mode, N.A., Bodogai, M., et al., 2018. Altered Extracellular Vesicle Concentration, Cargo, and Function in Diabetes. Diabetes 67(11):2377–2388.

[22] Zhang, H., Liu, J., Qu, D., Wang, L., Wong, C.M., Lau, C.W., et al., 2018. Serum exosomes mediate delivery of arginase 1 as a novel mechanism for endothelial dysfunction in diabetes. Proc Natl Acad Sci U S A 115(29):E6927–E6936.

[23] Zeng, M., Wen, J., Ma, Z., Xiao, L., Liu, Y., Kwon, S., et al., 2021. FOXO1-Mediated Downregulation of RAB27B Leads to Decreased Exosome Secretion in Diabetic Kidneys. Diabetes 70(7):1536–1548.

[24] Nunez Lopez, Y.O., Garufi, G., Seyhan, A.A., 2016. Altered levels of circulating cytokines and microRNAs in lean and obese individuals with prediabetes and type 2 diabetes. Mol Biosyst 13(1):106–121.

[25] Grover, S.P., Mackman, N., 2018. Tissue Factor. Arteriosclerosis, Thrombosis, and Vascular Biology 38(4):709–725.

[26] Sliker, B.H., Goetz, B.T., Barnes, R., King, H., Maurer, H.C., Olive, K.P., et al., 2020. HLA-B influences integrin beta-1 expression and pancreatic cancer cell migration. Exp Cell Res 390(2):111960.

[27] Apostolopoulou, M., Mastrototaro, L., Hartwig, S., Pesta, D., Straßburger, K., Filippo, E.d., et al., 2021. Metabolic responsiveness to training depends on insulin sensitivity and protein content of exosomes in insulin-resistant males. Science Advances 7(41):eabi9551.

[28] Gonzales, P.A., Pisitkun, T., Hoffert, J.D., Tchapyjnikov, D., Star, R.A., Kleta, R., et al., 2009. Large-Scale Proteomics and Phosphoproteomics of Urinary Exosomes. Journal of the American Society of Nephrology 20(2):363–379.

[29] Sun, J., Han, S., Ma, L., Zhang, H., Zhan, Z., Aguilar, H.A., et al., 2021. Synergistically Bifunctional Paramagnetic Separation Enables Efficient Isolation of Urine Extracellular Vesicles and Downstream Phosphoproteomic Analysis. ACS Appl Mater Interfaces 13(3):3622–3630.

[30] Zheng, H., Guan, S., Wang, X., Zhao, J., Gao, M., Zhang, X., 2020. Deconstruction of Heterogeneity of Size-Dependent Exosome Subpopulations from Human Urine by Profiling N-Glycoproteomics and Phosphoproteomics Simultaneously. Anal Chem 92(13):9239–9246.

[31] James, D.E., Stöckli, J., Birnbaum, M.J., 2021. The aetiology and molecular landscape of insulin resistance. Nature Reviews Molecular Cell Biology 22(11):751–771.

[32] Yang, J.Y., Deng, W., Chen, Y., Fan, W., Baldwin, K.M., Jope, R.S., et al., 2013. Impaired translocation and activation of mitochondrial Akt1 mitigated mitochondrial oxidative phosphorylation Complex V activity in diabetic myocardium. J Mol Cell Cardiol 59:167–175.

[33] Yang, J.Y., Yeh, H.Y., Lin, K., Wang, P.H., 2009. Insulin stimulates Akt translocation to mitochondria: implications on dysregulation of mitochondrial oxidative phosphorylation in diabetic myocardium. J Mol Cell Cardiol 46(6):919–926.

[34] Hurrle, S., Hsu, W.H., 2017. The etiology of oxidative stress in insulin resistance. Biomedical journal 40(5):257–262.

[35] Kuzmenko, D.I., Udintsev, S.N., Klimentyeva, T.K., Serebrov, V.Y., 2016. Oxidative stress in adipose tissue as a primary link in pathogenesis of insulin resistance. Biochemistry (Moscow) Supplement Series B: Biomedical Chemistry 10(3):212–219.

[36] Olsson, A.H., Yang, B.T., Hall, E., Taneera, J., Salehi, A., Nitert, M.D., et al., 2011. Decreased expression of genes involved in oxidative phosphorylation in human pancreatic islets from patients with type 2 diabetes. Eur J Endocrinol 165(4):589–595.

[37] Wang, M., Wang, X.C., Zhang, Z.Y., Mou, B., Hu, R.M., 2010. Impaired mitochondrial oxidative phosphorylation in multiple insulin-sensitive tissues of humans with type 2 diabetes mellitus. J Int Med Res 38(3):769–781.

[38] Mootha, V.K., Handschin, C., Arlow, D., Xie, X., St Pierre, J., Sihag, S., et al., 2004. Erralpha and Gabpa/b specify PGC-1alpha-dependent oxidative phosphorylation gene expression that is altered in diabetic muscle. Proc Natl Acad Sci U S A 101(17):6570–6575.

[39] Mootha, V.K., Lindgren, C.M., Eriksson, K.F., Subramanian, A., Sihag, S., Lehar, J., et al., 2003. PGC-1alpha-responsive genes involved in oxidative phosphorylation are coordinately downregulated in human diabetes. Nat Genet 34(3):267–273.

[40] Ying, W., Tseng, A., Chang, R.C.-A., Wang, H., Lin, Y.-l., Kanameni, S., et al., 2016. miR-150 regulates obesity-associated insulin resistance by controlling B cell functions. Scientific reports 6(1):20176.

[41] Winer, D.A., Winer, S., Shen, L., Wadia, P.P., Yantha, J., Paltser, G., et al., 2011. B cells promote insulin resistance through modulation of T cells and production of pathogenic IgG antibodies. Nature Medicine 17(5):610–617.

[42] Winer, D.A., Winer, S., Shen, L., Chng, M.H.Y., Engleman, E.G., 2012. B lymphocytes as emerging mediators of insulin resistance. International Journal of Obesity Supplements 2(1):S4–S7.

[43] Foley, J.H., Conway, E.M., 2016. Cross Talk Pathways Between Coagulation and Inflammation. Circulation Research 118(9):1392–1408.

[44] Goldberg, R.B., 2009. Cytokine and cytokine-like inflammation markers, endothelial dysfunction, and imbalanced coagulation in development of diabetes and its complications. J Clin Endocrinol Metab 94(9):3171–3182.

[45] Aso, Y., Yoshida, N., Okumura, K., Wakabayashi, S., Matsutomo, R., Takebayashi, K., et al., 2004. Coagulation and inflammation in overt diabetic nephropathy: association with hyperhomocysteinemia. Clin Chim Acta 348(1-2):139–145.

[46] Lam, S.M., Zhang, C., Wang, Z., Ni, Z., Zhang, S., Yang, S., et al., 2021. A multi-omics investigation of the composition and function of extracellular vesicles along the temporal trajectory of COVID-19. Nat Metab 3(7):909–922.

[47] Barberis, E., Vanella, V.V., Falasca, M., Caneapero, V., Cappellano, G., Raineri, D., et al., 2021. Circulating Exosomes Are Strongly Involved in SARS-CoV-2 Infection. Front Mol Biosci 8:632290.

[48] Hoshino, A., Costa-Silva, B., Shen, T.-L., Rodrigues, G., Hashimoto, A., Tesic Mark, M., et al., 2015. Tumour exosome integrins determine organotropic metastasis. Nature 527(7578):329–335.

[49] Wortzel, I., Dror, S., Kenific, C.M., Lyden, D., 2019. Exosome-Mediated Metastasis: Communication from a Distance. Developmental Cell 49(3):347–360.

[50] Zong, H., Bastie, C.C., Xu, J., Fassler, R., Campbell, K.P., Kurland, I.J., et al., 2009. Insulin resistance in striated muscle-specific integrin receptor beta1-deficient mice. J Biol Chem 284(7):4679–4688.

[51] Kang, L., Ayala, J.E., Lee-Young, R.S., Zhang, Z., James, F.D., Neufer, P.D., et al., 2011. Diet-induced muscle insulin resistance is associated with extracellular matrix remodeling and interaction with integrin alpha2beta1 in mice. Diabetes 60(2):416–426.

[52] de Fougerolles, A.R., Sprague, A.G., Nickerson-Nutter, C.L., Chi-Rosso, G., Rennert, P.D., Gardner, H., et al., 2000. Regulation of inflammation by collagen-binding integrins alpha1beta1 and alpha2beta1 in models of hypersensitivity and arthritis. J Clin Invest 105(6):721–729.

[53] Zhang, Z., Ramirez, N.E., Yankeelov, T.E., Li, Z., Ford, L.E., Qi, Y., et al., 2008. alpha2beta1 integrin expression in the tumor microenvironment enhances tumor angiogenesis in a tumor cell-specific manner. Blood 111(4):1980–1988.

[54] Roest, M., Banga, J.D., Grobbee, D.E., de Groot, P.G., Sixma, J.J., Tempelman, M.J., et al., 2000. Homozygosity for 807 T polymorphism in alpha(2) subunit of platelet alpha(2)beta(1) is associated with increased risk of cardiovascular mortality in high-risk women. Circulation 102(14):1645–1650.

[55] Wertheimer, E., Taylor, S.I., Tennenbaum, T., 1998. Insulin receptor regulation of cell surface integrins: a possible mechanism contributing to the development of diabetic complications. Proc Assoc Am Physicians 110(4):333–339.

[56] Yu, L., Su, Y., Paueksakon, P., Cheng, H., Chen, X., Wang, H., et al., 2012. Integrin alpha1/Akita double-knockout mice on a Balb/c background develop advanced features of human diabetic nephropathy. Kidney Int 81(11):1086–1097.

[57] Roth, T., Podesta, F., Stepp, M.A., Boeri, D., Lorenzi, M., 1993. Integrin overexpression induced by high glucose and by human diabetes: potential pathway to cell dysfunction in diabetic microangiopathy. Proc Natl Acad Sci U S A 90(20):9640–9644.

[58] Lay, A.C., Hale, L.J., Stowell-Connolly, H., Pope, R.J.P., Nair, V., Ju, W., et al., 2021. IGFBP-1 expression is reduced in human type 2 diabetic glomeruli and modulates beta1-integrin/FAK signalling in human podocytes. Diabetologia 64(7):1690–1702.

[59] Kanasaki, K., Kanda, Y., Palmsten, K., Tanjore, H., Bong Lee, S., Lebleu, V.S., et al., 2008. Integrin β1-mediated matrix assembly and signaling are critical for the normal development and function of the kidney glomerulus. Developmental Biology 313(2):584–593.

[60] Yu, C.-C., Fornoni, A., Weins, A., Hakroush, S., Maiguel, D., Sageshima, J., et al., 2013. Abatacept in B7-1–Positive Proteinuric Kidney Disease. New England Journal of Medicine 369(25):2416–2423.

[61] Taylor, R., 2008. Pathogenesis of type 2 diabetes: tracing the reverse route from cure to cause. Diabetologia 51(10):1781–1789.

[62] Taylor, R., 2013. Banting Memorial lecture 2012: reversing the twin cycles of type 2 diabetes. Diabet Med 30(3):267–275.

[63] Gastaldelli, A., Cusi, K., 2019. From NASH to diabetes and from diabetes to NASH: Mechanisms and treatment options. JHEP Reports 1(4):312–328.

[64] Targher, G., Chonchol, M., Zoppini, G., Abaterusso, C., Bonora, E., 2011. Risk of chronic kidney disease in patients with non-alcoholic fatty liver disease: Is there a link? Journal of Hepatology 54(5):1020–1029.

[65] Alicic, R.Z., Rooney, M.T., Tuttle, K.R., 2017. Diabetic Kidney Disease: Challenges, Progress, and Possibilities. Clinical journal of the American Society of Nephrology : CJASN 12(12):2032–2045.

[66] Le, M.H., Yeo, Y.H., Henry, L., Nguyen, M.H., 2019. Nonalcoholic Fatty Liver Disease and Renal Function Impairment: A Cross-Sectional Population-Based Study on Its Relationship From 1999 to 2016. Hepatol Commun 3(10):1334–1346.

[67] Pais, R., Bourron, O., 2017. Fatty liver and renal function impairment -Time for awareness? J Hepatol.

[68] Association, A.D., 2014. Standards of Medical Care in Diabetes—2014. Diabetes Care 37(Supplement 1):S14–S80.

[69] Ho, D.E., Imai, K., King, G., Stuart, E.A., 2011. MatchIt: Nonparametric Preprocessing for Parametric Causal Inference. Journal of Statistical Software 42(8).

[70] Welch, S., Gebhart, S.S., Bergman, R.N., Phillips, L.S., 1990. Minimal model analysis of intravenous glucose tolerance test-derived insulin sensitivity in diabetic subjects. J Clin Endocrinol Metab 71(6):1508–1518.

[71] Pacini, G., Bergman, R.N., 1986. MINMOD: a computer program to calculate insulin sensitivity and pancreatic responsivity from the frequently sampled intravenous glucose tolerance test. Comput Methods Programs Biomed 23(2):113–122.

[72] Uhlén, M., Fagerberg, L., Hallström, B.M., Lindskog, C., Oksvold, P., Mardinoglu, A., et al., 2015. Tissue-based map of the human proteome. Science 347(6220):1260419.

[73] Pathan, M., Fonseka, P., Chitti, S.V., Kang, T., Sanwlani, R., Van Deun, J., et al., 2019. Vesiclepedia 2019: a compendium of RNA, proteins, lipids and metabolites in extracellular vesicles. Nucleic Acids Res 47(D1):D516–d519.

[74] Wu, T., Hu, E., Xu, S., Chen, M., Guo, P., Dai, Z., et al., 2021. clusterProfiler 4.0: A universal enrichment tool for interpreting omics data. Innovation (N Y) 2(3):100141.

[75] Smyth, G.K., 2004. Linear Models and Empirical Bayes Methods for Assessing Differential Expression in Microarray Experiments. Statistical Applications in Genetics and Molecular Biology 3(1).

[76] Schwämmle, V., León, I.R., Jensen, O.N., 2013. Assessment and Improvement of Statistical Tools for Comparative Proteomics Analysis of Sparse Data Sets with Few Experimental Replicates. Journal of Proteome Research 12(9):3874–3883.

[77] Ting, L., Cowley, M.J., Hoon, S.L., Guilhaus, M., Raftery, M.J., Cavicchioli, R., 2009. Normalization and Statistical Analysis of Quantitative Proteomics Data Generated by Metabolic Labeling*. Molecular & Cellular Proteomics 8(10):2227–2242.

[78] Margolin, A.A., Ong, S.-E., Schenone, M., Gould, R., Schreiber, S.L., Carr, S.A., et al., 2009. Empirical Bayes Analysis of Quantitative Proteomics Experiments. PLoS One 4(10):e7454.

[79] Ritchie, M.E., Phipson, B., Wu, D., Hu, Y.F., Law, C.W., Shi, W., et al., 2015. limma powers differential expression analyses for RNA-sequencing and microarray studies. Nucleic Acids Research 43(7).

[80] Kammers, K., Cole, R.N., Tiengwe, C., Ruczinski, I., 2015. Detecting Significant Changes in Protein Abundance. EuPA open proteomics 7:11–19.

