## Supplementary Tables for "Defining the Proteomic and Phosphoproteomic Landscape of Circulating Extracellular Vesicles in the Diabetes Spectrum"

**Supplementary Table ST1. Differentially expressed proteins in PDM vs NGT**

| **Accession** | **UNIPROT** | **SYMBOL** | **logFC** | **AveExpr** | **P.Value** | **adj.P.Val** |
| --- | --- | --- | --- | --- | --- | --- |
| Q5TZA2 | Q5TZA2 | CROCC | 11.38 | -12.97 | 6.69E-30 | 1.25E-26 |
| Q7L5N7 | Q7L5N7 | LPCAT2 | 9.76 | -13.99 | 1.63E-28 | 1.53E-25 |
| Q13336-2 | Q13336 | SLC14A1 | 4.95 | -5.21 | 2.56E-20 | 1.60E-17 |
| P09497-1 | P09497 | CLTB | 6.31 | -5.52 | 3.63E-20 | 1.70E-17 |
| Q96C24 | Q96C24 | SYTL4 | -2.32 | -7.22 | 2.09E-12 | 7.82E-10 |
| A0A087WSY6 | A0A087WSY6 | NA | -3.67 | -7.82 | 3.11E-11 | 9.69E-09 |
| Q99426 | Q99426 | TBCB | -12.06 | -11.05 | 8.21E-11 | 2.19E-08 |
| Q86WI1 | Q86WI1 | PKHD1L1 | -1.72 | -6.42 | 2.83E-10 | 6.61E-08 |
| Q7Z6K5 | Q7Z6K5 | ARPIN | -10.79 | -15.04 | 3.98E-10 | 8.27E-08 |
| Q92736 | Q92736 | RYR2 | 9.26 | -12.38 | 1.05E-09 | 1.96E-07 |
| Q9HCM2 | Q9HCM2 | PLXNA4 | 1.77 | -7.10 | 2.03E-09 | 3.28E-07 |
| Q8IZP7 | Q8IZP7 | HS6ST3 | -8.15 | -17.01 | 2.10E-09 | 3.28E-07 |
| Q15084-1 | Q15084 | PDIA6 | -1.91 | -6.41 | 2.71E-09 | 3.90E-07 |
| P21741 | P21741 | MDK | -10.21 | -15.00 | 7.28E-09 | 9.73E-07 |
| P21730 | P21730 | C5AR1 | 8.26 | -13.77 | 1.93E-08 | 2.40E-06 |
| Q8TB69 | Q8TB69 | ZNF519 | 7.82 | -17.88 | 4.93E-08 | 5.76E-06 |
| Q27J81-2 | Q27J81 | INF2 | -1.68 | -6.66 | 5.80E-08 | 6.39E-06 |
| P51665 | P51665 | PSMD7 | 7.57 | -15.04 | 1.77E-07 | 1.84E-05 |
| P08174-5 | P08174 | CD55 | 2.13 | -6.95 | 3.16E-07 | 3.11E-05 |
| Q16181 | Q16181 | SEPTIN7 | -1.18 | -5.88 | 3.78E-07 | 3.53E-05 |
| O75150 | O75150 | RNF40 | 7.45 | -7.61 | 4.16E-07 | 3.61E-05 |
| P13716-1 | P13716 | ALAD | 1.36 | -4.49 | 4.24E-07 | 3.61E-05 |
| Q96JM2 | Q96JM2 | ZNF462 | -9.31 | -14.79 | 4.67E-07 | 3.71E-05 |
| Q9NS84 | Q9NS84 | CHST7 | -4.60 | -17.84 | 4.76E-07 | 3.71E-05 |
| P98160 | P98160 | HSPG2 | -1.66 | -3.51 | 1.65E-06 | 1.23E-04 |
| Q15323 | Q15323 | KRT31 | 7.32 | -12.14 | 2.06E-06 | 1.48E-04 |
| P10644 | P10644 | PRKAR1A | -1.55 | -6.47 | 2.44E-06 | 1.69E-04 |
| P11586 | P11586 | MTHFD1 | 1.06 | -7.52 | 2.88E-06 | 0.000192747 |
| P19971 | P19971 | TYMP | -1.36 | -8.12 | 5.92E-06 | 0.000382047 |
| P80511 | P80511 | S100A12 | -6.57 | -14.68 | 8.45E-06 | 0.000501318 |
| O94910 | O94910 | ADGRL1 | 3.62 | -18.86 | 8.57E-06 | 0.000501318 |
| Q9UHN6-1 | Q9UHN6 | CEMIP2 | -6.74 | -18.18 | 1.01E-05 | 0.000574757 |
| Q4L235-1 | Q4L235 | AASDH | 6.27 | -15.68 | 1.47E-05 | 0.000806845 |
| P05362 | P05362 | ICAM1 | 2.02 | -8.07 | 1.63E-05 | 0.000872345 |
| Q99497 | Q99497 | PARK7 | 1.32 | -6.95 | 1.82E-05 | 0.000938601 |
| O75882-1 | O75882 | ATRN | 2.12 | -5.38 | 1.86E-05 | 0.000938601 |
| Q9NZN3 | Q9NZN3 | EHD3 | -1.09 | -5.06 | 2.22E-05 | 0.001093358 |
| Q9ULV0 | Q9ULV0 | MYO5B | -6.80 | -17.50 | 2.35E-05 | 0.001116017 |
| P48681 | P48681 | NES | -8.00 | -16.45 | 2.39E-05 | 0.001116017 |
| Q6NXE6-1 | Q6NXE6 | ARMC6 | 3.75 | -18.31 | 2.84E-05 | 0.001296681 |
| Q96H15-1 | Q96H15 | TIMD4 | 6.10 | -15.91 | 3.60E-05 | 0.001579284 |
| NA | NA | NA | -8.32 | -14.90 | 3.63E-05 | 0.001579284 |
| Q9UJJ9 | Q9UJJ9 | GNPTG | 6.07 | -18.03 | 3.74E-05 | 0.001579853 |
| NA | NA | NA | 5.29 | -15.73 | 3.80E-05 | 0.001579853 |
| Q15691 | Q15691 | MAPRE1 | 1.38 | -8.35 | 4.04E-05 | 0.001642003 |
| Q9UQ80 | Q9UQ80 | PA2G4 | -1.35 | -9.86 | 5.07E-05 | 0.002019816 |
| P13073 | P13073 | COX4I1 | 6.54 | -12.96 | 6.02E-05 | 0.002299207 |
| P17010 | P17010 | ZFX | 7.90 | -16.77 | 6.27E-05 | 0.002345338 |
| P19838 | P19838 | NFKB1 | -6.78 | -17.79 | 7.35E-05 | 0.002698115 |
| Q12860 | Q12860 | CNTN1 | 8.57 | -14.10 | 8.12E-05 | 0.002867015 |
| Q13586 | Q13586 | STIM1 | -5.87 | -12.64 | 8.34E-05 | 0.002888564 |
| Q9UHQ9 | Q9UHQ9 | CYB5R1 | -5.10 | -18.64 | 1.08E-04 | 0.003594854 |
| Q6ZN44-1 | Q6ZN44 | UNC5A | -7.62 | -17.90 | 1.22E-04 | 0.003978636 |
| Q8IY82 | Q8IY82 | DRC7 | -4.82 | -18.74 | 1.27E-04 | 0.003978636 |
| P29401-2 | P29401 | TKT | 1.44 | -8.12 | 1.28E-04 | 0.003978636 |
| O60506 | O60506 | SYNCRIP | -5.18 | -18.70 | 1.30E-04 | 0.003978636 |
| P08397-1 | P08397 | HMBS | -5.31 | -16.37 | 1.68E-04 | 0.004972462 |
| O60229-6 | O60229 | KALRN | -6.21 | -18.37 | 1.75E-04 | 0.004972462 |
| O95602 | O95602 | POLR1A | 5.00 | -18.72 | 1.76E-04 | 0.004972462 |
| Q96RD7-1 | Q96RD7 | PANX1 | 6.75 | -16.63 | 1.77E-04 | 0.004972462 |
| P35232 | P35232 | PHB | -3.50 | -10.18 | 1.77E-04 | 0.004972462 |
| P17252 | P17252 | PRKCA | -2.83 | -7.55 | 1.93E-04 | 0.005229795 |
| A0A1B0GVM6 | A0A1B0GVM6 | C11orf97 | -7.99 | -13.41 | 2.08E-04 | 0.005485982 |
| Q9Y4A5 | Q9Y4A5 | TRRAP | 5.92 | -17.80 | 2.30E-04 | 0.005896222 |
| Q13315 | Q13315 | ATM | -1.56 | -8.07 | 2.47E-04 | 0.006233311 |
| P41218 | P41218 | MNDA | -7.48 | -15.31 | 2.64E-04 | 0.006567202 |
| P28799 | P28799 | GRN | 6.08 | -11.68 | 2.67E-04 | 0.006567202 |
| P14923 | P14923 | JUP | -2.54 | -5.51 | 2.77E-04 | 0.006733082 |
| P09105 | P09105 | HBQ1 | 5.95 | -16.84 | 3.12E-04 | 0.007431086 |
| P31641 | P31641 | SLC6A6 | 5.93 | -17.36 | 0.000325064 | 0.007602432 |
| Q9UK55 | Q9UK55 | SERPINA10 | -1.08 | -6.11 | 0.00033545 | 0.007710667 |
| Q9HC10 | Q9HC10 | OTOF | 1.02 | -9.21 | 0.000341167 | 0.007710667 |
| P22692 | P22692 | IGFBP4 | -5.56 | -18.05 | 0.000342245 | 0.007710667 |
| O00410 | O00410 | IPO5 | 5.12 | -10.58 | 0.000346176 | 0.007710667 |
| Q13867 | Q13867 | BLMH | 1.28 | -7.41 | 0.000354446 | 0.00780199 |
| P01920 | P01920 | HLA-DQB1 | 4.96 | -18.31 | 0.000363846 | 0.007915766 |
| O14618 | O14618 | CCS | -5.62 | -12.78 | 0.000376855 | 0.008104538 |
| Q5T749 | Q5T749 | KPRP | -1.52 | -6.58 | 0.000382981 | 0.008142691 |
| Q7L7X3 | Q7L7X3 | TAOK1 | -4.65 | -17.68 | 0.000444418 | 0.009133074 |
| O60884 | O60884 | DNAJA2 | -6.10 | -15.04 | 0.000449088 | 0.009133074 |
| P49257 | P49257 | LMAN1 | 4.64 | -18.31 | 0.000473998 | 0.009536018 |
| P49736 | P49736 | MCM2 | -6.03 | -17.69 | 0.000515686 | 0.010211543 |
| O43504 | O43504 | LAMTOR5 | -6.84 | -14.97 | 0.000518491 | 0.010211543 |
| P56645 | P56645 | PER3 | -6.74 | -17.35 | 0.000550521 | 0.010729432 |
| P32942 | P32942 | ICAM3 | -1.22 | -8.86 | 0.000558889 | 0.010780221 |
| Q5JPH6 | Q5JPH6 | EARS2 | 5.18 | -15.35 | 0.000604906 | 0.011317785 |
| Q00688 | Q00688 | FKBP3 | 5.23 | -18.04 | 0.000632942 | 0.011725089 |
| Q8TEP8-3 | Q8TEP8 | CEP192 | -6.02 | -17.63 | 0.000776249 | 0.013588669 |
| P26572 | P26572 | MGAT1 | 5.64 | -15.07 | 0.000777118 | 0.013588669 |
| P50416 | P50416 | CPT1A | 5.49 | -17.23 | 0.000793704 | 0.013750194 |
| O75915 | O75915 | ARL6IP5 | 5.61 | -13.56 | 0.000853911 | 0.014524249 |
| P00488 | P00488 | F13A1 | -1.37 | -2.17 | 0.000872322 | 0.014631247 |
| Q9NRV9 | Q9NRV9 | HEBP1 | 1.35 | -8.28 | 0.000905056 | 0.014848643 |
| P40197 | P40197 | GP5 | -1.65 | -8.68 | 0.000929736 | 0.014925669 |
| Q9UDX4 | Q9UDX4 | SEC14L3 | -6.04 | -17.60 | 0.000939385 | 0.014925669 |
| P49327 | P49327 | FASN | 1.18 | -7.63 | 0.00094133 | 0.014925669 |
| P01137 | P01137 | TGFB1 | -1.62 | -7.17 | 0.001048251 | 0.016201211 |
| P12111-2 | P12111 | COL6A3 | -1.23 | -5.13 | 0.001056412 | 0.016201211 |
| O15144 | O15144 | ARPC2 | -1.47 | -4.75 | 0.001094236 | 0.016429263 |
| P22695 | P22695 | UQCRC2 | -8.06 | -16.07 | 0.001097034 | 0.016429263 |
| P11049-1 | P11049 | CD37 | -2.43 | -8.83 | 0.001097626 | 0.016429263 |
| P98082 | P98082 | DAB2 | -4.33 | -16.70 | 0.001120783 | 0.016511694 |
| P01877 | P01877 | NA | -5.79 | -17.59 | 0.001188058 | 0.017366064 |
| P27449 | P27449 | ATP6V0C | 4.88 | -13.07 | 0.001200333 | 0.017409474 |
| O00161 | O00161 | SNAP23 | -1.39 | -7.73 | 0.001253675 | 0.017984391 |
| Q53GL0-1 | Q53GL0 | PLEKHO1 | -3.81 | -18.85 | 0.001259196 | 0.017984391 |
| Q9BWD1 | Q9BWD1 | ACAT2 | 5.24 | -11.38 | 0.001274741 | 0.018068484 |
| O14745 | O14745 | SLC9A3R1 | 1.37 | -7.65 | 0.001299567 | 0.018217601 |
| Q16610 | Q16610 | ECM1 | -1.99 | -2.54 | 0.001304735 | 0.018217601 |
| O75094-1 | O75094 | SLIT3 | 5.19 | -13.07 | 0.001364136 | 0.018766904 |
| Q7LDG7 | Q7LDG7 | RASGRP2 | -1.27 | -7.22 | 0.001485882 | 0.020145548 |
| Q8NBQ5 | Q8NBQ5 | HSD17B11 | -5.68 | -17.92 | 0.001528465 | 0.020573802 |
| P21796 | P21796 | VDAC1 | 4.02 | -10.65 | 0.001547744 | 0.020684497 |
| P04080 | P04080 | CSTB | -1.31 | -8.45 | 0.001607831 | 0.021335113 |
| O00743 | O00743 | PPP6C | -6.42 | -17.13 | 0.001638632 | 0.021541591 |
| P09960-1 | P09960 | LTA4H | -2.09 | -8.07 | 0.001646418 | 0.021541591 |
| P30512 | P30512 | NA | 4.59 | -18.18 | 0.001662638 | 0.021602746 |
| Q9NX76 | Q9NX76 | CMTM6 | 5.80 | -12.81 | 0.001712178 | 0.022092997 |
| Q96SJ8 | Q96SJ8 | TSPAN18 | -5.61 | -16.72 | 0.001731038 | 0.022125363 |
| P02671-1 | P02671 | FGA | -1.76 | 0.38 | 0.001738337 | 0.022125363 |
| Q13043-1 | Q13043 | STK4 | -1.75 | -9.91 | 0.001805656 | 0.022687787 |
| P36969 | P36969 | GPX4 | -2.02 | -9.72 | 0.001806777 | 0.022687787 |
| P40189-1 | P40189 | IL6ST | -4.60 | -17.52 | 0.00191104 | 0.023679179 |
| Q14573 | Q14573 | ITPR3 | -5.72 | -17.02 | 0.001941604 | 0.023899614 |
| Q15185 | Q15185 | PTGES3 | -4.48 | -11.53 | 0.002059427 | 0.024859271 |
| Q9NYL9 | Q9NYL9 | TMOD3 | 5.30 | -14.41 | 0.00207878 | 0.02493203 |
| P51812 | P51812 | RPS6KA3 | -4.77 | -17.81 | 0.002148107 | 0.025599414 |
| P0CG38 | P0CG38 | POTEI | 4.85 | -18.28 | 0.00240085 | 0.027933739 |
| Q5D862 | Q5D862 | FLG2 | -2.39 | -5.86 | 0.002476015 | 0.028247708 |
| Q5VW32 | Q5VW32 | BROX | 5.30 | -17.26 | 0.002781336 | 0.030445507 |
| A0A075B6K5 | A0A075B6K5 | NA | -1.35 | -4.67 | 0.002787516 | 0.030445507 |
| P62140 | P62140 | PPP1CB | 3.02 | -9.57 | 0.002798839 | 0.030445507 |
| Q92619 | Q92619 | ARHGAP45 | -1.51 | -6.76 | 0.002832623 | 0.030634904 |
| Q8IVF2-1 | Q8IVF2 | AHNAK2 | -5.86 | -16.71 | 0.002918854 | 0.031206719 |
| Q86UP2-1 | Q86UP2 | KTN1 | 4.12 | -17.92 | 0.003005484 | 0.031769835 |
| Q07021 | Q07021 | C1QBP | 4.92 | -14.38 | 0.003024828 | 0.031794683 |
| P48637 | P48637 | GSS | -4.46 | -11.97 | 0.003098553 | 0.032242969 |
| P15907 | P15907 | ST6GAL1 | 3.78 | -17.47 | 0.003116596 | 0.032242969 |
| P23368-1 | P23368 | ME2 | -5.55 | -14.64 | 0.003156419 | 0.032275404 |
| P35030 | P35030 | PRSS3 | -5.07 | -18.10 | 0.003156814 | 0.032275404 |
| O95716 | O95716 | RAB3D | -5.48 | -16.01 | 0.003204827 | 0.032412057 |
| Q92558 | Q92558 | WASF1 | 3.14 | -18.20 | 0.003245237 | 0.032625111 |
| Q9HBH0 | Q9HBH0 | RHOF | 1.13 | -8.57 | 0.003260767 | 0.032625111 |
| P43405-1 | P43405 | SYK | 2.73 | -9.99 | 0.003418018 | 0.033836573 |
| Q8IY47 | Q8IY47 | KBTBD2 | -4.62 | -18.14 | 0.003473889 | 0.034029562 |
| Q15911-1 | Q15911 | ZFHX3 | 6.00 | -16.48 | 0.003512677 | 0.034208972 |
| P49758-3 | P49758 | RGS6 | 4.24 | -13.23 | 0.003528772 | 0.034208972 |
| O43488 | O43488 | AKR7A2 | 2.74 | -7.61 | 0.003557978 | 0.034314316 |
| Q6S545-1 | Q6S545 | POTEH | -5.45 | -17.33 | 0.003588003 | 0.034426427 |
| P08493-1 | P08493 | MGP | -4.58 | -11.44 | 0.003624127 | 0.034595624 |
| Q96H79 | Q96H79 | ZC3HAV1L | -4.58 | -18.20 | 0.003787873 | 0.035613624 |
| O75964 | O75964 | ATP5MG | 4.37 | -14.12 | 0.003908399 | 0.036563071 |
| P02675 | P02675 | FGB | -1.63 | 0.53 | 0.00398276 | 0.036728872 |
| Q92686 | Q92686 | NRGN | 4.84 | -17.25 | 0.003985014 | 0.036728872 |
| Q12907 | Q12907 | LMAN2 | 3.26 | -11.00 | 0.004010157 | 0.036779433 |
| P02679 | P02679 | FGG | -1.76 | 0.76 | 0.004037319 | 0.036847926 |
| P14317 | P14317 | HCLS1 | -3.92 | -11.29 | 0.004074228 | 0.03692946 |
| O95477 | O95477 | ABCA1 | -1.51 | -9.77 | 0.004096764 | 0.03692946 |
| Q5SNT2-1 | Q5SNT2 | TMEM201 | 3.90 | -18.16 | 0.00413455 | 0.037013122 |
| A0A0A0MRZ9 | A0A0A0MRZ9 | NA | -4.66 | -18.23 | 0.004342425 | 0.03836575 |
| Q5TCS8 | Q5TCS8 | AK9 | 5.08 | -13.88 | 0.004347161 | 0.03836575 |
| P10606 | P10606 | COX5B | 4.01 | -11.76 | 0.004484202 | 0.038842326 |
| O76074 | O76074 | PDE5A | -1.49 | -4.74 | 0.004554889 | 0.039145539 |
| O60749 | O60749 | SNX2 | -4.78 | -17.51 | 0.004561052 | 0.039145539 |
| P30273 | P30273 | FCER1G | 4.36 | -9.65 | 0.004598691 | 0.039215643 |
| Q5THJ4 | Q5THJ4 | VPS13D | 5.14 | -16.71 | 0.004623967 | 0.039215643 |
| Q9Y6C2 | Q9Y6C2 | EMILIN1 | -1.52 | -8.27 | 0.00466547 | 0.039215643 |
| P98095 | P98095 | FBLN2 | -4.90 | -11.57 | 0.004748755 | 0.039488539 |
| Q93100-1 | Q93100 | PHKB | 7.08 | -16.20 | 0.004780775 | 0.039578896 |
| O75348 | O75348 | ATP6V1G1 | -5.92 | -16.58 | 0.004869173 | 0.039957117 |
| Q9C0F0 | Q9C0F0 | ASXL3 | -5.14 | -17.41 | 0.004932899 | 0.040147246 |
| Q8WZA0-2 | Q8WZA0 | LZIC | -2.79 | -7.90 | 0.004935257 | 0.040147246 |
| Q6UXG3 | Q6UXG3 | CD300LG | -4.85 | -16.92 | 0.005024433 | 0.040695734 |
| Q8IW75 | Q8IW75 | SERPINA12 | 5.79 | -16.23 | 0.005110775 | 0.040864361 |
| O75122-3 | O75122 | CLASP2 | -5.34 | -17.26 | 0.005201921 | 0.04124065 |
| Q96LP6 | Q96LP6 | C12orf42 | -5.38 | -17.27 | 0.005252161 | 0.041418685 |
| Q9HB71 | Q9HB71 | CACYBP | -5.27 | -15.87 | 0.005268652 | 0.041418685 |
| NA | NA | NA | -3.75 | -18.45 | 0.00535799 | 0.041484702 |
| P15169 | P15169 | CPN1 | -1.08 | -7.87 | 0.005379113 | 0.041484702 |
| Q96A08 | Q96A08 | H2BC1 | -5.62 | -17.09 | 0.005391161 | 0.041484702 |
| P15529-2 | P15529 | CD46 | 3.02 | -11.20 | 0.005427267 | 0.041484702 |
| P60903 | P60903 | S100A10 | -4.58 | -15.73 | 0.005427671 | 0.041484702 |
| P28065 | P28065 | PSMB9 | 3.74 | -16.06 | 0.005431121 | 0.041484702 |
| P36871-1 | P36871 | PGM1 | 1.10 | -7.07 | 0.005478131 | 0.041496289 |
| P12821-1 | P12821 | ACE | 5.45 | -11.64 | 0.005511687 | 0.041582124 |
| Q9BX46-2 | Q9BX46 | RBM24 | 4.71 | -16.54 | 0.005629654 | 0.042132334 |
| P24593 | P24593 | IGFBP5 | -5.20 | -17.45 | 0.005692045 | 0.042429545 |
| O00237 | O00237 | RNF103 | -4.03 | -18.35 | 0.005889328 | 0.043725925 |
| P42356 | P42356 | PI4KA | 4.49 | -14.12 | 0.005943326 | 0.043952425 |
| O94823-1 | O94823 | ATP10B | 5.86 | -13.00 | 0.006189067 | 0.045233377 |
| Q96FJ2 | Q96FJ2 | DYNLL2 | -1.43 | -8.10 | 0.006286435 | 0.045766224 |
| Q14699 | Q14699 | RFTN1 | -5.44 | -17.48 | 0.006629276 | 0.047760385 |
| O14617-1 | O14617 | AP3D1 | -4.73 | -11.13 | 0.006662459 | 0.047760385 |
| Q9BPZ7 | Q9BPZ7 | MAPKAP1 | -4.52 | -17.55 | 0.00677612 | 0.048216439 |
| Q13094 | Q13094 | LCP2 | 4.64 | -14.42 | 0.006777618 | 0.048216439 |
| A0A075B6J9 | A0A075B6J9 | NA | -4.82 | -16.75 | 0.006804842 | 0.04822674 |

**Supplementary Table ST2. Differentially expressed proteins in T2DM vs NGT.**

| **Accession** | **UNIPROT** | **SYMBOL** | **logFC** | **AveExpr** | **P.Value** | **adj.P.Val** |
| --- | --- | --- | --- | --- | --- | --- |
| Q5TZA2 | Q5TZA2 | CROCC | 11.02 | -12.97 | 2.32E-29 | 4.34E-26 |
| Q7L5N7 | Q7L5N7 | LPCAT2 | 9.52 | -13.99 | 4.72E-28 | 4.42E-25 |
| P09497-1 | P09497 | CLTB | 5.97 | -5.52 | 2.12E-19 | 1.32E-16 |
| Q96C24 | Q96C24 | SYTL4 | -2.15 | -7.22 | 1.56E-11 | 7.31E-09 |
| Q15084-1 | Q15084 | PDIA6 | -2.37 | -6.41 | 4.98E-11 | 1.86E-08 |
| A0A087WSY6 | A0A087WSY6 | NA | -3.39 | -7.82 | 2.30E-10 | 7.18E-08 |
| Q86WI1 | Q86WI1 | PKHD1L1 | -1.76 | -6.42 | 2.69E-10 | 7.20E-08 |
| Q99426 | Q99426 | TBCB | -11.38 | -11.05 | 3.84E-10 | 8.97E-08 |
| Q27J81-2 | Q27J81 | INF2 | -2.18 | -6.66 | 6.06E-10 | 1.23E-07 |
| Q9HCM2 | Q9HCM2 | PLXNA4 | 1.90 | -7.10 | 6.60E-10 | 1.23E-07 |
| P30046 | P30046 | DDT | 5.12 | -8.47 | 7.78E-10 | 1.32E-07 |
| Q92736 | Q92736 | RYR2 | 9.49 | -12.38 | 9.05E-10 | 1.41E-07 |
| P21730 | P21730 | C5AR1 | 9.74 | -13.77 | 1.16E-09 | 1.66E-07 |
| P21741 | P21741 | MDK | -11.29 | -15.00 | 1.49E-09 | 2.00E-07 |
| Q9NS84 | Q9NS84 | CHST7 | -6.25 | -17.84 | 2.64E-09 | 3.30E-07 |
| Q7Z6K5 | Q7Z6K5 | ARPIN | -9.26 | -15.04 | 1.08E-08 | 1.19E-06 |
| P13716-1 | P13716 | ALAD | 1.63 | -4.49 | 2.36E-08 | 2.45E-06 |
| Q5JPH6 | Q5JPH6 | EARS2 | 10.69 | -15.35 | 2.52E-08 | 2.48E-06 |
| P05362 | P05362 | ICAM1 | 2.86 | -8.07 | 8.93E-08 | 8.35E-06 |
| Q4L235-1 | Q4L235 | AASDH | 8.48 | -15.68 | 1.81E-07 | 1.61E-05 |
| Q15691 | Q15691 | MAPRE1 | 1.99 | -8.35 | 2.13E-07 | 1.81E-05 |
| P08397-1 | P08397 | HMBS | -8.57 | -16.37 | 2.35E-07 | 1.91E-05 |
| O75150 | O75150 | RNF40 | 7.50 | -7.61 | 5.01E-07 | 3.63E-05 |
| P08174-5 | P08174 | CD55 | 2.10 | -6.95 | 5.05E-07 | 3.63E-05 |
| Q96JM2 | Q96JM2 | ZNF462 | -9.39 | -14.79 | 5.44E-07 | 3.74E-05 |
| Q8IZP7 | Q8IZP7 | HS6ST3 | -6.08 | -17.01 | 5.60E-07 | 3.74E-05 |
| P09493-8 | P09493 | TPM1 | 1.08 | -6.54 | 1.54E-06 | 9.92E-05 |
| P19971 | P19971 | TYMP | -1.51 | -8.12 | 1.63E-06 | 0.000101808 |
| Q9ULV0 | Q9ULV0 | MYO5B | -8.18 | -17.50 | 2.12E-06 | 0.000127919 |
| Q15323 | Q15323 | KRT31 | 7.23 | -12.14 | 3.30E-06 | 0.000192674 |
| Q9NZN3 | Q9NZN3 | EHD3 | -1.25 | -5.06 | 3.94E-06 | 0.000223389 |
| Q86YW5-2 | Q86YW5 | TREML1 | 7.82 | -16.23 | 4.10E-06 | 0.000225771 |
| Q96H15-1 | Q96H15 | TIMD4 | 7.24 | -15.91 | 4.23E-06 | 0.000225928 |
| Q6P3W7 | Q6P3W7 | SCYL2 | 6.80 | -16.96 | 5.01E-06 | 0.000260279 |
| Q9UHN6-1 | Q9UHN6 | CEMIP2 | -6.92 | -18.18 | 9.02E-06 | 0.000455945 |
| P09382 | P09382 | LGALS1 | 1.24 | -7.73 | 1.04E-05 | 0.000509743 |
| P28065 | P28065 | PSMB9 | 6.85 | -16.06 | 1.10E-05 | 0.000517597 |
| Q99497 | Q99497 | PARK7 | 1.39 | -6.95 | 1.11E-05 | 0.000517597 |
| Q7L7X3 | Q7L7X3 | TAOK1 | -6.38 | -17.68 | 1.16E-05 | 0.000517597 |
| P10644 | P10644 | PRKAR1A | -1.42 | -6.47 | 1.16E-05 | 0.000517597 |
| P40197 | P40197 | GP5 | -2.42 | -8.68 | 1.27E-05 | 0.000550797 |
| P51665 | P51665 | PSMD7 | 5.85 | -15.04 | 1.30E-05 | 0.000550797 |
| P42768 | P42768 | WAS | -8.41 | -13.85 | 1.35E-05 | 0.000550797 |
| P17252 | P17252 | PRKCA | -3.55 | -7.55 | 1.35E-05 | 0.000550797 |
| P13073 | P13073 | COX4I1 | 7.37 | -12.96 | 1.58E-05 | 0.000629584 |
| P32942 | P32942 | ICAM3 | -1.63 | -8.86 | 2.28E-05 | 0.000852987 |
| P09960-1 | P09960 | LTA4H | -3.10 | -8.07 | 2.62E-05 | 0.000961468 |
| O75915 | O75915 | ARL6IP5 | 7.72 | -13.56 | 2.70E-05 | 0.000969767 |
| P11049-1 | P11049 | CD37 | -3.42 | -8.83 | 2.84E-05 | 0.000986537 |
| Q30KQ8 | Q30KQ8 | NA | -8.67 | -15.98 | 2.85E-05 | 0.000986537 |
| P35613 | P35613 | BSG | 1.22 | -6.78 | 3.25E-05 | 0.001105593 |
| P29401-2 | P29401 | TKT | 1.64 | -8.12 | 3.32E-05 | 0.001108316 |
| Q99439 | Q99439 | CNN2 | 1.01 | -7.82 | 3.64E-05 | 0.001194247 |
| O75964 | O75964 | ATP5MG | 6.95 | -14.12 | 3.97E-05 | 0.001282201 |
| Q16610 | Q16610 | ECM1 | -2.74 | -2.54 | 4.91E-05 | 0.001506339 |
| P48506 | P48506 | GCLC | 7.00 | -13.71 | 5.09E-05 | 0.001536684 |
| O15394 | O15394 | NCAM2 | 8.68 | -16.61 | 5.19E-05 | 0.001541023 |
| P16402 | P16402 | H1-3 | -2.37 | -8.60 | 5.30E-05 | 0.00154871 |
| Q9Y6Z7 | Q9Y6Z7 | COLEC10 | -2.04 | -8.51 | 5.53E-05 | 0.001590556 |
| P41226 | P41226 | UBA7 | -1.11 | -7.76 | 5.86E-05 | 0.001644727 |
| Q4KWH8-2 | Q4KWH8 | PLCH1 | -1.66 | -6.13 | 5.95E-05 | 0.001644727 |
| P19013 | P19013 | NA | -2.08 | -7.92 | 6.18E-05 | 0.001675475 |
| O95716 | O95716 | RAB3D | -8.19 | -16.01 | 6.41E-05 | 0.001713036 |
| Q15833-1 | Q15833 | STXBP2 | 1.03 | -5.71 | 7.63E-05 | 0.001985829 |
| Q3ZCW2 | Q3ZCW2 | LGALSL | 1.72 | -8.86 | 7.70E-05 | 0.001985829 |
| P21589-1 | P21589 | NT5E | -7.42 | -12.29 | 9.42E-05 | 0.002345978 |
| Q6ZN44-1 | Q6ZN44 | UNC5A | -7.91 | -17.90 | 9.59E-05 | 0.002345978 |
| P20701-2 | P20701 | ITGAL | -1.93 | -7.30 | 9.65E-05 | 0.002345978 |
| O60506 | O60506 | SYNCRIP | -5.39 | -18.70 | 9.84E-05 | 0.002360481 |
| Q15942 | Q15942 | ZYX | 1.50 | -6.29 | 0.000101009 | 0.002385587 |
| A0A075B6K5 | A0A075B6K5 | NA | -1.90 | -4.67 | 0.000102003 | 0.002385587 |
| P49736 | P49736 | MCM2 | -7.08 | -17.69 | 0.000107332 | 0.002469148 |
| Q96HC4-4 | Q96HC4 | PDLIM5 | -1.29 | -8.69 | 0.000108215 | 0.002469148 |
| P56645 | P56645 | PER3 | -7.92 | -17.35 | 0.000113201 | 0.002551802 |
| P00488 | P00488 | F13A1 | -1.68 | -2.17 | 0.000118945 | 0.002649352 |
| P04085 | P04085 | PDGFA | 8.06 | -16.25 | 0.000121629 | 0.002677262 |
| Q9Y251 | Q9Y251 | HPSE | -2.47 | -8.33 | 0.000127861 | 0.002781722 |
| Q03164 | Q03164 | KMT2A | -1.03 | -5.77 | 0.000129355 | 0.00278188 |
| Q9BTE3-1 | Q9BTE3 | MCMBP | 5.76 | -17.63 | 0.000131517 | 0.002796231 |
| O00571 | O00571 | DDX3X | -10.09 | -14.84 | 0.000140688 | 0.002904716 |
| Q9UHQ9 | Q9UHQ9 | CYB5R1 | -5.06 | -18.64 | 0.000145085 | 0.002904716 |
| P62140 | P62140 | PPP1CB | 4.14 | -9.57 | 0.000145268 | 0.002904716 |
| O60229-6 | O60229 | KALRN | -6.40 | -18.37 | 0.000149635 | 0.002904716 |
| Q8TEP8-3 | Q8TEP8 | CEP192 | -7.14 | -17.63 | 0.000152081 | 0.002904716 |
| O76074 | O76074 | PDE5A | -2.17 | -4.74 | 0.000152144 | 0.002904716 |
| Q9UDX4 | Q9UDX4 | SEC14L3 | -7.23 | -17.60 | 0.000169662 | 0.003142955 |
| P48681 | P48681 | NES | -6.93 | -16.45 | 0.000175059 | 0.003211124 |
| O75882-1 | O75882 | ATRN | 1.80 | -5.38 | 0.00018233 | 0.003302738 |
| P42356 | P42356 | PI4KA | 6.67 | -14.12 | 0.000183584 | 0.003302738 |
| O75094-1 | O75094 | SLIT3 | 6.35 | -13.07 | 0.000211528 | 0.003733673 |
| Q96JM3 | Q96JM3 | CHAMP1 | -8.45 | -16.74 | 0.000225712 | 0.003946796 |
| P14923 | P14923 | JUP | -2.63 | -5.51 | 0.000232852 | 0.004033939 |
| Q6S545-1 | Q6S545 | POTEH | -7.39 | -17.33 | 0.00023902 | 0.004077471 |
| Q8IY82 | Q8IY82 | DRC7 | -4.63 | -18.74 | 0.000247121 | 0.004129684 |
| O43504 | O43504 | LAMTOR5 | -7.46 | -14.97 | 0.000247207 | 0.004129684 |
| Q9Y336 | Q9Y336 | SIGLEC9 | 4.37 | -18.91 | 0.000251127 | 0.004158038 |
| Q9UK55 | Q9UK55 | SERPINA10 | -1.13 | -6.11 | 0.000254899 | 0.004168846 |
| Q8TCD6 | Q8TCD6 | PHOSPHO2 | -9.64 | -14.94 | 0.000256236 | 0.004168846 |
| P02751 | P02751 | FN1 | -2.56 | 2.19 | 0.000272433 | 0.0043566 |
| O75368 | O75368 | SH3BGRL | 6.41 | -11.79 | 0.000281871 | 0.004449229 |
| P13797 | P13797 | PLS3 | -6.76 | -11.73 | 0.000282981 | 0.004449229 |
| P61758 | P61758 | VBP1 | -5.53 | -17.50 | 0.000285726 | 0.004454949 |
| Q9BWD1 | Q9BWD1 | ACAT2 | 6.18 | -11.38 | 0.000291289 | 0.004504148 |
| Q9H0B8 | Q9H0B8 | CRISPLD2 | -1.54 | -8.24 | 0.00029474 | 0.004520144 |
| Q9C0F0 | Q9C0F0 | ASXL3 | -7.10 | -17.41 | 0.000304395 | 0.004630265 |
| O75122-3 | O75122 | CLASP2 | -7.42 | -17.26 | 0.000310714 | 0.004688276 |
| Q08830 | Q08830 | FGL1 | -1.24 | -6.06 | 0.000314539 | 0.004708015 |
| Q96LP6 | Q96LP6 | C12orf42 | -7.47 | -17.27 | 0.00031951 | 0.004724132 |
| Q96A08 | Q96A08 | H2BC1 | -7.83 | -17.09 | 0.000320665 | 0.004724132 |
| Q8TCG5-2 | Q8TCG5 | CPT1C | -5.60 | -15.20 | 0.00032714 | 0.004771538 |
| P23229-6 | P23229 | ITGA6 | 1.13 | -4.51 | 0.000341419 | 0.004913803 |
| O75396 | O75396 | SEC22B | 5.80 | -15.44 | 0.000345461 | 0.004927191 |
| P02671-1 | P02671 | FGA | -2.11 | 0.38 | 0.000356778 | 0.004944971 |
| Q53GL0-1 | Q53GL0 | PLEKHO1 | -4.40 | -18.85 | 0.000358332 | 0.004944971 |
| Q01518-1 | Q01518 | CAP1 | 1.03 | -4.36 | 0.000366468 | 0.005004825 |
| P51003 | P51003 | PAPOLA | -7.84 | -16.69 | 0.000373065 | 0.005058011 |
| Q6F5E8 | Q6F5E8 | CARMIL2 | -8.16 | -16.67 | 0.000380085 | 0.005080221 |
| O14745 | O14745 | SLC9A3R1 | 1.58 | -7.65 | 0.000380134 | 0.005080221 |
| O15511-1 | O15511 | ARPC5 | 1.14 | -6.38 | 0.000383109 | 0.005083664 |
| P01877 | P01877 | NA | -6.60 | -17.59 | 0.00038635 | 0.005090566 |
| P02675 | P02675 | FGB | -2.13 | 0.53 | 0.000401231 | 0.005249667 |
| NA | NA | NA | -1.02 | -8.26 | 0.000417677 | 0.005426907 |
| Q6PIU2 | Q6PIU2 | NCEH1 | -7.76 | -16.73 | 0.000422453 | 0.005451096 |
| Q9NRV9 | Q9NRV9 | HEBP1 | 1.48 | -8.28 | 0.000450033 | 0.005767198 |
| P19838 | P19838 | NFKB1 | -5.87 | -17.79 | 0.000465292 | 0.005922183 |
| P46109 | P46109 | CRKL | -1.02 | -8.01 | 0.00047302 | 0.005979869 |
| P27449 | P27449 | ATP6V0C | 5.44 | -13.07 | 0.00049195 | 0.006121267 |
| P02679 | P02679 | FGG | -2.27 | 0.76 | 0.000493868 | 0.006121267 |
| A0A0C4DH67 | A0A0C4DH67 | NA | 1.50 | -6.90 | 0.000496655 | 0.006121267 |
| Q9BUL8 | Q9BUL8 | PDCD10 | 1.00 | -8.54 | 0.000500804 | 0.006124214 |
| P19320-1 | P19320 | VCAM1 | -6.04 | -17.18 | 0.00051503 | 0.006251179 |
| Q75N90 | Q75N90 | FBN3 | -7.52 | -16.93 | 0.000517869 | 0.006251179 |
| P98082 | P98082 | DAB2 | -4.76 | -16.70 | 0.000526289 | 0.006312089 |
| P20813 | P20813 | CYP2B6 | -6.77 | -13.90 | 0.000533224 | 0.006354538 |
| O00555-1 | O00555 | CACNA1A | -8.25 | -16.60 | 0.00054128 | 0.006409711 |
| P05534 | P05534 | NA | -1.23 | -4.99 | 0.000576793 | 0.006779928 |
| Q9NRY5 | Q9NRY5 | FAM114A2 | 6.62 | -17.32 | 0.000579791 | 0.006779928 |
| P07911 | P07911 | UMOD | -7.84 | -15.83 | 0.000583992 | 0.006784998 |
| O60884 | O60884 | DNAJA2 | -6.03 | -15.04 | 0.00060207 | 0.006874352 |
| P31749 | P31749 | AKT1 | -9.21 | -15.45 | 0.000602562 | 0.006874352 |
| P62136-1 | P62136 | PPP1CA | 1.59 | -9.48 | 0.00061769 | 0.006961312 |
| P04632 | P04632 | CAPNS1 | 1.00 | -7.11 | 0.000621346 | 0.006961312 |
| P43405-1 | P43405 | SYK | 3.36 | -9.99 | 0.000625511 | 0.006966257 |
| P61960 | P61960 | UFM1 | 5.55 | -16.45 | 0.000638561 | 0.007066982 |
| A0A0A0MRZ9 | A0A0A0MRZ9 | NA | -5.90 | -18.23 | 0.00064211 | 0.007066982 |
| Q9Y2L9-1 | Q9Y2L9 | LRCH1 | -8.05 | -16.34 | 0.000694192 | 0.007465939 |
| Q8NI99 | Q8NI99 | ANGPTL6 | -2.07 | -10.02 | 0.00069432 | 0.007465939 |
| Q96H79 | Q96H79 | ZC3HAV1L | -5.63 | -18.20 | 0.000715342 | 0.007648024 |
| A0A075B6J9 | A0A075B6J9 | NA | -6.40 | -16.75 | 0.000725187 | 0.007709228 |
| Q8WZA0-2 | Q8WZA0 | LZIC | -3.54 | -7.90 | 0.000735356 | 0.007773165 |
| P21796 | P21796 | VDAC1 | 4.41 | -10.65 | 0.000772933 | 0.008065183 |
| Q8N335 | Q8N335 | GPD1L | 4.33 | -18.77 | 0.000775913 | 0.008065183 |
| P09668 | P09668 | CTSH | -6.48 | -16.04 | 0.000797276 | 0.008241458 |
| P15907 | P15907 | ST6GAL1 | 4.48 | -17.47 | 0.000804286 | 0.008268234 |
| Q99102-16 | Q99102 | MUC4 | -8.56 | -15.80 | 0.000817536 | 0.008313098 |
| P15529-2 | P15529 | CD46 | 3.83 | -11.20 | 0.000838317 | 0.008414623 |
| Q9GZX5 | Q9GZX5 | ZNF350 | 6.45 | -17.30 | 0.000849913 | 0.008458442 |
| Q8IY47 | Q8IY47 | KBTBD2 | -5.51 | -18.14 | 0.000858592 | 0.008493147 |
| P49327 | P49327 | FASN | 1.21 | -7.63 | 0.000867018 | 0.008493147 |
| Q8NBF2-1 | Q8NBF2 | NHLRC2 | -5.43 | -10.46 | 0.000887727 | 0.008650714 |
| P06132 | P06132 | UROD | 6.46 | -15.45 | 0.000923205 | 0.008885765 |
| P61952 | P61952 | GNG11 | 1.16 | -9.49 | 0.000930844 | 0.008885765 |
| NA | NA | NA | -4.68 | -18.45 | 0.000956137 | 0.009058439 |
| Q13315 | Q13315 | ATM | -1.38 | -8.07 | 0.001003341 | 0.009386256 |
| Q9H061-1 | Q9H061 | TMEM126A | -6.54 | -15.35 | 0.001029945 | 0.009508249 |
| P30626-1 | P30626 | SRI | 1.24 | -6.89 | 0.001036359 | 0.009508249 |
| Q02318 | Q02318 | CYP27A1 | 7.56 | -11.68 | 0.001038535 | 0.009508249 |
| P23276 | P23276 | KEL | -2.62 | -5.97 | 0.001038962 | 0.009508249 |
| O15144 | O15144 | ARPC2 | -1.50 | -4.75 | 0.001083123 | 0.009816886 |
| P36871-1 | P36871 | PGM1 | 1.36 | -7.07 | 0.001092507 | 0.009816886 |
| P35030 | P35030 | PRSS3 | -5.83 | -18.10 | 0.001100214 | 0.009816886 |
| Q9NX76 | Q9NX76 | CMTM6 | 6.19 | -12.81 | 0.001106385 | 0.009816886 |
| Q92530 | Q92530 | PSMF1 | 5.90 | -12.72 | 0.001107089 | 0.009816886 |
| Q9UNF0 | Q9UNF0 | PACSIN2 | -1.43 | -8.09 | 0.001136612 | 0.00999715 |
| Q9Y4D8-1 | Q9Y4D8 | NA | 3.01 | -19.00 | 0.001138104 | 0.00999715 |
| Q96BY6-1 | Q96BY6 | DOCK10 | -1.57 | -6.43 | 0.001219006 | 0.010657762 |
| P15169 | P15169 | CPN1 | -1.32 | -7.87 | 0.001234033 | 0.010738955 |
| O00237 | O00237 | RNF103 | -4.96 | -18.35 | 0.001243804 | 0.010773874 |
| P69905 | P69905 | HBA1 | 1.02 | 2.18 | 0.001260384 | 0.010817334 |
| Q13094 | Q13094 | LCP2 | 5.80 | -14.42 | 0.00128226 | 0.010954833 |
| O00743 | O00743 | PPP6C | -6.65 | -17.13 | 0.001383569 | 0.011538234 |
| Q13586 | Q13586 | STIM1 | -4.58 | -12.64 | 0.001387548 | 0.011538234 |
| Q9BQ90 | Q9BQ90 | KLHDC3 | -8.08 | -15.93 | 0.001450911 | 0.011906381 |
| P68133 | P68133 | ACTA1 | 1.04 | -3.42 | 0.001549877 | 0.012607911 |
| P04275 | P04275 | VWF | -1.06 | -2.83 | 0.001558437 | 0.012622661 |
| P31415 | P31415 | CASQ1 | 5.56 | -12.79 | 0.001582013 | 0.012758393 |
| Q5D862 | Q5D862 | FLG2 | -2.54 | -5.86 | 0.001686938 | 0.013430897 |
| P00915 | P00915 | CA1 | 1.10 | -3.26 | 0.001737461 | 0.013774532 |
| O60268-1 | O60268 | KIAA0513 | 4.90 | -12.04 | 0.001772724 | 0.013994794 |
| P56192 | P56192 | MARS1 | -6.27 | -16.47 | 0.001800711 | 0.014142514 |
| Q12860 | Q12860 | CNTN1 | 6.48 | -14.10 | 0.001806553 | 0.014142514 |
| P22695 | P22695 | UQCRC2 | -7.74 | -16.07 | 0.00185325 | 0.014387679 |
| Q7RTS7 | Q7RTS7 | KRT74 | -8.15 | -14.48 | 0.001940269 | 0.014939271 |
| Q9ULH1-2 | Q9ULH1 | ASAP1 | -3.62 | -10.53 | 0.001991185 | 0.015166468 |
| P10606 | P10606 | COX5B | 4.51 | -11.76 | 0.001999602 | 0.015166468 |
| Q13043-1 | Q13043 | STK4 | -1.76 | -9.91 | 0.002002201 | 0.015166468 |
| P11836 | P11836 | MS4A1 | -2.87 | -10.47 | 0.002020677 | 0.015244702 |
| Q6ZMW3 | Q6ZMW3 | EML6 | 4.66 | -13.53 | 0.00203176 | 0.015266759 |
| P43681 | P43681 | CHRNA4 | -8.73 | -15.81 | 0.002055203 | 0.015381136 |
| Q15828 | Q15828 | CST6 | 5.60 | -15.80 | 0.002099418 | 0.0155202 |
| P07359 | P07359 | GP1BA | 1.05 | -5.44 | 0.002106965 | 0.0155202 |
| P51812 | P51812 | RPS6KA3 | -4.85 | -17.81 | 0.00214744 | 0.01569477 |
| Q96AG4 | Q96AG4 | LRRC59 | 4.47 | -18.06 | 0.00217402 | 0.015765859 |
| Q562R1 | Q562R1 | ACTBL2 | 1.16 | -2.23 | 0.002190992 | 0.015827588 |
| Q658P3-1 | Q658P3 | STEAP3 | -1.56 | -9.81 | 0.002217409 | 0.015956819 |
| P30273 | P30273 | FCER1G | 4.84 | -9.65 | 0.00229908 | 0.016413844 |
| Q8NCA5 | Q8NCA5 | FAM98A | -4.09 | -18.79 | 0.002303934 | 0.016413844 |
| O00560-1 | O00560 | SDCBP | -5.21 | -18.29 | 0.002314094 | 0.016413844 |
| Q9H8W4 | Q9H8W4 | PLEKHF2 | 4.96 | -17.50 | 0.00231601 | 0.016413844 |
| P78415 | P78415 | IRX3 | -6.69 | -16.75 | 0.002336626 | 0.016497461 |
| Q8NFW1 | Q8NFW1 | COL22A1 | -7.27 | -14.93 | 0.002362188 | 0.016566429 |
| P09972 | P09972 | ALDOC | 1.10 | -8.37 | 0.002364103 | 0.016566429 |
| O00501 | O00501 | CLDN5 | 5.11 | -12.08 | 0.002384739 | 0.016586794 |
| O00410 | O00410 | IPO5 | 4.26 | -10.58 | 0.002402585 | 0.01658759 |
| P63261 | P63261 | ACTG1 | 1.15 | 0.27 | 0.002412448 | 0.016594447 |
| Q6ZVT6-1 | Q6ZVT6 | CFAP20DC | 4.71 | -15.41 | 0.002503419 | 0.017141273 |
| Q92619 | Q92619 | ARHGAP45 | -1.56 | -6.76 | 0.002510267 | 0.017141273 |
| Q96LB3-1 | Q96LB3 | IFT74 | -7.04 | -15.88 | 0.002541407 | 0.017290811 |
| Q9Y6C2 | Q9Y6C2 | EMILIN1 | -1.66 | -8.27 | 0.002596856 | 0.017540496 |
| Q96FZ7 | Q96FZ7 | CHMP6 | 6.33 | -14.40 | 0.002622418 | 0.017638667 |
| P38646 | P38646 | HSPA9 | -4.88 | -9.22 | 0.002630245 | 0.017638667 |
| O94823-1 | O94823 | ATP10B | 6.64 | -13.00 | 0.002734781 | 0.018032101 |
| O43681 | O43681 | GET3 | 5.82 | -16.17 | 0.002737101 | 0.018032101 |
| Q3T906 | Q3T906 | GNPTAB | -5.22 | -14.34 | 0.002756185 | 0.018094111 |
| O95865 | O95865 | DDAH2 | 5.70 | -14.95 | 0.002920208 | 0.018775631 |
| O15400-2 | O15400 | STX7 | -1.80 | -7.62 | 0.002951665 | 0.018912895 |
| Q9HBI6 | Q9HBI6 | CYP4F11 | 5.51 | -17.10 | 0.003007552 | 0.01920522 |
| P55145 | P55145 | MANF | 5.63 | -17.45 | 0.00302751 | 0.019266909 |
| Q13459-1 | Q13459 | MYO9B | 6.27 | -16.04 | 0.003149225 | 0.019971521 |
| P18859 | P18859 | ATP5PF | 5.31 | -17.69 | 0.003170252 | 0.019971521 |
| Q8TC12 | Q8TC12 | RDH11 | 5.16 | -12.71 | 0.003192309 | 0.020042988 |
| P67936 | P67936 | TPM4 | 1.10 | -5.58 | 0.003262657 | 0.020416159 |
| P14384 | P14384 | CPM | -5.45 | -17.80 | 0.003287256 | 0.020433407 |
| Q4FZB7-1 | Q4FZB7 | KMT5B | -5.47 | -16.19 | 0.003323496 | 0.020590271 |
| Q9Y2I6-1 | Q9Y2I6 | NINL | 4.76 | -15.38 | 0.003551901 | 0.021788877 |
| P22531 | P22531 | SPRR2E | 4.75 | -12.95 | 0.003575608 | 0.021791407 |
| Q6IAA8 | Q6IAA8 | LAMTOR1 | 4.90 | -13.00 | 0.00361721 | 0.021939268 |
| P14406 | P14406 | COX7A2 | 4.51 | -15.84 | 0.003631282 | 0.021939268 |
| A0A1B0GVM6 | A0A1B0GVM6 | C11orf97 | -6.02 | -13.41 | 0.003642344 | 0.021939268 |
| P67775 | P67775 | PPP2CA | 4.95 | -14.62 | 0.00378554 | 0.022628577 |
| Q6ULP2 | Q6ULP2 | AFTPH | 4.83 | -17.27 | 0.003832457 | 0.022836076 |
| Q9NVA2-1 | Q9NVA2 | SEPTIN11 | 1.08 | -8.66 | 0.003905247 | 0.023195928 |
| P40763-2 | P40763 | STAT3 | -3.87 | -11.82 | 0.003936365 | 0.023306767 |
| Q58EX7 | Q58EX7 | PLEKHG4 | 5.83 | -13.93 | 0.004097608 | 0.024033307 |
| O15265-2 | O15265 | ATXN7 | 5.89 | -12.23 | 0.004208478 | 0.024606447 |
| Q96N67 | Q96N67 | DOCK7 | 5.55 | -14.98 | 0.004269419 | 0.024668768 |
| P22692 | P22692 | IGFBP4 | -4.30 | -18.05 | 0.004270127 | 0.024668768 |
| P17813 | P17813 | ENG | 3.12 | -10.91 | 0.004271876 | 0.024668768 |
| Q9P126 | Q9P126 | CLEC1B | 4.11 | -11.36 | 0.004335314 | 0.024958067 |
| P07203 | P07203 | GPX1 | -1.39 | -5.63 | 0.004372363 | 0.024966846 |
| P01701 | P01701 | NA | -2.82 | -3.32 | 0.004373229 | 0.024966846 |
| P22303-1 | P22303 | ACHE | -1.67 | -8.25 | 0.004376871 | 0.024966846 |
| Q6NXE6-1 | Q6NXE6 | ARMC6 | 2.34 | -18.31 | 0.004419767 | 0.025132174 |
| O15173 | O15173 | PGRMC2 | 5.20 | -16.04 | 0.004432719 | 0.025132174 |
| P13861 | P13861 | PRKAR2A | 4.36 | -17.94 | 0.004511539 | 0.025424969 |
| P40925 | P40925 | MDH1 | 1.59 | -7.95 | 0.004592827 | 0.025584775 |
| Q9BXR6 | Q9BXR6 | CFHR5 | -1.11 | -4.82 | 0.004594594 | 0.025584775 |
| P09105 | P09105 | HBQ1 | 4.49 | -16.84 | 0.004797185 | 0.026554829 |
| Q9BWP8-9 | Q9BWP8 | COLEC11 | -1.18 | -8.63 | 0.004911641 | 0.027059548 |
| Q9Y613 | Q9Y613 | FHOD1 | -1.27 | -4.23 | 0.004917288 | 0.027059548 |
| Q9NRW1 | Q9NRW1 | RAB6B | 1.11 | -7.94 | 0.004962703 | 0.027229376 |
| Q8NBL1 | Q8NBL1 | POGLUT1 | -5.36 | -17.51 | 0.005224749 | 0.028266266 |
| NA | NA | NA | -5.17 | -14.90 | 0.00522722 | 0.028266266 |
| O15264 | O15264 | MAPK13 | -5.62 | -17.48 | 0.005251426 | 0.028315325 |
| P35232 | P35232 | PHB | -2.47 | -10.18 | 0.005387504 | 0.028718008 |
| Q8TE77 | Q8TE77 | SSH3 | -6.65 | -14.86 | 0.005410921 | 0.02875212 |
| Q15911-1 | Q15911 | ZFHX3 | 5.76 | -16.48 | 0.005429359 | 0.02875212 |
| Q13423 | Q13423 | NNT | 5.87 | -15.26 | 0.005440006 | 0.02875212 |
| Q14C86-3 | Q14C86 | GAPVD1 | -1.01 | -7.93 | 0.005487098 | 0.028838088 |
| Q5XPI4 | Q5XPI4 | RNF123 | 3.28 | -10.62 | 0.005574545 | 0.029134003 |
| P06703 | P06703 | S100A6 | 1.09 | -7.33 | 0.005609334 | 0.029234159 |
| Q5T749 | Q5T749 | KPRP | -1.13 | -6.58 | 0.006139736 | 0.031369287 |
| Q8N1Q1 | Q8N1Q1 | CA13 | -6.47 | -14.89 | 0.006151998 | 0.031369287 |
| P78527 | P78527 | PRKDC | -2.24 | -8.70 | 0.006213015 | 0.031502849 |
| O95866-7 | O95866 | MPIG6B | -1.96 | -6.91 | 0.006339537 | 0.031885142 |
| P0DOX4 | P0DOX4 | NA | 4.32 | -12.50 | 0.006632236 | 0.033178914 |
| Q9UKE5-4 | Q9UKE5 | TNIK | -1.74 | -8.88 | 0.006710353 | 0.033480187 |
| A0A0C4DH32 | A0A0C4DH32 | NA | -5.25 | -16.06 | 0.006730222 | 0.033490013 |
| Q14699 | Q14699 | RFTN1 | -5.51 | -17.48 | 0.006847063 | 0.033981047 |
| Q96FJ2 | Q96FJ2 | DYNLL2 | -1.44 | -8.10 | 0.006912275 | 0.034213935 |
| Q15404-1 | Q15404 | RSU1 | 1.03 | -4.10 | 0.007029727 | 0.034612155 |
| P51452 | P51452 | DUSP3 | 1.11 | -8.14 | 0.007578443 | 0.036799422 |
| P13693 | P13693 | TPT1 | 4.61 | -12.02 | 0.008222484 | 0.039230043 |
| P01903 | P01903 | HLA-DRA | -1.52 | -8.25 | 0.008266332 | 0.039248797 |
| P27348 | P27348 | YWHAQ | 1.30 | -7.30 | 0.008288002 | 0.039248797 |
| Q9Y2Y8 | Q9Y2Y8 | PRG3 | 5.09 | -13.23 | 0.008328028 | 0.039248797 |
| Q5VTH9-1 | Q5VTH9 | DNAI4 | 5.21 | -16.17 | 0.008347754 | 0.039248797 |
| P07951-2 | P07951 | TPM2 | -3.45 | -11.86 | 0.008391463 | 0.039349444 |
| P20963-1 | P20963 | CD247 | -3.20 | -11.58 | 0.008528448 | 0.039792333 |
| Q17RC7 | Q17RC7 | EXOC3L4 | 4.64 | -13.74 | 0.008628853 | 0.040061002 |
| Q9Y2Q0-2 | Q9Y2Q0 | ATP8A1 | 3.82 | -11.66 | 0.008662641 | 0.040118319 |
| A0A0C4DH31 | A0A0C4DH31 | NA | -1.18 | -9.00 | 0.008832961 | 0.040758442 |
| Q15366-1 | Q15366 | PCBP2 | 4.93 | -14.80 | 0.008844429 | 0.040758442 |
| Q9HB71 | Q9HB71 | CACYBP | -4.95 | -15.87 | 0.009269922 | 0.042302497 |
| O94903 | O94903 | PLPBP | -5.04 | -17.58 | 0.009398022 | 0.042678884 |
| P68032 | P68032 | ACTC1 | 5.62 | -13.77 | 0.009928597 | 0.044159031 |
| P12821-1 | P12821 | ACE | 5.09 | -11.64 | 0.009973087 | 0.044159031 |
| O43790 | O43790 | KRT86 | 1.09 | -7.00 | 0.010007178 | 0.044159031 |
| P0C091 | P0C091 | FREM3 | 3.06 | -12.27 | 0.010497855 | 0.045998798 |
| Q5VW32 | Q5VW32 | BROX | 4.49 | -17.26 | 0.010745672 | 0.046886049 |
| Q6XQN6-1 | Q6XQN6 | NAPRT | 4.80 | -12.51 | 0.010750462 | 0.046886049 |
| P24903 | P24903 | CYP2F1 | 4.96 | -13.56 | 0.010844608 | 0.046968199 |
| Q15208 | Q15208 | STK38 | 4.23 | -17.60 | 0.010890928 | 0.047009805 |
| P53990-1 | P53990 | IST1 | 5.34 | -12.69 | 0.01093015 | 0.047012208 |
| Q12907 | Q12907 | LMAN2 | 2.88 | -11.00 | 0.011074249 | 0.047414006 |
| Q8NDL9 | Q8NDL9 | AGBL5 | -7.58 | -14.35 | 0.011144431 | 0.047497108 |
| Q92882 | Q92882 | OSTF1 | 4.52 | -13.69 | 0.011190426 | 0.047584745 |
| O14618 | O14618 | CCS | -3.78 | -12.78 | 0.011954305 | 0.049925236 |

**Supplementary Table ST3. Differentially expressed phosphoproteins in PDM vs. NGT**

| **Accession** | **UNIPROT** | **SYMBOL** | **logFC** | **AveExpr** | **P.Value** | **adj.P.Val** |
| --- | --- | --- | --- | --- | --- | --- |
| P14672 | P14672 | SLC2A4 | 8.48 | -10.37 | 2.65E-16 | 1.37E-13 |
| P02647 | P02647 | APOA1 | 3.50 | 1.79 | 8.42E-12 | 1.45E-09 |
| Q5T1N1-1 | Q5T1N1 | AKNAD1 | -7.38 | -14.95 | 8.45E-12 | 1.45E-09 |
| P08514-1 | P08514 | ITGA2B | -6.14 | -0.36 | 2.19E-11 | 2.83E-09 |
| Q5M775-1 | Q5M775 | SPECC1 | -10.48 | -8.81 | 6.33E-09 | 5.88E-07 |
| Q5JSH3 | Q5JSH3 | WDR44 | 3.25 | -3.54 | 6.84E-09 | 5.88E-07 |
| P02533 | P02533 | KRT14 | -10.12 | -12.10 | 1.92E-08 | 1.41E-06 |
| P12109 | P12109 | COL6A1 | -10.41 | -11.27 | 4.81E-07 | 3.10E-05 |
| Q9H7E9-2 | Q9H7E9 | C8orf33 | -8.25 | -11.76 | 5.41E-07 | 3.10E-05 |
| Q15404-1 | Q15404 | RSU1 | 5.73 | -11.05 | 3.27E-06 | 1.69E-04 |
| Q96Q42-1 | Q96Q42 | ALS2 | -7.15 | -12.31 | 4.46E-06 | 1.92E-04 |
| P07948-1 | P07948 | LYN | 2.07 | -3.62 | 4.47E-06 | 1.92E-04 |
| Q14247-1 | Q14247 | CTTN | 1.55 | -6.49 | 1.53E-05 | 6.07E-04 |
| P68363 | P68363 | TUBA1B | -2.33 | -6.12 | 1.77E-05 | 6.52E-04 |
| Q16610 | Q16610 | ECM1 | -2.94 | -6.43 | 4.13E-05 | 1.42E-03 |
| P05556-1 | P05556 | ITGB1 | -6.98 | -14.07 | 8.64E-05 | 2.79E-03 |
| P02545-3 | P02545 | LMNA | 5.57 | -10.46 | 1.41E-04 | 4.29E-03 |
| P20340-1 | P20340 | RAB6A | -4.96 | -14.53 | 1.77E-04 | 4.84E-03 |
| Q9ULI3 | Q9ULI3 | HEG1 | -5.76 | -13.47 | 1.78E-04 | 4.84E-03 |
| P69905 | P69905 | HBA1 | 1.37 | -2.68 | 2.30E-04 | 5.94E-03 |
| Q92625 | Q92625 | ANKS1A | 3.73 | -11.03 | 3.22E-04 | 7.61E-03 |
| Q9BXF9 | Q9BXF9 | TEKT3 | -4.36 | -15.52 | 3.24E-04 | 7.61E-03 |
| Q01518-1 | Q01518 | CAP1 | 1.72 | -6.37 | 4.26E-04 | 9.57E-03 |
| Q8NEZ4 | Q8NEZ4 | KMT2C | 6.71 | -12.37 | 5.98E-04 | 1.28E-02 |
| Q96RI0 | Q96RI0 | F2RL3 | 4.29 | -9.40 | 6.38E-04 | 1.32E-02 |
| Q8N163-1 | Q8N163 | CCAR2 | -5.00 | -15.11 | 7.09E-04 | 1.40E-02 |
| Q9Y5S2 | Q9Y5S2 | CDC42BPB | 7.08 | -10.29 | 7.33E-04 | 1.40E-02 |
| P55198 | P55198 | MLLT6 | -4.09 | -14.43 | 9.18E-04 | 1.69E-02 |
| O14639-1 | O14639 | ABLIM1 | 3.89 | -10.57 | 9.76E-04 | 1.74E-02 |
| Q13099-3 | Q13099 | IFT88 | -6.06 | -14.28 | 1.04E-03 | 1.79E-02 |
| P13645 | P13645 | KRT10 | -2.01 | -2.68 | 1.11E-03 | 1.85E-02 |
| Q0JRZ9-1 | Q0JRZ9 | FCHO2 | 4.89 | -13.42 | 1.26E-03 | 2.03E-02 |
| A2A3K4-1 | A2A3K4 | PTPDC1 | -3.65 | -8.14 | 1.36E-03 | 2.08E-02 |
| P04075 | P04075 | ALDOA | 1.09 | -4.72 | 1.37E-03 | 2.08E-02 |
| P15924-1 | P15924 | DSP | -4.12 | -14.41 | 1.57E-03 | 2.32E-02 |
| Q5TB80 | Q5TB80 | CEP162 | 4.65 | -14.06 | 2.29E-03 | 3.23E-02 |
| P02679 | P02679 | FGG | -1.57 | -1.32 | 2.37E-03 | 3.23E-02 |
| Q8IZF2 | Q8IZF2 | ADGRF5 | 4.31 | -14.76 | 2.38E-03 | 3.23E-02 |
| P60174-1 | P60174 | TPI1 | -4.28 | -15.30 | 2.58E-03 | 3.38E-02 |
| P16452-1 | P16452 | EPB42 | -1.11 | -5.26 | 2.78E-03 | 3.38E-02 |
| Q8TB72-1 | Q8TB72 | PUM2 | 3.08 | -12.63 | 2.82E-03 | 3.38E-02 |
| P02751-9 | P02751 | FN1 | -1.42 | 1.26 | 2.85E-03 | 3.38E-02 |
| Q01082-1 | Q01082 | SPTBN1 | 3.89 | -12.56 | 2.85E-03 | 3.38E-02 |
| Q7Z3Y8 | Q7Z3Y8 | KRT27 | -3.90 | -14.76 | 2.88E-03 | 3.38E-02 |
| Q8TEH3 | Q8TEH3 | DENND1A | 4.30 | -13.54 | 3.23E-03 | 3.70E-02 |
| P62328 | P62328 | TMSB4X | -4.72 | -9.63 | 3.41E-03 | 3.75E-02 |
| Q8WWA1 | Q8WWA1 | TMEM40 | -3.31 | -8.41 | 3.41E-03 | 3.75E-02 |
| P46063 | P46063 | RECQL | 4.33 | -13.26 | 3.57E-03 | 3.84E-02 |
| Q9BQE3 | Q9BQE3 | TUBA1C | 4.36 | -9.96 | 4.18E-03 | 4.40E-02 |
| O00187-1 | O00187 | MASP2 | -1.04 | -6.07 | 4.35E-03 | 4.48E-02 |
| P19878 | P19878 | NCF2 | -4.00 | -14.05 | 4.43E-03 | 4.48E-02 |
| Q08AD1 | Q08AD1 | CAMSAP2 | -4.82 | -14.48 | 4.66E-03 | 4.58E-02 |
| P46821 | P46821 | MAP1B | -4.18 | -11.88 | 4.70E-03 | 4.58E-02 |

**Supplementary Table ST4. Differentially expressed phosphoproteins in T2DM vs. NGT**

| **Accession** | **UNIPROT** | **SYMBOL** | **logFC** | **AveExpr** | **P.Value** | **adj.P.Val** |
| --- | --- | --- | --- | --- | --- | --- |
| P14672 | P14672 | SLC2A4 | 8.02 | -10.37 | 1.53E-15 | 7.90E-13 |
| Q5T1N1-1 | Q5T1N1 | AKNAD1 | -7.91 | -14.95 | 2.74E-12 | 7.07E-10 |
| Q5JSH3 | Q5JSH3 | WDR44 | 4.43 | -3.54 | 1.74E-11 | 2.99E-09 |
| P02647 | P02647 | APOA1 | 3.37 | 1.79 | 2.79E-11 | 3.60E-09 |
| Q5CZC0-1 | Q5CZC0 | FSIP2 | -10.47 | -7.86 | 9.15E-11 | 8.24E-09 |
| P08514-1 | P08514 | ITGA2B | -5.84 | -0.36 | 9.58E-11 | 8.24E-09 |
| Q9H7E9-2 | Q9H7E9 | C8orf33 | -12.62 | -11.76 | 2.59E-10 | 1.91E-08 |
| P02533 | P02533 | KRT14 | -12.66 | -12.10 | 3.35E-10 | 2.16E-08 |
| P07948-1 | P07948 | LYN | 3.03 | -3.62 | 8.70E-09 | 4.99E-07 |
| O00161 | O00161 | SNAP23 | -6.11 | -10.66 | 4.42E-08 | 2.28E-06 |
| Q01082-1 | Q01082 | SPTBN1 | 8.37 | -12.56 | 2.25E-07 | 1.06E-05 |
| P68363 | P68363 | TUBA1B | -3.02 | -6.12 | 4.23E-07 | 1.82E-05 |
| Q15404-1 | Q15404 | RSU1 | 6.45 | -11.05 | 6.56E-07 | 2.61E-05 |
| P55198 | P55198 | MLLT6 | -7.16 | -14.43 | 8.34E-07 | 3.07E-05 |
| P20340-1 | P20340 | RAB6A | -7.22 | -14.53 | 1.31E-06 | 4.50E-05 |
| P62070-4 | P62070 | RRAS2 | -6.40 | -10.93 | 1.44E-06 | 4.66E-05 |
| P67936 | P67936 | TPM4 | -4.24 | -9.79 | 1.98E-06 | 6.02E-05 |
| P02545-3 | P02545 | LMNA | 7.63 | -10.46 | 2.53E-06 | 7.25E-05 |
| P02675 | P02675 | FGB | -3.77 | -1.67 | 3.49E-06 | 9.47E-05 |
| Q14247-1 | Q14247 | CTTN | 1.70 | -6.49 | 5.31E-06 | 1.37E-04 |
| P02748 | P02748 | C9 | -1.90 | -6.53 | 6.65E-06 | 1.60E-04 |
| Q15942 | Q15942 | ZYX | 1.49 | -6.19 | 6.81E-06 | 1.60E-04 |
| Q96FS4 | Q96FS4 | SIPA1 | -7.63 | -10.78 | 7.58E-06 | 1.70E-04 |
| P01031 | P01031 | C5 | -6.27 | -9.94 | 8.46E-06 | 1.82E-04 |
| P06702 | P06702 | S100A9 | 1.62 | -1.69 | 1.27E-05 | 2.54E-04 |
| Q7RTP6-5 | Q7RTP6 | MICAL3 | -1.94 | -6.47 | 1.31E-05 | 2.54E-04 |
| Q5M775-1 | Q5M775 | SPECC1 | -6.75 | -8.81 | 1.33E-05 | 2.54E-04 |
| Q01433-1 | Q01433 | AMPD2 | 7.10 | -13.74 | 1.49E-05 | 0.000268663 |
| Q2M2I8-1 | Q2M2I8 | AAK1 | 5.94 | -11.84 | 1.51E-05 | 0.000268663 |
| Q16512-1 | Q16512 | PKN1 | -5.76 | -10.31 | 1.65E-05 | 0.00028459 |
| P68871 | P68871 | HBB | -1.35 | -0.33 | 1.74E-05 | 0.000290029 |
| Q96RI0 | Q96RI0 | F2RL3 | 5.77 | -9.40 | 2.39E-05 | 0.000384778 |
| Q08495-1 | Q08495 | DMTN | 1.51 | -0.24 | 2.46E-05 | 0.000385374 |
| P98082 | P98082 | DAB2 | 4.09 | -15.30 | 2.61E-05 | 0.000388994 |
| P46821 | P46821 | MAP1B | -7.01 | -11.88 | 2.64E-05 | 0.000388994 |
| Q16610 | Q16610 | ECM1 | -3.09 | -6.43 | 2.75E-05 | 0.000393917 |
| P04275 | P04275 | VWF | -4.77 | -8.07 | 2.85E-05 | 0.000397904 |
| P29692-2 | P29692 | EEF1D | -1.92 | -6.73 | 2.96E-05 | 0.000402004 |
| Q92625 | Q92625 | ANKS1A | 4.62 | -11.03 | 3.06E-05 | 0.000404723 |
| Q8NE71-1 | Q8NE71 | ABCF1 | -1.31 | -6.37 | 3.20E-05 | 0.000412333 |
| P60709 | P60709 | ACTB | -1.28 | -0.96 | 4.16E-05 | 0.000522986 |
| Q9H4B7 | Q9H4B7 | TUBB1 | -6.88 | -5.84 | 5.47E-05 | 0.000671617 |
| P02656 | P02656 | APOC3 | -1.91 | -1.92 | 5.83E-05 | 0.000699589 |
| Q05655-2 | Q05655 | PRKCD | 1.31 | -5.04 | 6.34E-05 | 0.000737564 |
| P11171-5 | P11171 | EPB41 | 1.69 | -3.08 | 6.43E-05 | 0.000737564 |
| Q5T0N5-4 | Q5T0N5 | FNBP1L | 6.29 | -11.97 | 6.90E-05 | 0.000774368 |
| P04792 | P04792 | HSPB1 | -2.25 | -6.23 | 7.11E-05 | 0.000780482 |
| Q9BXF9 | Q9BXF9 | TEKT3 | -5.03 | -15.52 | 7.68E-05 | 0.000825179 |
| P08697-1 | P08697 | SERPINF2 | -2.41 | -6.74 | 8.03E-05 | 0.000845126 |
| Q8TB72-1 | Q8TB72 | PUM2 | 4.40 | -12.63 | 9.11E-05 | 0.000940556 |
| P04004 | P04004 | VTN | -1.83 | -5.32 | 9.97E-05 | 0.000992995 |
| P01042-2 | P01042 | KNG1 | -1.53 | 2.31 | 1.02E-04 | 0.000992995 |
| Q92508 | Q92508 | PIEZO1 | 1.55 | -4.65 | 1.02E-04 | 0.000992995 |
| P17301 | P17301 | ITGA2 | 5.67 | -10.77 | 1.08E-04 | 0.001028458 |
| P60174-1 | P60174 | TPI1 | -5.90 | -15.30 | 1.21E-04 | 0.001133449 |
| P05556-1 | P05556 | ITGB1 | -6.89 | -14.07 | 1.25E-04 | 0.001150319 |
| Q9Y3X0 | Q9Y3X0 | CCDC9 | 3.69 | -15.13 | 1.35E-04 | 0.00122611 |
| Q13442 | Q13442 | PDAP1 | 5.66 | -13.04 | 1.57E-04 | 0.001399947 |
| O15231-3 | O15231 | ZNF185 | 5.19 | -13.11 | 1.67E-04 | 0.001458748 |
| O14639-1 | O14639 | ABLIM1 | 4.68 | -10.57 | 1.70E-04 | 0.001463708 |
| Q9BQE3 | Q9BQE3 | TUBA1C | 6.15 | -9.96 | 1.84E-04 | 0.001560487 |
| O15194-1 | O15194 | CTDSPL | 5.32 | -12.16 | 2.05E-04 | 0.001708754 |
| Q7Z3Y8 | Q7Z3Y8 | KRT27 | -5.18 | -14.76 | 2.17E-04 | 0.001775345 |
| P23142 | P23142 | FBLN1 | -2.37 | -6.95 | 2.26E-04 | 0.001818188 |
| Q13107-2 | Q13107 | USP4 | 6.15 | -11.78 | 2.40E-04 | 0.001890698 |
| P13646 | P13646 | KRT13 | -3.30 | -6.73 | 2.43E-04 | 0.001890698 |
| O00264 | O00264 | PGRMC1 | 1.44 | -3.45 | 2.45E-04 | 0.001890698 |
| P12259 | P12259 | F5 | -2.04 | -6.38 | 2.86E-04 | 0.002167265 |
| P02679 | P02679 | FGG | -1.98 | -1.32 | 2.92E-04 | 0.002183714 |
| Q96C24 | Q96C24 | SYTL4 | 1.70 | -5.89 | 0.000325831 | 0.002401838 |
| P22466 | P22466 | GAL | -5.80 | -12.83 | 0.000341143 | 0.002479292 |
| Q5T1M5 | Q5T1M5 | FKBP15 | -5.63 | -14.82 | 0.000350127 | 0.002509243 |
| Q14644 | Q14644 | RASA3 | 1.79 | -5.48 | 0.000427496 | 0.003021754 |
| P04003 | P04003 | C4BPA | -2.00 | -6.09 | 0.000454468 | 0.003168994 |
| O94769-2 | O94769 | ECM2 | -5.60 | -14.14 | 0.000484811 | 0.003335499 |
| P35612-1 | P35612 | ADD2 | 1.76 | -2.66 | 0.000494498 | 0.003357384 |
| Q86UE4 | Q86UE4 | MTDH | 5.98 | -9.49 | 0.000514973 | 0.003416857 |
| Q9Y4D1-2 | Q9Y4D1 | DAAM1 | 5.58 | -10.84 | 0.000516502 | 0.003416857 |
| O60841 | O60841 | EIF5B | 5.17 | -11.19 | 0.000527027 | 0.003442351 |
| P42566-1 | P42566 | EPS15 | 1.16 | -7.17 | 0.000586752 | 0.003741125 |
| P21333 | P21333 | FLNA | 1.56 | -1.17 | 0.00058727 | 0.003741125 |
| P49407-1 | P49407 | ARRB1 | 1.49 | -6.78 | 0.00059508 | 0.003744649 |
| O43306 | O43306 | ADCY6 | 1.42 | -6.77 | 0.000635506 | 0.003950854 |
| Q5JSP0-2 | Q5JSP0 | FGD3 | 1.15 | -6.72 | 0.000652502 | 0.004008228 |
| O43432-3 | O43432 | EIF4G3 | 3.77 | -12.68 | 0.000681219 | 0.004135401 |
| Q0ZGT2 | Q0ZGT2 | NEXN | 1.62 | -5.31 | 0.00072071 | 0.004289759 |
| Q8TEW0 | Q8TEW0 | PARD3 | 5.44 | -11.63 | 0.000739956 | 0.004289759 |
| Q9Y210-1 | Q9Y210 | TRPC6 | 2.90 | -8.86 | 0.000741569 | 0.004289759 |
| P23528 | P23528 | CFL1 | 1.05 | -2.10 | 0.000746058 | 0.004289759 |
| Q16204 | Q16204 | CCDC6 | 2.54 | -15.93 | 0.00075046 | 0.004289759 |
| P08603-1 | P08603 | CFH | -2.37 | -8.04 | 0.000756527 | 0.004289759 |
| O14713-1 | O14713 | ITGB1BP1 | 4.90 | -12.13 | 0.000783049 | 0.004391881 |
| Q9UIL4 | Q9UIL4 | KIF25 | -6.90 | -14.54 | 0.000810858 | 0.004498954 |
| P01859 | P01859 | NA | -1.90 | -5.66 | 0.00085774 | 0.004701098 |
| P06396 | P06396 | GSN | -1.70 | -6.88 | 0.000872004 | 0.004701098 |
| Q7LDG7-4 | Q7LDG7 | RASGRP2 | 5.75 | -11.16 | 0.000874623 | 0.004701098 |
| O95292 | O95292 | VAPB | 3.94 | -11.82 | 0.000885153 | 0.004708651 |
| P19474 | P19474 | TRIM21 | -6.11 | -15.11 | 0.001039244 | 0.005471936 |
| Q8N163-1 | Q8N163 | CCAR2 | -4.87 | -15.11 | 0.001076648 | 0.005555504 |
| Q8WWA1 | Q8WWA1 | TMEM40 | -3.82 | -8.41 | 0.001151783 | 0.005884356 |
| Q08830 | Q08830 | FGL1 | -4.85 | -14.29 | 0.001203949 | 0.006090565 |
| Q0JRZ9-1 | Q0JRZ9 | FCHO2 | 4.98 | -13.42 | 0.001250606 | 0.006214707 |
| P04921-1 | P04921 | GYPC | -5.64 | -13.34 | 0.001350532 | 0.006636898 |
| P68366 | P68366 | TUBA4A | -5.71 | -12.16 | 0.00138983 | 0.006765588 |
| P01008 | P01008 | SERPINC1 | -1.79 | -7.57 | 0.001407754 | 0.006766817 |
| Q9Y3P9 | Q9Y3P9 | RABGAP1 | 4.11 | -13.40 | 0.001424003 | 0.006766817 |
| Q9UJ41-1 | Q9UJ41 | RABGEF1 | 4.44 | -13.04 | 0.001429424 | 0.006766817 |
| P30273 | P30273 | FCER1G | 1.01 | -4.42 | 0.001555521 | 0.007296806 |
| Q8TEQ6 | Q8TEQ6 | GEMIN5 | 3.48 | -15.18 | 0.001580523 | 0.007347295 |
| Q9Y5S2 | Q9Y5S2 | CDC42BPB | -6.63 | -10.29 | 0.001623422 | 0.007479335 |
| Q15311 | Q15311 | RALBP1 | 4.07 | -13.08 | 0.001684243 | 0.007623416 |
| Q9Y624 | Q9Y624 | F11R | 1.33 | -5.72 | 0.001742431 | 0.007818213 |
| O00159-1 | O00159 | MYO1C | 3.52 | -13.90 | 0.001817548 | 0.008084956 |
| P13798 | P13798 | APEH | -4.34 | -14.25 | 0.001902274 | 0.008375546 |
| Q9NRX5 | Q9NRX5 | SERINC1 | 2.75 | -9.31 | 0.001926541 | 0.008375546 |
| P10412 | P10412 | H1-4 | -5.43 | -14.27 | 0.00193157 | 0.008375546 |
| Q8NEZ4 | Q8NEZ4 | KMT2C | 6.02 | -12.37 | 0.001953217 | 0.008398833 |
| Q96BY6-1 | Q96BY6 | DOCK10 | 4.77 | -11.54 | 0.001971722 | 0.008408334 |
| Q12913-1 | Q12913 | PTPRJ | 1.51 | -5.55 | 0.001996849 | 0.008437643 |
| Q7Z6I6-1 | Q7Z6I6 | ARHGAP30 | -4.49 | -11.36 | 0.002057565 | 0.008562127 |
| Q6PJF5-1 | Q6PJF5 | RHBDF2 | 1.18 | -5.65 | 0.002097458 | 0.008658308 |
| Q8TCJ2 | Q8TCJ2 | STT3B | 3.69 | -12.34 | 0.002134974 | 0.008675872 |
| P13647 | P13647 | KRT5 | -3.15 | -7.86 | 0.002135341 | 0.008675872 |
| P49841-1 | P49841 | GSK3B | 1.00 | -6.38 | 0.002185245 | 0.008809268 |
| Q0VD83-1 | Q0VD83 | APOBR | -4.89 | -14.61 | 0.002230141 | 0.008832986 |
| P14618 | P14618 | PKM | -2.31 | -8.45 | 0.002238736 | 0.008832986 |
| Q13099-3 | Q13099 | IFT88 | -5.66 | -14.28 | 0.0022443 | 0.008832986 |
| P17936 | P17936 | IGFBP3 | -6.97 | -13.12 | 0.002271019 | 0.008832986 |
| P19823 | P19823 | ITIH2 | -1.65 | -4.91 | 0.002306956 | 0.008832986 |
| P49006 | P49006 | MARCKSL1 | 4.21 | -9.71 | 0.00232737 | 0.008832986 |
| Q92954-6 | Q92954 | PRG4 | -1.54 | -4.15 | 0.002328074 | 0.008832986 |
| Q562R1 | Q562R1 | ACTBL2 | 3.09 | -15.79 | 0.002354496 | 0.00886803 |
| Q8N9U0 | Q8N9U0 | TC2N | 2.05 | -8.38 | 0.002472108 | 0.009208621 |
| Q8N1G4 | Q8N1G4 | LRRC47 | -5.22 | -13.67 | 0.002480617 | 0.009208621 |
| P16615-4 | P16615 | ATP2A2 | 3.22 | -8.95 | 0.002504485 | 0.009230817 |
| P10909-2 | P10909 | CLU | -1.12 | -3.53 | 0.002614334 | 0.009499974 |
| Q9BSQ5-1 | Q9BSQ5 | CCM2 | -4.86 | -13.94 | 0.002693753 | 0.009720118 |
| P19634-1 | P19634 | SLC9A1 | 1.12 | -5.90 | 0.002733903 | 0.009789973 |
| P20701-2 | P20701 | ITGAL | -4.69 | -13.25 | 0.002751058 | 0.009789973 |
| Q9Y613 | Q9Y613 | FHOD1 | 1.26 | -6.96 | 0.003056572 | 0.010802678 |
| E9PAV3 | E9PAV3 | NA | 4.40 | -12.15 | 0.003126121 | 0.010931914 |
| P16298 | P16298 | PPP3CB | 2.85 | -15.65 | 0.003155908 | 0.010931914 |
| P36507 | P36507 | MAP2K2 | 1.24 | -6.86 | 0.003156696 | 0.010931914 |
| Q14156-1 | Q14156 | EFR3A | 3.47 | -15.03 | 0.00330749 | 0.011302418 |
| P02724 | P02724 | GYPA | 3.20 | -1.19 | 0.004029091 | 0.013412974 |
| Q9BUN1 | Q9BUN1 | C1orf56 | -1.22 | -7.36 | 0.004250564 | 0.014057846 |
| Q13459-1 | Q13459 | MYO9B | 1.22 | -6.21 | 0.004286308 | 0.014057846 |
| P13861 | P13861 | PRKAR2A | 4.14 | -13.81 | 0.004304534 | 0.014057846 |
| P08575-2 | P08575 | PTPRC | -1.05 | -3.92 | 0.004408471 | 0.014176816 |
| P02671-1 | P02671 | FGA | -1.97 | 0.09 | 0.004415454 | 0.014176816 |
| P22059 | P22059 | OSBP | 4.81 | -11.16 | 0.004423387 | 0.014176816 |
| Q92556-1 | Q92556 | ELMO1 | 4.60 | -12.25 | 0.004542367 | 0.01446828 |
| P23588 | P23588 | EIF4B | 3.05 | -12.66 | 0.004973158 | 0.015743249 |
| Q9Y490 | Q9Y490 | TLN1 | 1.27 | -4.44 | 0.005065057 | 0.0159364 |
| Q8N370-1 | Q8N370 | SLC43A2 | -4.90 | -11.28 | 0.005098919 | 0.015945711 |
| P02549-1 | P02549 | SPTA1 | 1.54 | 2.06 | 0.005276263 | 0.016400915 |
| P37802 | P37802 | TAGLN2 | 4.32 | -9.82 | 0.005663871 | 0.017396174 |
| Q15746-4 | Q15746 | MYLK | 3.01 | -7.05 | 0.00577991 | 0.017647536 |
| P54277 | P54277 | PMS1 | 5.04 | -12.33 | 0.006123258 | 0.018585888 |
| Q92609-1 | Q92609 | TBC1D5 | 2.13 | -14.97 | 0.006314656 | 0.019054752 |
| P04406-1 | P04406 | GAPDH | -1.56 | -4.09 | 0.006607739 | 0.019823218 |
| Q96TC7 | Q96TC7 | RMDN3 | -4.06 | -11.32 | 0.007553079 | 0.022144254 |
| P21731 | P21731 | TBXA2R | -1.60 | -8.84 | 0.007672635 | 0.022367683 |
| P00450 | P00450 | CP | -1.26 | -6.56 | 0.009107719 | 0.02632237 |
| Q9ULI3 | Q9ULI3 | HEG1 | -3.78 | -13.47 | 0.009175637 | 0.02632237 |
| P0C7M8 | P0C7M8 | CLEC2L | 4.06 | -13.29 | 0.009182222 | 0.02632237 |
| Q6IQ22 | Q6IQ22 | RAB12 | 3.32 | -13.94 | 0.009310416 | 0.026490344 |
| O95183 | O95183 | VAMP5 | 3.05 | -14.92 | 0.009343494 | 0.026490344 |
| Q92539 | Q92539 | LPIN2 | 2.45 | -15.95 | 0.009741486 | 0.027327841 |
| Q13404 | Q13404 | UBE2V1 | 2.68 | -15.90 | 0.009806304 | 0.027351636 |
| O60232 | O60232 | ZNRD2 | -5.13 | -11.96 | 0.010400489 | 0.028852969 |
| Q9P107 | Q9P107 | GMIP | 1.04 | -6.49 | 0.01058678 | 0.029212718 |
| Q9P270 | Q9P270 | SLAIN2 | 3.54 | -12.75 | 0.010770732 | 0.029562223 |
| P11836 | P11836 | MS4A1 | -1.12 | -7.16 | 0.010850498 | 0.029623583 |
| O15417-1 | O15417 | TNRC18 | 1.87 | -8.48 | 0.01096197 | 0.029770404 |
| P16402 | P16402 | H1-3 | -4.68 | -14.11 | 0.011020045 | 0.029771432 |
| P15374 | P15374 | UCHL3 | -5.25 | -12.60 | 0.011113629 | 0.029867877 |
| Q7Z401-1 | Q7Z401 | DENND4A | 1.23 | -4.28 | 0.011802521 | 0.031554928 |
| Q8NC44 | Q8NC44 | RETREG2 | 3.38 | -11.25 | 0.013009999 | 0.034426458 |
| Q9UNE0 | Q9UNE0 | EDAR | 3.42 | -14.69 | 0.014031183 | 0.036751729 |
| P19827-1 | P19827 | ITIH1 | -1.60 | -7.13 | 0.014480967 | 0.037594972 |
| P35611-3 | P35611 | ADD1 | 1.08 | -0.60 | 0.014498836 | 0.037594972 |
| Q05209-1 | Q05209 | PTPN12 | 4.27 | -10.65 | 0.015009629 | 0.038724842 |
| P13224 | P13224 | GP1BB | -1.35 | -4.44 | 0.015534892 | 0.039880619 |
| Q86UX7 | Q86UX7 | FERMT3 | -4.53 | -12.13 | 0.015616532 | 0.039891735 |
| Q8TAD7 | Q8TAD7 | C12orf75 | 1.71 | -8.01 | 0.016213939 | 0.040909829 |
| P31749 | P31749 | AKT1 | 2.93 | -12.51 | 0.016252936 | 0.040909829 |
| Q9Y666 | Q9Y666 | SLC12A7 | 3.09 | -11.74 | 0.01759095 | 0.043639087 |
| Q04656-1 | Q04656 | ATP7A | 2.99 | -15.45 | 0.017792159 | 0.043927053 |
| Q8WWN9 | Q8WWN9 | IPCEF1 | 3.74 | -12.01 | 0.018570249 | 0.045629755 |
| O00421 | O00421 | CCRL2 | 3.47 | -13.31 | 0.019090324 | 0.046685343 |

**Supplementary Table ST5. List of differentially expressed EV proteins: T2DM vs. PDM**

| **Accession** | **UNIPROT** | **SYMBOL** | **logFC** | **AveExpr** | **P.Value** | **adj.P.Val** |
| --- | --- | --- | --- | --- | --- | --- |
| Q13336-2 | Q13336 | SLC14A1 | -4.98 | -5.21 | 2.24E-20 | 4.19E-17 |
| P30046 | P30046 | DDT | 4.89 | -8.47 | 1.36E-09 | 1.27E-06 |
| P80511 | P80511 | S100A12 | 8.84 | -14.68 | 7.58E-08 | 4.12E-05 |
| Q8TB69 | Q8TB69 | ZNF519 | -7.57 | -17.88 | 8.80E-08 | 4.12E-05 |
| Q8TCG5-2 | Q8TCG5 | CPT1C | -9.46 | -15.20 | 1.69E-07 | 6.33E-05 |
| P68133 | P68133 | ACTA1 | 1.46 | -3.42 | 3.20E-05 | 0.0099891 |
| O75390 | O75390 | CS | 1.73 | -9.44 | 3.91E-05 | 0.0104519 |
| P49257 | P49257 | LMAN1 | -5.67 | -18.31 | 4.79E-05 | 0.0112101 |
| P43686 | P43686 | PSMC4 | 5.97 | -17.04 | 7.24E-05 | 0.0150474 |
| O95602 | O95602 | POLR1A | -5.30 | -18.72 | 9.04E-05 | 0.0169168 |
| P36969 | P36969 | GPX4 | 2.66 | -9.72 | 0.0001063 | 0.0180701 |
| Q9NZT1 | Q9NZT1 | CALML5 | -1.18 | -8.00 | 0.0001159 | 0.0180701 |
| P80723 | P80723 | BASP1 | 0.81 | -7.99 | 0.0001395 | 0.0200743 |
| P60903 | P60903 | S100A10 | 6.52 | -15.73 | 0.0002112 | 0.026432 |
| Q9Y336 | Q9Y336 | SIGLEC9 | 4.37 | -18.91 | 0.0002119 | 0.026432 |
| Q5JPH6 | Q5JPH6 | EARS2 | 5.51 | -15.35 | 0.0003195 | 0.0373656 |
| Q8N335 | Q8N335 | GPD1L | 4.52 | -18.77 | 0.0004306 | 0.0473879 |
| P05387 | P05387 | RPLP2 | -1.26 | -7.87 | 0.0006016 | 0.062528 |
| O94910 | O94910 | ADGRL1 | -2.54 | -18.86 | 0.0006544 | 0.0628449 |
| Q00688 | Q00688 | FKBP3 | -5.17 | -18.04 | 0.0007253 | 0.0628449 |
| P11586 | P11586 | MTHFD1 | -0.68 | -7.52 | 0.0007398 | 0.0628449 |
| P61077-1 | P61077 | UBE2D3 | 6.41 | -17.00 | 0.0007725 | 0.0628449 |
| Q9HC10 | Q9HC10 | OTOF | -0.94 | -9.21 | 0.0008114 | 0.0632543 |
| Q9UJJ9 | Q9UJJ9 | GNPTG | -4.56 | -18.03 | 0.0009394 | 0.0703023 |
| Q86YW5-2 | Q86YW5 | TREML1 | 4.89 | -16.23 | 0.001006 | 0.0723912 |
| P22792 | P22792 | CPN2 | -0.69 | -3.05 | 0.001085 | 0.0751873 |
| P56192 | P56192 | MARS1 | -6.45 | -16.47 | 0.0012046 | 0.0804912 |
| Q14766-4 | Q14766 | LTBP1 | 0.62 | -5.06 | 0.0013327 | 0.0859818 |
| P22735 | P22735 | TGM1 | 5.95 | -12.18 | 0.0014065 | 0.0877161 |
| P13667 | P13667 | PDIA4 | 2.01 | -11.84 | 0.0014993 | 0.0885993 |
| P62495 | P62495 | ETF1 | 4.00 | -11.58 | 0.0015153 | 0.0885993 |
| Q6UXG3 | Q6UXG3 | CD300LG | 5.56 | -16.92 | 0.0016714 | 0.0928609 |
| P15311 | P15311 | EZR | -0.42 | -7.94 | 0.0017343 | 0.0928609 |
| P01833 | P01833 | PIGR | -0.68 | -6.68 | 0.0017371 | 0.0928609 |
| P29508 | P29508 | SERPINB3 | 0.99 | -8.27 | 0.0018212 | 0.0938469 |
| P42768 | P42768 | WAS | -5.29 | -13.85 | 0.0020256 | 0.0938469 |
| Q96SJ8 | Q96SJ8 | TSPAN18 | 5.52 | -16.72 | 0.0020358 | 0.0938469 |
| P01137 | P01137 | TGFB1 | 1.50 | -7.17 | 0.0020408 | 0.0938469 |
| Q6P3W7 | Q6P3W7 | SCYL2 | 3.97 | -16.96 | 0.0021032 | 0.0938469 |
| P35813-3 | P35813 | PPM1A | 1.05 | -10.35 | 0.0021951 | 0.0938469 |
| Q96M91 | Q96M91 | CFAP53 | -3.51 | -15.15 | 0.0022154 | 0.0938469 |
| Q5VTH9-1 | Q5VTH9 | WDR78 | 6.07 | -16.17 | 0.0023019 | 0.0938469 |
| NA | NA | NA | -0.84 | -8.26 | 0.0023328 | 0.0938469 |
| P36955 | P36955 | SERPINF1 | -0.91 | -4.80 | 0.0023519 | 0.0938469 |
| Q08050 | Q08050 | FOXM1 | 6.22 | -16.67 | 0.0023525 | 0.0938469 |
| P09668 | P09668 | CTSH | -5.66 | -16.04 | 0.0023575 | 0.0938469 |
| P04406-1 | P04406 | GAPDH | 0.70 | -4.08 | 0.0025347 | 0.0987997 |
| Q13867 | Q13867 | BLMH | -1.02 | -7.41 | 0.0029273 | 0.1101539 |
| P01782 | P01782 | NA | 0.79 | -5.09 | 0.0029437 | 0.1101539 |
| O15173 | O15173 | PGRMC2 | 5.35 | -16.04 | 0.0030792 | 0.1122574 |
| O14617-1 | O14617 | AP3D1 | 5.23 | -11.13 | 0.0031199 | 0.1122574 |
| O15394 | O15394 | NCAM2 | 5.72 | -16.61 | 0.0031922 | 0.112689 |
| P40763-2 | P40763 | STAT3 | -3.90 | -11.82 | 0.003295 | 0.1141647 |
| P01127-2 | P01127 | PDGFB | 0.70 | -8.85 | 0.0035748 | 0.1200549 |
| Q15389-2 | Q15389 | ANGPT1 | -1.06 | -9.02 | 0.0036494 | 0.1200549 |
| P08493-1 | P08493 | MGP | 4.57 | -11.44 | 0.0037072 | 0.1200549 |
| P11836 | P11836 | MS4A1 | -2.63 | -10.47 | 0.0037216 | 0.1200549 |
| P21283 | P21283 | ATP6V1C1 | 3.67 | -18.66 | 0.00387 | 0.1227241 |
| P35998 | P35998 | PSMC2 | -0.89 | -8.77 | 0.0045188 | 0.1409122 |
| Q14258 | Q14258 | TRIM25 | 0.80 | -9.10 | 0.0047921 | 0.1469853 |
| P38646 | P38646 | HSPA9 | -4.41 | -9.22 | 0.0051151 | 0.1499051 |
| Q08499-6 | Q08499 | PDE4D | -0.44 | -6.94 | 0.0051605 | 0.1499051 |
| Q7Z794 | Q7Z794 | KRT77 | -1.01 | -5.89 | 0.0051787 | 0.1499051 |
| P07451 | P07451 | CA3 | 4.10 | -18.36 | 0.0052078 | 0.1499051 |
| Q8N4M1 | Q8N4M1 | SLC44A3 | 5.12 | -15.46 | 0.0054309 | 0.1532398 |
| P19013 | P19013 | NA | -1.29 | -7.92 | 0.0056157 | 0.1532398 |
| P30491 | P30491 | NA | -0.56 | -3.28 | 0.0056501 | 0.1532398 |
| O15259 | O15259 | NPHP1 | -5.65 | -10.73 | 0.0056513 | 0.1532398 |
| P04080 | P04080 | CSTB | 1.12 | -8.45 | 0.0057575 | 0.153891 |
| P51452 | P51452 | DUSP3 | 1.13 | -8.14 | 0.0059623 | 0.1571194 |
| P05109 | P05109 | S100A8 | 0.52 | -4.67 | 0.0060759 | 0.157854 |
| Q9ULH1-2 | Q9ULH1 | ASAP1 | -3.09 | -10.53 | 0.0061605 | 0.157854 |
| P01718 | P01718 | NA | -5.20 | -13.84 | 0.0062433 | 0.157854 |
| O00571 | O00571 | DDX3X | -6.57 | -14.84 | 0.0064728 | 0.161475 |
| P06729 | P06729 | CD2 | -5.37 | -17.21 | 0.0068293 | 0.1664038 |
| P04792 | P04792 | HSPB1 | 0.60 | -8.05 | 0.0069693 | 0.1664038 |
| Q96AG4 | Q96AG4 | LRRC59 | 3.78 | -18.06 | 0.0070126 | 0.1664038 |
| P26038 | P26038 | MSN | -0.38 | -2.92 | 0.0070261 | 0.1664038 |
| P52566 | P52566 | ARHGDIB | 0.70 | -5.40 | 0.0072062 | 0.1671465 |
| Q14005-1 | Q14005 | IL16 | -0.47 | -7.25 | 0.0072422 | 0.1671465 |
| P21589-1 | P21589 | NT5E | -4.59 | -12.29 | 0.0073411 | 0.1671465 |
| P19320-1 | P19320 | VCAM1 | -4.33 | -17.18 | 0.0075558 | 0.1671465 |
| P00403 | P00403 | COX2 | 4.91 | -17.29 | 0.007584 | 0.1671465 |
| Q9Y4D8-1 | Q9Y4D8 | NA | 2.34 | -19.00 | 0.0075935 | 0.1671465 |
| A6QL63-1 | A6QL63 | BTBD11 | 6.57 | -15.86 | 0.007741 | 0.1684122 |
| P53396-1 | P53396 | ACLY | 0.48 | -6.61 | 0.007944 | 0.170841 |
| P07339 | P07339 | CTSD | -0.48 | -6.58 | 0.0081626 | 0.1734589 |
| P41218 | P41218 | MNDA | 5.06 | -15.31 | 0.0082511 | 0.1734589 |
| P09493-5 | P09493 | TPM1 | 1.07 | -7.82 | 0.0085877 | 0.1760555 |
| A0A0C4DH31 | A0A0C4DH31 | NA | -1.17 | -9.00 | 0.0085934 | 0.1760555 |
| P27824 | P27824 | CANX | 0.65 | -6.25 | 0.0086642 | 0.1760555 |
| Q8IZT6-1 | Q8IZT6 | ASPM | 0.69 | -6.60 | 0.008751 | 0.1760555 |
| P20963-1 | P20963 | CD247 | -3.12 | -11.58 | 0.0089952 | 0.1767802 |
| Q8N1N4 | Q8N1N4 | KRT78 | -0.76 | -6.87 | 0.0090679 | 0.1767802 |
| Q9UI42-1 | Q9UI42 | CPA4 | -3.95 | -18.30 | 0.009148 | 0.1767802 |
| P20701-2 | P20701 | ITGAL | -1.16 | -7.30 | 0.009165 | 0.1767802 |
| A2RUR9-3 | A2RUR9 | CCDC144A | 0.63 | -9.59 | 0.0092619 | 0.1768265 |
| P0DMM9 | P0DMM9 | SULT1A3 | 5.09 | -14.01 | 0.0096224 | 0.1805141 |
| Q9BX46-2 | Q9BX46 | RBM24 | -4.35 | -16.54 | 0.0097899 | 0.1805141 |
| Q9Y4A5 | Q9Y4A5 | TRRAP | -3.85 | -17.80 | 0.009932 | 0.1805141 |
| P49411 | P49411 | TUFM | -0.71 | -9.21 | 0.0100869 | 0.1805141 |
| Q9NRW1 | Q9NRW1 | RAB6B | 0.99 | -7.94 | 0.0101074 | 0.1805141 |
| Q30KQ8 | Q30KQ8 | NA | -4.65 | -15.98 | 0.0101143 | 0.1805141 |
| P00915 | P00915 | CA1 | 0.86 | -3.26 | 0.0102252 | 0.1805141 |
| O75113 | O75113 | N4BP1 | -4.26 | -17.96 | 0.0103341 | 0.1805141 |
| P50416 | P50416 | CPT1A | -3.99 | -17.23 | 0.0103594 | 0.1805141 |
| P01619 | P01619 | NA | 0.68 | -5.49 | 0.0104198 | 0.1805141 |
| Q15555-4 | Q15555 | MAPRE2 | 0.74 | -6.76 | 0.0106032 | 0.1820056 |
| Q14573 | Q14573 | ITPR3 | 4.53 | -17.02 | 0.0111752 | 0.1863211 |
| P13473-3 | P13473 | LAMP2 | 3.40 | -9.57 | 0.0112256 | 0.1863211 |
| O94919 | O94919 | ENDOD1 | 0.77 | -7.35 | 0.0113027 | 0.1863211 |
| O95373 | O95373 | IPO7 | -0.45 | -6.54 | 0.0113232 | 0.1863211 |
| Q3ZCW2 | Q3ZCW2 | LGALSL | 0.98 | -8.86 | 0.0113525 | 0.1863211 |
| P36222 | P36222 | CHI3L1 | -5.03 | -15.15 | 0.0117871 | 0.189553 |
| P08397-1 | P08397 | HMBS | -3.26 | -16.37 | 0.0119678 | 0.189553 |
| P01920 | P01920 | HLA-DQB1 | -3.27 | -18.31 | 0.0120688 | 0.189553 |
| P17010 | P17010 | ZFX | -4.46 | -16.77 | 0.0121164 | 0.189553 |
| P22748 | P22748 | CA4 | 0.49 | -7.81 | 0.0121203 | 0.189553 |
| P15090 | P15090 | FABP4 | -0.71 | -8.33 | 0.0121573 | 0.189553 |
| Q99714-1 | Q99714 | HSD17B10 | 2.37 | -10.50 | 0.0122872 | 0.1899953 |
| Q13387-1 | Q13387 | MAPK8IP2 | -4.56 | -17.29 | 0.0130315 | 0.1965086 |
| P55145 | P55145 | MANF | 4.50 | -17.45 | 0.013096 | 0.1965086 |
| P04632 | P04632 | CAPNS1 | 0.67 | -7.11 | 0.0131112 | 0.1965086 |
| Q96PL5 | Q96PL5 | ERMAP | -0.69 | -8.26 | 0.0131286 | 0.1965086 |
| Q9H0B8 | Q9H0B8 | CRISPLD2 | -0.96 | -8.24 | 0.0133377 | 0.1980548 |
| Q15907 | Q15907 | RAB11B | 0.63 | -6.28 | 0.013606 | 0.2004471 |
| Q96HC4-4 | Q96HC4 | PDLIM5 | -0.73 | -8.69 | 0.0140398 | 0.2052229 |
| Q9Y6Z7 | Q9Y6Z7 | COLEC10 | -1.09 | -8.51 | 0.0142134 | 0.2061488 |
| Q3T906 | Q3T906 | GNPTAB | -4.07 | -14.34 | 0.0144009 | 0.2072626 |
| Q9BPZ7 | Q9BPZ7 | MAPKAP1 | 4.00 | -17.55 | 0.0152258 | 0.2174616 |
| Q92954-3 | Q92954 | PRG4 | -0.39 | -2.51 | 0.0156898 | 0.2179589 |
| Q8NI99 | Q8NI99 | ANGPTL6 | -1.36 | -10.02 | 0.0157136 | 0.2179589 |
| O75037 | O75037 | KIF21B | -4.36 | -17.28 | 0.0157386 | 0.2179589 |
| Q8TBZ0-1 | Q8TBZ0 | CCDC110 | -0.70 | -8.87 | 0.0158268 | 0.2179589 |
| P16402 | P16402 | H1-3 | -1.24 | -8.60 | 0.0158431 | 0.2179589 |
| O43681 | O43681 | GET3 | 4.44 | -16.17 | 0.0163646 | 0.2234896 |
| Q9GZV3 | Q9GZV3 | SLC5A7 | -0.58 | -10.24 | 0.0165381 | 0.2234949 |
| P04424 | P04424 | ASL | 3.57 | -17.14 | 0.0167724 | 0.2234949 |
| P06753-2 | P06753 | TPM3 | 0.55 | -3.83 | 0.0168312 | 0.2234949 |
| O43488 | O43488 | AKR7A2 | -2.18 | -7.61 | 0.0168427 | 0.2234949 |
| O00299 | O00299 | CLIC1 | 0.55 | -5.08 | 0.0172431 | 0.2271956 |
| Q96DA0 | Q96DA0 | ZG16B | 1.25 | -8.94 | 0.0176443 | 0.2287461 |
| Q8WWZ8-1 | Q8WWZ8 | OIT3 | -0.89 | -6.67 | 0.0176965 | 0.2287461 |
| A0A0J9YX35 | A0A0J9YX35 | NA | -0.49 | -5.63 | 0.0177275 | 0.2287461 |
| Q6DD88 | Q6DD88 | ATL3 | -0.29 | -6.53 | 0.0180173 | 0.2308923 |
| P98160 | P98160 | HSPG2 | 0.67 | -3.51 | 0.0182419 | 0.2321804 |
| P28065 | P28065 | PSMB9 | 3.10 | -16.06 | 0.018532 | 0.2342789 |
| Q9C0D6 | Q9C0D6 | FHDC1 | -0.49 | -9.59 | 0.0186717 | 0.2344616 |
| P28289 | P28289 | TMOD1 | 0.81 | -6.10 | 0.0190573 | 0.2377076 |
| P20813 | P20813 | CYP2B6 | -4.19 | -13.90 | 0.0193074 | 0.2378661 |
| P60174 | P60174 | TPI1 | 0.58 | -5.71 | 0.0193242 | 0.2378661 |
| Q9UBW5-1 | Q9UBW5 | BIN2 | -0.88 | -5.65 | 0.0194543 | 0.2379019 |
| Q8N8Q8-1 | Q8N8Q8 | COX18 | -4.98 | -13.91 | 0.0198311 | 0.2399944 |
| Q0ZGT2 | Q0ZGT2 | NEXN | -0.46 | -5.59 | 0.019882 | 0.2399944 |
| P43250-2 | P43250 | GRK6 | -5.20 | -15.40 | 0.0201404 | 0.2415555 |
| O75947-1 | O75947 | ATP5PD | 4.06 | -15.99 | 0.0203741 | 0.2418743 |
| P16157 | P16157 | ANK1 | 0.54 | -0.13 | 0.0204255 | 0.2418743 |
| Q9UBC2-4 | Q9UBC2 | EPS15L1 | -0.87 | -10.01 | 0.0207573 | 0.2430414 |
| P49748-1 | P49748 | ACADVL | -0.45 | -7.17 | 0.0208885 | 0.2430414 |
| P06703 | P06703 | S100A6 | 0.87 | -7.33 | 0.0209561 | 0.2430414 |
| Q6UXI9 | Q6UXI9 | NPNT | -3.77 | -12.14 | 0.0211561 | 0.2430414 |
| NA | NA | NA | 4.67 | -16.77 | 0.0211736 | 0.2430414 |
| P59534 | P59534 | TAS2R39 | -4.18 | -16.32 | 0.021379 | 0.2439031 |
| P11142-1 | P11142 | HSPA8 | 0.50 | -3.91 | 0.0222686 | 0.2525127 |
| O43865 | O43865 | AHCYL1 | -0.40 | -8.62 | 0.0226404 | 0.2547215 |
| P02549-1 | P02549 | SPTA1 | 0.54 | 0.22 | 0.0229408 | 0.2547215 |
| P48506 | P48506 | GCLC | 3.42 | -13.71 | 0.0230213 | 0.2547215 |
| A0A075B6I1 | A0A075B6I1 | NA | -3.55 | -14.59 | 0.0231599 | 0.2547215 |
| Q9Y4D1-2 | Q9Y4D1 | DAAM1 | 0.57 | -7.61 | 0.0232816 | 0.2547215 |
| P12273 | P12273 | PIP | -3.93 | -12.06 | 0.0234213 | 0.2547215 |
| P0C7P3-1 | P0C7P3 | SLFN14 | 0.88 | -5.10 | 0.0236808 | 0.2547215 |
| P04075 | P04075 | ALDOA | 0.51 | -4.78 | 0.0237108 | 0.2547215 |
| Q8NBF2-1 | Q8NBF2 | NHLRC2 | -3.41 | -10.46 | 0.0237275 | 0.2547215 |
| Q9NS84 | Q9NS84 | CHST7 | -1.65 | -17.84 | 0.0238248 | 0.2547215 |
| P25787 | P25787 | PSMA2 | -0.40 | -6.87 | 0.0240093 | 0.2552356 |
| Q13790 | Q13790 | APOF | -0.34 | -5.52 | 0.0246735 | 0.2608143 |
| Q9Y613 | Q9Y613 | FHOD1 | -0.97 | -4.23 | 0.0250216 | 0.2625339 |
| Q8NCT3 | Q8NCT3 | KIAA0895 | 2.70 | -18.43 | 0.025211 | 0.2625339 |
| Q9Y210-1 | Q9Y210 | TRPC6 | 0.41 | -7.59 | 0.0252571 | 0.2625339 |
| Q9BRG1 | Q9BRG1 | VPS25 | -4.12 | -16.09 | 0.0256127 | 0.2631593 |
| P00736 | P00736 | C1R | -0.45 | -3.16 | 0.0259337 | 0.2631593 |
| Q15828 | Q15828 | CST6 | 3.81 | -15.80 | 0.0259537 | 0.2631593 |
| P02749 | P02749 | APOH | 0.40 | -2.37 | 0.0260777 | 0.2631593 |
| Q06830 | Q06830 | PRDX1 | 0.52 | -5.42 | 0.0261386 | 0.2631593 |
| Q7RTS7 | Q7RTS7 | KRT74 | -5.47 | -14.48 | 0.0261612 | 0.2631593 |
| O00187-1 | O00187 | MASP2 | -0.57 | -7.89 | 0.0264067 | 0.2642085 |
| P07476 | P07476 | IVL | -4.32 | -13.80 | 0.0277288 | 0.2713424 |
| Q6ULP2 | Q6ULP2 | AFTPH | 3.49 | -17.27 | 0.0278024 | 0.2713424 |
| P0CG38 | P0CG38 | POTEI | -3.36 | -18.28 | 0.0279576 | 0.2713424 |
| Q9NQ75-1 | Q9NQ75 | CASS4 | -0.33 | -6.33 | 0.0280326 | 0.2713424 |
| Q86YZ3 | Q86YZ3 | HRNR | -0.57 | -8.06 | 0.0280786 | 0.2713424 |
| P35749-4 | P35749 | MYH11 | -0.70 | -8.46 | 0.0280942 | 0.2713424 |
| P31749 | P31749 | AKT1 | -5.36 | -15.45 | 0.028594 | 0.2713424 |
| P23284 | P23284 | PPIB | 0.66 | -8.13 | 0.0285964 | 0.2713424 |
| Q9NYK6-1 | Q9NYK6 | C21orf91 | 3.68 | -18.29 | 0.0287101 | 0.2713424 |
| P00742 | P00742 | F10 | 0.49 | -6.22 | 0.0287546 | 0.2713424 |
| P61106 | P61106 | RAB14 | -1.13 | -4.93 | 0.02877 | 0.2713424 |
| P14317 | P14317 | HCLS1 | 2.88 | -11.29 | 0.0288885 | 0.2713424 |
| P18859 | P18859 | ATP5PF | 3.71 | -17.69 | 0.0290051 | 0.2713424 |
| P63261 | P63261 | ACTG1 | 0.77 | 0.27 | 0.02957 | 0.2738598 |
| P01871 | P01871 | NA | -0.55 | 0.58 | 0.029792 | 0.2738598 |
| P29972-1 | P29972 | AQP1 | 0.53 | -6.30 | 0.0299413 | 0.2738598 |
| Q8IWA5-3 | Q8IWA5 | SLC44A2 | -0.67 | -7.67 | 0.0299478 | 0.2738598 |
| Q8IZP7 | Q8IZP7 | HS6ST3 | 2.07 | -17.01 | 0.0302061 | 0.2738598 |
| P02655 | P02655 | APOC2 | -0.56 | -0.30 | 0.030233 | 0.2738598 |
| Q15366-1 | Q15366 | PCBP2 | 3.92 | -14.80 | 0.0303749 | 0.2738598 |
| Q6P2E9-1 | Q6P2E9 | EDC4 | -0.39 | -8.77 | 0.030541 | 0.2738598 |
| P35606 | P35606 | COPB2 | -3.43 | -15.13 | 0.030702 | 0.2738598 |
| Q9Y3I1 | Q9Y3I1 | FBXO7 | 3.06 | -17.56 | 0.0307379 | 0.2738598 |
| Q86VP6-1 | Q86VP6 | CAND1 | 0.79 | -6.71 | 0.0310022 | 0.2749059 |
| Q9BTE3-1 | Q9BTE3 | MCMBP | 2.87 | -17.63 | 0.0311947 | 0.2753076 |
| P61204 | P61204 | ARF3 | 0.51 | -6.31 | 0.0313966 | 0.2755728 |
| NA | NA | NA | -2.17 | -19.00 | 0.0315193 | 0.2755728 |
| O60234 | O60234 | GMFG | -0.45 | -6.95 | 0.0319545 | 0.2768076 |
| P67936 | P67936 | TPM4 | 0.75 | -5.58 | 0.0319585 | 0.2768076 |
| P30626-1 | P30626 | SRI | 0.75 | -6.89 | 0.0324249 | 0.2768076 |
| Q07954 | Q07954 | LRP1 | -0.52 | -4.36 | 0.0324582 | 0.2768076 |
| Q9Y4L1 | Q9Y4L1 | HYOU1 | -0.53 | -6.90 | 0.0325153 | 0.2768076 |
| Q16181 | Q16181 | SEPTIN7 | 0.39 | -5.88 | 0.0325482 | 0.2768076 |
| Q6UX06 | Q6UX06 | OLFM4 | 0.71 | -7.80 | 0.0331366 | 0.2805365 |
| P36543 | P36543 | ATP6V1E1 | 2.83 | -18.41 | 0.0333743 | 0.2812759 |
| F5H4B4 | F5H4B4 | FAM227A | 1.82 | -19.35 | 0.0338863 | 0.2843108 |
| Q27J81-2 | Q27J81 | INF2 | -0.50 | -6.66 | 0.0344215 | 0.2875116 |
| Q14761 | Q14761 | PTPRCAP | -3.42 | -13.63 | 0.0350662 | 0.2893589 |
| Q9Y251 | Q9Y251 | HPSE | -1.20 | -8.33 | 0.0352831 | 0.2893589 |
| Q9H299 | Q9H299 | SH3BGRL3 | 0.70 | -8.23 | 0.0353335 | 0.2893589 |
| P05362 | P05362 | ICAM1 | 0.85 | -8.07 | 0.0353783 | 0.2893589 |
| Q15485-1 | Q15485 | FCN2 | -0.93 | -7.65 | 0.0358135 | 0.2893589 |
| P07738 | P07738 | BPGM | 0.66 | -7.41 | 0.036075 | 0.2893589 |
| P09972 | P09972 | ALDOC | 0.71 | -8.37 | 0.0361233 | 0.2893589 |
| Q15691 | Q15691 | MAPRE1 | 0.61 | -8.35 | 0.036911 | 0.2893589 |
| Q9NRY5 | Q9NRY5 | FAM114A2 | 3.65 | -17.32 | 0.0369301 | 0.2893589 |
| O95445-2 | O95445 | APOM | -0.35 | -4.70 | 0.0369834 | 0.2893589 |
| Q9GZX5 | Q9GZX5 | ZNF350 | 3.69 | -17.30 | 0.0370686 | 0.2893589 |
| Q92786 | Q92786 | PROX1 | -4.42 | -14.06 | 0.0370929 | 0.2893589 |
| Q86UX7 | Q86UX7 | FERMT3 | 0.47 | -2.55 | 0.0372928 | 0.2893589 |
| P01768 | P01768 | NA | -0.50 | -6.18 | 0.0375092 | 0.2893589 |
| Q4KMQ2-3 | Q4KMQ2 | ANO6 | -0.99 | -3.63 | 0.0375979 | 0.2893589 |
| P00568 | P00568 | AK1 | -1.88 | -9.65 | 0.0376742 | 0.2893589 |
| P61960 | P61960 | UFM1 | 3.07 | -16.45 | 0.0377625 | 0.2893589 |
| Q08722 | Q08722 | CD47 | 0.51 | -5.70 | 0.0377642 | 0.2893589 |
| P01772 | P01772 | NA | -2.43 | -12.33 | 0.0378083 | 0.2893589 |
| O00453 | O00453 | LST1 | -3.93 | -16.83 | 0.0379186 | 0.2893589 |
| A0A0A0MRZ8 | A0A0A0MRZ8 | NA | -0.47 | -5.22 | 0.0381237 | 0.2893589 |
| P37235 | P37235 | HPCAL1 | 3.18 | -12.47 | 0.0381786 | 0.2893589 |
| P01602 | P01602 | NA | 2.23 | -10.26 | 0.038322 | 0.2893589 |
| Q8NBL1 | Q8NBL1 | POGLUT1 | -3.77 | -17.51 | 0.0383544 | 0.2893589 |
| Q96JG6 | Q96JG6 | VPS50 | -0.45 | -10.45 | 0.0388314 | 0.2911032 |
| P0DOY2 | P0DOY2 | NA | -0.43 | -3.05 | 0.0389908 | 0.2911032 |
| P35908 | P35908 | KRT2 | -0.34 | -1.06 | 0.0390523 | 0.2911032 |
| Q9Y2I6-1 | Q9Y2I6 | NINL | 3.17 | -15.38 | 0.0392337 | 0.2912948 |
| P11277 | P11277 | SPTB | 0.45 | 0.84 | 0.0394581 | 0.2918026 |
| Q8TCD6 | Q8TCD6 | PHOSPHO2 | -4.84 | -14.94 | 0.0396885 | 0.2918432 |
| Q92558 | Q92558 | WASF1 | -2.09 | -18.20 | 0.0397755 | 0.2918432 |
| Q13459-1 | Q13459 | MYO9B | 4.09 | -16.04 | 0.0400818 | 0.2925161 |
| Q9Y5F6-2 | Q9Y5F6 | PCDHGC5 | -3.83 | -14.77 | 0.0401799 | 0.2925161 |
| Q9H8W4 | Q9H8W4 | PLEKHF2 | 3.11 | -17.50 | 0.0404672 | 0.2934656 |
| P12814-1 | P12814 | ACTN1 | 0.53 | -3.56 | 0.0407515 | 0.2943862 |
| Q15084-1 | Q15084 | PDIA6 | -0.46 | -6.41 | 0.0411849 | 0.2963727 |
| O00161 | O00161 | SNAP23 | 0.82 | -7.73 | 0.0416008 | 0.2982191 |
| Q12805 | Q12805 | EFEMP1 | -0.43 | -6.86 | 0.0418013 | 0.2985124 |
| O75396 | O75396 | SEC22B | 2.95 | -15.44 | 0.0425155 | 0.3024583 |
| A0A075B6J1 | A0A075B6J1 | NA | 3.68 | -11.13 | 0.0432196 | 0.3063025 |
| P15170-2 | P15170 | GSPT1 | 3.31 | -16.04 | 0.0436435 | 0.3072236 |
| P01344 | P01344 | IGF2 | 2.22 | -10.80 | 0.0437326 | 0.3072236 |
| P01891 | P01891 | NA | 0.39 | -7.71 | 0.043884 | 0.3072236 |
| P31641 | P31641 | SLC6A6 | -3.03 | -17.36 | 0.0441352 | 0.3072236 |
| Q14974 | Q14974 | KPNB1 | 0.49 | -8.76 | 0.0441706 | 0.3072236 |
| P62820 | P62820 | RAB1A | 2.14 | -7.68 | 0.0446759 | 0.3095877 |
| P48668 | P48668 | KRT6C | -0.82 | -8.76 | 0.0449534 | 0.3103608 |
| P61604 | P61604 | HSPE1 | 2.01 | -9.18 | 0.0452836 | 0.311491 |
| Q5TZA2 | Q5TZA2 | CROCC | -0.36 | -12.97 | 0.0456332 | 0.3127462 |
| Q9UKY7 | Q9UKY7 | CDV3 | -0.44 | -6.56 | 0.0460628 | 0.3145382 |
| Q8NHL6-2 | Q8NHL6 | LILRB1 | -1.69 | -11.34 | 0.0464602 | 0.3160986 |
| P01714 | P01714 | NA | -0.47 | -6.02 | 0.0470606 | 0.3179849 |
| P55209 | P55209 | NAP1L1 | 0.58 | -8.86 | 0.0472579 | 0.3179849 |
| P00387-3 | P00387 | CYB5R3 | -0.47 | -7.01 | 0.0473547 | 0.3179849 |
| P24158 | P24158 | PRTN3 | 0.59 | -8.34 | 0.0474173 | 0.3179849 |
| P01130 | P01130 | LDLR | -0.44 | -6.72 | 0.047623 | 0.3182236 |
| P13639 | P13639 | EEF2 | -0.29 | -6.19 | 0.0482516 | 0.3210166 |
| Q4KWH8-2 | Q4KWH8 | PLCH1 | -0.70 | -6.13 | 0.0483841 | 0.3210166 |
| P29992 | P29992 | GNA11 | 3.22 | -17.78 | 0.0485794 | 0.3211733 |
| Q9BWP8-9 | Q9BWP8 | COLEC11 | -0.78 | -8.63 | 0.0488855 | 0.3220589 |
| P48740-1 | P48740 | MASP1 | -0.33 | -5.02 | 0.0491606 | 0.3227351 |
| P21333-2 | P21333 | FLNA | 0.32 | 0.02 | 0.0495265 | 0.3230834 |

**Supplementary Table ST6. List of differentially expressed EV phosphoproteins: T2DM vs. PDM**

| **Accession** | **UNIPROT** | **SYMBOL** | **logFC** | **AveExpr** | **P.Value** | **adj.P.Val** |
| --- | --- | --- | --- | --- | --- | --- |
| Q5CZC0-1 | Q5CZC0 | FSIP2 | -9.50 | -7.86 | 4.83E-10 | 2.49E-07 |
| Q9Y5S2 | Q9Y5S2 | CDC42BPB | -13.71 | -10.29 | 7.65E-08 | 1.88E-05 |
| P60709 | P60709 | ACTB | -1.85 | -0.96 | 1.09E-07 | 1.88E-05 |
| O00161 | O00161 | SNAP23 | -5.50 | -10.66 | 2.16E-07 | 2.79E-05 |
| P68871 | P68871 | HBB | -1.73 | -0.33 | 2.75E-07 | 2.84E-05 |
| Q7LDG7-4 | Q7LDG7 | RASGRP2 | 9.38 | -11.16 | 1.35E-06 | 0.000116297 |
| Q96Q42-1 | Q96Q42 | ALS2 | 7.57 | -12.31 | 1.90E-06 | 0.000119404 |
| P04792 | P04792 | HSPB1 | -2.85 | -6.23 | 2.07E-06 | 0.000119404 |
| Q01518-1 | Q01518 | CAP1 | -2.60 | -6.37 | 2.08E-06 | 0.000119404 |
| Q9H4B7 | Q9H4B7 | TUBB1 | -8.43 | -5.84 | 2.50E-06 | 0.000128969 |
| P04075 | P04075 | ALDOA | -1.81 | -4.72 | 2.78E-06 | 0.000130346 |
| P98082 | P98082 | DAB2 | 4.52 | -15.30 | 4.99E-06 | 0.000214378 |
| P62070-4 | P62070 | RRAS2 | -5.55 | -10.93 | 9.63E-06 | 0.000382186 |
| E9PAV3 | E9PAV3 | NA | 7.07 | -12.15 | 1.47E-05 | 0.000543115 |
| P20701-2 | P20701 | ITGAL | -7.34 | -13.25 | 1.72E-05 | 0.000546848 |
| A2A3K4-1 | A2A3K4 | PTPDC1 | 5.35 | -8.14 | 1.76E-05 | 0.000546848 |
| Q9Y3X0 | Q9Y3X0 | CCDC9 | 4.26 | -15.13 | 1.80E-05 | 0.000546848 |
| P02751-9 | P02751 | FN1 | 2.25 | 1.26 | 1.95E-05 | 0.000559338 |
| P67936 | P67936 | TPM4 | -3.50 | -9.79 | 2.55E-05 | 0.000692774 |
| P04004 | P04004 | VTN | -1.95 | -5.32 | 3.58E-05 | 0.000922784 |
| Q16204 | Q16204 | CCDC6 | 3.22 | -15.93 | 4.24E-05 | 0.00104144 |
| P04406-1 | P04406 | GAPDH | -2.53 | -4.09 | 4.99E-05 | 0.001141209 |
| Q92508 | Q92508 | PIEZO1 | 1.61 | -4.65 | 5.09E-05 | 0.001141209 |
| P02675 | P02675 | FGB | -3.04 | -1.67 | 5.56E-05 | 0.001195178 |
| P12109 | P12109 | COL6A1 | 7.49 | -11.27 | 6.16E-05 | 0.001271283 |
| P13224 | P13224 | GP1BB | -2.43 | -4.44 | 6.78E-05 | 0.00134459 |
| Q96FS4 | Q96FS4 | SIPA1 | -6.26 | -10.78 | 8.52E-05 | 0.001592772 |
| P06702 | P06702 | S100A9 | 1.38 | -1.69 | 8.64E-05 | 0.001592772 |
| P13646 | P13646 | KRT13 | -3.42 | -6.73 | 0.000134486 | 0.002392916 |
| P62328 | P62328 | TMSB4X | 6.26 | -9.63 | 0.00022916 | 0.003941549 |
| P69905 | P69905 | HBA1 | -1.34 | -2.68 | 0.000279243 | 0.004648044 |
| Q562R1 | Q562R1 | ACTBL2 | 3.76 | -15.79 | 0.000301445 | 0.004786066 |
| O00421 | O00421 | CCRL2 | 5.69 | -13.31 | 0.000306086 | 0.004786066 |
| Q01433-1 | Q01433 | AMPD2 | 5.43 | -13.74 | 0.000330992 | 0.005023288 |
| P31749 | P31749 | AKT1 | 4.62 | -12.51 | 0.000345172 | 0.005088829 |
| P11171-5 | P11171 | EPB41 | 1.42 | -3.08 | 0.000391941 | 0.005600135 |
| Q92530 | Q92530 | PSMF1 | -5.69 | -13.17 | 0.00040156 | 0.005600135 |
| P13645 | P13645 | KRT10 | 2.22 | -2.68 | 0.000416219 | 0.00565182 |
| O00187-1 | O00187 | MASP2 | 1.32 | -6.07 | 0.00055912 | 0.00739759 |
| Q96TC7 | Q96TC7 | RMDN3 | -5.37 | -11.32 | 0.000614359 | 0.007925228 |
| P29692-2 | P29692 | EEF1D | -1.45 | -6.73 | 0.000663379 | 0.008348869 |
| P19823 | P19823 | ITIH2 | -1.83 | -4.91 | 0.000772979 | 0.009496595 |
| Q01082-1 | Q01082 | SPTBN1 | 4.47 | -12.56 | 0.000824567 | 0.009838254 |
| P50502 | P50502 | ST13 | 1.14 | -3.57 | 0.000838921 | 0.009838254 |
| Q16512-1 | Q16512 | PKN1 | -4.04 | -10.31 | 0.000867534 | 0.009947718 |
| Q9NZH0-2 | Q9NZH0 | GPRC5B | 1.08 | -6.85 | 0.00091235 | 0.010025639 |
| O95183 | O95183 | VAMP5 | 4.00 | -14.92 | 0.000913188 | 0.010025639 |
| P62158 | P62158 | NA | 1.29 | -8.10 | 0.000944636 | 0.010154835 |
| Q92539 | Q92539 | LPIN2 | 3.15 | -15.95 | 0.001179188 | 0.012022453 |
| Q14644 | Q14644 | RASA3 | 1.58 | -5.48 | 0.001203496 | 0.012022453 |
| P10412 | P10412 | H1-4 | -5.63 | -14.27 | 0.001211565 | 0.012022453 |
| Q08357 | Q08357 | SLC20A2 | 3.89 | -14.93 | 0.001268607 | 0.012350965 |
| P23588 | P23588 | EIF4B | 3.51 | -12.66 | 0.001343284 | 0.012835827 |
| Q14761 | Q14761 | PTPRCAP | -3.53 | -15.24 | 0.001436345 | 0.013475526 |
| Q13404 | Q13404 | UBE2V1 | 3.37 | -15.90 | 0.001481207 | 0.013648269 |
| Q9H7E9-2 | Q9H7E9 | C8orf33 | -4.38 | -11.76 | 0.001694464 | 0.01533936 |
| O15231-3 | O15231 | ZNF185 | 4.06 | -13.11 | 0.001749301 | 0.015562745 |
| P16298 | P16298 | PPP3CB | 2.98 | -15.65 | 0.001915926 | 0.016615328 |
| P22466 | P22466 | GAL | -4.77 | -12.83 | 0.001932015 | 0.016615328 |
| P19878 | P19878 | NCF2 | 4.37 | -14.05 | 0.002161196 | 0.018281589 |
| P02748 | P02748 | C9 | -1.11 | -6.53 | 0.002512981 | 0.020668515 |
| O00159-1 | O00159 | MYO1C | 3.33 | -13.90 | 0.002523482 | 0.020668515 |
| P35612-1 | P35612 | ADD2 | 1.45 | -2.66 | 0.00269567 | 0.021399472 |
| Q8NBI5 | Q8NBI5 | SLC43A3 | 4.60 | -14.88 | 0.003463928 | 0.027081618 |
| Q2M2I8-1 | Q2M2I8 | AAK1 | 3.52 | -11.84 | 0.003631976 | 0.027560286 |
| Q9UJ41-1 | Q9UJ41 | RABGEF1 | 3.88 | -13.04 | 0.003860729 | 0.028732869 |
| Q9UJZ1 | Q9UJZ1 | STOML2 | 3.90 | -13.80 | 0.00389787 | 0.028732869 |
| Q8TEQ6 | Q8TEQ6 | GEMIN5 | 3.06 | -15.18 | 0.003957796 | 0.028763697 |
| P01031 | P01031 | C5 | -3.51 | -9.94 | 0.004108075 | 0.029441206 |
| Q99808 | Q99808 | SLC29A1 | 1.23 | -5.90 | 0.004212088 | 0.029773116 |
| P32246 | P32246 | CCR1 | -2.36 | -8.91 | 0.004416828 | 0.03008701 |
| P02656 | P02656 | APOC3 | -1.22 | -1.92 | 0.004431089 | 0.03008701 |
| P13861 | P13861 | PRKAR2A | 4.05 | -13.81 | 0.004463507 | 0.03008701 |
| Q96CV9-1 | Q96CV9 | OPTN | -4.49 | -15.24 | 0.004548036 | 0.03008701 |
| Q9Y490 | Q9Y490 | TLN1 | 1.25 | -4.44 | 0.005033225 | 0.03237177 |
| P51858-3 | P51858 | HDGF | -1.15 | -7.61 | 0.00510194 | 0.03237177 |
| Q5JSH3 | Q5JSH3 | WDR44 | 1.18 | -3.54 | 0.005144351 | 0.03237177 |
| P55196-1 | P55196 | AFDN | -3.34 | -12.80 | 0.005331907 | 0.033147757 |
| Q5M775-1 | Q5M775 | SPECC1 | 3.73 | -8.81 | 0.005796983 | 0.034839838 |
| P11836 | P11836 | MS4A1 | -1.20 | -7.16 | 0.006091115 | 0.036126616 |
| Q8TEW0 | Q8TEW0 | PARD3 | 4.16 | -11.63 | 0.006297052 | 0.036508752 |
| Q9Y3P9 | Q9Y3P9 | RABGAP1 | 3.35 | -13.40 | 0.00648295 | 0.037168911 |
| P22392-2 | P22392 | NME2 | 3.05 | -9.39 | 0.006557168 | 0.037181306 |
| Q13442 | Q13442 | PDAP1 | 3.69 | -13.04 | 0.007107663 | 0.039864718 |
| P0C7M8 | P0C7M8 | CLEC2L | 4.07 | -13.29 | 0.007969409 | 0.043286476 |
| Q7RTP6-5 | Q7RTP6 | MICAL3 | -1.02 | -6.47 | 0.008106992 | 0.043575083 |
| P59768 | P59768 | GNG2 | 2.76 | -8.62 | 0.008776364 | 0.046686638 |
| Q8IZF2 | Q8IZF2 | ADGRF5 | -3.62 | -14.76 | 0.008916428 | 0.046947721 |
| Q9UHY1 | Q9UHY1 | NRBP1 | 1.22 | -6.95 | 0.009223056 | 0.047753293 |
| Q9BSQ5-1 | Q9BSQ5 | CCM2 | -4.04 | -13.94 | 0.009372679 | 0.047753293 |
| P16949-1 | P16949 | STMN1 | -3.45 | -11.04 | 0.009410258 | 0.047753293 |
| P55198 | P55198 | MLLT6 | -3.07 | -14.43 | 0.009439604 | 0.047753293 |
| P23142 | P23142 | FBLN1 | -1.51 | -6.95 | 0.010054511 | 0.049741434 |
| O60232 | O60232 | ZNRD2 | -5.07 | -11.96 | 0.010121804 | 0.049741434 |

**Supplementary Table ST7. List of significant correlation between DE EV phosphoproteins and clinical measures**

| **var1** | **var2** | **cor** | **p.value** | **FDR** |
| --- | --- | --- | --- | --- |
| AAK1 | auc_glucose | 0.60 | 4.43E-04 | 0.0197786 |
| AAK1 | AIRg | -0.62 | 0.000837 | 0.0279025 |
| AAK1 | Glucose_bsl | 0.50 | 0.0051823 | 0.0829547 |
| AAK1 | DI | -0.48 | 0.0146152 | 0.1562258 |
| AAK1 | TSH | 0.42 | 0.0204295 | 0.1873728 |
| AAK1 | HbA1C | 0.36 | 0.0498498 | 0.3044623 |
| ABCF1 | auc_glucose | -0.79 | 2.144E-07 | 0.0004622 |
| ABCF1 | HbA1C | -0.66 | 7.027E-05 | 0.0063376 |
| ABCF1 | Glucose_bsl | -0.63 | 0.0002107 | 0.0129786 |
| ABCF1 | AIRg | 0.51 | 0.0086845 | 0.1129838 |
| ABCF1 | DI | 0.47 | 0.0175012 | 0.1701959 |
| ABCF1 | Sg | 0.41 | 0.0428862 | 0.2809977 |
| ABLIM1 | auc_glucose | 0.53 | 0.0025137 | 0.0543498 |
| ABLIM1 | Glucose_bsl | 0.50 | 0.0050979 | 0.0820098 |
| ABLIM1 | DEXA__Fat_Mass | 0.39 | 0.031562 | 0.2360319 |
| ABLIM1 | Waist_Circumference | 0.37 | 0.0412257 | 0.2759899 |
| ACTB | auc_glucose | -0.61 | 0.0003379 | 0.01694 |
| ACTB | Glucose_bsl | -0.52 | 0.0035077 | 0.0659425 |
| ACTB | HOMA_B | 0.49 | 0.0056497 | 0.0867166 |
| ACTB | auc_INSULIN | 0.48 | 0.0070407 | 0.1002907 |
| ACTB | Sg | 0.52 | 0.0072855 | 0.1015412 |
| ACTB | AIRg | 0.46 | 0.0215503 | 0.1924944 |
| ACTB | Si | -0.45 | 2.52E-02 | 0.209708 |
| ACTB | HbA1C | -0.37 | 0.0441965 | 0.2863918 |
| ACTBL2 | auc_glucose | 0.46 | 0.011251 | 0.132532 |
| ACTBL2 | TSH | 0.43 | 0.0171129 | 0.168626 |
| ACTBL2 | HOMA_B | -0.39 | 0.034293 | 0.2484036 |
| ADCY6 | auc_glucose | 0.67 | 5.174E-05 | 0.0053971 |
| ADCY6 | Glucose_bsl | 0.62 | 0.0002777 | 0.0148435 |
| ADCY6 | HbA1C | 0.50 | 0.0045423 | 0.0761007 |
| ADCY6 | Respiration_Rate | 0.40 | 0.0332049 | 0.2423661 |
| ADCY6 | HOMA_B | -0.36 | 0.0479931 | 0.2987215 |
| ADD1 | AIRg | -0.51 | 9.15E-03 | 0.1175981 |
| ADD1 | auc_glucose | 0.45 | 0.0134962 | 0.1497086 |
| ADD1 | DEXA__Lean_Mass | -0.41 | 0.0240583 | 0.2025844 |
| ADD1 | Glucose_bsl | 0.38 | 0.0409144 | 0.2751949 |
| ADD1 | TSH | 0.37 | 0.0439771 | 0.2852555 |
| ADD1 | DEXA_Weight | -0.36 | 0.0487384 | 0.3016187 |
| ADD2 | auc_glucose | 0.54 | 0.0020512 | 0.0479593 |
| ADD2 | Glucose_bsl | 0.48 | 0.0078072 | 0.105626 |
| ADD2 | HOMA_B | -0.44 | 0.0156477 | 0.162169 |
| ADD2 | AIRg | -0.47 | 0.0183359 | 0.1756711 |
| ADD2 | auc_INSULIN | -0.37 | 0.0444604 | 0.2872381 |
| AFDN | DEXA__Fat_Mass | 0.36 | 0.0482926 | 0.2991461 |
| AKNAD1 | auc_glucose | -0.57 | 0.0009143 | 0.0292369 |
| AKNAD1 | Glucose_bsl | -0.51 | 0.0040606 | 0.0711643 |
| AKNAD1 | HbA1C | -0.49 | 0.0056741 | 0.0867166 |
| AKNAD1 | Waist_Circumference | -0.43 | 0.0183239 | 0.1756711 |
| AKNAD1 | TSH | -0.41 | 0.0261085 | 0.2138998 |
| AKNAD1 | DEXA__Fat_Mass | -0.41 | 0.0262465 | 0.2145845 |
| AKT1 | auc_glucose | 0.59 | 0.0005649 | 0.0230088 |
| AKT1 | AIRg | -0.60 | 0.0015236 | 0.039093 |
| AKT1 | Glucose_bsl | 0.48 | 0.0072017 | 0.1014677 |
| AKT1 | DI | -0.45 | 0.0234794 | 0.2009239 |
| AKT1 | Sg | -0.45 | 0.0244244 | 0.2054001 |
| AKT1 | HOMA_B | -0.39 | 0.0329855 | 0.2413089 |
| AKT1 | HbA1C | 0.37 | 0.0446435 | 0.2881332 |
| ALDOA | auc_glucose | -0.49 | 0.0055278 | 0.0863486 |
| ALDOA | Si | -0.47 | 0.0169068 | 0.1676936 |
| ALDOA | Glucose_bsl | -0.37 | 0.045469 | 0.2908486 |
| ALS2 | HOMA_B | -0.42 | 0.0193725 | 0.1820961 |
| AMPD2 | auc_glucose | 0.67 | 5.65E-05 | 0.0058001 |
| AMPD2 | Glucose_bsl | 0.63 | 0.0001855 | 0.012051 |
| AMPD2 | HbA1C | 0.62 | 0.0002306 | 0.0135549 |
| AMPD2 | DI | -0.43 | 0.0300182 | 0.2300091 |
| AMPD2 | DEXA_Weight | 0.39 | 0.033809 | 0.2459425 |
| ANKS1A | Glucose_bsl | 0.62 | 0.0002974 | 0.0156885 |
| ANKS1A | auc_glucose | 0.59 | 0.0005806 | 0.0230088 |
| ANKS1A | DEXA__Fat_Mass | 0.47 | 0.0094765 | 0.120402 |
| ANKS1A | DEXA_Fat_Percentage | 0.47 | 0.0095491 | 0.1208496 |
| ANKS1A | HbA1C | 0.36 | 0.0491271 | 0.3030258 |
| APEH | HbA1C | -0.59 | 0.0006809 | 0.0250181 |
| APEH | auc_glucose | -0.56 | 0.0012902 | 0.0361205 |
| APEH | Glucose_bsl | -0.54 | 0.002063 | 0.0479917 |
| APEH | Respiration_Rate | -0.44 | 0.0173196 | 0.1691936 |
| APEH | LDL | 0.38 | 0.0434027 | 0.2823797 |
| APEH | DI | 0.41 | 0.0436241 | 0.2835345 |
| APOA1 | auc_glucose | 0.61 | 0.0003897 | 0.0186692 |
| APOA1 | Glucose_bsl | 0.55 | 0.0015211 | 0.039093 |
| APOA1 | HbA1C | 0.54 | 0.0018632 | 0.0447939 |
| APOA1 | Waist_Circumference | 0.53 | 2.68E-03 | 0.0557052 |
| APOA1 | DEXA_Weight | 0.45 | 0.0121174 | 0.1394364 |
| APOA1 | DEXA__Fat_Mass | 0.42 | 0.02185 | 0.1943658 |
| APOA1 | HDL | -0.40 | 0.027754 | 0.2210408 |
| APOA1 | HOMA_IR | 0.37 | 0.0471528 | 0.2957311 |
| APOBR | auc_glucose | -0.51 | 0.003715 | 0.0672968 |
| APOBR | Sg | 0.47 | 0.0166895 | 0.1666672 |
| APOBR | Glucose_bsl | -0.40 | 0.0288149 | 0.2253275 |
| APOBR | AIRg | 0.44 | 0.0294907 | 0.2278573 |
| APOBR | HbA1C | -0.40 | 0.0305931 | 0.2318537 |
| APOBR | DI | 0.41 | 0.0425788 | 0.2799815 |
| APOC3 | AIRg | 0.75 | 1.544E-05 | 0.0024361 |
| APOC3 | auc_glucose | -0.69 | 2.655E-05 | 0.0037206 |
| APOC3 | Glucose_bsl | -0.51 | 0.0041891 | 0.0726296 |
| APOC3 | HbA1C | -0.44 | 0.0139804 | 0.1532394 |
| APOC3 | TSH | -0.43 | 0.0164118 | 0.1653198 |
| APOC3 | DI | 0.40 | 0.0452416 | 0.2908321 |
| APOC3 | Ins30_glu30 | 0.37 | 0.0472097 | 0.2957311 |
| ARHGAP30 | auc_glucose | -0.57 | 0.0009492 | 0.0294022 |
| ARHGAP30 | Glucose_bsl | -0.54 | 0.0021273 | 0.0492123 |
| ARHGAP30 | Respiration_Rate | -0.47 | 0.0107361 | 0.1295342 |
| ARHGAP30 | HbA1C | -0.44 | 0.0149159 | 0.1578744 |
| ARHGAP30 | HDL | 0.44 | 0.0152466 | 0.1597001 |
| ARRB1 | auc_glucose | 0.67 | 4.427E-05 | 0.0047618 |
| ARRB1 | Glucose_bsl | 0.58 | 0.0008816 | 0.0289404 |
| ARRB1 | Ins30_glu30 | -0.47 | 0.0092293 | 0.118425 |
| ARRB1 | AIRg | -0.49 | 1.37E-02 | 0.1511292 |
| ARRB1 | HbA1C | 0.43 | 0.0167693 | 0.1666672 |
| ATP2A2 | Glucose_bsl | 0.57 | 0.0009218 | 0.0292369 |
| ATP2A2 | auc_glucose | 0.57 | 0.0011156 | 0.0326423 |
| ATP2A2 | DEXA_Fat_Percentage | 0.47 | 0.0083172 | 0.1099941 |
| ATP2A2 | MATSUDA | -0.43 | 0.018807 | 0.1785978 |
| ATP7A | DI | -0.58 | 0.0026344 | 0.0549571 |
| ATP7A | Sg | -0.54 | 0.0057358 | 0.0872788 |
| ATP7A | auc_glucose | 0.42 | 0.0205713 | 0.1873728 |
| ATP7A | Heart_Rate_Average | -0.39 | 0.0337829 | 0.2459425 |
| ATP7A | HbA1C | 0.38 | 0.0377081 | 0.2616508 |
| C12orf75 | Glucose_bsl | 0.53 | 0.0028852 | 0.0577668 |
| C12orf75 | DEXA_Fat_Percentage | 0.44 | 0.0142565 | 0.1539974 |
| C12orf75 | auc_glucose | 0.39 | 0.0351094 | 0.251164 |
| C1orf56 | auc_glucose | -0.55 | 0.0017361 | 0.0428533 |
| C1orf56 | Glucose_bsl | -0.42 | 0.0195208 | 0.1829576 |
| C4BPA | auc_glucose | -0.59 | 0.0005444 | 0.0225687 |
| C4BPA | Glucose_bsl | -0.58 | 0.0008248 | 0.0276361 |
| C4BPA | AIRg | 0.56 | 0.0036804 | 0.0670571 |
| C4BPA | Respiration_Rate | -0.49 | 0.0065884 | 0.0961788 |
| C4BPA | HbA1C | -0.45 | 0.0119409 | 0.1381423 |
| C5 | auc_glucose | -0.75 | 1.849E-06 | 0.0009199 |
| C5 | HbA1C | -0.68 | 3.816E-05 | 0.004407 |
| C5 | Glucose_bsl | -0.66 | 7.366E-05 | 0.0065178 |
| C5 | DI | 0.51 | 0.0095067 | 0.1205486 |
| C5 | AIRg | 0.42 | 0.034301 | 0.2484036 |
| C5 | Respiration_Rate | -0.39 | 0.0375279 | 0.2612506 |
| C5 | HOMA_IR | -0.38 | 0.0392611 | 0.2696004 |
| C8orf33 | auc_glucose | -0.70 | 1.832E-05 | 0.0026955 |
| C8orf33 | DI | 0.68 | 0.0001977 | 0.0124134 |
| C8orf33 | HbA1C | -0.62 | 0.0002332 | 0.0135845 |
| C8orf33 | Glucose_bsl | -0.59 | 0.0006713 | 0.0248071 |
| C8orf33 | HOMA_IR | -0.41 | 0.0235497 | 0.2009239 |
| C8orf33 | DEXA__Fat_Mass | -0.36 | 0.0482696 | 0.2991461 |
| C9 | auc_glucose | -0.73 | 4.084E-06 | 0.0013901 |
| C9 | Glucose_bsl | -0.64 | 0.000143 | 0.0103884 |
| C9 | HbA1C | -0.49 | 0.0055567 | 0.0865911 |
| C9 | TSH | -0.47 | 0.0093845 | 0.1194679 |
| C9 | AIRg | 0.45 | 0.0255793 | 0.211111 |
| C9 | HOMA_B | 0.38 | 0.0366328 | 0.2577573 |
| CAMSAP2 | QUICKI | 0.48 | 0.0074604 | 0.1026104 |
| CAMSAP2 | Waist_Circumference | -0.41 | 0.0234177 | 0.2009239 |
| CAMSAP2 | HbA1C | -0.37 | 0.0431834 | 0.2817601 |
| CAP1 | auc_glucose | -0.44 | 0.0140976 | 0.1537424 |
| CAP1 | MATSUDA | -0.44 | 0.0143107 | 0.1539974 |
| CAP1 | Si | -0.41 | 0.0395055 | 0.269893 |
| CCAR2 | DEXA__Fat_Mass | -0.48 | 0.0072746 | 0.1015412 |
| CCAR2 | auc_glucose | -0.41 | 0.0257695 | 0.2117553 |
| CCAR2 | Glucose_bsl | -0.37 | 0.0417726 | 0.2779185 |
| CCDC6 | auc_c_peptide | -0.69 | 0.0022304 | 0.0506115 |
| CCDC6 | auc_glucose | 0.45 | 0.0122499 | 0.1402127 |
| CCDC6 | QUICKI | 0.44 | 0.0147783 | 0.1573502 |
| CCDC6 | DI | -0.44 | 0.0260233 | 0.2135691 |
| CCDC6 | MATSUDA | 0.40 | 0.0266291 | 0.2154369 |
| CCDC6 | HbA1C | 0.39 | 0.0325055 | 0.2390041 |
| CCDC6 | Sg | -0.43 | 0.0330757 | 0.2416954 |
| CCDC9 | auc_glucose | 0.68 | 3.53E-05 | 0.0042274 |
| CCDC9 | Glucose_bsl | 0.58 | 7.43E-04 | 0.0263107 |
| CCDC9 | HbA1C | 0.58 | 0.0007688 | 0.0267522 |
| CCDC9 | DI | -0.62 | 0.0009223 | 0.0292369 |
| CCDC9 | AIRg | -0.59 | 0.0020468 | 0.0479593 |
| CCDC9 | HOMA_B | -0.42 | 0.0209026 | 0.1898552 |
| CCDC9 | auc_INSULIN | -0.37 | 0.0425876 | 0.2799815 |
| CCM2 | TSH | -0.59 | 0.0006071 | 0.0232318 |
| CCM2 | auc_glucose | -0.50 | 0.0046629 | 0.0776693 |
| CCM2 | HOMA_B | 0.46 | 0.0108001 | 0.1296972 |
| CCM2 | Glucose_bsl | -0.42 | 0.0196839 | 0.183688 |
| CCM2 | Ins30_glu30 | 0.42 | 0.0204738 | 0.1873728 |
| CCM2 | auc_INSULIN | 0.37 | 0.0427244 | 0.2802216 |
| CCM2 | AIRg | 0.40 | 0.0467453 | 0.2949723 |
| CCR1 | Glucose_bsl | -0.42 | 0.0195882 | 0.1833241 |
| CCR1 | Diastolic_Blood_Pressure_Average | -0.40 | 0.030408 | 0.2313511 |
| CCR1 | Temperature | -0.40 | 0.040528 | 0.2738708 |
| CCR1 | auc_glucose | -0.37 | 0.043304 | 0.2820212 |
| CCRL2 | auc_glucose | 0.41 | 0.0254063 | 0.211111 |
| CCRL2 | Glucose_bsl | 0.40 | 0.0266615 | 0.2154369 |
| CCRL2 | HbA1C | 0.40 | 0.0277887 | 0.2210453 |
| CCRL2 | auc_c_peptide | -0.49 | 0.0454324 | 0.2908486 |
| CDC42BPB | auc_glucose | -0.53 | 0.0027664 | 0.0566055 |
| CDC42BPB | HbA1C | -0.42 | 0.0213155 | 0.1919878 |
| CDC42BPB | Glucose_bsl | -0.37 | 0.0458594 | 0.2921898 |
| CEP162 | Waist_Circumference | 0.48 | 0.0074059 | 0.1023382 |
| CEP162 | Temperature | -0.50 | 0.0093277 | 0.1191674 |
| CEP162 | QUICKI | -0.42 | 2.23E-02 | 0.1958493 |
| CEP162 | HDL | -0.38 | 0.0403496 | 0.2735231 |
| CFH | auc_glucose | -0.52 | 0.0033541 | 0.0641738 |
| CFH | Respiration_Rate | -0.50 | 0.0056615 | 0.0867166 |
| CFH | HbA1C | -0.46 | 0.0108345 | 0.1296972 |
| CFH | AIRg | 0.48 | 0.015822 | 0.1629318 |
| CFH | Glucose_bsl | -0.44 | 0.0160258 | 0.1642362 |
| CFH | DI | 0.46 | 0.0219601 | 0.1948093 |
| CFL1 | Glucose_bsl | 0.68 | 2.984E-05 | 0.0038333 |
| CFL1 | auc_glucose | 0.68 | 4.107E-05 | 0.0045987 |
| CFL1 | HbA1C | 0.52 | 0.0030455 | 0.0600469 |
| CFL1 | Respiration_Rate | 0.51 | 0.0049644 | 0.0808677 |
| CLEC2L | auc_glucose | 0.55 | 0.0016094 | 0.0408159 |
| CLEC2L | DI | -0.56 | 0.0036366 | 0.0670571 |
| CLEC2L | HbA1C | 0.45 | 0.0119063 | 0.1380642 |
| CLEC2L | Glucose_bsl | 0.40 | 0.0281033 | 0.222141 |
| CLEC2L | Respiration_Rate | 0.40 | 0.0335325 | 0.2444814 |
| CLU | AIRg | 0.60 | 0.0014876 | 0.0385762 |
| CLU | auc_glucose | -0.52 | 0.003468 | 0.0657691 |
| CLU | TSH | -0.49 | 0.0060498 | 0.0904355 |
| CLU | HOMA_B | 0.47 | 0.0084658 | 0.111051 |
| CLU | Glucose_bsl | -0.44 | 0.0156962 | 0.1624119 |
| CLU | auc_INSULIN | 0.43 | 0.0172772 | 0.1690431 |
| COL6A1 | HDL | 0.48 | 0.0067749 | 0.0982364 |
| COL6A1 | QUICKI | 0.48 | 0.0075716 | 0.1033029 |
| COL6A1 | DEXA__Fat_Mass | -0.44 | 0.0162517 | 0.164475 |
| COL6A1 | MATSUDA | 0.43 | 0.0191102 | 0.180417 |
| COL6A1 | Heart_Rate_Average | -0.42 | 0.0197162 | 0.1837246 |
| COL6A1 | Waist_Circumference | -0.41 | 0.0237543 | 0.2016001 |
| CP | auc_glucose | -0.57 | 0.0009502 | 0.0294022 |
| CP | AIRg | 0.55 | 0.0044878 | 0.0757767 |
| CP | Glucose_bsl | -0.50 | 0.0047857 | 0.0787518 |
| CP | HbA1C | -0.43 | 0.018242 | 0.1755517 |
| CTDSPL | Glucose_bsl | 0.70 | 1.834E-05 | 0.0026955 |
| CTDSPL | auc_glucose | 0.66 | 7.458E-05 | 0.0065178 |
| CTDSPL | HbA1C | 0.53 | 0.0025594 | 0.0544573 |
| CTDSPL | HOMA_B | -0.43 | 0.0172781 | 0.1690431 |
| CTDSPL | AIRg | -0.41 | 0.044393 | 0.2870892 |
| CTTN | auc_glucose | 0.58 | 0.000805 | 0.0272549 |
| CTTN | HbA1C | 0.54 | 0.0020542 | 0.0479593 |
| CTTN | Glucose_bsl | 0.47 | 0.0092492 | 0.118445 |
| CTTN | Waist_Circumference | 0.41 | 0.0248762 | 0.20758 |
| CTTN | HOMA_IR | 0.39 | 0.0342286 | 0.2484036 |
| DAAM1 | Glucose_bsl | 0.63 | 0.0001681 | 0.0114449 |
| DAAM1 | auc_glucose | 0.57 | 0.000973 | 0.0298214 |
| DAAM1 | HOMA_B | -0.45 | 0.0123246 | 0.1403227 |
| DAAM1 | HbA1C | 0.40 | 0.0264532 | 0.2150698 |
| DAB2 | auc_glucose | 0.70 | 1.532E-05 | 0.0024361 |
| DAB2 | HbA1C | 0.59 | 0.0005713 | 0.0230088 |
| DAB2 | Glucose_bsl | 0.59 | 0.000645 | 0.0240484 |
| DAB2 | DI | -0.59 | 0.0018873 | 0.045205 |
| DAB2 | AIRg | -0.54 | 0.0056395 | 0.0867166 |
| DAB2 | auc_c_peptide | -0.60 | 0.0114298 | 0.1342239 |
| DAB2 | HOMA_B | -0.41 | 0.0230413 | 0.2000108 |
| DAB2 | Sg | -0.44 | 0.0263692 | 0.2150436 |
| DENND1A | auc_INSULIN | 0.38 | 0.037863 | 0.262444 |
| DENND1A | DEXA__Fat_Mass | 0.36 | 0.0488922 | 0.3022808 |
| DENND4A | auc_glucose | 0.45 | 0.0128214 | 0.1445307 |
| DENND4A | AIRg | -0.48 | 0.0143108 | 0.1539974 |
| DENND4A | HOMA_B | -0.43 | 0.0183636 | 0.1756768 |
| DENND4A | DEXA__Lean_Mass | -0.42 | 0.0198748 | 0.1846703 |
| DENND4A | TSH | 0.39 | 0.0319862 | 0.2371436 |
| DENND4A | DEXA_Weight | -0.39 | 0.0350389 | 0.2509376 |
| DENND4A | Glucose_bsl | 0.38 | 0.0406681 | 0.274531 |
| DMTN | auc_glucose | 0.71 | 1.281E-05 | 0.0022396 |
| DMTN | Glucose_bsl | 0.61 | 0.0003207 | 0.0162879 |
| DMTN | AIRg | -0.55 | 0.0042776 | 0.0733781 |
| DMTN | HbA1C | 0.45 | 1.34E-02 | 0.1490214 |
| DMTN | HOMA_B | -0.40 | 0.0306533 | 0.2318537 |
| DMTN | TSH | 0.38 | 0.0394252 | 0.269893 |
| DOCK10 | auc_glucose | 0.55 | 0.0014686 | 0.0383746 |
| DOCK10 | Glucose_bsl | 0.48 | 0.0077334 | 0.1052126 |
| DOCK10 | HbA1C | 0.41 | 0.0262936 | 0.2146979 |
| DSP | DEXA__Fat_Mass | -0.40 | 0.027302 | 0.2187885 |
| ECM1 | auc_glucose | -0.55 | 0.0016386 | 0.0412331 |
| ECM1 | Glucose_bsl | -0.46 | 0.0105618 | 0.1279081 |
| ECM1 | DEXA__Fat_Mass | -0.41 | 0.0255252 | 0.211111 |
| ECM1 | HbA1C | -0.38 | 0.0386283 | 0.2660376 |
| ECM1 | Waist_Circumference | -0.37 | 0.0465307 | 0.2944367 |
| ECM2 | auc_glucose | -0.64 | 0.0001326 | 0.0097423 |
| ECM2 | Glucose_bsl | -0.53 | 0.002456 | 0.0541766 |
| ECM2 | HbA1C | -0.49 | 0.006066 | 0.0904355 |
| ECM2 | DI | 0.40 | 0.0452052 | 0.2908321 |
| EDAR | Sg | -0.58 | 0.0024845 | 0.0542821 |
| EDAR | HOMA_B | -0.44 | 0.0148177 | 0.1573502 |
| EDAR | auc_c_peptide | -0.56 | 0.0205096 | 0.1873728 |
| EDAR | AIRg | -0.44 | 0.0283398 | 0.2229606 |
| EDAR | auc_glucose | 0.39 | 0.0349021 | 0.2507694 |
| EDAR | Insulin_bsl | -0.38 | 0.0403468 | 0.2735231 |
| EDAR | DI | -0.41 | 0.0410137 | 0.2751949 |
| EEF1D | auc_glucose | -0.74 | 3.727E-06 | 0.0013392 |
| EEF1D | Glucose_bsl | -0.69 | 2.762E-05 | 0.0037206 |
| EEF1D | HbA1C | -0.54 | 0.0022559 | 0.0510101 |
| EEF1D | HOMA_B | 0.45 | 0.0136352 | 0.1507327 |
| EEF1D | AIRg | 0.42 | 0.0369112 | 0.2588991 |
| EEF1D | auc_INSULIN | 0.37 | 0.045394 | 0.2908486 |
| EFR3A | auc_glucose | 0.56 | 0.0012463 | 0.0351959 |
| EFR3A | DEXA__Lean_Mass | 0.51 | 0.0036313 | 0.0670571 |
| EFR3A | DEXA_Fat_Percentage | -0.47 | 0.0082159 | 0.1095507 |
| EFR3A | HbA1C | 0.44 | 0.0143067 | 0.1539974 |
| EFR3A | Glucose_bsl | 0.39 | 0.0320601 | 0.2371436 |
| EFR3A | DEXA_Weight | 0.39 | 0.0354735 | 0.252589 |
| EIF4B | DI | -0.65 | 0.0004237 | 0.0192958 |
| EIF4B | AIRg | -0.64 | 0.0005131 | 0.0218314 |
| EIF4B | Sg | -0.56 | 0.003681 | 0.0670571 |
| EIF4B | auc_glucose | 0.46 | 0.0104658 | 0.127599 |
| EIF4B | HbA1C | 0.44 | 0.0150513 | 0.1589903 |
| EIF4B | Glucose_bsl | 0.39 | 0.0318318 | 0.2371436 |
| EIF4B | auc_INSULIN | -0.38 | 0.0391105 | 0.269072 |
| EIF4G3 | auc_glucose | 0.61 | 0.000381 | 0.0185256 |
| EIF4G3 | Glucose_bsl | 0.56 | 0.0011626 | 0.0335653 |
| EIF4G3 | HbA1C | 0.53 | 2.59E-03 | 0.0544573 |
| EIF5B | Glucose_bsl | 0.60 | 0.000434 | 0.0194929 |
| EIF5B | auc_glucose | 0.57 | 0.0009036 | 0.0292369 |
| EIF5B | HOMA_B | -0.53 | 0.0026952 | 0.0557052 |
| EIF5B | auc_c_peptide | -0.60 | 0.0110745 | 0.1311702 |
| EIF5B | AIRg | -0.49 | 0.0138522 | 0.1520923 |
| EIF5B | Ins30_glu30 | -0.43 | 0.0177216 | 0.1715648 |
| EIF5B | DEXA_Fat_Percentage | 0.38 | 0.0362831 | 0.2567209 |
| ELMO1 | auc_glucose | 0.56 | 0.0013981 | 0.0375155 |
| ELMO1 | Glucose_bsl | 0.49 | 0.0063822 | 0.0942328 |
| ELMO1 | HOMA_B | -0.46 | 0.0105339 | 0.1279081 |
| ELMO1 | AIRg | -0.48 | 0.0150705 | 0.1589903 |
| EPB41 | auc_glucose | 0.65 | 0.000109 | 0.0085941 |
| EPB41 | Glucose_bsl | 0.53 | 0.0026961 | 0.0557052 |
| EPB41 | AIRg | -0.57 | 0.0029035 | 0.0579541 |
| EPB41 | HbA1C | 0.40 | 0.0289582 | 0.2259022 |
| EPB41 | TSH | 0.37 | 0.0463557 | 0.2941926 |
| EPS15 | auc_glucose | 0.61 | 0.0003643 | 0.0178803 |
| EPS15 | Glucose_bsl | 0.59 | 0.000647 | 0.0240484 |
| EPS15 | AIRg | -0.46 | 0.0200143 | 0.1855599 |
| F11R | auc_glucose | 0.46 | 0.0110394 | 0.1309947 |
| F11R | TSH | 0.45 | 0.0116448 | 0.1361792 |
| F11R | Glucose_bsl | 0.43 | 0.0167581 | 0.1666672 |
| F11R | auc_c_peptide | -0.53 | 0.0283345 | 0.2229606 |
| F11R | Heart_Rate_Average | 0.37 | 0.047284 | 0.2957311 |
| F2RL3 | Glucose_bsl | 0.66 | 7.668E-05 | 0.0065245 |
| F2RL3 | auc_glucose | 0.65 | 0.0001124 | 0.0087578 |
| F2RL3 | HbA1C | 0.48 | 0.0070969 | 0.1007201 |
| F2RL3 | Heart_Rate_Average | 0.40 | 0.0267172 | 0.2154369 |
| F5 | auc_glucose | -0.71 | 1.023E-05 | 0.0019546 |
| F5 | Glucose_bsl | -0.58 | 0.0006926 | 0.0253069 |
| F5 | AIRg | 0.63 | 0.0007694 | 0.0267522 |
| F5 | HbA1C | -0.54 | 0.0021694 | 0.0497512 |
| F5 | DI | 0.49 | 0.0119728 | 0.1381569 |
| FBLN1 | Glucose_bsl | -0.75 | 2.167E-06 | 0.0010011 |
| FBLN1 | auc_glucose | -0.71 | 1.15E-05 | 0.0021242 |
| FBLN1 | HbA1C | -0.53 | 0.0028269 | 0.0568183 |
| FBLN1 | HOMA_IR | -0.41 | 0.0246772 | 0.2072276 |
| FCER1G | auc_glucose | 0.74 | 2.891E-06 | 0.001205 |
| FCER1G | Glucose_bsl | 0.71 | 1.028E-05 | 0.0019546 |
| FCER1G | HbA1C | 0.55 | 0.0018413 | 0.0444307 |
| FCER1G | Respiration_Rate | 0.48 | 0.0081829 | 0.1093368 |
| FCER1G | HDL | -0.39 | 0.0313485 | 0.2351862 |
| FCER1G | MATSUDA | -0.39 | 0.0314986 | 0.2360319 |
| FCER1G | AIRg | -0.43 | 0.0323484 | 0.2388097 |
| FCHO2 | DEXA__Fat_Mass | 0.41 | 0.0227186 | 0.1984087 |
| FCHO2 | TSH | 0.41 | 0.0227647 | 0.1984087 |
| FCHO2 | Heart_Rate_Average | 0.40 | 0.0298319 | 0.2288526 |
| FCHO2 | DI | -0.40 | 0.0469313 | 0.2953532 |
| FERMT3 | AIRg | 0.70 | 8.901E-05 | 0.0071955 |
| FERMT3 | auc_glucose | -0.49 | 0.0056855 | 0.0867166 |
| FERMT3 | Glucose_bsl | -0.47 | 0.0085534 | 0.1118136 |
| FERMT3 | HOMA_B | 0.44 | 0.0159587 | 0.1638173 |
| FERMT3 | auc_INSULIN | 0.42 | 0.0194699 | 0.1827459 |
| FGA | auc_glucose | -0.57 | 0.0009591 | 0.0295361 |
| FGA | Glucose_bsl | -0.53 | 0.0024169 | 0.0535285 |
| FGB | auc_glucose | -0.75 | 1.525E-06 | 0.0008219 |
| FGB | Glucose_bsl | -0.72 | 7.463E-06 | 0.0017357 |
| FGB | HOMA_B | 0.59 | 0.0006243 | 0.0235948 |
| FGB | HbA1C | -0.51 | 0.0042256 | 0.0728724 |
| FGB | auc_INSULIN | 0.46 | 0.0110224 | 0.1309947 |
| FGD3 | auc_glucose | 0.66 | 7.056E-05 | 0.0063376 |
| FGD3 | Glucose_bsl | 0.60 | 0.0004531 | 0.0199337 |
| FGD3 | HbA1C | 0.44 | 0.0141358 | 0.1538989 |
| FGG | auc_glucose | -0.60 | 0.0005099 | 0.0218314 |
| FGG | Glucose_bsl | -0.55 | 0.0015492 | 0.0394427 |
| FGG | HbA1C | -0.41 | 0.0256604 | 0.2113375 |
| FGG | HOMA_IR | -0.38 | 0.0404056 | 0.2736157 |
| FGG | DEXA__Fat_Mass | -0.36 | 0.0493298 | 0.3030258 |
| FGL1 | AIRg | 0.64 | 0.0006068 | 0.0232318 |
| FGL1 | auc_glucose | -0.52 | 0.0033167 | 0.0638359 |
| FGL1 | Sg | 0.54 | 0.0050117 | 0.0813985 |
| FGL1 | Glucose_bsl | -0.40 | 0.0273912 | 0.2189744 |
| FHOD1 | auc_glucose | 0.58 | 0.0007504 | 0.0263733 |
| FHOD1 | Glucose_bsl | 0.50 | 0.0050903 | 0.0820098 |
| FHOD1 | HbA1C | 0.45 | 0.0123065 | 0.1403227 |
| FKBP15 | auc_glucose | -0.51 | 0.0037662 | 0.0678443 |
| FKBP15 | AIRg | 0.52 | 0.0078904 | 0.1060851 |
| FKBP15 | Sg | 0.46 | 0.0221217 | 0.1954587 |
| FKBP15 | Temperature | 0.42 | 0.0307179 | 0.2320087 |
| FLNA | auc_glucose | 0.73 | 5.378E-06 | 0.0015122 |
| FLNA | Glucose_bsl | 0.63 | 0.0001854 | 0.012051 |
| FLNA | HbA1C | 0.54 | 0.002308 | 0.0520007 |
| FN1 | Sg | -0.63 | 0.0007424 | 0.0263107 |
| FN1 | AIRg | -0.48 | 0.0146565 | 0.1564091 |
| FN1 | DI | -0.48 | 0.0154635 | 0.1612943 |
| FN1 | auc_glucose | 0.42 | 0.0206731 | 0.1880352 |
| FNBP1L | auc_glucose | 0.55 | 0.0018412 | 0.0444307 |
| FNBP1L | AIRg | -0.50 | 0.0104771 | 0.127599 |
| FNBP1L | Glucose_bsl | 0.45 | 0.0118343 | 0.1378965 |
| FNBP1L | DI | -0.48 | 0.0141665 | 0.1539745 |
| FNBP1L | HbA1C | 0.42 | 0.0205216 | 0.1873728 |
| FSIP2 | auc_glucose | -0.82 | 2.154E-08 | 8.176E-05 |
| FSIP2 | Glucose_bsl | -0.67 | 4.492E-05 | 0.0047618 |
| FSIP2 | HbA1C | -0.65 | 9.948E-05 | 0.0079422 |
| FSIP2 | DI | 0.52 | 0.0072241 | 0.1015412 |
| FSIP2 | Sg | 0.51 | 0.0097384 | 0.1220507 |
| FSIP2 | AIRg | 0.43 | 0.0320193 | 0.2371436 |
| GAL | auc_glucose | -0.67 | 4.42E-05 | 0.0047618 |
| GAL | Glucose_bsl | -0.60 | 0.0004756 | 0.0206413 |
| GAL | HOMA_B | 0.52 | 0.0033388 | 0.0640714 |
| GAL | AIRg | 0.56 | 0.0036611 | 0.0670571 |
| GAL | HbA1C | -0.48 | 0.0074732 | 0.1026104 |
| GAL | Ins30_glu30 | 0.46 | 0.0101038 | 0.1256563 |
| GAL | auc_INSULIN | 0.40 | 0.027736 | 0.2210408 |
| GAL | DI | 0.41 | 0.0417768 | 0.2779185 |
| GAPDH | auc_glucose | -0.50 | 0.005163 | 0.0828518 |
| GEMIN5 | DI | -0.58 | 0.0025297 | 0.0543498 |
| GEMIN5 | auc_glucose | 0.47 | 0.0088518 | 0.1144886 |
| GEMIN5 | auc_c_peptide | -0.57 | 0.0166477 | 0.166657 |
| GEMIN5 | HbA1C | 0.39 | 0.0327803 | 0.2403519 |
| GEMIN5 | MATSUDA | 0.37 | 0.0418146 | 0.2779185 |
| GEMIN5 | LDL | -0.37 | 0.0457401 | 0.2917174 |
| GMIP | LDL | -0.52 | 3.66E-03 | 6.71E-02 |
| GMIP | auc_glucose | 0.48 | 7.28E-03 | 0.1015412 |
| GMIP | Glucose_bsl | 0.42 | 1.96E-02 | 0.1835136 |
| GMIP | AIRg | -0.46 | 0.021212 | 0.191589 |
| GMIP | DI | -0.44 | 0.0266191 | 0.2154369 |
| GMIP | HbA1C | 0.37 | 0.0453727 | 0.2908486 |
| GNG2 | AIRg | -0.61 | 0.0013072 | 0.0362832 |
| GNG2 | auc_glucose | 0.44 | 1.61E-02 | 0.1642502 |
| GNG2 | HOMA_B | -0.43 | 0.0176505 | 0.1713901 |
| GP1BB | auc_INSULIN | 0.58 | 0.0007968 | 0.0271201 |
| GP1BB | HOMA_B | 0.53 | 0.0027733 | 0.0566055 |
| GP1BB | auc_glucose | -0.42 | 0.0203427 | 0.1873728 |
| GP1BB | Glucose_bsl | -0.39 | 0.0325226 | 0.2390041 |
| GPRC5B | HOMA_B | -0.68 | 3.303E-05 | 0.0040308 |
| GPRC5B | auc_INSULIN | -0.62 | 0.0002368 | 0.0136706 |
| GPRC5B | Glucose_bsl | 0.45 | 0.0119128 | 0.1380642 |
| GPRC5B | auc_glucose | 0.44 | 0.0140771 | 0.1537424 |
| GPRC5B | DEXA_Fat_Percentage | 0.39 | 0.0347528 | 0.2502744 |
| GPRC5B | Insulin_bsl | -0.38 | 0.0405027 | 0.2738708 |
| GSK3B | auc_glucose | 0.66 | 6.422E-05 | 0.0061075 |
| GSK3B | Glucose_bsl | 0.53 | 0.0023562 | 0.0527244 |
| GSK3B | HbA1C | 0.44 | 0.0148767 | 0.1577172 |
| GSK3B | auc_c_peptide | -0.54 | 0.0254433 | 0.211111 |
| GSK3B | Respiration_Rate | 0.40 | 0.0296812 | 0.2287243 |
| GSK3B | AIRg | -0.43 | 0.0309903 | 0.2333111 |
| GSN | auc_glucose | -0.51 | 0.0039033 | 0.0697039 |
| GSN | AIRg | 0.51 | 0.0088493 | 0.1144886 |
| GSN | TSH | -0.37 | 0.0414205 | 0.2764359 |
| GSN | Glucose_bsl | -0.37 | 0.0443822 | 0.2870892 |
| GYPA | DEXA_Fat_Percentage | 0.40 | 0.0265054 | 0.2150698 |
| GYPA | auc_glucose | 0.39 | 0.0345214 | 0.2492513 |
| GYPA | DEXA__Fat_Mass | 0.37 | 0.046222 | 0.2939209 |
| GYPA | Glucose_bsl | 0.36 | 0.0481923 | 0.2991166 |
| GYPC | HOMA_B | 0.49 | 0.006546 | 0.0959931 |
| GYPC | TSH | -0.47 | 8.28E-03 | 0.1099354 |
| GYPC | auc_glucose | -0.45 | 1.19E-02 | 0.1380642 |
| GYPC | Glucose_bsl | -0.45 | 0.0128283 | 0.1445307 |
| GYPC | auc_INSULIN | 0.41 | 0.022663 | 0.1983246 |
| GYPC | HDL | 0.38 | 0.0394558 | 0.269893 |
| H1-3 | AIRg | 0.61 | 0.0011198 | 0.0326423 |
| H1-3 | Glucose_bsl | -0.43 | 0.0167775 | 0.1666672 |
| H1-3 | auc_glucose | -0.43 | 0.0176979 | 0.1715648 |
| H1-3 | HbA1C | -0.38 | 0.0383531 | 0.2647059 |
| H1-4 | Glucose_bsl | -0.66 | 7.641E-05 | 0.0065245 |
| H1-4 | auc_glucose | -0.62 | 0.0002483 | 0.0141833 |
| H1-4 | HOMA_B | 0.56 | 0.0013663 | 0.0369702 |
| H1-4 | HbA1C | -0.49 | 0.0057837 | 0.0873899 |
| H1-4 | AIRg | 0.48 | 0.0151603 | 0.1594173 |
| H1-4 | DI | 0.46 | 0.0221844 | 0.1954587 |
| HBA1 | QUICKI | -0.43 | 0.0163674 | 0.1653198 |
| HBA1 | Heart_Rate_Average | 0.40 | 0.0268535 | 0.2159972 |
| HBA1 | auc_INSULIN | 0.40 | 0.0287818 | 0.2253275 |
| HBA1 | HOMA_B | 0.36 | 0.0476657 | 0.2975428 |
| HBB | auc_glucose | -0.72 | 7.515E-06 | 0.0017357 |
| HBB | Glucose_bsl | -0.51 | 0.0042618 | 0.0733015 |
| HBB | HbA1C | -0.50 | 0.0045064 | 0.0757847 |
| HBB | Sg | 0.54 | 0.0053891 | 0.0850027 |
| HBB | AIRg | 0.43 | 0.0316437 | 0.2360319 |
| HDGF | Sg | 0.50 | 0.0108699 | 0.1296972 |
| HDGF | Waist_Circumference | 0.38 | 0.0371477 | 0.2599939 |
| HDGF | DEXA__Lean_Mass | 0.36 | 0.0478335 | 0.2983023 |
| HEG1 | TSH | -0.38 | 0.0398938 | 0.2716421 |
| HEG1 | DEXA_Weight | -0.37 | 0.0423885 | 0.2799815 |
| HSPB1 | auc_glucose | -0.52 | 0.0032338 | 0.0628823 |
| HSPB1 | Sg | 0.45 | 0.0237399 | 0.2016001 |
| HSPB1 | Glucose_bsl | -0.40 | 0.0294179 | 0.2278573 |
| HSPB1 | QUICKI | -0.38 | 0.0383493 | 0.2647059 |
| HSPB1 | DI | 0.40 | 0.0467978 | 0.2949723 |
| IFT88 | auc_glucose | -0.44 | 0.0143115 | 0.1539974 |
| IFT88 | Glucose_bsl | -0.41 | 0.0248019 | 0.2072276 |
| IGFBP3 | auc_glucose | -0.59 | 0.0005374 | 0.022518 |
| IGFBP3 | Glucose_bsl | -0.57 | 0.0009125 | 0.0292369 |
| IGFBP3 | HOMA_B | 0.45 | 0.0132793 | 0.1480642 |
| IGFBP3 | AIRg | 0.47 | 0.0182028 | 0.1754356 |
| IGFBP3 | DEXA_Fat_Percentage | -0.39 | 0.0320048 | 0.2371436 |
| IGFBP3 | HbA1C | -0.37 | 0.0419813 | 0.2786861 |
| IPCEF1 | auc_glucose | 0.53 | 0.0025101 | 0.0543498 |
| IPCEF1 | Glucose_bsl | 0.48 | 0.0073919 | 0.1023382 |
| IPCEF1 | LDL | -0.43 | 0.0211593 | 0.191589 |
| IPCEF1 | HbA1C | 0.41 | 0.0255932 | 0.211111 |
| IPCEF1 | auc_c_peptide | -0.52 | 0.0325119 | 0.2390041 |
| IPCEF1 | AIRg | -0.42 | 0.038399 | 0.2647403 |
| ITGA2 | auc_glucose | 0.58 | 0.0007884 | 0.0269777 |
| ITGA2 | Glucose_bsl | 0.57 | 0.0010925 | 0.0325579 |
| ITGA2 | HbA1C | 0.46 | 0.0110058 | 0.1309947 |
| ITGA2B | DEXA__Fat_Mass | -0.53 | 0.0025968 | 0.0544573 |
| ITGA2B | auc_glucose | -0.51 | 0.0044181 | 0.0751366 |
| ITGA2B | HbA1C | -0.47 | 0.0083668 | 0.1104249 |
| ITGA2B | Insulin_bsl | -0.46 | 0.0102271 | 0.12646 |
| ITGA2B | Glucose_bsl | -0.43 | 0.0188912 | 0.1787301 |
| ITGA2B | HOMA_IR | -0.42 | 0.0213027 | 0.1919878 |
| ITGA2B | Waist_Circumference | -0.40 | 0.0292569 | 0.2276824 |
| ITGA2B | DI | 0.40 | 0.0493602 | 0.3030258 |
| ITGAL | HOMA_B | 0.53 | 0.002502 | 0.0543498 |
| ITGAL | auc_glucose | -0.51 | 0.0036916 | 0.0670611 |
| ITGAL | auc_INSULIN | 0.46 | 9.76E-03 | 0.1221421 |
| ITGAL | Sg | 0.46 | 0.0222826 | 0.1957903 |
| ITGAL | Glucose_bsl | -0.41 | 0.0247718 | 0.2072276 |
| ITGAL | HbA1C | -0.39 | 0.0320189 | 0.2371436 |
| ITGB1 | Glucose_bsl | -0.51 | 0.0040024 | 0.0703351 |
| ITGB1 | auc_glucose | -0.50 | 0.0044381 | 0.0751366 |
| ITGB1 | DEXA__Fat_Mass | -0.49 | 0.0060831 | 0.0904355 |
| ITGB1 | Heart_Rate_Average | -0.45 | 0.0121504 | 0.1395683 |
| ITGB1 | HOMA_IR | -0.44 | 0.0138486 | 0.1520923 |
| ITGB1 | HbA1C | -0.43 | 0.0189039 | 0.1787301 |
| ITGB1 | Insulin_bsl | -0.42 | 0.0210024 | 0.1904945 |
| ITGB1BP1 | auc_glucose | 0.63 | 0.0002203 | 0.0131894 |
| ITGB1BP1 | HbA1C | 0.62 | 0.0002513 | 0.0141833 |
| ITGB1BP1 | DI | -0.63 | 0.0007324 | 0.0263107 |
| ITGB1BP1 | Glucose_bsl | 0.56 | 0.0012367 | 0.0350773 |
| ITGB1BP1 | AIRg | -0.43 | 0.0301184 | 0.2305039 |
| ITGB1BP1 | Respiration_Rate | 0.38 | 0.0439083 | 0.2850954 |
| ITIH1 | HOMA_B | 0.64 | 0.0001275 | 0.0095161 |
| ITIH1 | auc_INSULIN | 0.63 | 0.0001925 | 0.0123264 |
| ITIH1 | Glucose_bsl | -0.55 | 0.0016951 | 0.0421634 |
| ITIH1 | AIRg | 0.56 | 0.0032601 | 0.0631224 |
| ITIH1 | auc_glucose | -0.50 | 0.0052074 | 0.0831516 |
| ITIH2 | auc_glucose | -0.66 | 6.541E-05 | 0.0061305 |
| ITIH2 | Glucose_bsl | -0.58 | 0.0007753 | 0.0268131 |
| ITIH2 | AIRg | 0.56 | 0.0039239 | 0.0697143 |
| ITIH2 | HbA1C | -0.51 | 0.0039823 | 0.070174 |
| ITIH2 | HOMA_IR | -0.38 | 0.0357076 | 0.2537591 |
| ITIH2 | HOMA_B | 0.37 | 0.0420593 | 0.2786861 |
| KIF25 | auc_glucose | -0.57 | 0.0010712 | 0.0323698 |
| KIF25 | Glucose_bsl | -0.55 | 0.0017049 | 0.0422432 |
| KIF25 | HbA1C | -0.36 | 0.049134 | 0.3030258 |
| KMT2C | Waist_Circumference | 0.50 | 0.0050864 | 0.0820098 |
| KMT2C | HbA1C | 0.49 | 0.0056381 | 0.0867166 |
| KMT2C | HOMA_IR | 0.44 | 0.0162485 | 0.164475 |
| KMT2C | Insulin_bsl | 0.40 | 0.0269298 | 0.2163415 |
| KMT2C | Glucose_bsl | 0.37 | 0.0467561 | 0.2949723 |
| KNG1 | auc_glucose | -0.74 | 3.00E-06 | 0.001205 |
| KNG1 | HbA1C | -0.73 | 5.306E-06 | 0.0015122 |
| KNG1 | Glucose_bsl | -0.69 | 2.028E-05 | 0.0029143 |
| KNG1 | DI | 0.56 | 0.003418 | 0.0652046 |
| KNG1 | AIRg | 0.45 | 2.33E-02 | 0.2009239 |
| KNG1 | HOMA_IR | -0.41 | 0.0240134 | 0.2024703 |
| KRT10 | auc_INSULIN | -0.45 | 0.0137023 | 0.1511292 |
| KRT10 | HOMA_B | -0.44 | 0.015574 | 0.1619248 |
| KRT13 | auc_glucose | -0.61 | 0.000365 | 0.0178803 |
| KRT13 | auc_c_peptide | 0.66 | 0.0036498 | 0.0670571 |
| KRT13 | Glucose_bsl | -0.46 | 0.0104601 | 0.127599 |
| KRT13 | Sg | 0.43 | 0.0324309 | 0.2390041 |
| KRT13 | TSH | -0.37 | 0.04648 | 0.2944367 |
| KRT14 | auc_glucose | -0.68 | 3.023E-05 | 0.0038333 |
| KRT14 | Glucose_bsl | -0.59 | 0.0005762 | 0.0230088 |
| KRT14 | HbA1C | -0.56 | 0.0011447 | 0.0331956 |
| KRT14 | DI | 0.52 | 0.0075417 | 0.1031122 |
| KRT14 | HOMA_IR | -0.39 | 0.0339922 | 0.246997 |
| KRT14 | AIRg | 0.41 | 0.0392707 | 0.2696004 |
| KRT27 | auc_glucose | -0.53 | 0.002597 | 0.0544573 |
| KRT27 | Temperature | 0.52 | 0.0069052 | 0.0992349 |
| KRT27 | Glucose_bsl | -0.46 | 0.0107656 | 0.1296482 |
| KRT27 | Waist_Circumference | -0.42 | 0.0220769 | 0.1954587 |
| KRT27 | HbA1C | -0.41 | 0.0239947 | 0.2024703 |
| KRT5 | Glucose_bsl | -0.60 | 0.0004189 | 0.0192958 |
| KRT5 | auc_glucose | -0.59 | 0.0005807 | 0.0230088 |
| KRT5 | AIRg | 0.51 | 0.0087005 | 0.1129838 |
| KRT5 | HbA1C | -0.44 | 0.0153966 | 0.1608557 |
| KRT5 | Sg | 0.43 | 0.030559 | 0.2318537 |
| LMNA | auc_glucose | 0.58 | 0.0008871 | 0.0289756 |
| LMNA | DI | -0.54 | 0.0055858 | 0.0866261 |
| LMNA | HbA1C | 0.49 | 0.0059215 | 0.0890573 |
| LMNA | Glucose_bsl | 0.46 | 0.0104014 | 0.1273989 |
| LMNA | AIRg | -0.48 | 0.0161021 | 0.1642467 |
| LMNA | TSH | 0.42 | 0.0204212 | 0.1873728 |
| LMNA | HDL | -0.40 | 0.029759 | 0.2287243 |
| LPIN2 | Sg | -0.56 | 0.0034792 | 0.065789 |
| LPIN2 | TSH | 0.47 | 0.0093425 | 0.1191674 |
| LPIN2 | QUICKI | 0.40 | 0.0272616 | 0.2187355 |
| LPIN2 | Insulin_bsl | -0.36 | 0.0492962 | 0.3030258 |
| LRRC47 | Glucose_bsl | -0.57 | 0.0009354 | 0.0293662 |
| LRRC47 | auc_glucose | -0.56 | 0.0014422 | 0.0380678 |
| LRRC47 | HOMA_B | 0.44 | 0.0160503 | 0.1642362 |
| LRRC47 | AIRg | 0.48 | 0.0162482 | 0.164475 |
| LRRC47 | Ins30_glu30 | 0.41 | 0.0230958 | 0.2002156 |
| LRRC47 | HbA1C | -0.37 | 0.0453897 | 0.2908486 |
| LRRC47 | DEXA_Fat_Percentage | -0.36 | 0.0493875 | 0.3030258 |
| LYN | auc_glucose | 0.71 | 1.239E-05 | 0.0022256 |
| LYN | Glucose_bsl | 0.62 | 2.75E-04 | 0.0147954 |
| LYN | HbA1C | 0.48 | 0.0067122 | 0.097546 |
| LYN | AIRg | -0.49 | 0.0119848 | 0.1381569 |
| LYN | TSH | 0.43 | 0.017411 | 0.1697379 |
| MAP1B | auc_glucose | -0.49 | 0.0064889 | 0.0953727 |
| MAP1B | AIRg | 0.42 | 0.0345337 | 0.2492513 |
| MAP2K2 | auc_glucose | 0.54 | 0.0022144 | 0.0504245 |
| MAP2K2 | Glucose_bsl | 0.43 | 0.0171778 | 0.1686532 |
| MAP2K2 | Systolic_Blood_Pressure_Average | -0.38 | 0.038058 | 0.2632742 |
| MARCKSL1 | auc_glucose | 0.46 | 0.0108144 | 0.1296972 |
| MARCKSL1 | TSH | 0.39 | 0.0355038 | 0.252589 |
| MARCKSL1 | Glucose_bsl | 0.37 | 0.0470947 | 0.2957311 |
| MASP2 | HOMA_B | -0.63 | 0.0002172 | 0.0131265 |
| MASP2 | auc_INSULIN | -0.60 | 0.0004743 | 0.0206413 |
| MASP2 | Insulin_bsl | -0.52 | 0.0032808 | 0.0633334 |
| MASP2 | Sg | -0.49 | 0.0125028 | 0.1418732 |
| MASP2 | DEXA__Fat_Mass | -0.42 | 0.0200919 | 0.1856209 |
| MASP2 | AIRg | -0.42 | 0.03636 | 0.2569838 |
| MICAL3 | auc_glucose | -0.77 | 7.578E-07 | 0.0005864 |
| MICAL3 | Glucose_bsl | -0.69 | 2.853E-05 | 0.0037652 |
| MICAL3 | HbA1C | -0.57 | 0.000907 | 0.0292369 |
| MICAL3 | DI | 0.42 | 0.0374646 | 0.2612506 |
| MICAL3 | AIRg | 0.40 | 0.0450304 | 0.2900516 |
| MLLT6 | auc_glucose | -0.61 | 0.0003214 | 0.0162879 |
| MLLT6 | Glucose_bsl | -0.54 | 0.0020322 | 0.0479593 |
| MLLT6 | HbA1C | -0.50 | 0.0048932 | 0.0799106 |
| MLLT6 | AIRg | 0.53 | 0.0069269 | 0.0993268 |
| MLLT6 | DI | 0.50 | 0.0102024 | 0.12646 |
| MLLT6 | Sg | 0.42 | 0.0369651 | 0.2589959 |
| MLLT6 | Ins30_glu30 | 0.37 | 0.0431584 | 0.2817601 |
| MS4A1 | Temperature | 0.59 | 0.0014497 | 0.0381098 |
| MS4A1 | Sg | 0.53 | 0.0060807 | 0.0904355 |
| MS4A1 | auc_glucose | -0.43 | 0.0182705 | 0.1755651 |
| MTDH | auc_glucose | 0.63 | 0.0001863 | 0.012051 |
| MTDH | Glucose_bsl | 0.49 | 0.0057756 | 0.0873899 |
| MTDH | Waist_Circumference | 0.43 | 0.0167584 | 0.1666672 |
| MTDH | HbA1C | 0.41 | 0.0247761 | 0.2072276 |
| MYLK | auc_glucose | 0.58 | 0.0007445 | 0.0263107 |
| MYLK | Glucose_bsl | 0.46 | 0.01115 | 0.1315818 |
| MYO1C | auc_glucose | 0.59 | 0.0006071 | 0.0232318 |
| MYO1C | HbA1C | 0.59 | 0.0006134 | 0.0233347 |
| MYO1C | Glucose_bsl | 0.57 | 0.0009481 | 0.0294022 |
| MYO1C | DI | -0.58 | 0.002547 | 0.0544573 |
| MYO1C | AIRg | -0.55 | 0.0047181 | 0.0778367 |
| MYO1C | HOMA_B | -0.45 | 0.0122417 | 0.1402127 |
| MYO9B | auc_glucose | 0.59 | 0.0005397 | 0.022518 |
| MYO9B | HbA1C | 0.51 | 0.0037526 | 0.0677886 |
| MYO9B | Glucose_bsl | 0.51 | 4.31E-03 | 0.0736752 |
| MYO9B | Respiration_Rate | 0.38 | 4.32E-02 | 0.2817601 |
| NCF2 | DEXA__Fat_Mass | -0.41 | 0.0229613 | 0.1995841 |
| NCF2 | AIRg | -0.42 | 0.0353699 | 0.252589 |
| NCF2 | Waist_Circumference | -0.37 | 0.0412833 | 0.2759923 |
| NEXN | auc_glucose | 0.72 | 9.003E-06 | 0.0019546 |
| NEXN | Glucose_bsl | 0.60 | 0.0004215 | 0.0192958 |
| NEXN | HbA1C | 0.53 | 0.00241 | 0.0535285 |
| NEXN | Respiration_Rate | 0.43 | 0.0204625 | 0.1873728 |
| NME2 | AIRg | -0.62 | 0.0009906 | 0.030219 |
| NME2 | DI | -0.47 | 0.0174279 | 0.1697379 |
| NME2 | auc_glucose | 0.41 | 0.025686 | 0.2113375 |
| NRBP1 | DEXA__Lean_Mass | -0.57 | 0.0009311 | 0.0293662 |
| NRBP1 | DEXA_Weight | -0.48 | 0.0072596 | 0.1015412 |
| NRBP1 | AIRg | -0.49 | 0.0126971 | 0.1438036 |
| NRBP1 | Waist_Circumference | -0.44 | 0.0145956 | 0.1562258 |
| NRBP1 | HDL | 0.42 | 0.0214735 | 0.1923397 |
| NRBP1 | DEXA_Fat_Percentage | 0.42 | 0.0221553 | 0.1954587 |
| NRBP1 | auc_glucose | 0.41 | 0.0239478 | 0.2024703 |
| NRBP1 | HOMA_B | -0.40 | 0.0267088 | 0.2154369 |
| OPTN | HOMA_B | 0.50 | 0.0045908 | 0.0767157 |
| OPTN | auc_glucose | -0.48 | 0.0068294 | 0.0983648 |
| OPTN | AIRg | 0.49 | 0.0129913 | 0.1461126 |
| OPTN | Ins30_glu30 | 0.41 | 0.0234502 | 0.2009239 |
| OPTN | auc_INSULIN | 0.39 | 0.0313057 | 0.2351382 |
| OPTN | Glucose_bsl | -0.37 | 0.0425553 | 0.2799815 |
| OSBP | auc_glucose | 0.55 | 0.0018341 | 0.0444307 |
| OSBP | HbA1C | 0.45 | 0.0134961 | 0.1497086 |
| OSBP | Glucose_bsl | 0.41 | 0.0231896 | 0.2007589 |
| PARD3 | auc_glucose | 0.63 | 0.0002144 | 0.0130818 |
| PARD3 | Glucose_bsl | 0.52 | 0.003102 | 0.0609737 |
| PARD3 | HbA1C | 0.43 | 0.0184444 | 0.1761887 |
| PDAP1 | auc_glucose | 0.64 | 0.0001562 | 0.0111004 |
| PDAP1 | Glucose_bsl | 0.62 | 0.0002301 | 0.0135549 |
| PDAP1 | HbA1C | 0.59 | 0.000587 | 0.0230088 |
| PDAP1 | Respiration_Rate | 0.48 | 0.0088697 | 0.1144918 |
| PDAP1 | DI | -0.50 | 0.0105859 | 0.1279609 |
| PDAP1 | AIRg | -0.47 | 0.0164946 | 0.165768 |
| PDAP1 | LDL | -0.41 | 0.0255688 | 0.211111 |
| PGRMC1 | auc_glucose | 0.74 | 3.168E-06 | 0.001205 |
| PGRMC1 | Glucose_bsl | 0.67 | 5.811E-05 | 0.0058722 |
| PGRMC1 | HbA1C | 0.51 | 0.0040753 | 0.0711946 |
| PGRMC1 | Respiration_Rate | 0.42 | 0.0235777 | 0.2009239 |
| PGRMC1 | Heart_Rate_Average | 0.40 | 0.0267769 | 0.2156492 |
| PGRMC1 | AIRg | -0.41 | 0.0425464 | 0.2799815 |
| PIEZO1 | auc_glucose | 0.56 | 0.0012254 | 0.0349112 |
| PIEZO1 | HOMA_B | -0.52 | 0.0034912 | 0.0658239 |
| PIEZO1 | AIRg | -0.56 | 0.003542 | 0.0662027 |
| PIEZO1 | Glucose_bsl | 0.48 | 0.0078849 | 0.1060851 |
| PIEZO1 | auc_INSULIN | -0.48 | 0.0079124 | 0.1061603 |
| PIEZO1 | Sg | -0.51 | 0.0096227 | 0.121466 |
| PIEZO1 | TSH | 0.40 | 0.0277275 | 0.2210408 |
| PIEZO1 | HbA1C | 0.39 | 0.030984 | 0.2333111 |
| PIEZO1 | DI | -0.42 | 0.0366815 | 0.2577573 |
| PKM | auc_glucose | -0.57 | 0.000997 | 0.0302717 |
| PKM | Temperature | 0.52 | 0.0066114 | 0.0962967 |
| PKM | HbA1C | -0.48 | 0.0069442 | 0.0993538 |
| PKM | Glucose_bsl | -0.47 | 0.0085585 | 0.1118136 |
| PKM | DI | 0.47 | 0.0191636 | 0.1804537 |
| PKM | Sg | 0.44 | 0.0297366 | 0.2287243 |
| PKM | HOMA_IR | -0.40 | 0.0306455 | 0.2318537 |
| PKN1 | auc_glucose | -0.71 | 9.544E-06 | 0.0019546 |
| PKN1 | Glucose_bsl | -0.67 | 6.099E-05 | 0.0060456 |
| PKN1 | AIRg | 0.61 | 0.001288 | 0.0361205 |
| PKN1 | HbA1C | -0.53 | 0.0028185 | 0.0568183 |
| PKN1 | DI | 0.49 | 0.0130426 | 0.146278 |
| PKN1 | HOMA_IR | -0.37 | 0.0456597 | 0.2915137 |
| PMS1 | auc_glucose | 0.63 | 0.0001736 | 0.0115764 |
| PMS1 | Glucose_bsl | 0.54 | 0.00221 | 0.0504245 |
| PMS1 | HbA1C | 0.46 | 0.0103713 | 0.1273989 |
| PMS1 | AIRg | -0.47 | 0.0191699 | 0.1804537 |
| PMS1 | HOMA_B | -0.40 | 0.0294106 | 0.2278573 |
| PPP3CB | HbA1C | 0.63 | 0.0001957 | 0.0124077 |
| PPP3CB | auc_glucose | 0.60 | 0.0005207 | 0.0220073 |
| PPP3CB | Glucose_bsl | 0.57 | 0.0011206 | 0.0326423 |
| PPP3CB | auc_c_peptide | -0.58 | 0.0157403 | 0.1626081 |
| PPP3CB | DEXA__Lean_Mass | 0.40 | 0.0278792 | 0.2212209 |
| PPP3CB | DEXA_Weight | 0.38 | 3.61E-02 | 0.2559893 |
| PRG4 | auc_glucose | -0.58 | 0.0007406 | 0.0263107 |
| PRG4 | Glucose_bsl | -0.49 | 0.0055777 | 0.0866261 |
| PRG4 | HbA1C | -0.40 | 0.0297798 | 0.2287243 |
| PRKAR2A | AIRg | -0.63 | 0.0008244 | 0.0276361 |
| PRKAR2A | auc_glucose | 0.56 | 0.0013359 | 0.0367855 |
| PRKAR2A | DI | -0.56 | 0.0039126 | 0.0697039 |
| PRKAR2A | DEXA_Fat_Percentage | -0.48 | 0.0068118 | 0.0983297 |
| PRKAR2A | DEXA__Lean_Mass | 0.43 | 0.0188384 | 0.1786332 |
| PRKAR2A | Sg | -0.45 | 0.0223683 | 0.1960107 |
| PRKAR2A | Respiration_Rate | 0.41 | 0.0264859 | 0.2150698 |
| PRKCD | auc_glucose | 0.77 | 6.512E-07 | 0.0005864 |
| PRKCD | Glucose_bsl | 0.66 | 6.951E-05 | 0.0063376 |
| PRKCD | HbA1C | 0.56 | 0.0013385 | 0.0367855 |
| PRKCD | AIRg | -0.51 | 0.0096354 | 0.121466 |
| PSMF1 | auc_glucose | -0.44 | 0.0148132 | 0.1573502 |
| PSMF1 | Sg | 0.46 | 0.0200902 | 0.1856209 |
| PSMF1 | DEXA_Fat_Percentage | 0.38 | 0.0357981 | 0.2541235 |
| PSMF1 | DI | 0.42 | 0.0367086 | 0.2577573 |
| PTPDC1 | Ins30_glu30 | 0.42 | 0.0211971 | 0.191589 |
| PTPDC1 | Sg | -0.43 | 0.0307455 | 0.2320087 |
| PTPDC1 | auc_glucose | 0.39 | 0.0344191 | 0.2489803 |
| PTPN12 | AIRg | -0.57 | 0.00319 | 0.062325 |
| PTPN12 | auc_glucose | 0.52 | 0.0034421 | 0.0654704 |
| PTPN12 | Sg | -0.48 | 0.0159041 | 0.1635168 |
| PTPN12 | DI | -0.45 | 0.0228062 | 0.1985029 |
| PTPN12 | HbA1C | 0.39 | 0.0320861 | 0.2371436 |
| PTPN12 | Glucose_bsl | 0.38 | 0.0410068 | 0.2751949 |
| PTPRC | auc_glucose | -0.51 | 0.0036778 | 0.0670571 |
| PTPRC | Glucose_bsl | -0.50 | 0.0050221 | 0.0813985 |
| PTPRC | HOMA_B | 0.43 | 0.0177672 | 0.1717492 |
| PTPRC | DEXA_Fat_Percentage | -0.42 | 0.0205581 | 0.1873728 |
| PTPRC | HbA1C | -0.41 | 0.0235008 | 0.2009239 |
| PTPRC | auc_INSULIN | 0.36 | 0.0496374 | 0.3039818 |
| PTPRCAP | auc_glucose | -0.46 | 0.0108555 | 0.1296972 |
| PTPRJ | auc_glucose | 0.70 | 1.481E-05 | 0.0024361 |
| PTPRJ | Glucose_bsl | 0.56 | 0.0014088 | 0.0375565 |
| PTPRJ | HbA1C | 0.50 | 0.0048849 | 0.0799106 |
| PUM2 | auc_glucose | 0.54 | 0.0021307 | 0.0492123 |
| PUM2 | Glucose_bsl | 0.51 | 0.0043841 | 0.074808 |
| PUM2 | HbA1C | 0.48 | 0.0078901 | 0.1060851 |
| RAB12 | TSH | 0.62 | 0.0002986 | 0.0156885 |
| RAB12 | Ins30_glu30 | -0.50 | 0.004871 | 0.0799106 |
| RAB12 | auc_glucose | 0.44 | 0.0161533 | 0.1642502 |
| RAB12 | Glucose_bsl | 0.40 | 0.0286879 | 0.2251516 |
| RAB12 | HDL | -0.38 | 0.036644 | 0.2577573 |
| RAB12 | HOMA_B | -0.36 | 0.047459 | 0.2965385 |
| RAB6A | auc_glucose | -0.58 | 0.0007884 | 0.0269777 |
| RAB6A | Glucose_bsl | -0.54 | 0.0020499 | 0.0479593 |
| RAB6A | HbA1C | -0.53 | 0.0025952 | 0.0544573 |
| RAB6A | DEXA__Fat_Mass | -0.49 | 0.0064429 | 0.0949113 |
| RAB6A | DI | 0.46 | 0.0215979 | 0.1926534 |
| RABGAP1 | Glucose_bsl | 0.68 | 4.124E-05 | 0.0045987 |
| RABGAP1 | auc_glucose | 0.63 | 0.0001655 | 0.0113861 |
| RABGAP1 | HbA1C | 0.61 | 0.0003431 | 0.0170699 |
| RABGAP1 | DI | -0.51 | 0.0099702 | 0.1242336 |
| RABGAP1 | HOMA_B | -0.45 | 0.0117662 | 0.1373506 |
| RABGAP1 | Diastolic_Blood_Pressure_Average | 0.37 | 0.0465223 | 0.2944367 |
| RABGEF1 | auc_glucose | 0.69 | 2.758E-05 | 0.0037206 |
| RABGEF1 | Glucose_bsl | 0.55 | 0.0015294 | 0.039093 |
| RABGEF1 | HbA1C | 0.53 | 0.0027834 | 0.0566055 |
| RABGEF1 | AIRg | -0.51 | 0.0084338 | 0.1108567 |
| RABGEF1 | DI | -0.44 | 0.029239 | 0.2276824 |
| RABGEF1 | Respiration_Rate | 0.38 | 0.0403087 | 0.2735231 |
| RALBP1 | AIRg | -0.65 | 0.0003887 | 0.0186692 |
| RALBP1 | Glucose_bsl | 0.52 | 0.0035671 | 0.0664796 |
| RALBP1 | auc_glucose | 0.51 | 0.0042254 | 0.0728724 |
| RALBP1 | HbA1C | 0.46 | 0.0104015 | 0.1273989 |
| RALBP1 | DI | -0.50 | 0.0114361 | 0.1342239 |
| RALBP1 | HOMA_B | -0.42 | 0.0201624 | 0.1860062 |
| RASA3 | auc_glucose | 0.73 | 4.887E-06 | 0.0015122 |
| RASA3 | Glucose_bsl | 0.60 | 0.0004045 | 0.0190924 |
| RASA3 | HbA1C | 0.53 | 0.002463 | 0.0541766 |
| RASA3 | LDL | -0.38 | 0.042054 | 0.2786861 |
| RASGRP2 | Sg | -0.61 | 0.0013424 | 0.0367855 |
| RASGRP2 | auc_glucose | 0.56 | 0.0014112 | 0.0375565 |
| RASGRP2 | HbA1C | 0.42 | 0.020028 | 0.1855599 |
| RASGRP2 | DI | -0.43 | 0.0302889 | 0.2312006 |
| RASGRP2 | Glucose_bsl | 0.37 | 0.042237 | 0.2792908 |
| RECQL | Waist_Circumference | 0.56 | 0.0013551 | 0.0368204 |
| RECQL | HDL | -0.51 | 0.0038532 | 0.0690941 |
| RECQL | Temperature | -0.53 | 0.0054395 | 0.0853819 |
| RECQL | MATSUDA | -0.44 | 0.0152406 | 0.1597001 |
| RECQL | QUICKI | -0.41 | 0.0233522 | 0.2009239 |
| RECQL | Triglycerides | 0.37 | 0.0426012 | 0.2799815 |
| RETREG2 | AIRg | -0.54 | 0.0054213 | 0.0853036 |
| RETREG2 | Triglycerides | 0.37 | 0.044969 | 0.2899444 |
| RHBDF2 | auc_glucose | 0.60 | 0.0004988 | 0.0215039 |
| RHBDF2 | Glucose_bsl | 0.52 | 0.0031723 | 0.0621667 |
| RHBDF2 | HbA1C | 0.46 | 0.0096686 | 0.1216472 |
| RMDN3 | auc_glucose | -0.53 | 0.0024742 | 0.05424 |
| RMDN3 | HOMA_B | 0.49 | 0.0058203 | 0.0877389 |
| RMDN3 | AIRg | 0.50 | 0.011535 | 0.1351392 |
| RMDN3 | Glucose_bsl | -0.42 | 0.0214536 | 0.1923397 |
| RMDN3 | HbA1C | -0.41 | 0.0232497 | 0.2009239 |
| RMDN3 | auc_c_peptide | 0.54 | 0.0247404 | 0.2072276 |
| RMDN3 | auc_INSULIN | 0.40 | 0.0294553 | 0.2278573 |
| RMDN3 | Sg | 0.40 | 0.0461672 | 0.2938613 |
| RRAS2 | auc_glucose | -0.82 | 2.529E-08 | 8.176E-05 |
| RRAS2 | Glucose_bsl | -0.66 | 6.263E-05 | 0.0060456 |
| RRAS2 | HbA1C | -0.65 | 8.728E-05 | 0.0071448 |
| RRAS2 | AIRg | 0.61 | 0.0012192 | 0.0348863 |
| RRAS2 | DI | 0.55 | 0.0047165 | 0.0778367 |
| RRAS2 | Sg | 0.51 | 0.0097862 | 0.1221768 |
| RSU1 | auc_glucose | 0.58 | 0.0008752 | 0.0288774 |
| RSU1 | Glucose_bsl | 0.57 | 0.0010829 | 0.0325579 |
| RSU1 | HbA1C | 0.52 | 0.0035231 | 0.0660406 |
| RSU1 | Waist_Circumference | 0.51 | 0.0039356 | 0.0697191 |
| RSU1 | DEXA__Fat_Mass | 0.39 | 0.0348014 | 0.2503453 |
| RSU1 | HDL | -0.39 | 0.0354694 | 0.252589 |
| RSU1 | HOMA_IR | 0.37 | 0.0456631 | 0.2915137 |
| RSU1 | Heart_Rate_Average | 0.36 | 0.0492917 | 0.3030258 |
| S100A9 | auc_glucose | 0.78 | 4.833E-07 | 0.0005463 |
| S100A9 | AIRg | -0.70 | 0.0001147 | 0.0088336 |
| S100A9 | Glucose_bsl | 0.55 | 0.0014913 | 0.0385762 |
| S100A9 | HbA1C | 0.54 | 0.0023158 | 0.0520007 |
| S100A9 | Sg | -0.54 | 0.0053008 | 0.0840204 |
| S100A9 | DI | -0.47 | 0.01877 | 0.1785162 |
| SERINC1 | auc_glucose | 0.49 | 0.0056577 | 0.0867166 |
| SERINC1 | TSH | 0.38 | 0.0373598 | 0.2611952 |
| SERINC1 | Temperature | -0.39 | 0.0462736 | 0.2939599 |
| SERPINC1 | Glucose_bsl | -0.56 | 0.0013016 | 0.0362821 |
| SERPINC1 | auc_glucose | -0.56 | 0.0013853 | 0.0373269 |
| SERPINC1 | auc_c_peptide | 0.67 | 0.0032379 | 0.0628823 |
| SERPINC1 | HOMA_B | 0.44 | 0.0157941 | 0.162903 |
| SERPINC1 | TSH | -0.42 | 0.0198217 | 0.1844417 |
| SERPINC1 | HbA1C | -0.40 | 0.0264652 | 0.2150698 |
| SERPINC1 | AIRg | 0.42 | 0.0365559 | 0.2577573 |
| SERPINC1 | Respiration_Rate | -0.37 | 4.71E-02 | 0.2957311 |
| SERPINF2 | auc_glucose | -0.62 | 0.0002665 | 0.0147288 |
| SERPINF2 | Glucose_bsl | -0.57 | 0.0011164 | 0.0326423 |
| SERPINF2 | HbA1C | -0.41 | 0.0254925 | 0.211111 |
| SIPA1 | auc_glucose | -0.76 | 1.286E-06 | 0.0007561 |
| SIPA1 | Glucose_bsl | -0.66 | 8.134E-05 | 0.0068317 |
| SIPA1 | HbA1C | -0.53 | 0.0025228 | 0.0543498 |
| SIPA1 | HOMA_B | 0.44 | 0.016099 | 0.1642467 |
| SLAIN2 | Respiration_Rate | 0.49 | 0.0073883 | 0.1023382 |
| SLAIN2 | auc_glucose | 0.47 | 0.0083996 | 0.1106312 |
| SLAIN2 | Glucose_bsl | 0.45 | 0.0135725 | 0.1502973 |
| SLAIN2 | HbA1C | 0.44 | 0.0155153 | 0.1615735 |
| SLAIN2 | HOMA_IR | 0.39 | 0.0346303 | 0.2496704 |
| SLAIN2 | Waist_Circumference | 0.38 | 0.0375865 | 0.2612506 |
| SLC12A7 | AIRg | -0.56 | 0.0039458 | 0.0697191 |
| SLC12A7 | HOMA_B | -0.48 | 0.0071175 | 0.1007201 |
| SLC12A7 | auc_glucose | 0.44 | 0.0145809 | 0.1562258 |
| SLC12A7 | auc_INSULIN | -0.40 | 0.0303597 | 0.231256 |
| SLC12A7 | TSH | 0.38 | 0.039522 | 0.269893 |
| SLC12A7 | Glucose_bsl | 0.37 | 0.0421684 | 0.279123 |
| SLC29A1 | HOMA_B | -0.72 | 6.539E-06 | 0.0016915 |
| SLC29A1 | auc_INSULIN | -0.66 | 6.233E-05 | 0.0060456 |
| SLC29A1 | AIRg | -0.53 | 0.0063407 | 0.093833 |
| SLC29A1 | auc_glucose | 0.45 | 0.0132415 | 0.1478977 |
| SLC29A1 | Glucose_bsl | 0.43 | 0.0163924 | 0.1653198 |
| SLC29A1 | Ins30_glu30 | -0.37 | 0.0472586 | 0.2957311 |
| SLC2A4 | auc_glucose | 0.54 | 0.0019193 | 0.0458004 |
| SLC2A4 | Glucose_bsl | 0.53 | 0.0023776 | 0.0530196 |
| SLC2A4 | DEXA__Fat_Mass | 0.53 | 0.0025802 | 0.0544573 |
| SLC2A4 | HbA1C | 0.50 | 0.0052243 | 0.0832163 |
| SLC2A4 | Waist_Circumference | 0.40 | 0.027393 | 0.2189744 |
| SLC2A4 | DEXA_Weight | 0.40 | 0.0303138 | 0.2312006 |
| SLC2A4 | HOMA_IR | 0.38 | 0.0394395 | 0.269893 |
| SLC2A4 | DI | -0.40 | 0.0465842 | 0.2944867 |
| SLC43A2 | auc_glucose | -0.61 | 0.0003224 | 0.0162879 |
| SLC43A2 | HbA1C | -0.55 | 0.0014716 | 0.0383746 |
| SLC43A2 | HOMA_IR | -0.52 | 0.0029233 | 0.0581683 |
| SLC43A2 | Glucose_bsl | -0.47 | 0.0080479 | 0.1077555 |
| SLC43A2 | LDL | 0.48 | 0.0082957 | 0.1099354 |
| SLC43A2 | Insulin_bsl | -0.46 | 0.0103101 | 0.1272432 |
| SLC43A2 | QUICKI | 0.42 | 0.0215153 | 0.1924475 |
| SLC43A2 | Waist_Circumference | -0.40 | 0.0296702 | 0.2287243 |
| SLC43A3 | auc_glucose | 0.43 | 0.0169871 | 0.168094 |
| SLC9A1 | Respiration_Rate | 0.54 | 0.002602 | 0.0544573 |
| SLC9A1 | auc_glucose | 0.50 | 0.0047049 | 0.0778367 |
| SLC9A1 | Glucose_bsl | 0.38 | 0.0410207 | 0.2751949 |
| SNAP23 | auc_glucose | -0.77 | 8.16E-07 | 0.0005864 |
| SNAP23 | Glucose_bsl | -0.66 | 8.504E-05 | 0.0070507 |
| SNAP23 | HbA1C | -0.62 | 0.0002598 | 0.014486 |
| SNAP23 | AIRg | 0.53 | 0.0070028 | 0.0999719 |
| SNAP23 | Sg | 0.49 | 0.0130512 | 0.146278 |
| SNAP23 | HOMA_B | 0.43 | 0.0165076 | 0.165768 |
| SNAP23 | DI | 0.47 | 0.0170704 | 0.1685405 |
| SPECC1 | DEXA__Fat_Mass | -0.58 | 0.0008471 | 0.0280945 |
| SPECC1 | Heart_Rate_Average | -0.46 | 0.0105602 | 0.1279081 |
| SPECC1 | DEXA_Fat_Percentage | -0.37 | 0.0413113 | 0.2759923 |
| SPTA1 | auc_glucose | 0.44 | 1.44E-02 | 0.1550486 |
| SPTA1 | Glucose_bsl | 0.40 | 2.95E-02 | 0.2278573 |
| SPTA1 | AIRg | -0.40 | 0.0498571 | 0.3044623 |
| SPTBN1 | auc_glucose | 0.71 | 9.708E-06 | 0.0019546 |
| SPTBN1 | HbA1C | 0.61 | 0.0003008 | 0.0156885 |
| SPTBN1 | Glucose_bsl | 0.59 | 0.0005766 | 0.0230088 |
| SPTBN1 | DI | -0.59 | 0.0017663 | 0.0434321 |
| SPTBN1 | LDL | -0.46 | 0.0127976 | 0.1445307 |
| SPTBN1 | AIRg | -0.48 | 0.0142845 | 0.1539974 |
| SPTBN1 | HOMA_IR | 0.40 | 0.0303167 | 0.2312006 |
| ST13 | auc_INSULIN | -0.49 | 0.0061399 | 0.0910704 |
| ST13 | HOMA_B | -0.43 | 0.0171861 | 0.1686532 |
| ST13 | DEXA_Weight | -0.39 | 0.0326981 | 0.2400211 |
| STMN1 | auc_glucose | -0.40 | 0.028933 | 0.2259022 |
| STMN1 | HOMA_B | 0.40 | 0.0306395 | 0.2318537 |
| STOML2 | auc_glucose | 0.41 | 0.0234182 | 0.2009239 |
| STT3B | AIRg | -0.73 | 3.252E-05 | 0.0040308 |
| STT3B | auc_glucose | 0.54 | 0.001933 | 0.0459592 |
| STT3B | Glucose_bsl | 0.52 | 0.0030251 | 0.0598267 |
| STT3B | HOMA_B | -0.40 | 0.0279607 | 0.2215583 |
| STT3B | Ins30_glu30 | -0.39 | 0.0349757 | 0.2507694 |
| STT3B | DI | -0.42 | 3.50E-02 | 0.2507694 |
| STT3B | HbA1C | 0.38 | 0.0397389 | 0.2710879 |
| STT3B | auc_INSULIN | -0.38 | 0.0399041 | 0.2716421 |
| SYTL4 | Glucose_bsl | 0.63 | 0.0002009 | 0.0124905 |
| SYTL4 | HbA1C | 0.62 | 0.0002522 | 0.0141833 |
| SYTL4 | auc_glucose | 0.61 | 0.0003926 | 0.0186703 |
| SYTL4 | HOMA_IR | 0.48 | 0.0075164 | 0.1029839 |
| SYTL4 | Waist_Circumference | 0.48 | 0.0077441 | 0.1052126 |
| SYTL4 | DEXA__Fat_Mass | 0.41 | 0.0233782 | 0.2009239 |
| SYTL4 | DEXA_Weight | 0.41 | 0.0235815 | 0.2009239 |
| SYTL4 | Insulin_bsl | 0.36 | 0.0497502 | 0.3043849 |
| TAGLN2 | auc_glucose | 0.51 | 0.0040843 | 0.0711946 |
| TAGLN2 | Respiration_Rate | 0.43 | 0.0213545 | 0.1920129 |
| TAGLN2 | Glucose_bsl | 0.42 | 0.0221607 | 0.1954587 |
| TAGLN2 | HbA1C | 0.40 | 0.0279903 | 0.2215583 |
| TBC1D5 | HbA1C | 0.44 | 0.0152613 | 0.1597001 |
| TBC1D5 | HOMA_IR | 0.41 | 0.0238599 | 0.2022305 |
| TBC1D5 | Insulin_bsl | 0.41 | 2.61E-02 | 0.2138998 |
| TBC1D5 | auc_glucose | 0.39 | 0.031613 | 0.2360319 |
| TBC1D5 | DI | -0.43 | 0.0318933 | 0.2371436 |
| TBXA2R | auc_glucose | -0.49 | 0.0065733 | 0.0961753 |
| TBXA2R | Glucose_bsl | -0.48 | 0.0077815 | 0.105499 |
| TBXA2R | HbA1C | -0.43 | 0.0165582 | 0.1660187 |
| TBXA2R | AIRg | 0.44 | 0.0287979 | 0.2253275 |
| TC2N | DI | -0.59 | 0.001776 | 0.0435052 |
| TC2N | auc_glucose | 0.50 | 0.0045117 | 0.0757847 |
| TC2N | HbA1C | 0.44 | 0.0151143 | 0.1591928 |
| TC2N | Glucose_bsl | 0.38 | 0.0403331 | 0.2735231 |
| TC2N | Temperature | -0.40 | 0.0426482 | 0.2800057 |
| TC2N | auc_c_peptide | -0.50 | 0.0432203 | 0.2817601 |
| TEKT3 | auc_glucose | -0.51 | 0.0041276 | 0.0717563 |
| TEKT3 | Glucose_bsl | -0.49 | 0.0059635 | 0.0894807 |
| TEKT3 | HbA1C | -0.46 | 0.0103932 | 0.1273989 |
| TEKT3 | DI | 0.46 | 0.0219216 | 0.1947347 |
| TEKT3 | HOMA_IR | -0.37 | 0.0412245 | 0.2759899 |
| TLN1 | auc_glucose | 0.68 | 3.734E-05 | 0.0043905 |
| TLN1 | Glucose_bsl | 0.59 | 0.0005744 | 0.0230088 |
| TLN1 | HbA1C | 0.46 | 0.0102177 | 0.12646 |
| TMEM40 | auc_glucose | -0.43 | 0.0179435 | 0.1731947 |
| TMEM40 | TSH | -0.38 | 0.0410218 | 0.2751949 |
| TMEM40 | Waist_Circumference | -0.36 | 0.0495713 | 0.3038651 |
| TMSB4X | QUICKI | 0.42 | 0.0213777 | 0.1920129 |
| TNRC18 | Glucose_bsl | 0.45 | 0.0132075 | 0.1477735 |
| TNRC18 | DEXA_Fat_Percentage | 0.38 | 0.0375672 | 0.2612506 |
| TNRC18 | auc_glucose | 0.36 | 0.0479699 | 0.2987215 |
| TPI1 | auc_glucose | -0.56 | 0.0013506 | 0.0368204 |
| TPI1 | Glucose_bsl | -0.49 | 0.0057506 | 0.0872982 |
| TPI1 | HbA1C | -0.41 | 0.0239625 | 0.2024703 |
| TPM4 | auc_glucose | -0.64 | 0.000128 | 0.0095161 |
| TPM4 | AIRg | 0.68 | 0.0001705 | 0.0114876 |
| TPM4 | Sg | 0.65 | 0.0004186 | 0.0192958 |
| TPM4 | DI | 0.60 | 0.0014242 | 0.0377481 |
| TPM4 | Glucose_bsl | -0.53 | 0.0027338 | 0.0563036 |
| TPM4 | HbA1C | -0.46 | 0.0111311 | 0.1315818 |
| TRIM21 | auc_glucose | -0.59 | 0.0005844 | 0.0230088 |
| TRIM21 | Glucose_bsl | -0.55 | 1.62E-03 | 0.0410485 |
| TRIM21 | DEXA__Fat_Mass | -0.38 | 0.0374422 | 0.2612506 |
| TRPC6 | auc_glucose | 0.57 | 0.0011121 | 0.0326423 |
| TRPC6 | Respiration_Rate | 0.41 | 0.0282525 | 0.2228158 |
| TRPC6 | Glucose_bsl | 0.40 | 0.0293718 | 0.2278573 |
| TRPC6 | Si | -0.40 | 0.0481709 | 0.2991166 |
| TRPC6 | HbA1C | 0.36 | 0.0481954 | 0.2991166 |
| TUBA1B | auc_glucose | -0.62 | 0.0002714 | 0.0147954 |
| TUBA1B | Glucose_bsl | -0.49 | 0.0054802 | 0.0858128 |
| TUBA1B | HbA1C | -0.48 | 0.0068042 | 0.0983297 |
| TUBA1B | HOMA_IR | -0.47 | 0.0086894 | 0.1129838 |
| TUBA1B | Insulin_bsl | -0.45 | 0.0125047 | 0.1418732 |
| TUBA1B | DI | 0.45 | 0.023724 | 0.2016001 |
| TUBA1B | Heart_Rate_Average | -0.39 | 0.0355007 | 0.252589 |
| TUBA1B | Waist_Circumference | -0.36 | 0.0493819 | 0.3030258 |
| TUBA1C | auc_glucose | 0.55 | 0.0018121 | 0.0442217 |
| TUBA1C | Glucose_bsl | 0.50 | 0.0046719 | 0.0776693 |
| TUBA4A | AIRg | 0.60 | 0.0016599 | 0.0414468 |
| TUBA4A | auc_glucose | -0.50 | 0.0044383 | 0.0751366 |
| TUBA4A | Glucose_bsl | -0.50 | 0.0052863 | 0.083997 |
| TUBA4A | TSH | -0.48 | 0.0071043 | 0.1007201 |
| TUBA4A | HOMA_B | 0.47 | 0.0082776 | 0.1099354 |
| TUBA4A | auc_INSULIN | 0.44 | 0.0156221 | 0.1621637 |
| TUBA4A | Ins30_glu30 | 0.43 | 0.0171312 | 0.168626 |
| TUBA4A | DEXA_Fat_Percentage | -0.38 | 0.040847 | 0.2751949 |
| TUBB1 | auc_glucose | -0.72 | 7.405E-06 | 0.0017357 |
| TUBB1 | Glucose_bsl | -0.71 | 9.776E-06 | 0.0019546 |
| TUBB1 | HbA1C | -0.60 | 0.0004178 | 0.0192958 |
| TUBB1 | AIRg | 0.52 | 0.0073123 | 0.1016962 |
| TUBB1 | DI | 0.51 | 0.0097256 | 0.1220507 |
| TUBB1 | HOMA_B | 0.42 | 0.0217063 | 0.1933536 |
| TUBB1 | auc_INSULIN | 0.38 | 0.0362128 | 0.256504 |
| UBE2V1 | HbA1C | 0.50 | 0.0053873 | 0.0850027 |
| UBE2V1 | auc_glucose | 0.47 | 0.0091305 | 0.1175981 |
| UBE2V1 | Glucose_bsl | 0.43 | 0.0187188 | 0.1785162 |
| UCHL3 | auc_glucose | -0.40 | 0.0285742 | 0.2245313 |
| USP4 | auc_glucose | 0.64 | 0.0001627 | 0.0113861 |
| USP4 | Glucose_bsl | 0.60 | 0.0004337 | 0.0194929 |
| USP4 | HbA1C | 0.44 | 0.0140853 | 0.1537424 |
| USP4 | Waist_Circumference | 0.38 | 0.0365874 | 0.2577573 |
| USP4 | Temperature | -0.40 | 0.0415364 | 0.2769234 |
| USP4 | HOMA_B | -0.37 | 0.0429889 | 0.281386 |
| VAMP5 | auc_glucose | 0.54 | 0.0021403 | 0.0492571 |
| VAMP5 | HbA1C | 0.53 | 0.0028291 | 0.0568183 |
| VAMP5 | Glucose_bsl | 0.51 | 0.003857 | 0.0690941 |
| VAMP5 | auc_c_peptide | -0.52 | 0.031202 | 0.2346316 |
| VAPB | AIRg | -0.57 | 0.0027941 | 0.056645 |
| VAPB | auc_glucose | 0.52 | 0.0029732 | 0.0589811 |
| VAPB | Glucose_bsl | 0.45 | 0.0122848 | 0.1403227 |
| VAPB | Ins30_glu30 | -0.41 | 0.025487 | 0.211111 |
| VTN | auc_glucose | -0.64 | 0.0001242 | 0.0094466 |
| VTN | HOMA_B | 0.64 | 0.000146 | 0.0104909 |
| VTN | AIRg | 0.67 | 0.0002729 | 0.0147954 |
| VTN | auc_INSULIN | 0.58 | 0.0007263 | 0.0263107 |
| VTN | Glucose_bsl | -0.56 | 0.0011843 | 0.0340405 |
| VTN | HbA1C | -0.40 | 0.0278729 | 0.2212209 |
| VTN | TSH | -0.38 | 0.0376101 | 0.2612506 |
| VWF | auc_glucose | -0.77 | 5.069E-07 | 0.0005463 |
| VWF | Glucose_bsl | -0.76 | 1.283E-06 | 0.0007561 |
| VWF | HbA1C | -0.70 | 1.456E-05 | 0.0024361 |
| VWF | HOMA_IR | -0.55 | 0.0016527 | 0.0414265 |
| VWF | LDL | 0.49 | 0.0074501 | 0.1026104 |
| WDR44 | auc_glucose | 0.70 | 1.747E-05 | 0.0026905 |
| WDR44 | Glucose_bsl | 0.63 | 1.64E-04 | 0.0113861 |
| WDR44 | HbA1C | 0.53 | 0.0027833 | 0.0566055 |
| WDR44 | TSH | 0.43 | 0.0187708 | 0.1785162 |
| WDR44 | DI | -0.44 | 0.0281326 | 0.222141 |
| WDR44 | DEXA__Fat_Mass | 0.39 | 0.0329136 | 0.2410561 |
| ZNF185 | auc_glucose | 0.72 | 6.239E-06 | 0.0016811 |
| ZNF185 | Glucose_bsl | 0.59 | 0.0006036 | 0.0232318 |
| ZNF185 | HbA1C | 0.59 | 0.0006275 | 0.0235948 |
| ZNF185 | DI | -0.61 | 0.0010924 | 0.0325579 |
| ZNF185 | AIRg | -0.47 | 0.0169991 | 0.168094 |
| ZNRD2 | auc_glucose | -0.60 | 0.0004522 | 0.0199337 |
| ZNRD2 | HbA1C | -0.48 | 0.0071931 | 0.1014677 |
| ZNRD2 | Sg | 0.45 | 0.0227485 | 0.1984087 |
| ZNRD2 | Glucose_bsl | -0.39 | 0.0315896 | 0.2360319 |
| ZNRD2 | AIRg | 0.42 | 0.0380642 | 0.2632742 |
| ZYX | auc_glucose | 0.78 | 4.72E-07 | 0.0005463 |
| ZYX | Glucose_bsl | 0.73 | 5.26E-06 | 0.0015122 |
| ZYX | HbA1C | 0.61 | 0.000315 | 0.0162879 |
| ZYX | DI | -0.46 | 0.0222634 | 0.1957903 |
| ZYX | Respiration_Rate | 0.37 | 0.0469496 | 0.2953532 |

**Supplementary Table ST8. List of significant correlation between DE EV proteins and clinical measures**

| **var1** | **var2** | **cor** | **p.value** | **FDR** |
| --- | --- | --- | --- | --- |
| AASDH | auc_glucose | 0.66 | 6.95E-05 | 0.0266984 |
| AASDH | Waist_Circumference | 0.62 | 0.0002491 | 0.0376125 |
| AASDH | HbA1C | 0.56 | 0.0011647 | 0.0649714 |
| AASDH | DEXA_Weight | 0.56 | 0.0014024 | 0.0650071 |
| AASDH | Glucose_bsl | 0.54 | 0.0022244 | 0.0782674 |
| AASDH | DEXA__Lean_Mass | 0.50 | 0.0051484 | 0.1157374 |
| AASDH | DI | -0.42 | 0.0376411 | 0.3212163 |
| AASDH | HDL | -0.38 | 0.0384966 | 0.3236742 |
| AASDH | Temperature | -0.40 | 0.0437275 | 0.344782 |
| ABCA1 | DEXA__Fat_Mass | -0.57 | 0.0010711 | 0.0649714 |
| ABCA1 | DEXA_Fat_Percentage | -0.37 | 0.0458129 | 0.3509813 |
| ACAT2 | auc_glucose | 0.54 | 0.00224 | 0.0782674 |
| ACAT2 | Glucose_bsl | 0.51 | 0.0042296 | 0.1069495 |
| ACAT2 | DEXA__Fat_Mass | 0.45 | 0.0120568 | 0.1783509 |
| ACAT2 | HbA1C | 0.39 | 0.0329459 | 0.3022503 |
| ACE | DEXA__Fat_Mass | 0.46 | 0.0099524 | 0.1628255 |
| ACE | Diastolic_Blood_Pressure_Average | -0.44 | 0.0149416 | 0.1996867 |
| ACHE | Glucose_bsl | -0.59 | 0.0005538 | 0.0500213 |
| ACHE | auc_glucose | -0.54 | 0.0019772 | 0.0740807 |
| ACHE | Heart_Rate_Average | -0.45 | 0.0135806 | 0.1889134 |
| ACHE | DEXA__Fat_Mass | -0.41 | 0.0241195 | 0.2595502 |
| ACHE | HbA1C | -0.38 | 0.0410478 | 0.3343959 |
| ACTA1 | auc_glucose | 0.68 | 4.19E-05 | 0.0244807 |
| ACTA1 | Glucose_bsl | 0.52 | 0.0031273 | 0.0937729 |
| ACTA1 | HbA1C | 0.44 | 0.0147253 | 0.1991444 |
| ACTA1 | DI | -0.45 | 0.0256503 | 0.2678803 |
| ACTBL2 | auc_glucose | 0.61 | 0.000355 | 0.0425831 |
| ACTBL2 | Glucose_bsl | 0.54 | 0.0020881 | 0.0767053 |
| ACTBL2 | HbA1C | 0.39 | 0.0353631 | 0.3117143 |
| ACTC1 | auc_glucose | 0.55 | 0.0016795 | 0.0688865 |
| ACTC1 | Glucose_bsl | 0.50 | 0.0049135 | 0.1156933 |
| ACTC1 | HbA1C | 0.37 | 0.0448312 | 0.3468037 |
| ACTG1 | auc_glucose | 0.65 | 9.02E-05 | 0.0288756 |
| ACTG1 | Glucose_bsl | 0.59 | 0.0006014 | 0.0500213 |
| ACTG1 | HbA1C | 0.51 | 0.0043171 | 0.1083608 |
| ACTG1 | LDL | -0.43 | 0.0201298 | 0.2378052 |
| ADGRL1 | Waist_Circumference | 0.51 | 0.0040213 | 0.1042481 |
| ADGRL1 | HDL | -0.37 | 0.0441779 | 0.345115 |
| AFTPH | auc_glucose | 0.58 | 0.000688 | 0.0525108 |
| AFTPH | HbA1C | 0.49 | 0.0060022 | 0.1258187 |
| AFTPH | Glucose_bsl | 0.44 | 0.0140824 | 0.1940539 |
| AGBL5 | auc_glucose | -0.55 | 0.0017561 | 0.0690509 |
| AGBL5 | Glucose_bsl | -0.48 | 0.0070583 | 0.1377228 |
| AGBL5 | HbA1C | -0.40 | 0.0269428 | 0.2743311 |
| AK9 | Waist_Circumference | 0.43 | 0.0174362 | 0.2174972 |
| AK9 | Glucose_bsl | 0.38 | 0.0368997 | 0.3178999 |
| AKR7A2 | MATSUDA | -0.49 | 0.0065398 | 0.1319006 |
| AKR7A2 | DEXA__Fat_Mass | 0.38 | 0.0367332 | 0.3178999 |
| AKT1 | auc_glucose | -0.64 | 0.0001411 | 0.0344917 |
| AKT1 | Glucose_bsl | -0.57 | 0.0010252 | 0.0639281 |
| AKT1 | HbA1C | -0.45 | 0.012659 | 0.1809996 |
| ALAD | auc_glucose | 0.51 | 0.003997 | 0.1042278 |
| ALAD | HbA1C | 0.47 | 0.0088141 | 0.15655 |
| ALAD | DI | -0.50 | 0.0105122 | 0.1662058 |
| ALAD | Glucose_bsl | 0.40 | 0.0275791 | 0.2786679 |
| ALAD | DEXA__Fat_Mass | 0.38 | 0.0360996 | 0.3151561 |
| ALAD | Waist_Circumference | 0.38 | 0.0366583 | 0.3178999 |
| ALAD | TSH | 0.38 | 0.0382287 | 0.3236742 |
| ALDOC | auc_glucose | 0.48 | 0.0070741 | 0.1377228 |
| ALDOC | HbA1C | 0.39 | 0.0350418 | 0.3101201 |
| ALDOC | auc_c_peptide | -0.50 | 0.0404249 | 0.3317364 |
| ANGPTL6 | auc_glucose | -0.63 | 0.0002183 | 0.0355725 |
| ANGPTL6 | Glucose_bsl | -0.55 | 0.0017183 | 0.0690509 |
| ANGPTL6 | HbA1C | -0.40 | 0.0289805 | 0.2868927 |
| ANGPTL6 | Systolic_Blood_Pressure_Average | 0.40 | 0.0302387 | 0.290967 |
| AP3D1 | Systolic_Blood_Pressure_Average | 0.42 | 0.0217847 | 0.2465322 |
| AP3D1 | auc_INSULIN | -0.39 | 0.0310275 | 0.2938775 |
| AP3D1 | HOMA_B | -0.37 | 0.0445954 | 0.3460245 |
| ARHGAP45 | Heart_Rate_Average | -0.52 | 0.0035419 | 0.0993105 |
| ARHGAP45 | auc_glucose | -0.44 | 0.0149869 | 0.1998119 |
| ARL6IP5 | auc_glucose | 0.61 | 0.0003277 | 0.041706 |
| ARL6IP5 | Glucose_bsl | 0.55 | 0.0017941 | 0.0697277 |
| ARL6IP5 | HbA1C | 0.48 | 0.0067388 | 0.1338112 |
| ARL6IP5 | DEXA__Fat_Mass | 0.39 | 0.0340449 | 0.3049782 |
| ARL6IP5 | HOMA_IR | 0.37 | 0.0440505 | 0.345115 |
| ARMC6 | DEXA__Fat_Mass | 0.47 | 0.0093495 | 0.159257 |
| ARMC6 | Triglycerides | 0.46 | 0.0105235 | 0.1662058 |
| ARMC6 | DEXA_Weight | 0.43 | 0.017227 | 0.2163114 |
| ARMC6 | Waist_Circumference | 0.39 | 0.0344552 | 0.3070374 |
| ARPC2 | auc_glucose | -0.41 | 0.0257407 | 0.2681521 |
| ARPC5 | auc_glucose | 0.66 | 6.82E-05 | 0.0266984 |
| ARPC5 | Glucose_bsl | 0.64 | 0.0001392 | 0.0344917 |
| ARPC5 | HbA1C | 0.55 | 0.0014913 | 0.0658243 |
| ARPC5 | HOMA_IR | 0.39 | 0.0321484 | 0.2997596 |
| ARPC5 | LDL | -0.38 | 0.0395963 | 0.329413 |
| ARPIN | Waist_Circumference | -0.55 | 0.0017802 | 0.0694882 |
| ARPIN | DEXA_Weight | -0.54 | 0.0018841 | 0.0713732 |
| ARPIN | HDL | 0.50 | 0.0044788 | 0.1104921 |
| ARPIN | auc_glucose | -0.42 | 0.019532 | 0.233157 |
| ARPIN | TSH | -0.40 | 0.0292724 | 0.2881211 |
| ARPIN | DEXA__Lean_Mass | -0.39 | 0.0315459 | 0.2957537 |
| ARPIN | DEXA__Fat_Mass | -0.39 | 0.0344841 | 0.3070374 |
| ASAP1 | auc_glucose | -0.57 | 0.0010205 | 0.0639281 |
| ASAP1 | HbA1C | -0.51 | 0.0038367 | 0.1015905 |
| ASAP1 | Glucose_bsl | -0.45 | 0.0120547 | 0.1783509 |
| ASAP1 | Systolic_Blood_Pressure_Average | 0.39 | 0.0335556 | 0.3045814 |
| ASXL3 | auc_glucose | -0.59 | 0.00058 | 0.0500213 |
| ASXL3 | Glucose_bsl | -0.56 | 0.0013898 | 0.0649899 |
| ASXL3 | HbA1C | -0.40 | 0.0304032 | 0.2916211 |
| ATM | Heart_Rate_Average | -0.53 | 0.002459 | 0.0813784 |
| ATM | auc_glucose | -0.45 | 0.0125046 | 0.1809996 |
| ATM | Glucose_bsl | -0.42 | 0.0218333 | 0.2465322 |
| ATP10B | Glucose_bsl | 0.49 | 0.0055913 | 0.1201645 |
| ATP10B | auc_glucose | 0.49 | 0.0059082 | 0.1248021 |
| ATP10B | Sg | -0.50 | 0.0105475 | 0.166329 |
| ATP10B | Waist_Circumference | 0.42 | 0.0202631 | 0.2388305 |
| ATP5MG | auc_glucose | 0.66 | 7.49E-05 | 0.0266984 |
| ATP5MG | Glucose_bsl | 0.60 | 0.0004798 | 0.0483802 |
| ATP5MG | HbA1C | 0.48 | 0.0074499 | 0.1408198 |
| ATP5MG | Heart_Rate_Average | 0.36 | 0.0488341 | 0.3623594 |
| ATP5PF | auc_glucose | 0.42 | 0.0222457 | 0.2498328 |
| ATP5PF | Glucose_bsl | 0.38 | 0.0391518 | 0.3274178 |
| ATP5PF | DI | -0.41 | 0.0442124 | 0.345115 |
| ATP6V0C | auc_glucose | 0.53 | 0.0026207 | 0.0836356 |
| ATP6V0C | Glucose_bsl | 0.51 | 0.0037803 | 0.1015905 |
| ATP6V0C | DEXA__Fat_Mass | 0.38 | 0.0410747 | 0.3343959 |
| ATP6V1G1 | DEXA_Weight | -0.50 | 0.0044476 | 0.1104921 |
| ATP6V1G1 | DEXA__Fat_Mass | -0.46 | 0.0113034 | 0.1726793 |
| ATP6V1G1 | HDL | 0.45 | 0.012466 | 0.1807564 |
| ATP6V1G1 | QUICKI | 0.42 | 0.020871 | 0.2410138 |
| ATP6V1G1 | Waist_Circumference | -0.39 | 0.0322989 | 0.2999408 |
| ATP8A1 | MATSUDA | -0.55 | 0.0015208 | 0.0664575 |
| ATP8A1 | auc_glucose | 0.50 | 0.0051031 | 0.1157374 |
| ATP8A1 | Glucose_bsl | 0.38 | 0.0379624 | 0.3233079 |
| ATRN | auc_glucose | 0.47 | 0.0092926 | 0.159257 |
| ATRN | TSH | 0.45 | 0.0129065 | 0.1832612 |
| ATRN | DEXA_Weight | 0.43 | 0.0164766 | 0.2119394 |
| ATRN | HOMA_IR | 0.40 | 0.0301832 | 0.290967 |
| ATRN | MATSUDA | -0.39 | 0.0328397 | 0.3022503 |
| ATRN | Waist_Circumference | 0.38 | 0.0369428 | 0.3178999 |
| ATRN | Insulin_bsl | 0.37 | 0.0418989 | 0.3370961 |
| ATRN | HDL | -0.37 | 0.046092 | 0.3515474 |
| ATXN7 | auc_glucose | 0.50 | 0.0047039 | 0.1130913 |
| ATXN7 | Glucose_bsl | 0.47 | 0.0086169 | 0.1535778 |
| ATXN7 | Waist_Circumference | 0.37 | 0.0425544 | 0.3401353 |
| BLMH | DEXA__Fat_Mass | 0.61 | 0.0003069 | 0.0403865 |
| BLMH | QUICKI | -0.53 | 0.0026487 | 0.0836356 |
| BLMH | DEXA_Fat_Percentage | 0.51 | 0.0040462 | 0.1043677 |
| BLMH | Si | -0.53 | 0.0062202 | 0.1272053 |
| BLMH | MATSUDA | -0.45 | 0.0122222 | 0.1789919 |
| BLMH | Waist_Circumference | 0.39 | 0.0346748 | 0.3084676 |
| BROX | DEXA__Fat_Mass | 0.57 | 0.0010942 | 0.0649714 |
| BROX | HOMA_IR | 0.50 | 0.0047687 | 0.1141151 |
| BROX | Insulin_bsl | 0.47 | 0.0079934 | 0.1470621 |
| BSG | auc_glucose | 0.56 | 0.0013661 | 0.0649714 |
| BSG | Glucose_bsl | 0.51 | 0.0037893 | 0.1015905 |
| BSG | DEXA__Fat_Mass | 0.48 | 0.0076126 | 0.1423517 |
| BSG | HbA1C | 0.45 | 0.0129414 | 0.1832757 |
| BSG | DEXA_Fat_Percentage | 0.37 | 0.041637 | 0.3361797 |
| C12orf42 | auc_glucose | -0.59 | 0.0005667 | 0.0500213 |
| C12orf42 | Glucose_bsl | -0.55 | 0.0016315 | 0.068365 |
| C12orf42 | HbA1C | -0.38 | 0.036853 | 0.3178999 |
| C1QBP | QUICKI | -0.50 | 0.0051689 | 0.1157374 |
| C1QBP | DEXA__Fat_Mass | 0.46 | 0.0098305 | 0.1620637 |
| C1QBP | MATSUDA | -0.42 | 0.0198328 | 0.2351077 |
| C1QBP | Respiration_Rate | 0.42 | 0.0224838 | 0.2505118 |
| C1QBP | Glucose_bsl | 0.41 | 0.0255796 | 0.2678803 |
| C1QBP | HbA1C | 0.41 | 0.0256065 | 0.2678803 |
| C1QBP | HOMA_IR | 0.40 | 0.0295441 | 0.2889909 |
| C1QBP | Insulin_bsl | 0.39 | 0.0321897 | 0.2998728 |
| C3orf67 | auc_glucose | 0.51 | 0.0036129 | 0.100242 |
| C3orf67 | Glucose_bsl | 0.39 | 0.0332842 | 0.3031901 |
| C3orf67 | HbA1C | 0.37 | 0.0441308 | 0.345115 |
| C5AR1 | auc_glucose | 0.66 | 6.29E-05 | 0.0266984 |
| C5AR1 | Glucose_bsl | 0.60 | 0.0004015 | 0.0463165 |
| C5AR1 | HbA1C | 0.48 | 0.0066764 | 0.1329405 |
| C5AR1 | DEXA__Fat_Mass | 0.46 | 0.0097494 | 0.1616918 |
| C5AR1 | HOMA_IR | 0.41 | 0.0257018 | 0.2681447 |
| C5AR1 | Waist_Circumference | 0.36 | 0.0490678 | 0.3634416 |
| CA1 | auc_glucose | 0.50 | 0.0049928 | 0.1157374 |
| CA1 | AIRg | -0.49 | 0.0120896 | 0.1784699 |
| CA1 | Glucose_bsl | 0.45 | 0.0123958 | 0.1807232 |
| CA1 | DEXA__Lean_Mass | -0.44 | 0.015696 | 0.2061106 |
| CA1 | DI | -0.45 | 0.023645 | 0.2571389 |
| CA1 | DEXA_Fat_Percentage | 0.41 | 0.0248442 | 0.2648503 |
| CA13 | auc_glucose | -0.53 | 0.0025157 | 0.0822484 |
| CA13 | Glucose_bsl | -0.42 | 0.0197839 | 0.2350715 |
| CA13 | HbA1C | -0.40 | 0.0280539 | 0.2804296 |
| CA13 | Waist_Circumference | -0.37 | 0.0456431 | 0.3502661 |
| CACNA1A | auc_glucose | -0.61 | 0.0003461 | 0.0422968 |
| CACNA1A | Glucose_bsl | -0.54 | 0.0022491 | 0.0782674 |
| CACNA1A | HbA1C | -0.38 | 0.0407888 | 0.3340179 |
| CACYBP | Sg | 0.40 | 0.0494349 | 0.3656332 |
| CALML5 | HOMA_B | 0.50 | 0.0045498 | 0.1115441 |
| CALML5 | HDL | -0.49 | 0.0056491 | 0.1208206 |
| CALML5 | auc_INSULIN | 0.42 | 0.0203733 | 0.2395788 |
| CALML5 | Insulin_bsl | 0.42 | 0.0218531 | 0.2465322 |
| CALML5 | auc_glucose | -0.39 | 0.0312711 | 0.294793 |
| CALML5 | QUICKI | -0.39 | 0.0354926 | 0.3117143 |
| CAP1 | auc_glucose | 0.66 | 8.57E-05 | 0.0288756 |
| CAP1 | Glucose_bsl | 0.56 | 0.0014394 | 0.0650391 |
| CAP1 | HbA1C | 0.48 | 0.0074363 | 0.1408198 |
| CAP1 | Respiration_Rate | 0.37 | 0.0494936 | 0.3656336 |
| CAPNS1 | auc_glucose | 0.52 | 0.0033077 | 0.0951158 |
| CAPNS1 | Glucose_bsl | 0.45 | 0.0124544 | 0.1807564 |
| CAPNS1 | DEXA__Lean_Mass | -0.41 | 0.0260352 | 0.269705 |
| CAPNS1 | DEXA_Fat_Percentage | 0.37 | 0.0472021 | 0.3560947 |
| CAPNS1 | HbA1C | 0.36 | 0.0476393 | 0.3585229 |
| CARMIL2 | auc_glucose | -0.60 | 0.0005076 | 0.0486997 |
| CARMIL2 | Glucose_bsl | -0.56 | 0.0013927 | 0.0649899 |
| CARMIL2 | HbA1C | -0.40 | 0.0290148 | 0.2868927 |
| CARMIL2 | LDL | 0.38 | 0.0418241 | 0.3367575 |
| CASQ1 | auc_glucose | 0.54 | 0.0021 | 0.0767053 |
| CASQ1 | HbA1C | 0.49 | 0.006191 | 0.1271133 |
| CASQ1 | Glucose_bsl | 0.48 | 0.0072811 | 0.1395492 |
| CASQ1 | LDL | -0.46 | 0.0121951 | 0.1788499 |
| CCS | Triglycerides | -0.50 | 0.0046219 | 0.112172 |
| CCS | Waist_Circumference | -0.47 | 0.0095928 | 0.1598692 |
| CCS | Temperature | 0.40 | 0.0443887 | 0.345485 |
| CCS | Ins30_glu30 | 0.37 | 0.0446335 | 0.3460245 |
| CCS | DI | 0.40 | 0.0453644 | 0.3491632 |
| CD247 | auc_glucose | -0.59 | 0.0005343 | 0.0500213 |
| CD247 | HbA1C | -0.56 | 0.001446 | 0.0650391 |
| CD247 | Glucose_bsl | -0.53 | 0.0025888 | 0.0833109 |
| CD247 | HOMA_IR | -0.49 | 0.0062791 | 0.1281537 |
| CD247 | LDL | 0.39 | 0.0389566 | 0.3267388 |
| CD300LG | QUICKI | 0.50 | 0.0050217 | 0.1157374 |
| CD300LG | DEXA__Fat_Mass | -0.47 | 0.0086157 | 0.1535778 |
| CD300LG | DEXA_Fat_Percentage | -0.45 | 0.012965 | 0.1833318 |
| CD37 | auc_glucose | -0.56 | 0.0012017 | 0.0649714 |
| CD37 | Glucose_bsl | -0.51 | 0.0038394 | 0.1015905 |
| CD37 | DEXA_Fat_Percentage | -0.41 | 0.0243303 | 0.2609974 |
| CD46 | auc_glucose | 0.56 | 0.0011454 | 0.0649714 |
| CD46 | Glucose_bsl | 0.52 | 0.0031331 | 0.0937729 |
| CD46 | HbA1C | 0.43 | 0.0168496 | 0.2140823 |
| CD46 | HOMA_IR | 0.43 | 0.0186742 | 0.226339 |
| CD46 | Heart_Rate_Average | 0.40 | 0.0270048 | 0.2744867 |
| CD46 | Insulin_bsl | 0.38 | 0.0376381 | 0.3212163 |
| CD55 | auc_glucose | 0.56 | 0.0013143 | 0.0649714 |
| CD55 | Glucose_bsl | 0.52 | 0.0031664 | 0.093781 |
| CD55 | DEXA__Fat_Mass | 0.48 | 0.0073779 | 0.1405228 |
| CD55 | HbA1C | 0.44 | 0.0151645 | 0.2008754 |
| CD55 | Waist_Circumference | 0.41 | 0.0251616 | 0.2660239 |
| CEMIP2 | auc_glucose | -0.50 | 0.0052383 | 0.1157374 |
| CEMIP2 | Glucose_bsl | -0.48 | 0.0066152 | 0.1329405 |
| CEMIP2 | DEXA__Fat_Mass | -0.47 | 0.0082341 | 0.1491405 |
| CEMIP2 | HbA1C | -0.43 | 0.0183559 | 0.2246032 |
| CEMIP2 | Waist_Circumference | -0.41 | 0.0249396 | 0.2651852 |
| CEP192 | auc_glucose | -0.55 | 0.0015407 | 0.0664575 |
| CEP192 | Glucose_bsl | -0.53 | 0.0026625 | 0.0836356 |
| CEP192 | DEXA__Fat_Mass | -0.37 | 0.0425749 | 0.3401353 |
| CEP192 | Heart_Rate_Average | -0.37 | 0.0466168 | 0.3542328 |
| CFHR5 | DI | 0.45 | 0.0232482 | 0.254441 |
| CFHR5 | auc_c_peptide | 0.54 | 0.0265906 | 0.2732061 |
| CFHR5 | Glucose_bsl | -0.37 | 0.043906 | 0.345115 |
| CHAMP1 | auc_glucose | -0.63 | 0.0002008 | 0.0355725 |
| CHAMP1 | Glucose_bsl | -0.57 | 0.0011118 | 0.0649714 |
| CHAMP1 | HbA1C | -0.40 | 0.0291592 | 0.2872829 |
| CHAMP1 | Heart_Rate_Average | -0.38 | 0.0367122 | 0.3178999 |
| CHMP6 | auc_glucose | 0.56 | 0.0014508 | 0.0650391 |
| CHMP6 | Glucose_bsl | 0.48 | 0.0071969 | 0.1386185 |
| CHMP6 | HbA1C | 0.40 | 0.0293566 | 0.2882921 |
| CHRNA4 | auc_glucose | -0.56 | 0.0013206 | 0.0649714 |
| CHRNA4 | Glucose_bsl | -0.53 | 0.0026415 | 0.0836356 |
| CHRNA4 | HbA1C | -0.46 | 0.0106671 | 0.167958 |
| CHRNA4 | LDL | 0.39 | 0.0339636 | 0.3049782 |
| CHST7 | auc_glucose | -0.66 | 7.54E-05 | 0.0266984 |
| CHST7 | Glucose_bsl | -0.57 | 0.0011073 | 0.0649714 |
| CHST7 | HbA1C | -0.50 | 0.0050862 | 0.1157374 |
| CHST7 | AIRg | 0.48 | 0.0162573 | 0.2101496 |
| CLASP2 | auc_glucose | -0.58 | 0.0006905 | 0.0525108 |
| CLASP2 | Glucose_bsl | -0.56 | 0.00132 | 0.0649714 |
| CLASP2 | HbA1C | -0.40 | 0.0272941 | 0.2766055 |
| CLASP2 | DEXA__Fat_Mass | -0.37 | 0.0452597 | 0.3490882 |
| CLDN5 | Glucose_bsl | 0.58 | 0.0008105 | 0.057382 |
| CLDN5 | auc_glucose | 0.57 | 0.0010053 | 0.0637593 |
| CLDN5 | DEXA__Fat_Mass | 0.44 | 0.0151913 | 0.2009721 |
| CLDN5 | DEXA_Fat_Percentage | 0.40 | 0.028388 | 0.282121 |
| CLEC1B | auc_glucose | 0.44 | 0.0139939 | 0.1933526 |
| CLEC1B | Si | -0.48 | 0.0150936 | 0.2005235 |
| CLEC1B | Temperature | -0.44 | 0.0238448 | 0.2581288 |
| CLTB | auc_glucose | 0.57 | 0.0010061 | 0.0637593 |
| CLTB | Glucose_bsl | 0.53 | 0.0023847 | 0.0802676 |
| CLTB | Waist_Circumference | 0.52 | 0.0032943 | 0.0951158 |
| CLTB | HbA1C | 0.52 | 0.0035708 | 0.0996146 |
| CLTB | DEXA__Fat_Mass | 0.47 | 0.0082971 | 0.1496983 |
| CLTB | DEXA_Weight | 0.44 | 0.0138466 | 0.1915751 |
| CLTB | HDL | -0.42 | 0.0212935 | 0.2434285 |
| CLTB | HOMA_IR | 0.38 | 0.0381122 | 0.3236742 |
| CMTM6 | auc_glucose | 0.56 | 0.0012315 | 0.0649714 |
| CMTM6 | Glucose_bsl | 0.54 | 0.001898 | 0.071635 |
| CMTM6 | MATSUDA | -0.45 | 0.0118611 | 0.176847 |
| CMTM6 | DEXA__Fat_Mass | 0.37 | 0.0453715 | 0.3491632 |
| CNN2 | auc_glucose | 0.58 | 0.0008408 | 0.0586257 |
| CNN2 | Glucose_bsl | 0.54 | 0.0021471 | 0.0770705 |
| CNN2 | HbA1C | 0.51 | 0.0037326 | 0.1015759 |
| CNN2 | DEXA__Fat_Mass | 0.48 | 0.0070968 | 0.1377228 |
| CNN2 | HOMA_IR | 0.46 | 0.0114699 | 0.1741857 |
| CNN2 | Insulin_bsl | 0.43 | 0.0169754 | 0.214439 |
| CNTN1 | DEXA_Weight | 0.53 | 0.0026444 | 0.0836356 |
| CNTN1 | DEXA__Fat_Mass | 0.38 | 0.03587 | 0.3141995 |
| CNTN1 | DEXA__Lean_Mass | 0.38 | 0.0388284 | 0.3259952 |
| COL22A1 | auc_glucose | -0.59 | 0.0005834 | 0.0500213 |
| COL22A1 | Glucose_bsl | -0.57 | 0.0010755 | 0.0649714 |
| COL22A1 | HbA1C | -0.43 | 0.0186346 | 0.2261257 |
| COL6A3 | DEXA_Weight | -0.53 | 0.002664 | 0.0836356 |
| COL6A3 | DEXA__Fat_Mass | -0.50 | 0.0051723 | 0.1157374 |
| COL6A3 | HOMA_IR | -0.40 | 0.0302417 | 0.290967 |
| COL6A3 | Glucose_bsl | -0.39 | 0.0310309 | 0.2938775 |
| COL6A3 | Insulin_bsl | -0.37 | 0.0443076 | 0.345115 |
| COLEC10 | auc_glucose | -0.70 | 1.95E-05 | 0.0200487 |
| COLEC10 | Glucose_bsl | -0.59 | 0.0006003 | 0.0500213 |
| COLEC10 | HbA1C | -0.45 | 0.0136257 | 0.1892842 |
| COLEC10 | auc_c_peptide | 0.51 | 0.0379806 | 0.3233079 |
| COLEC10 | TSH | -0.38 | 0.0401962 | 0.3311831 |
| COLEC11 | auc_glucose | -0.56 | 0.0012093 | 0.0649714 |
| COLEC11 | Glucose_bsl | -0.43 | 0.0166882 | 0.2124584 |
| COX4I1 | auc_glucose | 0.55 | 0.0018067 | 0.0697277 |
| COX4I1 | Glucose_bsl | 0.51 | 0.0043005 | 0.1083608 |
| COX4I1 | DEXA__Fat_Mass | 0.43 | 0.0189033 | 0.2283077 |
| COX4I1 | Waist_Circumference | 0.42 | 0.020775 | 0.2406903 |
| COX4I1 | HbA1C | 0.39 | 0.0329309 | 0.3022503 |
| COX4I1 | Heart_Rate_Average | 0.37 | 0.0421447 | 0.3382781 |
| COX5B | auc_glucose | 0.51 | 0.0041659 | 0.1061023 |
| COX5B | MATSUDA | -0.46 | 0.0110566 | 0.1712859 |
| COX5B | Glucose_bsl | 0.45 | 0.0118181 | 0.176847 |
| COX5B | DEXA__Fat_Mass | 0.37 | 0.0429572 | 0.3418596 |
| COX7A2 | auc_glucose | 0.49 | 0.0059774 | 0.1257449 |
| COX7A2 | Glucose_bsl | 0.38 | 0.0369138 | 0.3178999 |
| CPM | auc_glucose | -0.57 | 0.0009418 | 0.0615845 |
| CPM | Respiration_Rate | -0.54 | 0.0024219 | 0.0812522 |
| CPM | Glucose_bsl | -0.46 | 0.0100281 | 0.1628255 |
| CPM | HbA1C | -0.36 | 0.048779 | 0.3623594 |
| CPN1 | auc_glucose | -0.59 | 0.0006139 | 0.0500213 |
| CPN1 | Glucose_bsl | -0.56 | 0.0014254 | 0.0650391 |
| CPN1 | HOMA_IR | -0.41 | 0.0229926 | 0.2535364 |
| CPN1 | HbA1C | -0.40 | 0.0276859 | 0.2787634 |
| CPN1 | Heart_Rate_Average | -0.40 | 0.030443 | 0.2916211 |
| CPT1A | DEXA_Weight | 0.54 | 0.0020682 | 0.0763761 |
| CPT1A | auc_INSULIN | 0.48 | 0.0073391 | 0.1403038 |
| CPT1A | Waist_Circumference | 0.46 | 0.0112228 | 0.1724743 |
| CPT1A | DEXA__Lean_Mass | 0.44 | 0.0157858 | 0.2069695 |
| CPT1A | Insulin_bsl | 0.42 | 0.019499 | 0.2330342 |
| CPT1A | Respiration_Rate | 0.43 | 0.019626 | 0.2334652 |
| CPT1A | HOMA_B | 0.42 | 0.0209236 | 0.2413497 |
| CPT1A | MATSUDA | -0.37 | 0.0412754 | 0.3349669 |
| CPT1A | HDL | -0.37 | 0.0442946 | 0.345115 |
| CPT1C | auc_glucose | -0.52 | 0.0032884 | 0.0951158 |
| CPT1C | auc_INSULIN | 0.46 | 0.0100142 | 0.1628255 |
| CPT1C | HOMA_B | 0.44 | 0.0144049 | 0.1966504 |
| CPT1C | AIRg | 0.47 | 0.0187219 | 0.2266501 |
| CPT1C | Glucose_bsl | -0.42 | 0.022539 | 0.2505118 |
| CPT1C | HbA1C | -0.39 | 0.0338343 | 0.3049782 |
| CPT1C | Sg | 0.40 | 0.0455445 | 0.3502317 |
| CRISPLD2 | auc_glucose | -0.56 | 0.0011753 | 0.0649714 |
| CRISPLD2 | Glucose_bsl | -0.48 | 0.0068414 | 0.1346977 |
| CRISPLD2 | DEXA__Lean_Mass | 0.43 | 0.0170031 | 0.214439 |
| CRISPLD2 | DEXA_Fat_Percentage | -0.40 | 0.0268557 | 0.2743311 |
| CRISPLD2 | HbA1C | -0.36 | 0.0475179 | 0.3579007 |
| CRKL | auc_glucose | -0.58 | 0.0007276 | 0.0540857 |
| CRKL | HbA1C | -0.58 | 0.0008067 | 0.057382 |
| CRKL | Glucose_bsl | -0.56 | 0.0012643 | 0.0649714 |
| CRKL | DEXA_Fat_Percentage | -0.42 | 0.0216441 | 0.2463397 |
| CRKL | HOMA_IR | -0.41 | 0.0247834 | 0.2647046 |
| CRKL | DEXA__Fat_Mass | -0.40 | 0.0298249 | 0.2899454 |
| CRKL | Systolic_Blood_Pressure_Average | 0.38 | 0.0381363 | 0.3236742 |
| CROCC | auc_glucose | 0.59 | 0.0006696 | 0.0520753 |
| CROCC | Glucose_bsl | 0.55 | 0.0017623 | 0.0690509 |
| CROCC | HbA1C | 0.53 | 0.0023256 | 0.0793185 |
| CROCC | Waist_Circumference | 0.51 | 0.0037158 | 0.1015759 |
| CROCC | DEXA__Fat_Mass | 0.51 | 0.003939 | 0.1036863 |
| CROCC | DEXA_Weight | 0.47 | 0.0080897 | 0.1480368 |
| CROCC | HOMA_IR | 0.41 | 0.0260174 | 0.269705 |
| CROCC | HDL | -0.40 | 0.0304518 | 0.2916211 |
| CROCC | Insulin_bsl | 0.37 | 0.0450298 | 0.3478372 |
| CS | auc_glucose | 0.48 | 0.0075502 | 0.1420816 |
| CS | AIRg | -0.42 | 0.0368344 | 0.3178999 |
| CST6 | auc_glucose | 0.60 | 0.0004864 | 0.0483802 |
| CST6 | Glucose_bsl | 0.55 | 0.001604 | 0.0678718 |
| CST6 | HbA1C | 0.50 | 0.0052041 | 0.1157374 |
| CSTB | Triglycerides | -0.53 | 0.0026263 | 0.0836356 |
| CSTB | DEXA__Fat_Mass | -0.42 | 0.0199996 | 0.2365396 |
| CSTB | DEXA_Weight | -0.42 | 0.0222035 | 0.2496612 |
| CTSH | auc_glucose | -0.60 | 0.0004995 | 0.0483802 |
| CTSH | HbA1C | -0.58 | 0.0008017 | 0.057382 |
| CTSH | Glucose_bsl | -0.55 | 0.0014726 | 0.0657273 |
| CTSH | DI | 0.49 | 0.0131497 | 0.1851784 |
| CYB5R1 | auc_glucose | -0.47 | 0.0094482 | 0.159257 |
| CYB5R1 | Glucose_bsl | -0.44 | 0.016036 | 0.2085655 |
| CYB5R1 | DEXA__Fat_Mass | -0.42 | 0.0219855 | 0.2477537 |
| CYB5R1 | HbA1C | -0.36 | 0.0496546 | 0.3659398 |
| CYP27A1 | auc_glucose | 0.53 | 0.0024713 | 0.0813784 |
| CYP27A1 | Glucose_bsl | 0.40 | 0.0298628 | 0.2900394 |
| CYP27A1 | Triglycerides | 0.36 | 0.0477053 | 0.3585229 |
| CYP2B6 | auc_glucose | -0.64 | 0.0001369 | 0.0344917 |
| CYP2B6 | AIRg | 0.62 | 0.0008941 | 0.0595715 |
| CYP2B6 | Glucose_bsl | -0.54 | 0.0021149 | 0.0767053 |
| CYP2B6 | HbA1C | -0.48 | 0.0077831 | 0.1437078 |
| CYP2B6 | Sg | 0.48 | 0.014831 | 0.1992869 |
| CYP2B6 | Waist_Circumference | -0.43 | 0.017603 | 0.2185153 |
| CYP2B6 | HDL | 0.39 | 0.0318247 | 0.2975521 |
| CYP2F1 | auc_glucose | 0.50 | 0.0053662 | 0.1169622 |
| CYP2F1 | Glucose_bsl | 0.43 | 0.0163787 | 0.2109711 |
| CYP4F11 | auc_glucose | 0.55 | 0.0016963 | 0.0688865 |
| CYP4F11 | DEXA_Weight | 0.44 | 0.014771 | 0.1991444 |
| CYP4F11 | HOMA_IR | 0.41 | 0.0236305 | 0.2571389 |
| CYP4F11 | Glucose_bsl | 0.41 | 0.0252049 | 0.2662075 |
| CYP4F11 | Respiration_Rate | 0.39 | 0.0347341 | 0.3087275 |
| CYP4F11 | DEXA__Lean_Mass | 0.39 | 0.0350202 | 0.3101201 |
| CYP4F11 | HbA1C | 0.37 | 0.044063 | 0.345115 |
| DAB2 | Waist_Circumference | -0.60 | 0.0004896 | 0.0483802 |
| DAB2 | HbA1C | -0.52 | 0.0030867 | 0.0934195 |
| DAB2 | DEXA_Weight | -0.50 | 0.0046862 | 0.1129307 |
| DAB2 | DEXA__Fat_Mass | -0.45 | 0.0116575 | 0.1765125 |
| DAB2 | auc_glucose | -0.45 | 0.0124285 | 0.1807232 |
| DAB2 | Glucose_bsl | -0.45 | 0.0133624 | 0.186892 |
| DAB2 | HOMA_IR | -0.41 | 0.0232829 | 0.2545495 |
| DAB2 | HDL | 0.41 | 0.0233993 | 0.2552341 |
| DAB2 | Insulin_bsl | -0.38 | 0.0395433 | 0.329413 |
| DAB2 | QUICKI | 0.37 | 0.0471372 | 0.3560947 |
| DDAH2 | auc_glucose | 0.49 | 0.0055891 | 0.1201645 |
| DDAH2 | Glucose_bsl | 0.38 | 0.0383132 | 0.3236742 |
| DDAH2 | AIRg | -0.41 | 0.0441963 | 0.345115 |
| DDT | auc_glucose | 0.88 | 1.50E-10 | 1.54E-06 |
| DDT | HbA1C | 0.74 | 3.69E-06 | 0.0075796 |
| DDT | Glucose_bsl | 0.71 | 9.34E-06 | 0.0159773 |
| DDT | AIRg | -0.52 | 0.0076833 | 0.1428945 |
| DDT | DI | -0.51 | 0.0093914 | 0.159257 |
| DDT | Sg | -0.49 | 0.0135591 | 0.1889134 |
| DDT | HOMA_B | -0.37 | 0.044629 | 0.3460245 |
| DDX3X | auc_glucose | -0.68 | 3.67E-05 | 0.0244807 |
| DDX3X | Glucose_bsl | -0.53 | 0.0026835 | 0.083991 |
| DDX3X | HbA1C | -0.40 | 0.0279493 | 0.2799297 |
| DNAJA2 | Waist_Circumference | -0.50 | 0.0044881 | 0.1104921 |
| DNAJA2 | AIRg | 0.46 | 0.0198113 | 0.2351077 |
| DNAJA2 | DI | 0.46 | 0.0215014 | 0.2452596 |
| DNAJA2 | HbA1C | -0.41 | 0.0226207 | 0.2507823 |
| DNAJA2 | auc_glucose | -0.41 | 0.0229142 | 0.2529436 |
| DNAJA2 | Triglycerides | -0.39 | 0.0312453 | 0.294793 |
| DNAJA2 | Respiration_Rate | -0.40 | 0.0338606 | 0.3049782 |
| DNAJA2 | HDL | 0.39 | 0.034208 | 0.3053732 |
| DNAJA2 | Glucose_bsl | -0.39 | 0.0349799 | 0.3101064 |
| DOCK10 | auc_glucose | -0.59 | 0.0006054 | 0.0500213 |
| DOCK10 | Glucose_bsl | -0.54 | 0.002218 | 0.0782674 |
| DOCK10 | HbA1C | -0.39 | 0.0315438 | 0.2957537 |
| DOCK7 | auc_glucose | 0.59 | 0.0005439 | 0.0500213 |
| DOCK7 | Glucose_bsl | 0.51 | 0.0040633 | 0.104547 |
| DOCK7 | HbA1C | 0.43 | 0.0184046 | 0.2247543 |
| DRC7 | auc_glucose | -0.52 | 0.0033975 | 0.0966174 |
| DRC7 | Glucose_bsl | -0.40 | 0.027898 | 0.2796888 |
| DRC7 | HbA1C | -0.40 | 0.0302242 | 0.290967 |
| DUSP3 | AIRg | -0.54 | 0.0052356 | 0.1157374 |
| DUSP3 | auc_glucose | 0.49 | 0.0057515 | 0.1222452 |
| DUSP3 | DEXA__Lean_Mass | -0.40 | 0.027684 | 0.2787634 |
| DUSP3 | Glucose_bsl | 0.37 | 0.0426293 | 0.3402009 |
| DUSP3 | Ins30_glu30 | -0.37 | 0.0432599 | 0.3429389 |
| DUSP3 | auc_c_peptide | -0.49 | 0.0469135 | 0.3556091 |
| EARS2 | auc_glucose | 0.74 | 2.64E-06 | 0.0067654 |
| EARS2 | Glucose_bsl | 0.70 | 1.89E-05 | 0.0200487 |
| EARS2 | HbA1C | 0.59 | 0.0006693 | 0.0520753 |
| EARS2 | DI | -0.40 | 0.0453369 | 0.3491632 |
| ECM1 | auc_glucose | -0.63 | 0.0002021 | 0.0355725 |
| ECM1 | Glucose_bsl | -0.56 | 0.0014327 | 0.0650391 |
| ECM1 | HbA1C | -0.42 | 0.020791 | 0.2406903 |
| EHD3 | auc_glucose | -0.46 | 0.0110208 | 0.1711638 |
| EHD3 | HbA1C | -0.39 | 0.0352756 | 0.3117143 |
| EHD3 | Glucose_bsl | -0.39 | 0.0354211 | 0.3117143 |
| EMILIN1 | auc_glucose | -0.52 | 0.0030465 | 0.0925306 |
| EMILIN1 | Glucose_bsl | -0.48 | 0.0070896 | 0.1377228 |
| EMILIN1 | HbA1C | -0.40 | 0.0281733 | 0.2805768 |
| EMILIN1 | MATSUDA | 0.36 | 0.0480533 | 0.3598213 |
| EMILIN1 | HOMA_IR | -0.36 | 0.0483846 | 0.3612482 |
| EML6 | auc_glucose | 0.54 | 0.0019645 | 0.073875 |
| EML6 | Glucose_bsl | 0.51 | 0.0039759 | 0.1042278 |
| EML6 | DEXA__Fat_Mass | 0.41 | 0.0230298 | 0.2536734 |
| EXOC3L4 | auc_glucose | 0.57 | 0.0011345 | 0.0649714 |
| EXOC3L4 | Glucose_bsl | 0.46 | 0.0103127 | 0.164907 |
| F13A1 | auc_glucose | -0.59 | 0.0005952 | 0.0500213 |
| F13A1 | Glucose_bsl | -0.55 | 0.0017226 | 0.0690509 |
| F13A1 | DEXA__Fat_Mass | -0.42 | 0.0208705 | 0.2410138 |
| F13A1 | HbA1C | -0.38 | 0.036724 | 0.3178999 |
| FAM114A2 | auc_glucose | 0.45 | 0.0136694 | 0.1896349 |
| FAM114A2 | Glucose_bsl | 0.42 | 0.0203211 | 0.2392393 |
| FAM114A2 | HbA1C | 0.37 | 0.0422598 | 0.3384078 |
| FAM98A | Glucose_bsl | -0.50 | 0.0050571 | 0.1157374 |
| FAM98A | auc_glucose | -0.48 | 0.0074486 | 0.1408198 |
| FASN | auc_glucose | 0.50 | 0.0053122 | 0.1166323 |
| FASN | Glucose_bsl | 0.47 | 0.0090113 | 0.1581892 |
| FBLN2 | MATSUDA | 0.42 | 0.0207192 | 0.2406903 |
| FBLN2 | QUICKI | 0.42 | 0.0218408 | 0.2465322 |
| FBLN2 | Temperature | 0.40 | 0.0442971 | 0.345115 |
| FBN3 | auc_glucose | -0.60 | 0.0004394 | 0.0477167 |
| FBN3 | Glucose_bsl | -0.53 | 0.0025575 | 0.0826354 |
| FBN3 | HbA1C | -0.38 | 0.0384454 | 0.3236742 |
| FCER1G | Glucose_bsl | 0.53 | 0.0025595 | 0.0826354 |
| FCER1G | DEXA__Fat_Mass | 0.52 | 0.0031543 | 0.093781 |
| FCER1G | auc_glucose | 0.47 | 0.00957 | 0.1597494 |
| FCER1G | HbA1C | 0.38 | 0.0403834 | 0.3317364 |
| FGA | auc_glucose | -0.61 | 0.0003567 | 0.0425831 |
| FGA | Glucose_bsl | -0.57 | 0.0011079 | 0.0649714 |
| FGA | HbA1C | -0.45 | 0.0131808 | 0.1851964 |
| FGA | HOMA_IR | -0.42 | 0.0225003 | 0.2505118 |
| FGA | DEXA__Fat_Mass | -0.39 | 0.0336939 | 0.3047945 |
| FGB | auc_glucose | -0.62 | 0.0002845 | 0.0403865 |
| FGB | Glucose_bsl | -0.58 | 0.0008635 | 0.0587056 |
| FGB | HbA1C | -0.44 | 0.014801 | 0.1991444 |
| FGG | auc_glucose | -0.59 | 0.0006384 | 0.0509925 |
| FGG | Glucose_bsl | -0.56 | 0.0013353 | 0.0649714 |
| FGG | HbA1C | -0.42 | 0.0205097 | 0.2402263 |
| FGL1 | auc_glucose | -0.61 | 0.0002987 | 0.0403865 |
| FGL1 | Glucose_bsl | -0.55 | 0.0017442 | 0.0690509 |
| FGL1 | HbA1C | -0.40 | 0.0301093 | 0.290967 |
| FHOD1 | auc_glucose | -0.59 | 0.0005885 | 0.0500213 |
| FHOD1 | Glucose_bsl | -0.49 | 0.0060761 | 0.1264891 |
| FKBP3 | Systolic_Blood_Pressure_Average | -0.39 | 0.0336567 | 0.3047945 |
| FLG2 | auc_glucose | -0.51 | 0.0038991 | 0.1028994 |
| FLG2 | Glucose_bsl | -0.49 | 0.0061369 | 0.1271133 |
| FLG2 | Heart_Rate_Average | -0.39 | 0.0328838 | 0.3022503 |
| FN1 | auc_glucose | -0.67 | 4.29E-05 | 0.0244807 |
| FN1 | Glucose_bsl | -0.57 | 0.0009939 | 0.0637593 |
| FN1 | HbA1C | -0.41 | 0.0238617 | 0.2581288 |
| FREM3 | MATSUDA | -0.49 | 0.0061817 | 0.1271133 |
| FREM3 | Glucose_bsl | 0.46 | 0.0101191 | 0.1628255 |
| FREM3 | auc_glucose | 0.45 | 0.0132321 | 0.1855749 |
| FREM3 | DEXA__Fat_Mass | 0.39 | 0.0323351 | 0.2999408 |
| FREM3 | DEXA_Fat_Percentage | 0.38 | 0.0404071 | 0.3317364 |
| GAPVD1 | auc_glucose | -0.41 | 0.0248048 | 0.2647046 |
| GCLC | auc_glucose | 0.63 | 0.0002003 | 0.0355725 |
| GCLC | Glucose_bsl | 0.59 | 0.000636 | 0.0509925 |
| GCLC | HbA1C | 0.54 | 0.0021295 | 0.0767053 |
| GCLC | DEXA__Fat_Mass | 0.49 | 0.0061669 | 0.1271133 |
| GET3 | auc_glucose | 0.49 | 0.0056067 | 0.1201645 |
| GET3 | Glucose_bsl | 0.46 | 0.0112525 | 0.1725156 |
| GET3 | Diastolic_Blood_Pressure_Average | 0.41 | 0.0250306 | 0.2651852 |
| GET3 | auc_c_peptide | -0.50 | 0.0392653 | 0.3277218 |
| GNG11 | auc_glucose | 0.62 | 0.0002831 | 0.0403865 |
| GNG11 | Glucose_bsl | 0.58 | 0.0006994 | 0.052794 |
| GNG11 | HbA1C | 0.45 | 0.0119691 | 0.1775781 |
| GNG11 | Respiration_Rate | 0.41 | 0.0272752 | 0.2766055 |
| GNPTAB | HbA1C | -0.52 | 0.003014 | 0.0922732 |
| GNPTAB | DI | 0.55 | 0.0041049 | 0.1050905 |
| GNPTAB | auc_glucose | -0.50 | 0.0046355 | 0.1122358 |
| GNPTAB | Glucose_bsl | -0.48 | 0.0076918 | 0.1428945 |
| GNPTAB | AIRg | 0.46 | 0.0222844 | 0.2498328 |
| GNPTAB | Triglycerides | -0.40 | 0.0265715 | 0.2732061 |
| GNPTAB | Sg | 0.43 | 0.0340435 | 0.3049782 |
| GNPTG | Insulin_bsl | 0.52 | 0.0033053 | 0.0951158 |
| GNPTG | auc_INSULIN | 0.48 | 0.0066277 | 0.1329405 |
| GNPTG | QUICKI | -0.47 | 0.0088972 | 0.1572097 |
| GNPTG | HDL | -0.43 | 0.0172358 | 0.2163114 |
| GNPTG | HOMA_IR | 0.43 | 0.0177506 | 0.2195471 |
| GNPTG | Heart_Rate_Average | 0.42 | 0.0209842 | 0.2415061 |
| GNPTG | HOMA_B | 0.41 | 0.0250017 | 0.2651852 |
| GNPTG | MATSUDA | -0.38 | 0.0362212 | 0.315928 |
| GP1BA | auc_glucose | 0.65 | 9.56E-05 | 0.0288756 |
| GP1BA | Glucose_bsl | 0.56 | 0.0012916 | 0.0649714 |
| GP1BA | HbA1C | 0.45 | 0.0126253 | 0.1809996 |
| GP1BA | Heart_Rate_Average | 0.40 | 0.0281164 | 0.2805768 |
| GP5 | auc_glucose | -0.68 | 3.18E-05 | 0.0244807 |
| GP5 | Glucose_bsl | -0.57 | 0.0009058 | 0.0596079 |
| GP5 | HbA1C | -0.47 | 0.0092506 | 0.159257 |
| GPD1L | auc_glucose | 0.47 | 0.0094707 | 0.159257 |
| GPD1L | auc_c_peptide | -0.59 | 0.01356 | 0.1889134 |
| GPD1L | DI | -0.47 | 0.0171355 | 0.2158442 |
| GPD1L | Glucose_bsl | 0.40 | 0.0266508 | 0.2732061 |
| GPD1L | Sg | -0.43 | 0.030293 | 0.2911872 |
| GPD1L | AIRg | -0.43 | 0.0337156 | 0.3047945 |
| GPX1 | auc_glucose | -0.55 | 0.0018057 | 0.0697277 |
| GPX1 | Glucose_bsl | -0.49 | 0.0064894 | 0.131295 |
| GPX1 | Heart_Rate_Average | -0.36 | 0.0482365 | 0.3605789 |
| GPX4 | HOMA_B | -0.40 | 0.027529 | 0.2784357 |
| GPX4 | auc_INSULIN | -0.39 | 0.032253 | 0.2999408 |
| GRN | TSH | 0.38 | 0.0372352 | 0.3196128 |
| GSS | DEXA_Weight | -0.60 | 0.0004448 | 0.0477167 |
| GSS | HOMA_IR | -0.49 | 0.006497 | 0.131295 |
| GSS | Waist_Circumference | -0.45 | 0.0121517 | 0.1787236 |
| GSS | Insulin_bsl | -0.44 | 0.0145054 | 0.197497 |
| GSS | DEXA__Lean_Mass | -0.44 | 0.0162432 | 0.2101496 |
| GSS | DEXA__Fat_Mass | -0.43 | 0.0177663 | 0.2195471 |
| GSS | HDL | 0.39 | 0.0331242 | 0.3024743 |
| GSS | Triglycerides | -0.38 | 0.0366309 | 0.3178999 |
| H2BC1 | auc_glucose | -0.59 | 0.0006628 | 0.0520753 |
| H2BC1 | Glucose_bsl | -0.55 | 0.001621 | 0.0681998 |
| H2BC1 | HbA1C | -0.40 | 0.029335 | 0.2882921 |
| HBA1 | auc_glucose | 0.51 | 0.0040149 | 0.1042481 |
| HBA1 | Glucose_bsl | 0.42 | 0.0204892 | 0.2402263 |
| HBQ1 | Diastolic_Blood_Pressure_Average | -0.43 | 0.0183049 | 0.2244628 |
| HBQ1 | Insulin_bsl | 0.42 | 0.0210638 | 0.2421509 |
| HBQ1 | DEXA__Fat_Mass | 0.41 | 0.0256388 | 0.2678803 |
| HBQ1 | HOMA_IR | 0.40 | 0.0300501 | 0.290967 |
| HCLS1 | DEXA__Fat_Mass | -0.45 | 0.0121439 | 0.1787236 |
| HCLS1 | HOMA_B | -0.44 | 0.0151492 | 0.2008754 |
| HCLS1 | auc_INSULIN | -0.43 | 0.0185339 | 0.2255363 |
| HCLS1 | Ins30_glu30 | 0.37 | 0.0460336 | 0.3515474 |
| HEBP1 | Heart_Rate_Average | 0.56 | 0.0012338 | 0.0649714 |
| HEBP1 | auc_glucose | 0.46 | 0.011367 | 0.1733937 |
| HEBP1 | Glucose_bsl | 0.37 | 0.0414826 | 0.33594 |
| HMBS | auc_glucose | -0.63 | 0.0001722 | 0.0355725 |
| HMBS | Glucose_bsl | -0.55 | 0.0016939 | 0.0688865 |
| HMBS | HbA1C | -0.50 | 0.0051789 | 0.1157374 |
| HMBS | Waist_Circumference | -0.44 | 0.0141924 | 0.1947855 |
| HMBS | HOMA_IR | -0.40 | 0.0305291 | 0.2920889 |
| HPSE | auc_glucose | -0.67 | 5.94E-05 | 0.0266984 |
| HPSE | Glucose_bsl | -0.58 | 0.0008452 | 0.0586257 |
| HPSE | HbA1C | -0.46 | 0.0114449 | 0.1740644 |
| HPSE | LDL | 0.38 | 0.042747 | 0.3404503 |
| HS6ST3 | DEXA_Weight | -0.41 | 0.0242999 | 0.2609441 |
| HS6ST3 | TSH | -0.38 | 0.040133 | 0.3310603 |
| HS6ST3 | Heart_Rate_Average | -0.36 | 0.0498503 | 0.3666084 |
| HSD17B11 | Insulin_bsl | -0.52 | 0.0034951 | 0.0985427 |
| HSD17B11 | HOMA_IR | -0.50 | 0.0048557 | 0.1148581 |
| HSD17B11 | QUICKI | 0.38 | 0.0365001 | 0.3178999 |
| HSD17B11 | Waist_Circumference | -0.38 | 0.0396712 | 0.3296245 |
| HSPA9 | auc_glucose | -0.48 | 0.0079478 | 0.1464857 |
| HSPA9 | Sg | 0.43 | 0.0337572 | 0.3047945 |
| HSPA9 | HbA1C | -0.36 | 0.0482598 | 0.3605789 |
| HSPG2 | Waist_Circumference | -0.51 | 0.0036647 | 0.1014068 |
| HSPG2 | HDL | 0.47 | 0.0090297 | 0.1581892 |
| HSPG2 | Insulin_bsl | -0.40 | 0.0284244 | 0.2822094 |
| HSPG2 | QUICKI | 0.39 | 0.0333187 | 0.3032355 |
| ICAM1 | auc_glucose | 0.63 | 0.0002029 | 0.0355725 |
| ICAM1 | Glucose_bsl | 0.60 | 0.0004695 | 0.0483802 |
| ICAM1 | HbA1C | 0.45 | 0.0125444 | 0.1809996 |
| ICAM1 | TSH | 0.43 | 0.0172822 | 0.2166286 |
| ICAM1 | DEXA__Fat_Mass | 0.43 | 0.018094 | 0.2229923 |
| ICAM1 | DEXA_Fat_Percentage | 0.38 | 0.0373292 | 0.3201517 |
| ICAM1 | DI | -0.40 | 0.0446263 | 0.3460245 |
| ICAM3 | auc_glucose | -0.53 | 0.0027307 | 0.0850297 |
| ICAM3 | Glucose_bsl | -0.43 | 0.0177098 | 0.2195471 |
| ICAM3 | HbA1C | -0.39 | 0.0321202 | 0.2997596 |
| ICAM3 | DEXA__Fat_Mass | -0.38 | 0.0381696 | 0.3236742 |
| ICAM3 | Sg | 0.40 | 0.0470744 | 0.3560947 |
| IFT74 | auc_glucose | -0.55 | 0.0017254 | 0.0690509 |
| IFT74 | Glucose_bsl | -0.47 | 0.0088335 | 0.156623 |
| IGFBP4 | HbA1C | -0.47 | 0.0081052 | 0.1480569 |
| IGFBP4 | Waist_Circumference | -0.47 | 0.0094146 | 0.159257 |
| IGFBP4 | auc_glucose | -0.37 | 0.0434464 | 0.343357 |
| IGFBP4 | DEXA_Weight | -0.37 | 0.0469365 | 0.3556091 |
| IGFBP5 | DEXA__Fat_Mass | -0.45 | 0.0118289 | 0.176847 |
| IGFBP5 | Waist_Circumference | -0.41 | 0.0237266 | 0.2577542 |
| IGFBP5 | HDL | 0.39 | 0.0330047 | 0.3022503 |
| IGFBP5 | DEXA_Fat_Percentage | -0.37 | 0.0448622 | 0.3468037 |
| IGFBP5 | QUICKI | 0.37 | 0.0472087 | 0.3560947 |
| IGFBP5 | Insulin_bsl | -0.36 | 0.0485594 | 0.3622897 |
| IL6ST | DEXA__Fat_Mass | -0.47 | 0.0083684 | 0.1506829 |
| IL6ST | auc_INSULIN | -0.46 | 0.0101094 | 0.1628255 |
| IL6ST | HOMA_B | -0.44 | 0.0148533 | 0.199326 |
| IL6ST | Insulin_bsl | -0.37 | 0.0416541 | 0.3361797 |
| IL6ST | Diastolic_Blood_Pressure_Average | 0.37 | 0.0462719 | 0.3521461 |
| INF2 | auc_glucose | -0.69 | 2.39E-05 | 0.0223041 |
| INF2 | Glucose_bsl | -0.66 | 6.33E-05 | 0.0266984 |
| INF2 | HbA1C | -0.56 | 0.0013763 | 0.0649714 |
| INF2 | DEXA_Weight | -0.52 | 0.0031882 | 0.093781 |
| INF2 | DEXA__Fat_Mass | -0.52 | 0.0034765 | 0.0984157 |
| INF2 | Waist_Circumference | -0.48 | 0.0068636 | 0.1347265 |
| INF2 | HOMA_IR | -0.38 | 0.0409307 | 0.3340179 |
| IPO5 | Waist_Circumference | 0.42 | 0.0212342 | 0.243021 |
| IPO5 | Heart_Rate_Average | 0.37 | 0.0432589 | 0.3429389 |
| IPO5 | HDL | -0.36 | 0.0486445 | 0.3623594 |
| IRX3 | auc_glucose | -0.59 | 0.0006826 | 0.0525108 |
| IRX3 | Heart_Rate_Average | -0.44 | 0.0160215 | 0.2085655 |
| IRX3 | Glucose_bsl | -0.44 | 0.0162019 | 0.2101496 |
| IRX3 | HOMA_IR | -0.38 | 0.0402724 | 0.3315451 |
| IST1 | auc_glucose | 0.41 | 0.0247485 | 0.2647046 |
| IST1 | Glucose_bsl | 0.38 | 0.0384849 | 0.3236742 |
| ITGA6 | auc_glucose | 0.63 | 0.0001829 | 0.0355725 |
| ITGA6 | Glucose_bsl | 0.55 | 0.0017426 | 0.0690509 |
| ITGA6 | HbA1C | 0.40 | 0.0302221 | 0.290967 |
| ITGAL | auc_glucose | -0.62 | 0.0002442 | 0.0374102 |
| ITGAL | Glucose_bsl | -0.54 | 0.0018653 | 0.0710068 |
| ITGAL | HbA1C | -0.39 | 0.0333623 | 0.3033635 |
| ITPR3 | HOMA_B | -0.46 | 0.0100244 | 0.1628255 |
| ITPR3 | auc_INSULIN | -0.38 | 0.0384172 | 0.3236742 |
| JUP | auc_glucose | -0.50 | 0.0044733 | 0.1104921 |
| JUP | Glucose_bsl | -0.47 | 0.0091246 | 0.1586068 |
| JUP | HbA1C | -0.43 | 0.0188431 | 0.2278485 |
| KBTBD2 | auc_glucose | -0.50 | 0.0048369 | 0.1146784 |
| KBTBD2 | Glucose_bsl | -0.48 | 0.0070261 | 0.1374692 |
| KEL | auc_glucose | -0.56 | 0.0012608 | 0.0649714 |
| KEL | Glucose_bsl | -0.54 | 0.0020234 | 0.0749882 |
| KEL | DEXA__Fat_Mass | -0.38 | 0.0411446 | 0.3345073 |
| KEL | Heart_Rate_Average | -0.36 | 0.0486958 | 0.3623594 |
| KIAA0513 | auc_glucose | 0.46 | 0.0101098 | 0.1628255 |
| KIAA0513 | Glucose_bsl | 0.45 | 0.0117684 | 0.176847 |
| KIAA0513 | DEXA_Fat_Percentage | 0.41 | 0.0240612 | 0.2591942 |
| KLHDC3 | auc_glucose | -0.60 | 0.0004987 | 0.0483802 |
| KLHDC3 | Glucose_bsl | -0.54 | 0.0018555 | 0.0710068 |
| KLHDC3 | LDL | 0.39 | 0.0390614 | 0.3270837 |
| KLHDC3 | HbA1C | -0.37 | 0.043585 | 0.3439229 |
| KMT2A | auc_glucose | -0.56 | 0.0011758 | 0.0649714 |
| KMT2A | Glucose_bsl | -0.55 | 0.0017573 | 0.0690509 |
| KMT2A | HbA1C | -0.45 | 0.0129432 | 0.1832757 |
| KPRP | DEXA__Fat_Mass | -0.43 | 0.0180746 | 0.2229923 |
| KRT31 | Glucose_bsl | 0.57 | 0.0011002 | 0.0649714 |
| KRT31 | auc_glucose | 0.55 | 0.0015599 | 0.0670046 |
| KRT31 | HbA1C | 0.48 | 0.0068158 | 0.1346977 |
| KRT31 | DEXA__Fat_Mass | 0.45 | 0.0117546 | 0.176847 |
| KRT31 | MATSUDA | -0.39 | 0.0334933 | 0.3042847 |
| KRT31 | HDL | -0.39 | 0.0340043 | 0.3049782 |
| KRT31 | DEXA_Weight | 0.38 | 0.0400332 | 0.330902 |
| KRT31 | Waist_Circumference | 0.37 | 0.0427443 | 0.3404503 |
| KRT74 | auc_glucose | -0.62 | 0.000242 | 0.0374102 |
| KRT74 | Glucose_bsl | -0.56 | 0.0011807 | 0.0649714 |
| KRT74 | HbA1C | -0.46 | 0.0104617 | 0.1662058 |
| KRT86 | Glucose_bsl | 0.41 | 0.0258128 | 0.268485 |
| KRT86 | auc_glucose | 0.39 | 0.0319515 | 0.2984663 |
| KRT86 | DEXA__Fat_Mass | 0.37 | 0.0418211 | 0.3367575 |
| KTN1 | DEXA__Fat_Mass | 0.44 | 0.0145668 | 0.197808 |
| KTN1 | Heart_Rate_Average | 0.39 | 0.0341169 | 0.3053032 |
| KTN1 | QUICKI | -0.38 | 0.0390592 | 0.3270837 |
| LAMTOR1 | auc_glucose | 0.51 | 0.0040345 | 0.1043271 |
| LAMTOR1 | Glucose_bsl | 0.50 | 0.0050155 | 0.1157374 |
| LAMTOR1 | DEXA__Fat_Mass | 0.41 | 0.025853 | 0.2686307 |
| LAMTOR5 | auc_glucose | -0.52 | 0.0033291 | 0.0951987 |
| LAMTOR5 | Sg | 0.47 | 0.018564 | 0.2255363 |
| LAMTOR5 | AIRg | 0.44 | 0.0260818 | 0.269915 |
| LAMTOR5 | Glucose_bsl | -0.40 | 0.0276971 | 0.2787634 |
| LAMTOR5 | DI | 0.40 | 0.0459333 | 0.3514013 |
| LCP2 | auc_glucose | 0.53 | 0.0025597 | 0.0826354 |
| LCP2 | HbA1C | 0.48 | 0.0070301 | 0.1374692 |
| LCP2 | Glucose_bsl | 0.45 | 0.0127874 | 0.1820739 |
| LCP2 | LDL | -0.39 | 0.0377232 | 0.3216496 |
| LGALS1 | auc_glucose | 0.61 | 0.0003045 | 0.0403865 |
| LGALS1 | Glucose_bsl | 0.56 | 0.0011958 | 0.0649714 |
| LGALS1 | DEXA__Fat_Mass | 0.53 | 0.0023691 | 0.0800027 |
| LGALS1 | HbA1C | 0.47 | 0.0092267 | 0.159257 |
| LGALSL | auc_glucose | 0.68 | 4.06E-05 | 0.0244807 |
| LGALSL | Glucose_bsl | 0.60 | 0.000412 | 0.0469948 |
| LGALSL | HbA1C | 0.50 | 0.00457 | 0.1115441 |
| LMAN1 | Sg | 0.51 | 0.009835 | 0.1620637 |
| LMAN1 | AIRg | 0.49 | 0.0120997 | 0.1784699 |
| LMAN1 | DEXA_Weight | 0.45 | 0.0126832 | 0.1810933 |
| LMAN1 | Waist_Circumference | 0.39 | 0.0325967 | 0.3012043 |
| LMAN1 | DEXA__Fat_Mass | 0.38 | 0.0375359 | 0.3208524 |
| LMAN1 | MATSUDA | -0.37 | 0.0421094 | 0.3382594 |
| LMAN1 | DI | 0.40 | 0.0495062 | 0.3656336 |
| LMAN2 | Respiration_Rate | 0.42 | 0.0223161 | 0.2498328 |
| LPCAT2 | auc_glucose | 0.59 | 0.0006408 | 0.0509925 |
| LPCAT2 | HbA1C | 0.54 | 0.0021877 | 0.0780827 |
| LPCAT2 | Glucose_bsl | 0.53 | 0.0024449 | 0.0813784 |
| LPCAT2 | Waist_Circumference | 0.50 | 0.0045741 | 0.1115441 |
| LPCAT2 | DEXA__Fat_Mass | 0.49 | 0.0059247 | 0.1248932 |
| LPCAT2 | DEXA_Weight | 0.46 | 0.011062 | 0.1712859 |
| LPCAT2 | HOMA_IR | 0.39 | 0.0327133 | 0.3020079 |
| LPCAT2 | HDL | -0.37 | 0.0414935 | 0.33594 |
| LPCAT2 | Insulin_bsl | 0.36 | 0.0487845 | 0.3623594 |
| LRCH1 | auc_glucose | -0.62 | 0.0002784 | 0.0403865 |
| LRCH1 | Glucose_bsl | -0.54 | 0.0022413 | 0.0782674 |
| LRCH1 | HbA1C | -0.37 | 0.0433848 | 0.3431346 |
| LRRC59 | Glucose_bsl | 0.63 | 0.0002043 | 0.0355725 |
| LRRC59 | auc_glucose | 0.60 | 0.0004324 | 0.0477167 |
| LRRC59 | Waist_Circumference | 0.44 | 0.0147815 | 0.1991444 |
| LRRC59 | HbA1C | 0.41 | 0.0232183 | 0.2543856 |
| LRRC59 | Diastolic_Blood_Pressure_Average | 0.40 | 0.0269628 | 0.2743311 |
| LRRC59 | Respiration_Rate | 0.38 | 0.0442867 | 0.345115 |
| LTA4H | auc_glucose | -0.62 | 0.0002969 | 0.0403865 |
| LTA4H | Glucose_bsl | -0.56 | 0.0013504 | 0.0649714 |
| LTA4H | DEXA__Fat_Mass | -0.37 | 0.04225 | 0.3384078 |
| LTA4H | HbA1C | -0.36 | 0.0495744 | 0.3658739 |
| LZIC | auc_glucose | -0.56 | 0.0012144 | 0.0649714 |
| LZIC | Glucose_bsl | -0.50 | 0.0049556 | 0.1157374 |
| LZIC | HbA1C | -0.43 | 0.0165918 | 0.2124584 |
| LZIC | HOMA_IR | -0.37 | 0.0430066 | 0.3419875 |
| MANF | TSH | 0.43 | 0.0177716 | 0.2195471 |
| MANF | auc_glucose | 0.38 | 0.0398077 | 0.3296245 |
| MAPK13 | auc_glucose | -0.54 | 0.0021217 | 0.0767053 |
| MAPK13 | Glucose_bsl | -0.50 | 0.0050588 | 0.1157374 |
| MAPK13 | DEXA_Fat_Percentage | -0.46 | 0.0099956 | 0.1628255 |
| MAPK13 | DEXA__Lean_Mass | 0.38 | 0.0383755 | 0.3236742 |
| MAPRE1 | auc_glucose | 0.63 | 0.0001765 | 0.0355725 |
| MAPRE1 | Glucose_bsl | 0.56 | 0.0012707 | 0.0649714 |
| MAPRE1 | HbA1C | 0.52 | 0.003094 | 0.0934195 |
| MARS1 | auc_glucose | -0.61 | 0.0003378 | 0.0422842 |
| MARS1 | Glucose_bsl | -0.54 | 0.0021905 | 0.0780827 |
| MARS1 | Sg | 0.55 | 0.0046113 | 0.112172 |
| MARS1 | DI | 0.55 | 0.0047262 | 0.1133636 |
| MARS1 | HbA1C | -0.46 | 0.0107195 | 0.1685238 |
| MCM2 | auc_glucose | -0.56 | 0.0012261 | 0.0649714 |
| MCM2 | Glucose_bsl | -0.54 | 0.0021291 | 0.0767053 |
| MCM2 | DEXA__Fat_Mass | -0.39 | 0.0310619 | 0.2939004 |
| MCM2 | DEXA_Fat_Percentage | -0.37 | 0.0462736 | 0.3521461 |
| MCMBP | auc_glucose | 0.51 | 0.0037401 | 0.1015759 |
| MCMBP | Glucose_bsl | 0.50 | 0.0051673 | 0.1157374 |
| MCMBP | AIRg | -0.53 | 0.0062939 | 0.1281999 |
| MCMBP | DI | -0.53 | 0.0068286 | 0.1346977 |
| MCMBP | HbA1C | 0.43 | 0.0169356 | 0.214439 |
| MDH1 | auc_glucose | 0.43 | 0.0166447 | 0.2124584 |
| MDH1 | DEXA_Fat_Percentage | 0.41 | 0.0246684 | 0.2643483 |
| MDK | auc_glucose | -0.60 | 0.0004769 | 0.0483802 |
| MDK | Glucose_bsl | -0.55 | 0.0014886 | 0.0658243 |
| MDK | HbA1C | -0.55 | 0.0016977 | 0.0688865 |
| MDK | Waist_Circumference | -0.37 | 0.0447738 | 0.3466428 |
| ME2 | HOMA_IR | -0.51 | 0.0039822 | 0.1042278 |
| ME2 | Waist_Circumference | -0.50 | 0.0051723 | 0.1157374 |
| ME2 | Glucose_bsl | -0.47 | 0.0093939 | 0.159257 |
| ME2 | Insulin_bsl | -0.44 | 0.0162752 | 0.2101496 |
| ME2 | HDL | 0.40 | 0.0269397 | 0.2743311 |
| ME2 | HbA1C | -0.40 | 0.0290539 | 0.2868927 |
| MGAT1 | QUICKI | -0.48 | 0.006849 | 0.1346977 |
| MGAT1 | Glucose_bsl | 0.45 | 0.011869 | 0.176847 |
| MGAT1 | DEXA__Fat_Mass | 0.44 | 0.0141873 | 0.1947855 |
| MGAT1 | MATSUDA | -0.44 | 0.0150988 | 0.2005235 |
| MGAT1 | Heart_Rate_Average | 0.42 | 0.0224666 | 0.2505118 |
| MGAT1 | Waist_Circumference | 0.37 | 0.0460922 | 0.3515474 |
| MGP | Ins30_glu30 | 0.42 | 0.0199816 | 0.2365396 |
| MGP | QUICKI | 0.40 | 0.0291145 | 0.2871173 |
| MNDA | MATSUDA | 0.56 | 0.0012569 | 0.0649714 |
| MNDA | QUICKI | 0.53 | 0.0027945 | 0.0866466 |
| MPIG6B | auc_glucose | -0.49 | 0.0055657 | 0.1201645 |
| MPIG6B | Glucose_bsl | -0.47 | 0.0091929 | 0.1591473 |
| MPIG6B | Heart_Rate_Average | -0.37 | 0.0438653 | 0.345115 |
| MS4A1 | TSH | -0.50 | 0.0050572 | 0.1157374 |
| MS4A1 | QUICKI | -0.47 | 0.0089545 | 0.1579496 |
| MS4A1 | auc_glucose | -0.42 | 0.021805 | 0.2465322 |
| MTHFD1 | Waist_Circumference | 0.50 | 0.004471 | 0.1104921 |
| MTHFD1 | DEXA_Weight | 0.47 | 0.0080442 | 0.1474676 |
| MTHFD1 | HDL | -0.39 | 0.0307887 | 0.2931794 |
| MTHFD1 | auc_INSULIN | 0.38 | 0.0398648 | 0.3297757 |
| MTHFD1 | DEXA__Fat_Mass | 0.37 | 0.0445963 | 0.3460245 |
| MUC4 | auc_glucose | -0.57 | 0.0009965 | 0.0637593 |
| MUC4 | Glucose_bsl | -0.53 | 0.0023564 | 0.0800027 |
| MUC4 | HbA1C | -0.37 | 0.0465351 | 0.3538739 |
| MYO5B | auc_glucose | -0.51 | 0.0043106 | 0.1083608 |
| MYO5B | Glucose_bsl | -0.49 | 0.0056631 | 0.1208669 |
| MYO5B | HbA1C | -0.47 | 0.0091462 | 0.1586068 |
| MYO5B | Waist_Circumference | -0.41 | 0.0255311 | 0.2678803 |
| MYO5B | DI | 0.41 | 0.039756 | 0.3296245 |
| MYO5B | DEXA__Fat_Mass | -0.38 | 0.0398093 | 0.3296245 |
| MYO9B | auc_glucose | 0.58 | 0.0007323 | 0.0540857 |
| MYO9B | Glucose_bsl | 0.53 | 0.0024509 | 0.0813784 |
| MYO9B | HbA1C | 0.49 | 0.0054902 | 0.1194112 |
| MYO9B | Heart_Rate_Average | 0.46 | 0.0110044 | 0.1711638 |
| NAPRT | auc_glucose | 0.46 | 0.0101579 | 0.1629389 |
| NAPRT | Temperature | -0.45 | 0.0207316 | 0.2406903 |
| NCAM2 | HbA1C | 0.62 | 0.0002894 | 0.0403865 |
| NCAM2 | auc_glucose | 0.60 | 0.0004342 | 0.0477167 |
| NCAM2 | Glucose_bsl | 0.51 | 0.0043722 | 0.1092101 |
| NCAM2 | DI | -0.50 | 0.0108294 | 0.1694737 |
| NCAM2 | AIRg | -0.48 | 0.0147611 | 0.1991444 |
| NCEH1 | auc_glucose | -0.64 | 0.0001496 | 0.0355725 |
| NCEH1 | Glucose_bsl | -0.57 | 0.0010013 | 0.0637593 |
| NCEH1 | HbA1C | -0.42 | 0.0193236 | 0.2322403 |
| NES | Heart_Rate_Average | -0.42 | 0.0205677 | 0.2403181 |
| NHLRC2 | auc_glucose | -0.51 | 0.0038355 | 0.1015905 |
| NHLRC2 | HbA1C | -0.45 | 0.0121804 | 0.1788499 |
| NHLRC2 | Glucose_bsl | -0.43 | 0.0167011 | 0.2124584 |
| NINL | auc_glucose | 0.57 | 0.0008994 | 0.0595715 |
| NINL | Glucose_bsl | 0.44 | 0.0144599 | 0.197139 |
| NINL | HbA1C | 0.37 | 0.0433619 | 0.3431346 |
| NNT | auc_glucose | 0.48 | 0.0073666 | 0.1405228 |
| NNT | TSH | 0.42 | 0.0217597 | 0.2465322 |
| NRGN | DEXA__Fat_Mass | 0.56 | 0.0011689 | 0.0649714 |
| NRGN | auc_INSULIN | 0.41 | 0.0262849 | 0.2717434 |
| NRGN | Insulin_bsl | 0.40 | 0.030037 | 0.290967 |
| NT5E | auc_glucose | -0.75 | 1.54E-06 | 0.0067654 |
| NT5E | Glucose_bsl | -0.62 | 0.0002431 | 0.0374102 |
| NT5E | HbA1C | -0.61 | 0.0003291 | 0.041706 |
| NT5E | DI | 0.50 | 0.0112759 | 0.1725156 |
| NT5E | HOMA_IR | -0.38 | 0.0360563 | 0.3151561 |
| OSTF1 | auc_glucose | 0.47 | 0.0091331 | 0.1586068 |
| OSTF1 | Glucose_bsl | 0.46 | 0.0114178 | 0.1739098 |
| OSTF1 | HbA1C | 0.38 | 0.0391651 | 0.3274178 |
| OTOF | HDL | -0.56 | 0.0013797 | 0.0649714 |
| OTOF | Waist_Circumference | 0.55 | 0.001494 | 0.0658243 |
| OTOF | MATSUDA | -0.51 | 0.0035992 | 0.1001333 |
| OTOF | DEXA_Weight | 0.47 | 0.009494 | 0.159257 |
| OTOF | QUICKI | -0.45 | 0.0124196 | 0.1807232 |
| OTOF | Insulin_bsl | 0.41 | 0.0237737 | 0.2579923 |
| OTOF | DEXA__Lean_Mass | 0.36 | 0.0478589 | 0.3594142 |
| PA2G4 | Waist_Circumference | -0.46 | 0.0111856 | 0.1722962 |
| PA2G4 | Triglycerides | -0.43 | 0.0183226 | 0.2244628 |
| PA2G4 | MATSUDA | 0.43 | 0.0191051 | 0.2304731 |
| PA2G4 | HDL | 0.38 | 0.040421 | 0.3317364 |
| PA2G4 | QUICKI | 0.37 | 0.0460023 | 0.3515474 |
| PA2G4 | Temperature | 0.39 | 0.0471969 | 0.3560947 |
| PACSIN2 | DEXA__Fat_Mass | -0.50 | 0.0048691 | 0.1149114 |
| PACSIN2 | auc_glucose | -0.49 | 0.0060867 | 0.1264891 |
| PACSIN2 | DEXA_Fat_Percentage | -0.49 | 0.0063864 | 0.1295699 |
| PACSIN2 | Glucose_bsl | -0.44 | 0.0143168 | 0.1957074 |
| PANX1 | Waist_Circumference | 0.48 | 0.0076973 | 0.1428945 |
| PANX1 | Insulin_bsl | 0.47 | 0.009483 | 0.159257 |
| PANX1 | HOMA_IR | 0.43 | 0.0182006 | 0.2237695 |
| PAPOLA | auc_glucose | -0.62 | 0.0002654 | 0.0394906 |
| PAPOLA | Glucose_bsl | -0.55 | 0.0015347 | 0.0664575 |
| PAPOLA | HbA1C | -0.38 | 0.0356836 | 0.3128336 |
| PARK7 | auc_glucose | 0.55 | 0.0015386 | 0.0664575 |
| PARK7 | Glucose_bsl | 0.52 | 0.0031318 | 0.0937729 |
| PARK7 | DEXA__Fat_Mass | 0.50 | 0.0051803 | 0.1157374 |
| PARK7 | Waist_Circumference | 0.44 | 0.0154197 | 0.2037306 |
| PARK7 | HbA1C | 0.42 | 0.0224972 | 0.2505118 |
| PCBP2 | auc_glucose | 0.53 | 0.0025411 | 0.0826354 |
| PCBP2 | Glucose_bsl | 0.45 | 0.0126586 | 0.1809996 |
| PCBP2 | HbA1C | 0.42 | 0.0207961 | 0.2406903 |
| PCBP2 | auc_c_peptide | -0.51 | 0.0368094 | 0.3178999 |
| PDCD10 | auc_glucose | 0.53 | 0.0023638 | 0.0800027 |
| PDCD10 | Glucose_bsl | 0.52 | 0.0035503 | 0.0993105 |
| PDCD10 | HbA1C | 0.51 | 0.0037324 | 0.1015759 |
| PDCD10 | Temperature | -0.40 | 0.041003 | 0.334342 |
| PDE5A | auc_glucose | -0.57 | 0.0010337 | 0.0639281 |
| PDE5A | Glucose_bsl | -0.50 | 0.0050231 | 0.1157374 |
| PDE5A | HbA1C | -0.44 | 0.0147972 | 0.1991444 |
| PDGFA | auc_glucose | 0.61 | 0.0003642 | 0.0425919 |
| PDGFA | Glucose_bsl | 0.48 | 0.0077569 | 0.1434825 |
| PDGFA | AIRg | -0.46 | 0.0194551 | 0.2328065 |
| PDIA6 | auc_glucose | -0.65 | 0.0001031 | 0.0302333 |
| PDIA6 | Glucose_bsl | -0.60 | 0.0004462 | 0.0477167 |
| PDIA6 | HbA1C | -0.57 | 0.0010317 | 0.0639281 |
| PDIA6 | DI | 0.55 | 0.0047997 | 0.1144431 |
| PDIA6 | DEXA__Fat_Mass | -0.43 | 0.0165293 | 0.212112 |
| PDIA6 | HOMA_IR | -0.37 | 0.0462259 | 0.3521461 |
| PDLIM5 | auc_glucose | -0.71 | 1.16E-05 | 0.0170346 |
| PDLIM5 | Glucose_bsl | -0.68 | 4.22E-05 | 0.0244807 |
| PDLIM5 | HbA1C | -0.54 | 0.0022684 | 0.0784083 |
| PDLIM5 | DEXA_Fat_Percentage | -0.42 | 0.0225649 | 0.2505118 |
| PDLIM5 | Sg | 0.43 | 0.0330338 | 0.3022503 |
| PDLIM5 | DI | 0.41 | 0.0421816 | 0.3383097 |
| PER3 | auc_glucose | -0.56 | 0.0012291 | 0.0649714 |
| PER3 | Glucose_bsl | -0.54 | 0.0020083 | 0.0749631 |
| PER3 | DEXA__Fat_Mass | -0.39 | 0.0312284 | 0.294793 |
| PER3 | DEXA_Fat_Percentage | -0.37 | 0.0467596 | 0.3550545 |
| PGM1 | auc_glucose | 0.52 | 0.0035036 | 0.0985427 |
| PGM1 | Glucose_bsl | 0.51 | 0.0037641 | 0.1015905 |
| PGRMC2 | auc_glucose | 0.58 | 0.0007703 | 0.0560825 |
| PGRMC2 | Glucose_bsl | 0.41 | 0.0259373 | 0.2692342 |
| PGRMC2 | HbA1C | 0.40 | 0.0284553 | 0.2822433 |
| PHB | DEXA__Fat_Mass | -0.60 | 0.0004928 | 0.0483802 |
| PHB | DEXA_Fat_Percentage | -0.46 | 0.0109239 | 0.1702964 |
| PHB | Systolic_Blood_Pressure_Average | 0.40 | 0.0269479 | 0.2743311 |
| PHB | Glucose_bsl | -0.40 | 0.0285033 | 0.2824464 |
| PHB | MATSUDA | 0.37 | 0.041355 | 0.335348 |
| PHKB | DEXA__Fat_Mass | 0.47 | 0.0080327 | 0.1474676 |
| PHOSPHO2 | auc_glucose | -0.67 | 5.80E-05 | 0.0266984 |
| PHOSPHO2 | Glucose_bsl | -0.58 | 0.0008527 | 0.0587056 |
| PHOSPHO2 | HbA1C | -0.52 | 0.00302 | 0.0922732 |
| PHOSPHO2 | HOMA_IR | -0.38 | 0.0398144 | 0.3296245 |
| PHOSPHO2 | LDL | 0.37 | 0.0480483 | 0.3598213 |
| PI4KA | auc_glucose | 0.59 | 0.0006093 | 0.0500213 |
| PI4KA | Glucose_bsl | 0.50 | 0.0051219 | 0.1157374 |
| PI4KA | HbA1C | 0.46 | 0.0104854 | 0.1662058 |
| PKHD1L1 | auc_glucose | -0.52 | 0.0033002 | 0.0951158 |
| PKHD1L1 | HbA1C | -0.48 | 0.0067946 | 0.1346582 |
| PKHD1L1 | DEXA__Fat_Mass | -0.43 | 0.0184947 | 0.2254945 |
| PKHD1L1 | Glucose_bsl | -0.42 | 0.0206447 | 0.2406903 |
| PKHD1L1 | DEXA_Weight | -0.42 | 0.0212265 | 0.243021 |
| PKHD1L1 | Waist_Circumference | -0.41 | 0.0255945 | 0.2678803 |
| PKHD1L1 | DI | 0.44 | 0.0263921 | 0.2725771 |
| PKHD1L1 | HOMA_IR | -0.40 | 0.0277902 | 0.2791771 |
| PKHD1L1 | Insulin_bsl | -0.39 | 0.0309304 | 0.2937383 |
| PLCH1 | auc_glucose | -0.67 | 4.71E-05 | 0.0254715 |
| PLCH1 | Glucose_bsl | -0.63 | 0.0001669 | 0.0355725 |
| PLCH1 | HbA1C | -0.47 | 0.0091312 | 0.1586068 |
| PLCH1 | DEXA__Fat_Mass | -0.37 | 0.041588 | 0.3361753 |
| PLEKHF2 | auc_glucose | 0.53 | 0.0026464 | 0.0836356 |
| PLEKHF2 | Ins30_glu30 | -0.50 | 0.0048047 | 0.1144431 |
| PLEKHF2 | Glucose_bsl | 0.48 | 0.0077476 | 0.1434825 |
| PLEKHF2 | Si | 0.48 | 0.014949 | 0.1996867 |
| PLEKHF2 | AIRg | -0.47 | 0.0185631 | 0.2255363 |
| PLEKHF2 | HbA1C | 0.39 | 0.0313829 | 0.2950241 |
| PLEKHG4 | auc_glucose | 0.58 | 0.0008608 | 0.0587056 |
| PLEKHG4 | Glucose_bsl | 0.50 | 0.0050529 | 0.1157374 |
| PLEKHO1 | auc_glucose | -0.54 | 0.0018675 | 0.0710068 |
| PLEKHO1 | Glucose_bsl | -0.47 | 0.0088812 | 0.1571973 |
| PLEKHO1 | MATSUDA | 0.39 | 0.0343219 | 0.306124 |
| PLEKHO1 | DEXA__Fat_Mass | -0.38 | 0.0366946 | 0.3178999 |
| PLPBP | auc_glucose | -0.50 | 0.005236 | 0.1157374 |
| PLPBP | Glucose_bsl | -0.42 | 0.0223012 | 0.2498328 |
| PLPBP | HOMA_IR | -0.39 | 0.033713 | 0.3047945 |
| PLPBP | DEXA__Fat_Mass | -0.38 | 0.0388362 | 0.3259952 |
| PLS3 | auc_glucose | -0.56 | 0.0013529 | 0.0649714 |
| PLXNA4 | auc_glucose | 0.59 | 0.000563 | 0.0500213 |
| PLXNA4 | Glucose_bsl | 0.51 | 0.0040921 | 0.1050226 |
| PLXNA4 | HbA1C | 0.46 | 0.009698 | 0.161361 |
| PLXNA4 | Waist_Circumference | 0.42 | 0.0204828 | 0.2402263 |
| PLXNA4 | HDL | -0.38 | 0.0375332 | 0.3208524 |
| PLXNA4 | HOMA_IR | 0.36 | 0.0487126 | 0.3623594 |
| PLXNA4 | TSH | 0.36 | 0.0488511 | 0.3623594 |
| POGLUT1 | auc_glucose | -0.59 | 0.0005507 | 0.0500213 |
| POGLUT1 | Glucose_bsl | -0.49 | 0.0056858 | 0.1211011 |
| POLR1A | HOMA_B | 0.49 | 0.0061803 | 0.1271133 |
| POLR1A | AIRg | 0.50 | 0.0117119 | 0.176847 |
| POLR1A | auc_INSULIN | 0.45 | 0.0118657 | 0.176847 |
| POLR1A | Sg | 0.47 | 0.0174167 | 0.2174972 |
| POTEH | auc_glucose | -0.59 | 0.0005798 | 0.0500213 |
| POTEH | Glucose_bsl | -0.56 | 0.0012917 | 0.0649714 |
| POTEH | HbA1C | -0.39 | 0.0330096 | 0.3022503 |
| POTEH | DEXA__Fat_Mass | -0.37 | 0.0417018 | 0.3363004 |
| POTEI | Insulin_bsl | 0.65 | 9.53E-05 | 0.0288756 |
| POTEI | HOMA_IR | 0.61 | 0.0003027 | 0.0403865 |
| POTEI | LDL | -0.51 | 0.0044297 | 0.1103767 |
| POTEI | QUICKI | -0.47 | 0.0091315 | 0.1586068 |
| POTEI | DEXA_Weight | 0.45 | 0.0119093 | 0.1771894 |
| POTEI | HDL | -0.45 | 0.0124104 | 0.1807232 |
| POTEI | Waist_Circumference | 0.44 | 0.0142745 | 0.19539 |
| POTEI | auc_INSULIN | 0.44 | 0.0162243 | 0.2101496 |
| POTEI | HOMA_B | 0.42 | 0.019234 | 0.231485 |
| POTEI | HbA1C | 0.38 | 0.0404734 | 0.3318692 |
| PPP1CA | Glucose_bsl | 0.50 | 0.0049993 | 0.1157374 |
| PPP1CA | auc_glucose | 0.50 | 0.0050123 | 0.1157374 |
| PPP1CA | HbA1C | 0.39 | 0.0354829 | 0.3117143 |
| PPP1CA | DEXA__Fat_Mass | 0.37 | 0.0456513 | 0.3502661 |
| PPP1CA | auc_c_peptide | -0.48 | 0.0499058 | 0.3666092 |
| PPP1CB | auc_glucose | 0.51 | 0.0041376 | 0.105664 |
| PPP1CB | Glucose_bsl | 0.42 | 0.0201845 | 0.2381769 |
| PPP1CB | TSH | 0.41 | 0.0250242 | 0.2651852 |
| PPP1CB | HbA1C | 0.41 | 0.0257547 | 0.2681521 |
| PPP1CB | Sg | -0.41 | 0.0411862 | 0.3345073 |
| PPP2CA | auc_glucose | 0.55 | 0.0015912 | 0.0677815 |
| PPP2CA | Glucose_bsl | 0.47 | 0.0086584 | 0.1540499 |
| PPP2CA | HbA1C | 0.39 | 0.0308144 | 0.2931794 |
| PPP6C | auc_glucose | -0.47 | 0.0090652 | 0.1585408 |
| PPP6C | DEXA__Fat_Mass | -0.46 | 0.010078 | 0.1628255 |
| PPP6C | DEXA_Fat_Percentage | -0.42 | 0.0209681 | 0.2415061 |
| PPP6C | Glucose_bsl | -0.41 | 0.0228448 | 0.2526758 |
| PPP6C | Heart_Rate_Average | -0.39 | 0.0339843 | 0.3049782 |
| PRG3 | AIRg | -0.50 | 0.0108105 | 0.1694358 |
| PRG3 | auc_glucose | 0.39 | 0.0354498 | 0.3117143 |
| PRKAR1A | auc_glucose | -0.56 | 0.0012949 | 0.0649714 |
| PRKAR1A | Glucose_bsl | -0.56 | 0.0013756 | 0.0649714 |
| PRKAR1A | HOMA_IR | -0.47 | 0.009028 | 0.1581892 |
| PRKAR1A | DEXA__Fat_Mass | -0.47 | 0.0094787 | 0.159257 |
| PRKAR1A | QUICKI | 0.46 | 0.0099909 | 0.1628255 |
| PRKAR1A | Waist_Circumference | -0.46 | 0.0112596 | 0.1725156 |
| PRKAR1A | Insulin_bsl | -0.44 | 0.0142603 | 0.19539 |
| PRKAR1A | HbA1C | -0.43 | 0.0164952 | 0.2119394 |
| PRKAR1A | Heart_Rate_Average | -0.42 | 0.0196185 | 0.2334652 |
| PRKAR1A | MATSUDA | 0.41 | 0.0228654 | 0.2526758 |
| PRKAR1A | HDL | 0.40 | 0.0295578 | 0.2889909 |
| PRKAR2A | auc_glucose | 0.57 | 0.0010468 | 0.0643517 |
| PRKAR2A | HbA1C | 0.49 | 0.0060054 | 0.1258187 |
| PRKAR2A | DI | -0.51 | 0.0094892 | 0.159257 |
| PRKAR2A | Glucose_bsl | 0.45 | 0.0122986 | 0.1798542 |
| PRKCA | Waist_Circumference | -0.46 | 0.0097221 | 0.1615008 |
| PRKCA | auc_glucose | -0.42 | 0.0196177 | 0.2334652 |
| PRKCA | TSH | -0.40 | 0.0299118 | 0.2902406 |
| PRKCA | HbA1C | -0.37 | 0.0451089 | 0.3481866 |
| PRKCA | Glucose_bsl | -0.37 | 0.0459362 | 0.3514013 |
| PRKDC | auc_glucose | -0.56 | 0.0013297 | 0.0649714 |
| PRKDC | Glucose_bsl | -0.52 | 0.0032248 | 0.0943171 |
| PRKDC | Heart_Rate_Average | -0.42 | 0.0194572 | 0.2328065 |
| PRKDC | Triglycerides | 0.38 | 0.039243 | 0.3277218 |
| PRSS3 | Waist_Circumference | -0.49 | 0.0055787 | 0.1201645 |
| PRSS3 | DI | 0.42 | 0.0348088 | 0.3091237 |
| PRSS3 | HbA1C | -0.38 | 0.0394551 | 0.329038 |
| PSMB9 | auc_glucose | 0.63 | 0.0002172 | 0.0355725 |
| PSMB9 | HbA1C | 0.56 | 0.0014205 | 0.0650391 |
| PSMB9 | Glucose_bsl | 0.50 | 0.0053397 | 0.1166323 |
| PSMB9 | DEXA__Fat_Mass | 0.47 | 0.0082747 | 0.1495573 |
| PSMB9 | Si | -0.43 | 0.0323431 | 0.2999408 |
| PSMC4 | auc_glucose | 0.40 | 0.0264841 | 0.2729776 |
| PSMD7 | Waist_Circumference | 0.68 | 4.13E-05 | 0.0244807 |
| PSMD7 | DEXA_Weight | 0.60 | 0.0004221 | 0.0476161 |
| PSMD7 | HDL | -0.55 | 0.0016761 | 0.0688865 |
| PSMD7 | DEXA__Lean_Mass | 0.44 | 0.0150276 | 0.2000948 |
| PSMD7 | HbA1C | 0.43 | 0.0166774 | 0.2124584 |
| PSMD7 | DEXA__Fat_Mass | 0.42 | 0.020795 | 0.2406903 |
| PSMD7 | MATSUDA | -0.41 | 0.022732 | 0.251744 |
| PSMD7 | Glucose_bsl | 0.40 | 0.0266118 | 0.2732061 |
| PSMD7 | auc_glucose | 0.40 | 0.0297274 | 0.2895456 |
| PSMD7 | QUICKI | -0.39 | 0.0349265 | 0.3099011 |
| PSMD7 | HOMA_IR | 0.38 | 0.0383096 | 0.3236742 |
| PSMF1 | auc_glucose | 0.51 | 0.0038396 | 0.1015905 |
| PSMF1 | Glucose_bsl | 0.48 | 0.0071406 | 0.1383124 |
| PSMF1 | DEXA_Fat_Percentage | 0.41 | 0.0230717 | 0.2536913 |
| PSMF1 | DEXA__Fat_Mass | 0.39 | 0.0331531 | 0.3024743 |
| PTGES3 | HOMA_IR | -0.39 | 0.0330193 | 0.3022503 |
| RAB3D | auc_glucose | -0.65 | 0.0001131 | 0.0311114 |
| RAB3D | Glucose_bsl | -0.61 | 0.0003651 | 0.0425919 |
| RAB3D | HbA1C | -0.46 | 0.0098321 | 0.1620637 |
| RAB3D | DEXA__Fat_Mass | -0.40 | 0.0296969 | 0.2895238 |
| RAB6B | auc_glucose | 0.66 | 7.41E-05 | 0.0266984 |
| RAB6B | Glucose_bsl | 0.57 | 0.0011417 | 0.0649714 |
| RAB6B | HbA1C | 0.46 | 0.0099223 | 0.1628255 |
| RAB6B | Heart_Rate_Average | 0.38 | 0.0368666 | 0.3178999 |
| RASGRP2 | Systolic_Blood_Pressure_Average | 0.50 | 0.0052179 | 0.1157374 |
| RASGRP2 | Ins30_glu30 | 0.48 | 0.0075566 | 0.1420816 |
| RASGRP2 | DEXA_Fat_Percentage | -0.40 | 0.0277926 | 0.2791771 |
| RASGRP2 | Diastolic_Blood_Pressure_Average | 0.38 | 0.0401379 | 0.3310603 |
| RBM24 | MATSUDA | -0.54 | 0.002245 | 0.0782674 |
| RBM24 | QUICKI | -0.52 | 0.0033251 | 0.0951987 |
| RBM24 | HDL | -0.46 | 0.0111944 | 0.1722962 |
| RBM24 | Respiration_Rate | 0.45 | 0.0145439 | 0.1977582 |
| RBM24 | Waist_Circumference | 0.39 | 0.0316639 | 0.2965888 |
| RDH11 | Respiration_Rate | 0.57 | 0.0013418 | 0.0649714 |
| RDH11 | auc_glucose | 0.53 | 0.0024845 | 0.0814892 |
| RDH11 | Glucose_bsl | 0.45 | 0.0115385 | 0.1749699 |
| RDH11 | HbA1C | 0.42 | 0.0207074 | 0.2406903 |
| RDH11 | LDL | -0.38 | 0.0408779 | 0.3340179 |
| RFTN1 | DEXA__Fat_Mass | -0.45 | 0.013303 | 0.1863152 |
| RFTN1 | auc_glucose | -0.38 | 0.0408447 | 0.3340179 |
| RFTN1 | Glucose_bsl | -0.37 | 0.0415427 | 0.3360732 |
| RGS6 | DEXA__Fat_Mass | 0.48 | 0.0066322 | 0.1329405 |
| RGS6 | Glucose_bsl | 0.41 | 0.0231099 | 0.2537121 |
| RHOF | DEXA__Fat_Mass | 0.36 | 0.0481571 | 0.3603361 |
| RNF103 | auc_glucose | -0.51 | 0.0042174 | 0.1069026 |
| RNF103 | Glucose_bsl | -0.48 | 0.0072172 | 0.1387495 |
| RNF123 | Temperature | -0.41 | 0.0354952 | 0.3117143 |
| RNF40 | auc_glucose | 0.56 | 0.0013624 | 0.0649714 |
| RNF40 | Glucose_bsl | 0.41 | 0.0240077 | 0.25889 |
| RNF40 | MATSUDA | -0.39 | 0.0325723 | 0.3012043 |
| RNF40 | HbA1C | 0.39 | 0.0327425 | 0.3020079 |
| RNF40 | Waist_Circumference | 0.38 | 0.0375134 | 0.3208524 |
| RNF40 | Si | -0.41 | 0.0426494 | 0.3402009 |
| RNF40 | TSH | 0.37 | 0.0456221 | 0.3502661 |
| RPS6KA3 | auc_glucose | -0.40 | 0.0306652 | 0.2924778 |
| RSU1 | auc_glucose | 0.39 | 0.0323113 | 0.2999408 |
| RYR2 | auc_glucose | 0.56 | 0.0011503 | 0.0649714 |
| RYR2 | HbA1C | 0.55 | 0.0016065 | 0.0678718 |
| RYR2 | Glucose_bsl | 0.51 | 0.0043387 | 0.1086368 |
| RYR2 | Waist_Circumference | 0.46 | 0.0100014 | 0.1628255 |
| RYR2 | HOMA_IR | 0.46 | 0.0110842 | 0.1713711 |
| RYR2 | DEXA_Weight | 0.42 | 0.0217831 | 0.2465322 |
| RYR2 | Insulin_bsl | 0.41 | 0.0238959 | 0.2582267 |
| RYR2 | DI | -0.43 | 0.0331556 | 0.3024743 |
| RYR2 | DEXA__Fat_Mass | 0.39 | 0.033737 | 0.3047945 |
| RYR2 | HDL | -0.36 | 0.0497007 | 0.3660166 |
| S100A10 | HDL | 0.55 | 0.0016431 | 0.0685711 |
| S100A10 | MATSUDA | 0.54 | 0.0020154 | 0.0749631 |
| S100A10 | AIRg | -0.56 | 0.0037008 | 0.1015759 |
| S100A10 | QUICKI | 0.47 | 0.0093534 | 0.159257 |
| S100A10 | auc_INSULIN | -0.46 | 0.0109189 | 0.1702964 |
| S100A10 | Si | 0.48 | 0.0149258 | 0.1996867 |
| S100A10 | HOMA_B | -0.41 | 0.0255929 | 0.2678803 |
| S100A10 | Ins30_glu30 | -0.38 | 0.0383261 | 0.3236742 |
| S100A10 | Respiration_Rate | -0.39 | 0.0384196 | 0.3236742 |
| S100A12 | auc_glucose | 0.47 | 0.0083811 | 0.1506829 |
| S100A12 | AIRg | -0.48 | 0.0160497 | 0.2085655 |
| S100A12 | HOMA_B | -0.43 | 0.0191888 | 0.2312115 |
| S100A12 | auc_INSULIN | -0.39 | 0.0313561 | 0.2950241 |
| S100A6 | auc_glucose | 0.46 | 0.0101007 | 0.1628255 |
| S100A6 | Glucose_bsl | 0.39 | 0.0325358 | 0.3011835 |
| SCYL2 | auc_glucose | 0.75 | 2.17E-06 | 0.0067654 |
| SCYL2 | DI | -0.68 | 0.0001861 | 0.0355725 |
| SCYL2 | Glucose_bsl | 0.59 | 0.0006139 | 0.0500213 |
| SCYL2 | HbA1C | 0.56 | 0.0012206 | 0.0649714 |
| SCYL2 | Sg | -0.57 | 0.0028021 | 0.0866466 |
| SCYL2 | AIRg | -0.41 | 0.0409165 | 0.3340179 |
| SDCBP | auc_glucose | -0.50 | 0.0053264 | 0.1166323 |
| SDCBP | Heart_Rate_Average | -0.43 | 0.016616 | 0.2124584 |
| SDCBP | Glucose_bsl | -0.42 | 0.020522 | 0.2402263 |
| SEC14L3 | auc_glucose | -0.55 | 0.0015798 | 0.0675769 |
| SEC14L3 | Glucose_bsl | -0.53 | 0.0028489 | 0.0877941 |
| SEC14L3 | DEXA__Fat_Mass | -0.38 | 0.0380345 | 0.3234978 |
| SEC22B | auc_glucose | 0.58 | 0.0008213 | 0.0577504 |
| SEC22B | Glucose_bsl | 0.47 | 0.0081645 | 0.1486119 |
| SEC22B | HbA1C | 0.38 | 0.036102 | 0.3151561 |
| SEPTIN11 | auc_glucose | 0.43 | 0.0169861 | 0.214439 |
| SEPTIN11 | HbA1C | 0.40 | 0.0266659 | 0.2732061 |
| SEPTIN11 | Glucose_bsl | 0.40 | 0.0296216 | 0.2890641 |
| SEPTIN11 | DEXA_Fat_Percentage | 0.39 | 0.0354012 | 0.3117143 |
| SEPTIN7 | Waist_Circumference | -0.65 | 8.75E-05 | 0.0288756 |
| SEPTIN7 | DEXA_Weight | -0.65 | 0.0001152 | 0.0311114 |
| SEPTIN7 | HDL | 0.62 | 0.0002281 | 0.0365858 |
| SEPTIN7 | DEXA__Lean_Mass | -0.52 | 0.0032096 | 0.0941435 |
| SEPTIN7 | MATSUDA | 0.49 | 0.0062117 | 0.1272053 |
| SEPTIN7 | QUICKI | 0.47 | 0.0095112 | 0.159285 |
| SEPTIN7 | HOMA_IR | -0.38 | 0.0359937 | 0.3150141 |
| SEPTIN7 | Triglycerides | -0.37 | 0.0411736 | 0.3345073 |
| SEPTIN7 | Insulin_bsl | -0.37 | 0.042427 | 0.339482 |
| SERPINA10 | auc_glucose | -0.44 | 0.0146762 | 0.1990304 |
| SERPINA10 | DEXA__Fat_Mass | -0.43 | 0.0169129 | 0.214439 |
| SERPINA12 | DEXA_Weight | 0.70 | 1.89E-05 | 0.0200487 |
| SERPINA12 | DEXA__Fat_Mass | 0.53 | 0.0028563 | 0.0877941 |
| SERPINA12 | DEXA__Lean_Mass | 0.48 | 0.006682 | 0.1329405 |
| SERPINA12 | Triglycerides | 0.39 | 0.0340397 | 0.3049782 |
| SH3BGRL | auc_glucose | 0.52 | 0.0033536 | 0.0956337 |
| SH3BGRL | Glucose_bsl | 0.47 | 0.0081334 | 0.148309 |
| SH3BGRL | HbA1C | 0.36 | 0.0486421 | 0.3623594 |
| SIGLEC9 | auc_glucose | 0.63 | 0.0002175 | 0.0355725 |
| SIGLEC9 | HbA1C | 0.58 | 0.0007431 | 0.0544917 |
| SIGLEC9 | Glucose_bsl | 0.56 | 0.0014278 | 0.0650391 |
| SIGLEC9 | DI | -0.48 | 0.014958 | 0.1996867 |
| SIGLEC9 | AIRg | -0.45 | 0.0250142 | 0.2651852 |
| SLC14A1 | HOMA_B | 0.43 | 0.0175909 | 0.2185153 |
| SLC14A1 | auc_INSULIN | 0.42 | 0.0194299 | 0.2328065 |
| SLC14A1 | DEXA_Weight | 0.42 | 0.0221595 | 0.24944 |
| SLC14A1 | QUICKI | -0.38 | 0.0371239 | 0.3189234 |
| SLC14A1 | Insulin_bsl | 0.38 | 0.0383509 | 0.3236742 |
| SLC14A1 | Waist_Circumference | 0.37 | 0.0432297 | 0.3429389 |
| SLC14A1 | HDL | -0.37 | 0.0441978 | 0.345115 |
| SLC6A6 | DEXA_Weight | 0.40 | 0.0293974 | 0.2882921 |
| SLC6A6 | DEXA__Fat_Mass | 0.40 | 0.0294237 | 0.2882921 |
| SLC6A6 | Insulin_bsl | 0.40 | 0.0304014 | 0.2916211 |
| SLC6A6 | HDL | -0.39 | 0.0308795 | 0.293527 |
| SLC6A6 | Heart_Rate_Average | 0.38 | 0.0370562 | 0.3186086 |
| SLC6A6 | QUICKI | -0.36 | 0.0492517 | 0.3645404 |
| SLC9A3R1 | auc_glucose | 0.55 | 0.0017536 | 0.0690509 |
| SLC9A3R1 | Glucose_bsl | 0.54 | 0.001859 | 0.0710068 |
| SLC9A3R1 | DEXA__Fat_Mass | 0.45 | 0.0117924 | 0.176847 |
| SLC9A3R1 | DEXA_Fat_Percentage | 0.37 | 0.0469116 | 0.3556091 |
| SLC9A3R1 | Heart_Rate_Average | 0.36 | 0.0479204 | 0.359613 |
| SLIT3 | auc_glucose | 0.59 | 0.0005912 | 0.0500213 |
| SLIT3 | Glucose_bsl | 0.56 | 0.0013581 | 0.0649714 |
| SLIT3 | DEXA__Fat_Mass | 0.41 | 0.023826 | 0.2581288 |
| SLIT3 | DEXA_Fat_Percentage | 0.40 | 0.026863 | 0.2743311 |
| SNAP23 | Triglycerides | -0.63 | 0.0002159 | 0.0355725 |
| SNX2 | DEXA__Fat_Mass | -0.52 | 0.0032798 | 0.0951158 |
| SNX2 | MATSUDA | 0.46 | 0.0109317 | 0.1702964 |
| SNX2 | DEXA_Fat_Percentage | -0.44 | 0.0157002 | 0.2061106 |
| SNX2 | QUICKI | 0.39 | 0.0341407 | 0.3053032 |
| SPRR2E | auc_glucose | 0.52 | 0.0034799 | 0.0984157 |
| SPRR2E | Glucose_bsl | 0.44 | 0.0159774 | 0.2085655 |
| SRI | DEXA_Fat_Percentage | 0.52 | 0.0031456 | 0.093781 |
| SRI | auc_glucose | 0.50 | 0.0045743 | 0.1115441 |
| SRI | Glucose_bsl | 0.48 | 0.0071804 | 0.1385601 |
| SRI | DEXA__Lean_Mass | -0.48 | 0.007286 | 0.1395492 |
| SRI | DEXA__Fat_Mass | 0.43 | 0.0182792 | 0.2244628 |
| SSH3 | auc_glucose | -0.45 | 0.0125882 | 0.1809996 |
| SSH3 | Glucose_bsl | -0.41 | 0.0247713 | 0.2647046 |
| ST6GAL1 | Waist_Circumference | 0.51 | 0.0037939 | 0.1015905 |
| ST6GAL1 | auc_glucose | 0.50 | 0.0053358 | 0.1166323 |
| ST6GAL1 | Insulin_bsl | 0.49 | 0.0056003 | 0.1201645 |
| ST6GAL1 | HOMA_IR | 0.48 | 0.0076399 | 0.1426019 |
| ST6GAL1 | HbA1C | 0.45 | 0.0125196 | 0.1809996 |
| ST6GAL1 | DEXA__Fat_Mass | 0.42 | 0.0205766 | 0.2403181 |
| ST6GAL1 | DEXA_Weight | 0.41 | 0.0230808 | 0.2536913 |
| ST6GAL1 | Glucose_bsl | 0.37 | 0.0441084 | 0.345115 |
| STAT3 | auc_glucose | -0.56 | 0.0014058 | 0.0650071 |
| STAT3 | Glucose_bsl | -0.39 | 0.0354078 | 0.3117143 |
| STAT3 | HOMA_B | 0.36 | 0.0496522 | 0.3659398 |
| STEAP3 | auc_glucose | -0.55 | 0.0016864 | 0.0688865 |
| STEAP3 | Glucose_bsl | -0.45 | 0.0117376 | 0.176847 |
| STEAP3 | MATSUDA | 0.44 | 0.014188 | 0.1947855 |
| STEAP3 | Sg | 0.41 | 0.0433101 | 0.3430719 |
| STIM1 | DEXA_Weight | -0.42 | 0.0216269 | 0.2463397 |
| STIM1 | Heart_Rate_Average | -0.40 | 0.0297574 | 0.2895631 |
| STK38 | LDL | -0.46 | 0.012639 | 0.1809996 |
| STK38 | HbA1C | 0.41 | 0.0239236 | 0.2582538 |
| STK38 | auc_glucose | 0.40 | 0.0278806 | 0.2796888 |
| STK4 | auc_glucose | -0.50 | 0.005254 | 0.1157448 |
| STK4 | Glucose_bsl | -0.46 | 0.0100488 | 0.1628255 |
| STK4 | MATSUDA | 0.46 | 0.0101545 | 0.1629389 |
| STK4 | DEXA__Fat_Mass | -0.36 | 0.0479978 | 0.3598213 |
| STX7 | auc_glucose | -0.54 | 0.0023168 | 0.0792826 |
| STX7 | Glucose_bsl | -0.43 | 0.0166357 | 0.2124584 |
| STXBP2 | auc_glucose | 0.61 | 0.0003419 | 0.0422842 |
| STXBP2 | Glucose_bsl | 0.58 | 0.0007851 | 0.0567598 |
| STXBP2 | HbA1C | 0.44 | 0.0140707 | 0.1940539 |
| STXBP2 | DEXA__Fat_Mass | 0.43 | 0.0175374 | 0.2182293 |
| STXBP2 | MATSUDA | -0.41 | 0.024995 | 0.2651852 |
| STXBP2 | Heart_Rate_Average | 0.39 | 0.0340199 | 0.3049782 |
| STXBP2 | Respiration_Rate | 0.38 | 0.0408061 | 0.3340179 |
| SYK | auc_glucose | 0.54 | 0.0022068 | 0.0782674 |
| SYK | Glucose_bsl | 0.47 | 0.0082372 | 0.1491405 |
| SYNCRIP | auc_glucose | -0.54 | 0.0022649 | 0.0784083 |
| SYNCRIP | Glucose_bsl | -0.50 | 0.0049401 | 0.1157374 |
| SYNCRIP | DEXA__Fat_Mass | -0.48 | 0.0075922 | 0.1422299 |
| SYNCRIP | MATSUDA | 0.41 | 0.0253605 | 0.2673004 |
| SYTL4 | auc_glucose | -0.54 | 0.0021105 | 0.0767053 |
| SYTL4 | Glucose_bsl | -0.52 | 0.0030313 | 0.0923418 |
| SYTL4 | DEXA__Fat_Mass | -0.50 | 0.0046827 | 0.1129307 |
| SYTL4 | HbA1C | -0.47 | 0.0085598 | 0.1533592 |
| SYTL4 | Waist_Circumference | -0.46 | 0.0102077 | 0.1634819 |
| SYTL4 | HDL | 0.43 | 0.0174017 | 0.2174972 |
| SYTL4 | TSH | -0.40 | 0.0271847 | 0.2760421 |
| SYTL4 | DEXA_Weight | -0.40 | 0.0302397 | 0.290967 |
| TAOK1 | auc_glucose | -0.56 | 0.0011541 | 0.0649714 |
| TAOK1 | HbA1C | -0.52 | 0.0031702 | 0.093781 |
| TAOK1 | Glucose_bsl | -0.51 | 0.0041755 | 0.1061023 |
| TAOK1 | LDL | 0.37 | 0.0470097 | 0.3559007 |
| TBCB | Waist_Circumference | -0.49 | 0.0055957 | 0.1201645 |
| TBCB | HbA1C | -0.48 | 0.0071721 | 0.1385601 |
| TBCB | auc_glucose | -0.47 | 0.0094024 | 0.159257 |
| TBCB | DEXA_Weight | -0.45 | 0.0126115 | 0.1809996 |
| TBCB | HDL | 0.40 | 0.0279882 | 0.2800458 |
| TBCB | DEXA__Fat_Mass | -0.40 | 0.0306372 | 0.2924778 |
| TBCB | Glucose_bsl | -0.39 | 0.0317734 | 0.2973439 |
| TBCB | Triglycerides | -0.39 | 0.0324753 | 0.3008944 |
| TBCB | TSH | -0.37 | 0.0472789 | 0.3563618 |
| TGFB1 | auc_INSULIN | -0.53 | 0.0024709 | 0.0813784 |
| TGFB1 | HOMA_B | -0.47 | 0.009544 | 0.1595743 |
| TGFB1 | Heart_Rate_Average | -0.40 | 0.0305911 | 0.2924101 |
| TGFB1 | Insulin_bsl | -0.38 | 0.0395756 | 0.329413 |
| TIMD4 | auc_glucose | 0.59 | 0.0005821 | 0.0500213 |
| TIMD4 | HbA1C | 0.47 | 0.009027 | 0.1581892 |
| TIMD4 | Glucose_bsl | 0.45 | 0.0126172 | 0.1809996 |
| TKT | auc_glucose | 0.61 | 0.0003249 | 0.041706 |
| TKT | Glucose_bsl | 0.56 | 0.0013708 | 0.0649714 |
| TKT | HbA1C | 0.47 | 0.0086088 | 0.1535778 |
| TKT | DEXA__Fat_Mass | 0.36 | 0.0476963 | 0.3585229 |
| TMEM126A | auc_glucose | -0.58 | 0.0008847 | 0.0593623 |
| TMEM126A | Glucose_bsl | -0.46 | 0.0097956 | 0.1620637 |
| TMEM126A | HOMA_IR | -0.39 | 0.0313865 | 0.2950241 |
| TMEM201 | TSH | 0.51 | 0.0038029 | 0.1015905 |
| TMEM201 | Waist_Circumference | 0.50 | 0.0052423 | 0.1157374 |
| TMEM201 | HDL | -0.40 | 0.0274216 | 0.2776236 |
| TMEM201 | DEXA_Weight | 0.39 | 0.0312326 | 0.294793 |
| TMEM201 | DEXA__Lean_Mass | 0.37 | 0.0446603 | 0.3460245 |
| TNIK | auc_glucose | -0.43 | 0.016952 | 0.214439 |
| TNIK | DEXA__Lean_Mass | 0.40 | 0.0296141 | 0.2890641 |
| TNIK | Glucose_bsl | -0.36 | 0.049924 | 0.3666092 |
| TPM1 | DEXA__Lean_Mass | -0.46 | 0.0104262 | 0.1659457 |
| TPM1 | DEXA_Weight | -0.43 | 0.017474 | 0.2177037 |
| TPM1 | auc_glucose | 0.40 | 0.0266427 | 0.2732061 |
| TPM1 | HOMA_B | -0.37 | 0.0419599 | 0.3373225 |
| TPM2 | TSH | -0.46 | 0.0103764 | 0.1654633 |
| TPM2 | auc_c_peptide | 0.59 | 0.013014 | 0.1837718 |
| TPM2 | Heart_Rate_Average | -0.40 | 0.0290637 | 0.2868927 |
| TPM2 | auc_glucose | -0.39 | 0.0331761 | 0.3024743 |
| TPM4 | auc_glucose | 0.63 | 0.00019 | 0.0355725 |
| TPM4 | Glucose_bsl | 0.56 | 0.0012543 | 0.0649714 |
| TPM4 | HbA1C | 0.47 | 0.0093918 | 0.159257 |
| TPT1 | auc_glucose | 0.44 | 0.0154447 | 0.2037991 |
| TREML1 | auc_glucose | 0.65 | 0.0001071 | 0.0305397 |
| TREML1 | Glucose_bsl | 0.48 | 0.0074621 | 0.1408198 |
| TREML1 | HbA1C | 0.45 | 0.0128911 | 0.1832612 |
| TREML1 | DI | -0.44 | 0.0268482 | 0.2743311 |
| TRRAP | TSH | 0.40 | 0.0306837 | 0.2924778 |
| TRRAP | LDL | 0.38 | 0.0435611 | 0.3439229 |
| TSPAN18 | HOMA_B | -0.52 | 0.003186 | 0.093781 |
| TSPAN18 | auc_INSULIN | -0.42 | 0.0217409 | 0.2465322 |
| TSPAN18 | Insulin_bsl | -0.38 | 0.0365798 | 0.3178999 |
| TYMP | DI | 0.55 | 0.0040002 | 0.1042278 |
| TYMP | auc_glucose | -0.43 | 0.0172151 | 0.2163114 |
| TYMP | AIRg | 0.47 | 0.017362 | 0.2173637 |
| TYMP | HbA1C | -0.39 | 0.0310097 | 0.2938775 |
| UBA7 | HbA1C | -0.60 | 0.0004848 | 0.0483802 |
| UBA7 | auc_glucose | -0.55 | 0.0016522 | 0.0686713 |
| UBA7 | DI | 0.57 | 0.0027333 | 0.0850297 |
| UBA7 | Waist_Circumference | -0.51 | 0.0037353 | 0.1015759 |
| UBA7 | Triglycerides | -0.49 | 0.0060387 | 0.126259 |
| UBA7 | Glucose_bsl | -0.46 | 0.0098635 | 0.162274 |
| UBA7 | DEXA_Weight | -0.46 | 0.0103797 | 0.1654633 |
| UBA7 | HOMA_IR | -0.45 | 0.0131285 | 0.185133 |
| UBA7 | Insulin_bsl | -0.40 | 0.0281348 | 0.2805768 |
| UFM1 | auc_glucose | 0.64 | 0.0001371 | 0.0344917 |
| UFM1 | Glucose_bsl | 0.64 | 0.0001602 | 0.0355725 |
| UFM1 | HbA1C | 0.58 | 0.0007104 | 0.0532352 |
| UFM1 | DI | -0.43 | 0.0314106 | 0.2950241 |
| UFM1 | DEXA_Fat_Percentage | 0.38 | 0.0397599 | 0.3296245 |
| UMOD | auc_glucose | -0.63 | 0.0002058 | 0.0355725 |
| UMOD | Glucose_bsl | -0.56 | 0.0013336 | 0.0649714 |
| UMOD | HbA1C | -0.49 | 0.0061786 | 0.1271133 |
| UMOD | LDL | 0.46 | 0.0118546 | 0.176847 |
| UNC5A | auc_glucose | -0.54 | 0.0022828 | 0.0786429 |
| UNC5A | Glucose_bsl | -0.50 | 0.0051697 | 0.1157374 |
| UNC5A | DEXA__Fat_Mass | -0.47 | 0.0081878 | 0.1487718 |
| UNC5A | MATSUDA | 0.42 | 0.0225719 | 0.2505118 |
| UQCRC2 | auc_glucose | -0.47 | 0.0092888 | 0.159257 |
| UQCRC2 | Glucose_bsl | -0.46 | 0.0107855 | 0.1693021 |
| UROD | auc_glucose | 0.49 | 0.0060538 | 0.1263181 |
| UROD | Glucose_bsl | 0.41 | 0.0231321 | 0.2537121 |
| UROD | HbA1C | 0.40 | 0.0264654 | 0.2729776 |
| VBP1 | auc_glucose | -0.56 | 0.0013005 | 0.0649714 |
| VBP1 | DEXA_Weight | -0.47 | 0.009397 | 0.159257 |
| VBP1 | Glucose_bsl | -0.42 | 0.0212031 | 0.243021 |
| VBP1 | Waist_Circumference | -0.41 | 0.0234201 | 0.2552341 |
| VBP1 | TSH | -0.41 | 0.0241564 | 0.2596745 |
| VBP1 | DEXA__Lean_Mass | -0.41 | 0.0254654 | 0.2678561 |
| VBP1 | HbA1C | -0.38 | 0.040149 | 0.3310603 |
| VCAM1 | DI | 0.60 | 0.001527 | 0.0664575 |
| VCAM1 | auc_glucose | -0.53 | 0.0024732 | 0.0813784 |
| VCAM1 | HbA1C | -0.51 | 0.003838 | 0.1015905 |
| VCAM1 | Glucose_bsl | -0.50 | 0.0048366 | 0.1146784 |
| VCAM1 | Sg | 0.45 | 0.0233455 | 0.2549632 |
| VDAC1 | Glucose_bsl | 0.44 | 0.0156308 | 0.2057612 |
| VDAC1 | auc_glucose | 0.42 | 0.019342 | 0.2322403 |
| VDAC1 | DEXA__Fat_Mass | 0.42 | 0.0224444 | 0.2505118 |
| VDAC1 | DEXA_Fat_Percentage | 0.36 | 0.0488864 | 0.3623594 |
| VPS13D | Diastolic_Blood_Pressure_Average | -0.48 | 0.0066642 | 0.1329405 |
| VPS13D | Systolic_Blood_Pressure_Average | -0.41 | 0.0254457 | 0.2678561 |
| VPS13D | DEXA_Fat_Percentage | 0.40 | 0.0276634 | 0.2787634 |
| VPS13D | DEXA__Lean_Mass | -0.36 | 0.0498525 | 0.3666084 |
| VWF | auc_glucose | -0.51 | 0.0037065 | 0.1015759 |
| VWF | Glucose_bsl | -0.48 | 0.0066607 | 0.1329405 |
| WAS | auc_glucose | -0.64 | 0.0001528 | 0.0355725 |
| WAS | Glucose_bsl | -0.60 | 0.0004578 | 0.0483802 |
| WAS | HbA1C | -0.58 | 0.0008832 | 0.0593623 |
| WAS | Triglycerides | -0.45 | 0.01197 | 0.1775781 |
| WAS | DI | 0.49 | 0.0131871 | 0.1851964 |
| WDR78 | Glucose_bsl | 0.49 | 0.0057697 | 0.1223802 |
| WDR78 | HbA1C | 0.48 | 0.0074311 | 0.1408198 |
| WDR78 | auc_glucose | 0.47 | 0.0084891 | 0.1523585 |
| WDR78 | HOMA_B | -0.45 | 0.0135685 | 0.1889134 |
| WDR78 | auc_INSULIN | -0.39 | 0.0341784 | 0.3053732 |
| YWHAQ | DEXA_Fat_Percentage | 0.46 | 0.0105093 | 0.1662058 |
| YWHAQ | Glucose_bsl | 0.44 | 0.0137546 | 0.1905595 |
| YWHAQ | auc_glucose | 0.43 | 0.0162945 | 0.2101496 |
| YWHAQ | DEXA__Lean_Mass | -0.40 | 0.0294302 | 0.2882921 |
| YWHAQ | Heart_Rate_Average | 0.38 | 0.0365106 | 0.3178999 |
| ZC3HAV1L | auc_glucose | -0.46 | 0.0111788 | 0.1722962 |
| ZC3HAV1L | Glucose_bsl | -0.43 | 0.0181623 | 0.2235661 |
| ZFHX3 | HOMA_IR | 0.49 | 0.0063863 | 0.1295699 |
| ZFHX3 | Glucose_bsl | 0.47 | 0.009257 | 0.159257 |
| ZFHX3 | auc_glucose | 0.45 | 0.0127849 | 0.1820739 |
| ZFHX3 | Insulin_bsl | 0.44 | 0.0158353 | 0.207353 |
| ZFHX3 | Heart_Rate_Average | 0.43 | 0.0184121 | 0.2247543 |
| ZFHX3 | HbA1C | 0.41 | 0.025095 | 0.2655929 |
| ZFHX3 | QUICKI | -0.37 | 0.0442816 | 0.345115 |
| ZFX | Heart_Rate_Average | 0.42 | 0.0213193 | 0.2434522 |
| ZNF350 | auc_glucose | 0.54 | 0.0023131 | 0.0792826 |
| ZNF350 | Glucose_bsl | 0.48 | 0.0075902 | 0.1422299 |
| ZNF350 | Ins30_glu30 | -0.40 | 0.0282826 | 0.2813463 |
| ZNF350 | AIRg | -0.43 | 0.0328973 | 0.3022503 |
| ZNF350 | auc_c_peptide | -0.51 | 0.0373861 | 0.3203721 |
| ZNF462 | DEXA__Fat_Mass | -0.67 | 5.33E-05 | 0.0266984 |
| ZNF462 | DEXA_Weight | -0.49 | 0.0058968 | 0.1248021 |
| ZNF462 | Glucose_bsl | -0.44 | 0.015997 | 0.2085655 |
| ZNF462 | auc_glucose | -0.42 | 0.0211259 | 0.2425934 |
| ZNF462 | HbA1C | -0.41 | 0.0253602 | 0.2673004 |
| ZNF462 | Waist_Circumference | -0.37 | 0.0457858 | 0.3509813 |
| ZNF519 | QUICKI | -0.39 | 0.0355734 | 0.3121335 |
| ZNF519 | MATSUDA | -0.37 | 0.0431082 | 0.3425303 |
| ZNF519 | HDL | -0.37 | 0.0439672 | 0.345115 |
| ZYX | auc_glucose | 0.64 | 0.0001598 | 0.0355725 |
| ZYX | Glucose_bsl | 0.63 | 0.0001828 | 0.0355725 |
| ZYX | DEXA_Fat_Percentage | 0.44 | 0.0156335 | 0.2057612 |
| ZYX | HbA1C | 0.44 | 0.0159919 | 0.2085655 |
| ZYX | Heart_Rate_Average | 0.40 | 0.0281779 | 0.2805768 |
